# Is there an association between clinician behavioural factors, and the experience of pain in a dental setting? A Scoping Review

**DOI:** 10.1101/2023.06.18.23291577

**Authors:** Belinda Tang, Jasmine Ting, Rachel Brown, Sandhya Nathan, Claire Ashton-James, Atieh Sadr, Ali Gholamrezaei

## Abstract

**Background:** Effective management of pain is critical in a dental setting, and insufficient pain management can lead to anxiety in patients and hesitancy to seek further dental help. Currently, there are no existing scoping or systematic reviews discussing the impact of clinician behavioural factors on pain perception, thus highlighting the need for it.

**Aims:** The aim of this scoping review is two-fold: to analyse the scope of current evidence surrounding the association between clinician behaviour and pain experienced in the dental setting, and to identify areas where further research is needed.

**Methodology:** A list of search terms and subject headings was developed for Pubmed, PsycINFO and Embase to conduct pilot searches. Duplicate articles were removed, and each article screened in accordance with pre-established inclusion and exclusion criteria. Relevant articles were assessed using a data extraction form. Information was analysed to determine the scope of current evidence and areas where further study is needed.

**Results:** Dentist behaviour has a significant impact on the patient’s pain experience, and techniques to minimise pain include empathy, being calm and non-judgemental, and providing clear information to patients about treatment. Following up the patient after treatment, via a phone call or text message, reduces pain experience. Increased levels of dental anxiety have been shown to increase dental pain, and therefore it is important that clinicians implement behaviours to reduce dental anxiety in order to reduce pain.

**Conclusion:** Numerous studies have shown the association between clinician behavioural factors and the perception of pain in the dental setting. There are various techniques dentists can utilise to improve the pain experience for their patients, including displaying a warm empathetic demeanour, appropriately managing dentally anxious patients, and utilising non-judgemental communication skills. However, there are some areas where further research is needed.

**Highlights:**

- Clinician behaviour can be modified to alter the perception of pain in a dental setting
- Various studies outline techniques that a dentist can use to improve a patient’s pain experience
- Further research is required for chronic orofacial pain, phobic patients and patients with special needs

## Background

Health care and pain management are often linked, with the effective management of the latter being a hallmark of ethical, professional medical practice.(1) Within the field of dentistry, there are many types of pain, with acute orofacial pain being a common factor driving patients to seek professional treatment. Other types of pain experienced in the dental setting include procedural pain and post-procedural pain, resulting in pain management challenges for dental practitioners. Inadequate pain management may lead to the development or progression of negative health outcomes including anxiety or poor wound healing. It may also affect the psychological and social health of both patients and their loved ones, resulting in future hesitancy when seeking further medical help.(2,3) Therefore, exploring different factors that can influence a patient’s experience of pain within the dental setting is needed as it would allow for a more positive dental care experience.

An important moderator of patients’ pain experience is the degree to which they feel supported, reassured, cared for, and respected by others. Support from others can also provide emotional or informational support.(4,5) All of these forms of social support have strong correlations with physical and mental health outcomes and pain.(6) In a dental setting, examples of social support include, but are not limited to; appointment accompaniment by trusted others, patient social network, patient previous dental experiences and clinician behaviour.(7,8,9,10) In particular, clinician behavioural attributes such as concordance behaviour, communicativeness and friendliness have demonstrated significant impact on a patient’s dental pain experience.(11,12,13) A study by Ashton-James et al.(14) found that dentist competence and warmth may increase trust in the dental clinician, which is associated with less reported pain and less postoperative pain during wisdom tooth extraction. Therefore, examining the role of clinician behaviour as an adjustable factor allows for better pain management and improved patient care.

However, the experience of pain is subjective, with inter-individual variation and a large emotional component (in addition to the sensory discriminative component).(15,16) This can complicate the evaluation of pain as the level of reported or expressed pain may differ from the physiological pain experienced by the patient.(17) Additionally, the extent of this discrepancy can also be influenced by clinician behaviour and other social factors.(18) This concept was explored by a study conducted by Gallant et al (19) that compared patient expressed pain with physiological pain as measured through facial expressions in the presence of family, a stranger and alone. They observed lower self-reported pain but higher non verbal pain expressiveness in the presence of a stranger, but the opposite when in the presence of family. Conversely, Lumley et al (20) mentions that patients tend to report less pain to avoid burdening any present family. Krahe et al (21) suggested that attachment style is a possible reason for this variability in a similar style investigation that looked at pain related neural processing to measure physiological pain level. Therefore, there is a need to consider that the literature focusing on patient reported pain will present a higher degree of analytical complexity compared to those that measure actual experienced physiological pain.

## Clinical question

The clinical question that is being considered is: **What is the association between clinician behaviour and patients’ pain experience in a dental setting?**

## Aims

This scoping review has two aims:

### Aim 1: Investigate the scope of current research

The scope of current research available about clinician behaviour influencing pain perception in a dental setting was investigated. Aspects of clinician behaviour that we aimed to investigate included: dentist-patient rapport, dentist communication, perceived dentist interpersonal skills, the patient’s perceptions of dental treatment shaped by the patient’s previous dental experiences, and the level of control experienced by the patient while in the dental setting.

### Aim 2: Identify gaps in current research

After investigating the current research available regarding the relationship between clinician behaviour and perception of dental pain, gaps in research were identified and potential areas for further research were revealed.

To the best of our knowledge, there is no existing scoping or systematic review performed on this topic area thus highlighting the need for it. This will be helpful in allowing dental practitioners to develop a deeper understanding and appreciation of the way their actions and patient relationships influence pain, and incorporate this information into clinical practice to enhance patient management and the patient experience.

## Methodology

The scoping review was carried out in accordance with Preferred Reporting Items for Systematic reviews and Meta-Analyses (PRISMA,2020) guidelines (Figure 1). A list of potential search terms was developed by reading relevant articles found during an initial search, examining current systematic reviews and scoping reviews regarding pain in general, and by using the MeSH database. Once relevant search terms were established, subject headings were developed for each of the following three databases: Pubmed, Embase and PsycINFO (Table 1).

**Figure 1:**
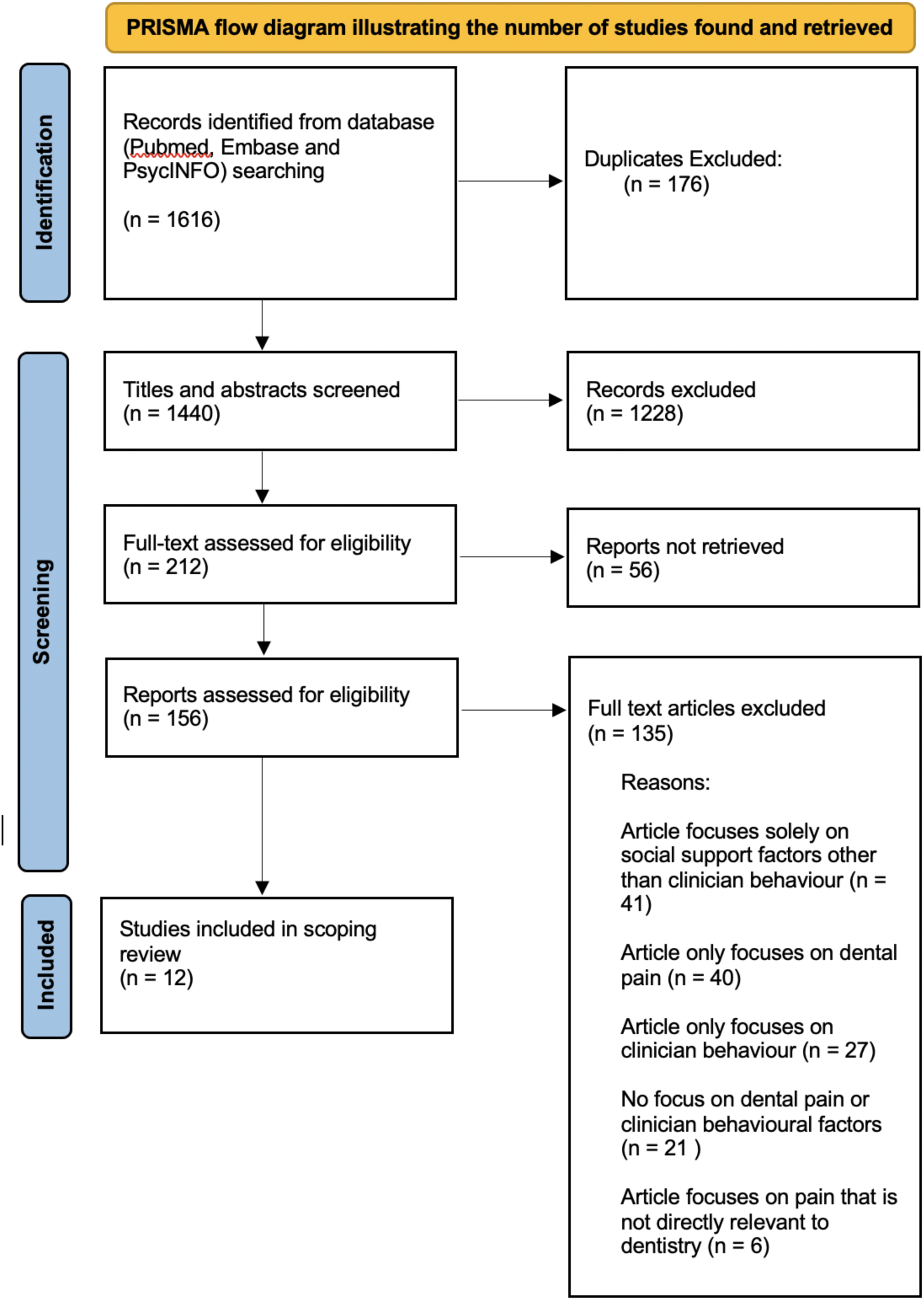
PRISMA Flow Diagram (22)

**Table 1:**
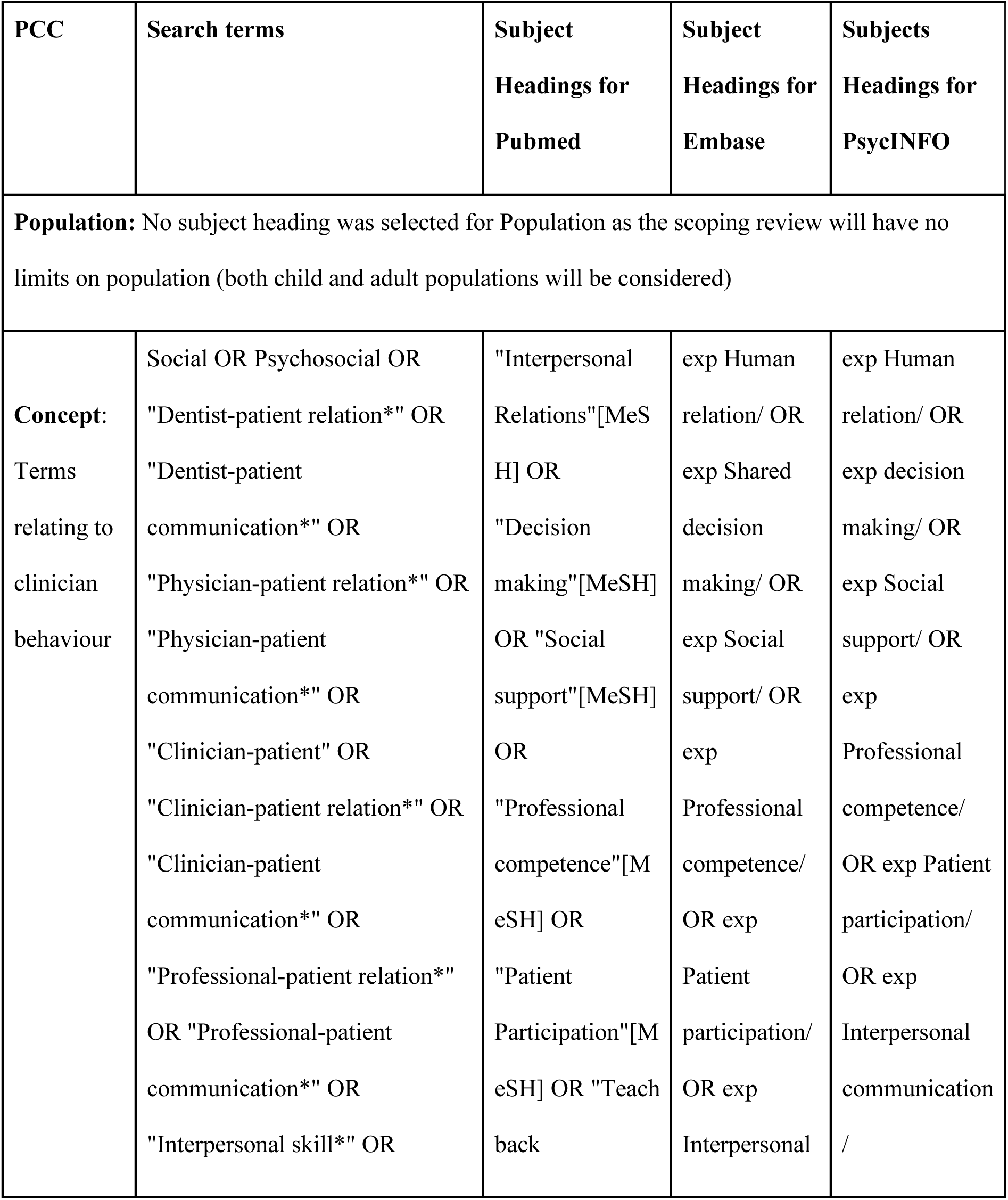
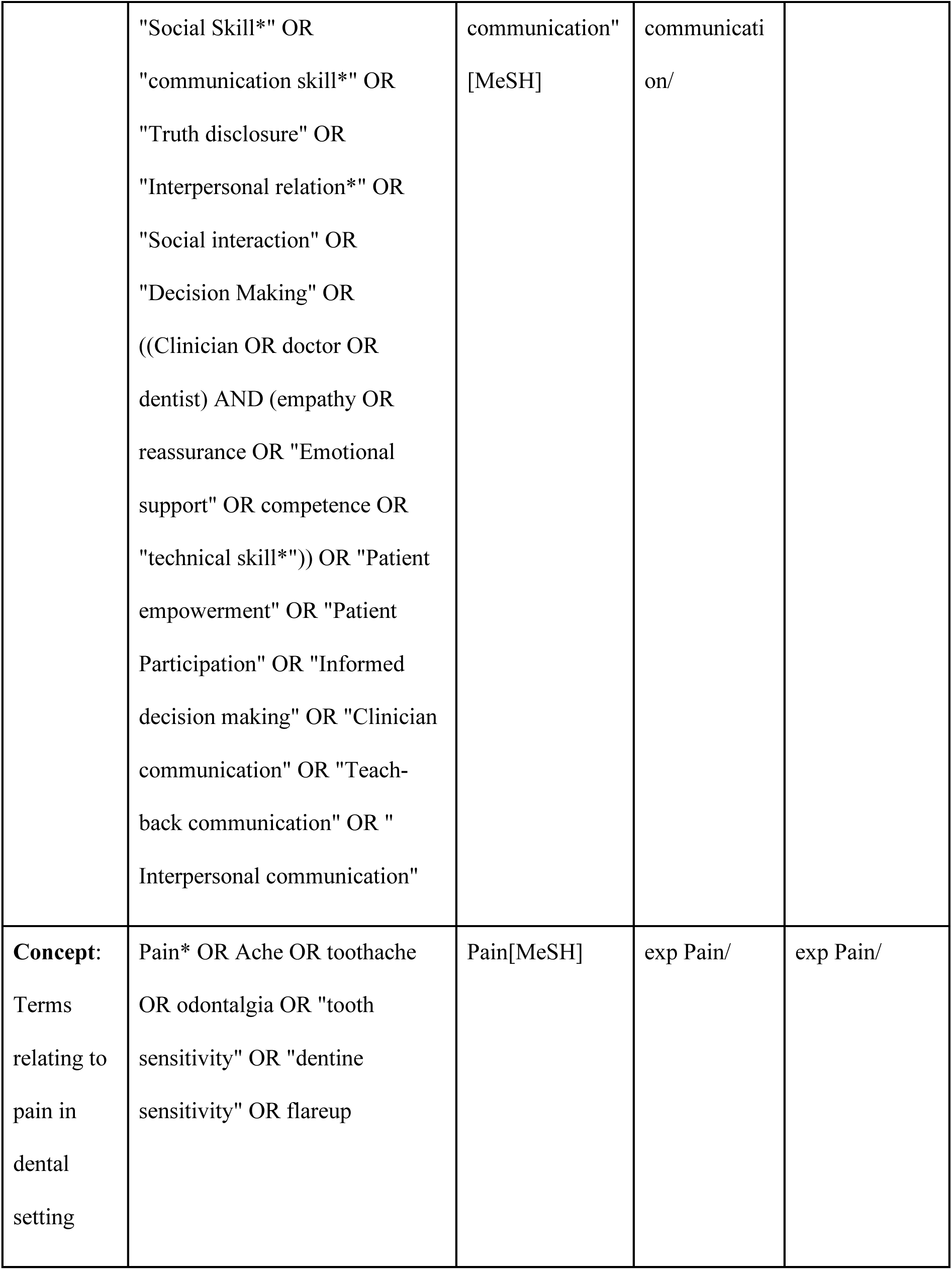
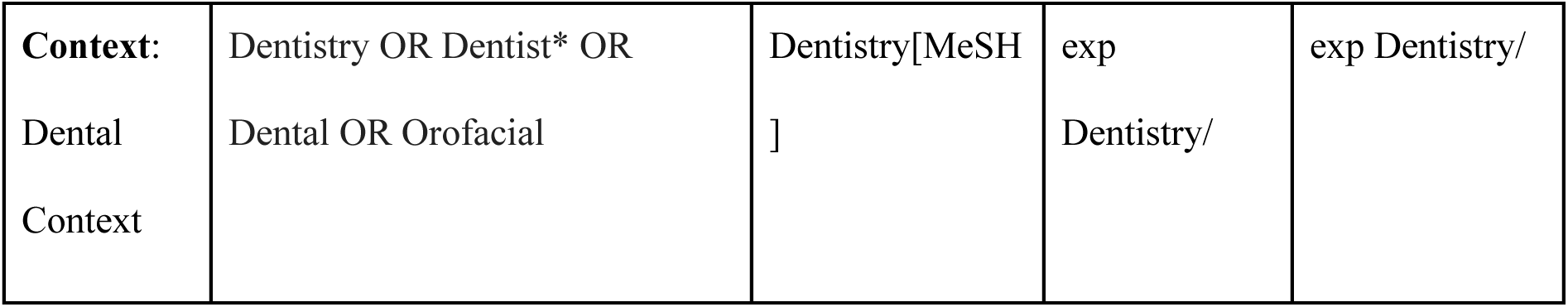
Search terms and subject headings for Pubmed, Embase and PsycINFO databases

Subsequently, a keyword and subject heading search was conducted using two of the three databases, Pubmed and Embase, from the beginning of each database until August 26th, 2022. Once keywords had been adjusted in order to identify a suitable number of articles (Table 2), a pilot search was conducted for Pubmed. In order to reduce bias, 100 articles were selected from Pubmed at random, with 50 articles being screened by two individuals, to determine if articles met the inclusion and exclusion criteria (Table 3, Figure 2). An agreement of 80% between each pair was achieved, and therefore the pilot search was deemed successful.

**Table 2:** Keyword and subject heading search

| Database | Number of articles identified |
| --- | --- |
| Pubmed | 856 |
| Embase | 536 |
| PsycINFO | 224 |

**Table 3:** Inclusion and exclusion criteria

| Inclusion criteria | Exclusion criteria |
| --- | --- |
| Articles that examine clinician behaviour as a social support factor<br>Articles relating to dental pain or pain in a dental setting | Articles not written in English<br>Articles not available in full text<br>Articles focusing solely on social support factors other than clinician behaviour<br>Articles focusing on pain that is not directly relevant to dentistry<br>Articles that do not focus on clinician behaviour or dental pain |

All of the articles from the three databases were exported to Endnote and duplicate articles removed. Subsequently, the articles were uploaded to Covidence for ease of collaboration, and the title and abstract of each article was assessed in accordance with the pre-established inclusion and exclusion criteria (Table 3). These articles then underwent an additional full text screening for eligibility through Covidence, with each article being screened by two individuals independently. This was to minimise errors and the introduction of the potential biases due to the nature of the data being extracted being affected by subjective interpretation. Any articles that had discrepancies underwent further review by an additional person. Any articles that could not be retrieved due to full text not being available were excluded, as well as any articles that did not meet the inclusion criteria.

The data extraction process was then carried out, with a data extraction form being created (Table 5) to extract details relating to the clinical question about each article in a systematic manner. The data extraction for each article was again done by two individuals for the same reasons stated previously (to minimise errors and potential biases), then reviewed again for consistency and accuracy. This was then uploaded to Covidence.

**Table 4:** Summary of data extracted from articles

| **Author** | **Study design** | **Location** | **Population/**<br>**procedure** | **Clinician behavioural**<br>**factors** | **Pain factors** |
| --- | --- | --- | --- | --- | --- |
| Bartlett et al. 2005 (23) | RCT | USA | Patients having orthodontic appliance placed | Phone call demonstrating care and assurance | Post-procedural orthodontic pain |
| Brown and Smith, 1979 (24) | Case reports | Australia | Paediatric dental patients having a dental extraction | Dentist breaking trust and going beyond child's limit/expectations, describing and demonstrating procedure, forewarning, being reassuring, being sympathetic, giving realistic explanations | Procedural pain |
| Corah et al. 1988 (25) | Cross-sectional study | Not specified | Public dental patients undergoing invasive procedures | Communication, empathy, paying attention to patient, not rushing, having a calm manner, taking patient seriously, not criticising, being reassuring, being understandable | Pre- and post-procedural pain |
| Council of | Reference | USA | Dentists | Dentist's attitude, non-verbal | Pre-procedural |
| Clinical Affairs (26) | manual |  | treating childhood dental diseases | communication (body language, posture, facial expression), communications skills (giving clear and specific instructions), being empathetic, verbal reassurance, appropriate physical contact, tell-show-do, ask-tell-ask, | and procedural pain |
| Curry et al., 1988 (27) | Cross sectional study | USA | Paediatric dental patients receiving restorative treatment | Focusing on positives, encouraging effective coping strategies, describing and demonstrating procedure | Procedural pain |
| Dangott et al. 1978 (28) | Narrative review | USA | Dentists | Non verbal communication, allowing for patient response to what they feel, emotional communication, using technical jargon, giving disconfirming messages | Dental pain in general |
| Epstein, 1984 (29) | Review article | N/A | Dental patients suffering from myofascial | Dentist-patient relationship can impact effect of placebo, suggestion and relaxation | Chronic pain (myofascial pain dysfunction) |
|  |  |  | pain<br>dysfunction<br>syndrome |  | syndrome) |
| Foreman,<br>1979 (30) | Narrative<br>review | England,<br>New<br>Zealand | Dental patients | Verbal encouragement<br>(anticipation reward), friendly<br>and empathetic manners,<br>explanations, establishing<br>good relationship, describing<br>and demonstrating procedure | Dental pain in<br>general |
| Foreman,<br>1987 (31) | Opinion piece | New<br>Zealand | Dental patients | Staying with patient during<br>recovery, communication,<br>building trust through initial<br>interview, asking questions<br>about fears, using<br>euphemistic, nontechnical<br>and non threatening language | Dental pain in<br>general |
| Gryll and<br>Katahn,<br>1978 (32) | RCT | USA | Oral surgery<br>patients<br>receiving<br>anaesthetic | Warm attitude | Pre- and post-<br>procedural pain |
| Harper et<br>al. 2004<br>(33) | Conference<br>paper | USA | Paediatric<br>dentists and<br>patients | Communication and building<br>rapport, acknowledging<br>patient's pain experience and interests, talking at an appropriate developmental level, appear relaxed, non verbal communication, describing and demonstrating procedure. | Dental pain in<br>general |
| Johnson et al. 1990 (34) | Cross sectional study | USA | Dental patients receiving treatment | Making comments that suggest negative evaluation of the patient by the dentist, asking personal questions while patient's mouth is full | Dental pain in general |
| Keith et al. 2013 (35) | RCT | USA | Orthodontic patients after placement of appliance | Text message showing concern and reassurance | Post-procedural orthodontic pain |
| Kuscu et al. 2021 (36) | Prospective cohort study | Turkey | Paediatric dental patients receiving various treatments (restorative, endodontic, extraction) | Showing empathy, self efficacy | Procedural pain |
| Loggia et al. 2008 (37) | Narrative review | Unspecified | Dental patients | Forewarning, describing and demonstrating procedure, explanation of steps | Dental pain in general |
| Milgrom, 1994 (38) | Cross sectional study | USA | Dentists treating school-aged children | Comforting manner, asking about distress and pain, communication, giving child control | Dental pain in general |
| Moore et al. 1998 (39) | Cross sectional | USA, Taiwan, China, Denmark, Sweden | Dentists conducting tooth-drilling procedures with local anaesthetic | Creating trust, one way communication, encouraging patients to express concerns and discomfort, phrasing of words | Dental pain in general |
| Radhakrishna et al. 2019 (40) | RCT | India | Paediatric dental patients requiring restorations | Describe and demonstrating procedure (through tell-show-playdoh, or tell-show-do) | Procedural pain |
| Riley et al. 2004 (41) | Prospective longitudinal study | USA | Dental patients | Establishing good relationship, supportive dentist | Pre- and post-procedural pain |
| Selden, 1993 (42) | Retrospective case | USA | Patients receiving root | Clear communication, distraction behaviour, next | Pre-procedural, procedural, and |
|  | controlled study |  | canal treatment | day telephone call, describing and demonstrating procedure | post-procedural pain |
| Smith and Freeman, 2010 (43) | Case controlled study | Scotland | Parents of paediatric dental patients | Attentiveness of the dentist to pain, taking the patient seriously | Dental pain in general |

Once data extraction was complete, the articles were evaluated by four people individually, and the results were synthesised into a table summarising the main clinician behavioural factors and the main dental pain concepts. These collated findings were then used to identify the scope of current research surrounding the clinical question, and potential areas for further study.

## Results

As stated above, PRISMA guidelines were utilised to carry out the scoping review (Figure 1). After the pilot search was deemed successful and all of the articles from Pubmed, Embase and PsycINFO were exported to Endnote, duplicate articles were removed, producing a total of 1440 articles. The articles were assessed against the pre-established inclusion and exclusion criteria, giving a total number of 212 articles. These articles underwent full text screening for eligibility by two independent individuals. A total of 56 articles were not retrieved due to the full text not being available, and a further 135 articles were excluded since they did not meet the inclusion criteria, leading to a final number of 21 articles for data extraction to be included in the scoping review.

Data extraction was conducted for these 21 articles, with each article being assessed by two individuals (Table 4, Table 6). Once data extraction was complete, the articles were evaluated by four people individually, and the findings collated in order to identify the scope of current research surrounding the clinical question, and potential areas for further study.

According to the summary of results (Table 4), it can be seen that studies ranged from the year 1978 to 2021, with only 4 (19%) articles being from the last 10 years, and the majority of the studies took place prior to the year 2000 (57%). In terms of study types, there seems to be a mix of both secondary knowledge synthesis papers (29%), as well as primary research papers (71%) consisting of randomised controlled trials, case reports, case controlled studies, cross sectional studies and cohort studies.

Regarding clinician behavioural factors, all of the 21 articles found that effective communication skills were crucial for clinicians in minimising dental pain. Examples of this include clearly explaining the procedure to the patient prior to treatment, encouraging patients to express their concerns, and providing verbal encouragement. In addition to that, 2 of the articles emphasised the importance of clinicians providing follow-up care to patients post-operatively, through the use of a follow-up phone call or text message. 10 of the articles discussed the importance of non-verbal communication skills, such as having open body language, posture, and facial expressions, and being reassuring and empathetic.

With regards to the population being investigated, 9 of the articles involved paediatric patients, dentists treating paediatric patients, or parents of paediatric patients. These articles found that clinician behaviours used to manage anxious paediatric patients included describing and demonstrating the procedure (including use of tell-show-do technique), forewarning, being reassuring and sympathetic, and giving realistic explanations.

20 of the 21 articles involved dental procedures such as restorative work, endodontics, orthodontics and extractions. Regarding the type of pain factors, 5 articles discussed pre-procedural pain, 6 articles discussed procedural pain, 7 articles discussed post-procedural pain, and 9 articles discussed dental pain in general (note that some articles discussed more than one type of pain). Only 1 of the 21 articles (5%) discussed chronic dental pain, namely myofascial pain dysfunction syndrome.

## Discussion

Upon assessment of 21 articles relevant to our research question, we found evidence of an association between clinician behaviour and patient pain perception in a dental setting. While many of the behavioural factors that were discussed in each of these articles are related to an extent, they can be categorised into a few overarching themes that each have important implications on patients’ pain experience.

### Non-verbal behaviour

Several clinician behaviours that were highlighted in the articles involved dentist demeanour, or non-verbal behaviour, which were noted to play a substantial role in modulating a patient’s perception of pain. Current literature discusses various dentist behaviours that can minimise patients’ dental pain, including friendliness and warmth,(32,26) displaying empathy,(36) and having a calm and non-judgemental demeanour,(25) in order to help develop rapport and foster a positive dentist-patient relationship.(29) Use of distraction techniques was also found to be effective in minimising dental pain.(37) This is similar to experimental findings by Ruben et al.,(47) where the participants’ objective and subjective pain scores to experimentally-induced pain were lower when interacting with highly supportive non-verbal physician behaviours such as nodding, smiling, leaning forward, making eye contact, and using warm tone; all of which are similar to those mentioned in the studies of this scoping review. Overall, these positive interactions not only create a sense of trust, but also result in a decreased perception of dental pain. Therefore, it is crucial for clinicians to be aware of how their non-verbal behaviour can be interpreted by patients.

### Verbal encouragement

Another specific aspect of dentist behaviour that has been associated with a reduction in dental pain experience is the use of verbal encouragement.(23,26) Foreman (30) discussed the concept of anticipation reward, where he encourages the dentist to verbally motivate patients before and during treatment about how the procedure will bring improvement, in order to reduce their pain experience. A similar concept of active verbal support was explored by Brown et al.(48) where they noted that verbal statements such as encouraging remarks, humorous comments, reassuring comments and advice given from another individual (stranger or not) helped to reduce experimental pain. Therefore, it is clear that some evidence exists in regards to the importance of verbal, in addition to non-verbal behaviours of dentists in improving patients’ pain experience.

### Patient empowerment

Additionally, dental pain can be minimised through the provision of information to patients about what is wrong with their tooth, describing steps in treatment, forewarning patients of what to expect post-operatively and how to prevent potential symptoms, and acknowledging patient’s concerns regarding their dental treatment.(28,42) Furthermore, receiving a follow-up phone call or text message after a dental procedure provided assurance to patients, and assisted in minimising post-operative pain experience - more so if the phone call or text contained elements of care and reassurance.(23,35,42) All of these behavioural traits are similar in that they involve informing the patient of their situation (patient empowerment) and making them feel supported and not alone. Thus, feelings of uncertainty and the unknown are eliminated, and the patient is allowed to have a sense of being in control. This is thought to increase patients’ coping strategies while healing or going through a painful experience, which could serve as a possible explanation as to why the pain experience is improved.(42,49,50) This parallels the findings in a medical context, where patients’ sense of control over their situation has also been shown to be positively associated with better pain control through similar mechanisms.(51,52)

### Anxiety and pain

However, just as a dentist’s communication approach can have positive effects on a patient’s pain perception, it can also heighten their pain experience, especially within the context of dental anxiety. It has been shown that anxious patients are more prone to pain, and therefore dentists can utilise behavioural techniques to reduce patient anxiety, which will then reduce the pain experience for that patient.(30,38,40) Further proof of this concept regarding the link between anxiety with reduced pain tolerance has come up extensively in other research in both the dental and medical field.(53,54) In particular, a detailed systematic review and meta-analysis by Lin et al.(55) has found that both dental anxiety and state anxiety had clear impacts on pain at different treatment stages. Other methods of alleviating patient anxiety include employing behavioural techniques, which encompasses concepts such as modelling (patient observes treatment on someone else), describing and demonstrating (e.g. tell-show-do), and distraction techniques.(24,27,30,40) Another way is to manage the patient environment, such as keeping anxious patients away from the sites and sounds of other patients, or filling the room with pleasant odours.(37) Foreman discusses how the experience of pain can be modulated both preoperatively and postoperatively, through the utilisation of effective communication and the creation of an empathic environment. Foreman suggests that the sedative effect of certain pharmacological interventions can be equivalent to clinician’s ability to effectively communicate, ultimately concluding that maximum patient benefit is derived by employing both pharmacological interventions as well as effective communication.(30)

### Effect of negative clinician behaviour

The power of effective communication and interpersonal skills is further reinforced by literature that highlights the negative implications of ineffective or inappropriate behaviour. Johnson et al. discusses how dentists who negatively judged patients’ oral health and hygiene by telling patients that they have “bad teeth”, or lecturing their patients about their poor oral hygiene practices, led to increased anxiety for the patient,(34) which (as discussed previously) can then lead to an increased susceptibility to pain.(30) Similar impacts of negative evaluation by the dentist were also mentioned by Corah et al., who also discusses how a calm and open demeanour, in-depth clear explanations, reassurance, and non-judgemental tone are traits that are most associated with patient satisfaction.(25)

### Paediatric patients

A review of the literature also showed a number of studies specifically addressing the management of dental pain and dental anxiety in paediatric patients. Several of the articles discussed the impact of parental dental anxiety and fear on their child, finding that parents who had negative past dental experiences were more likely to postpone their child’s dental treatment until they were experiencing pain.(43) Techniques that dentists and parents can use to minimise dental fear in children include speaking positively about past-dental experiences around children and modelling positive behaviour in the dental setting.(26,27) Additionally, dentists can reduce dental fear by utilising toys to explain dental procedures to children, and familiarise them with the instruments in a dental setting.(40) Ensuring that the child’s voice and concerns are heard is also essential, and this can be done by ensuring discussions about treatment are between the parent, child, and dentist, in order to minimise the child’s fears. Dentists should discuss treatment with a child truthfully and realistically, but also speak in a relaxed manner that is appropriate for the child’s development level.(33) A study by Milgrom found that dentists’ attitudes towards dental pain in children strongly influenced pain management practices, for example dentists who rated dental procedures as less painful were less likely to provide local anaesthetic if the child was in distress.(38) Therefore, it can be seen that clinician behaviour has a significant impact on dental anxiety and pain in children.

### Significance of patient satisfaction

An article by Riley demonstrated the broader impacts of negative dental experiences and found that, if a patient did not have overly positive views about the health system, they were more likely to consult with a layperson as opposed to a healthcare professional when experiencing dental pain. This may not only delay patients from receiving the care they need, but can also result in misinformation spreading amongst the community which can put other patients at risk of not receiving appropriate dental care. Therefore, it is critical that dentists understand their personal influence over patient satisfaction, in order to best manage individuals presenting with dental pain and to help ensure that their community receives appropriate care in a timely manner.(41)

### Acute vs chronic pain

It was found that most of the studies (95%) focused on either general dental pain, or pain experienced prior to, during, or after treatment procedures. This is unsurprising, because most dental pain that dentists have to deal with is often presented as acute in nature,(44) or associated with the dental treatment itself; for example, local anaesthetic injection or tooth drilling.(39) A study by Epstein (29) was the only study that looked into pain of a more chronic nature as it explored the potential therapeutic value of placeboes in treating myofascial pain dysfunction syndrome. It was found that the quality of the dentist-patient relationship had an effect on modifying patients’ response to treatment with placebo, with patients experiencing improved pain outcomes with placebo when the relationship was more positive. This is similar to other studies outside the dental context - Fuentes et al.(45) noticed that a supportive therapist-patient relationship enhanced the placebo response and improved pain intensity scores in patients with chronic lower back pain, and Kaptchuk et al.(46) also confirmed that a positive physician-patient relationship plays a significant role in the placebo effects of management of patients with irritable bowel syndrome. In each of these studies, the positive relationship was built on various clinician behavioural factors already discussed previously, such as warm/empathetic interactions, active listening, eye contact and physical touch. However, this highlights the need for more research into the area of chronic pain.

## Conclusion

Overall, this scoping review has demonstrated that there is an existing body of work exploring the association between clinician behavioural factors and the perception of pain within the dental setting. This review has highlighted many areas and learnings that dentists may incorporate into daily practice to improve the pain experience for their patients, such as the importance of a pleasant demeanour, appropriate management of dentally anxious patients and clear, non-judgemental communication skills. However, it is important to note that this study has also shown gaps in knowledge - only 19% of the articles were from the last 10 years, thus highlighting the need for more recent studies into this topic. Additionally, most articles explored the impact of clinician behaviour on pre-procedural, procedural and post-procedural pain, particularly in the areas of restorations, orthodontics and extractions, but there is less research surrounding chronic orofacial pain (such as temporomandibular disorder), thus highlighting an area where further research is needed. The studies explored pain experience in adults and children, but further studies could also be conducted to explore the impact of dentist behaviour on pain experienced by dental phobic patients and patients with special needs. Further research into this area would help dentists to improve their clinical practice and patient management, reducing barriers for patients obtaining dental care and improving oral health within the community.

## Data Availability

All data produced in the present work are contained in the manuscript

## Appendix

**Figure 2:**
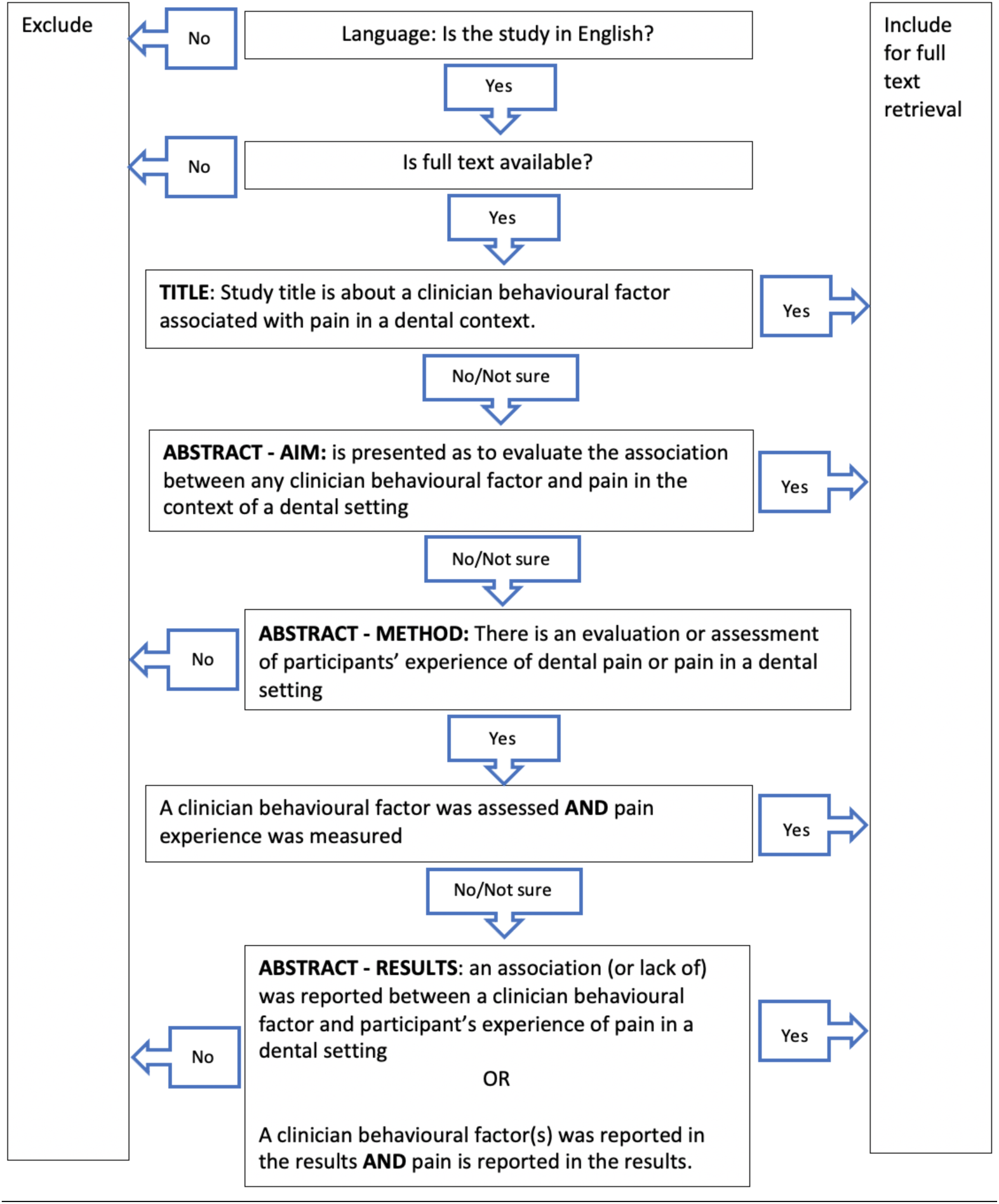
Flowchart showing our process of title and abstract screening, according to inclusion and exclusion criteria

**Figure 3:**
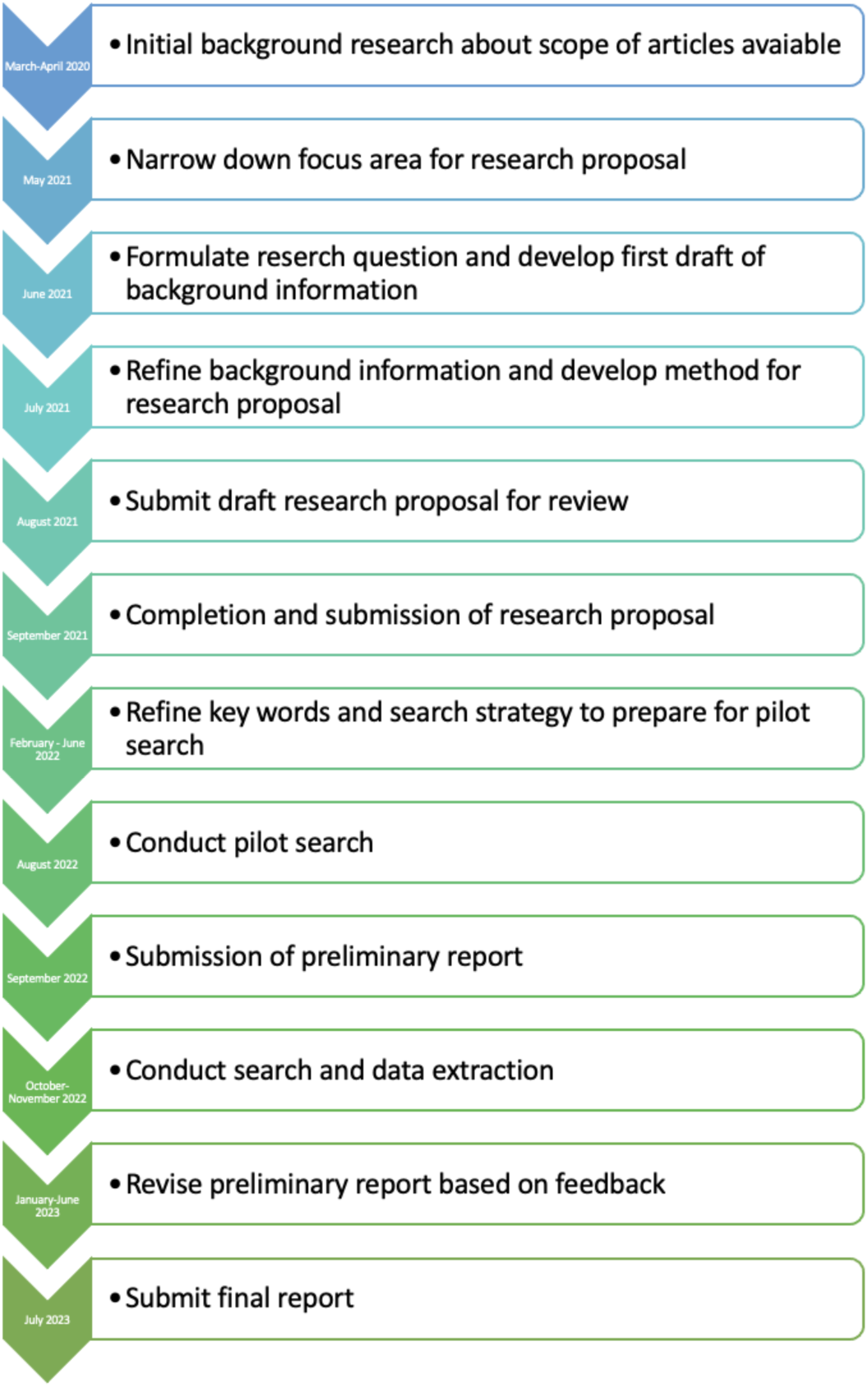
Project timeline

**Figure 4:**
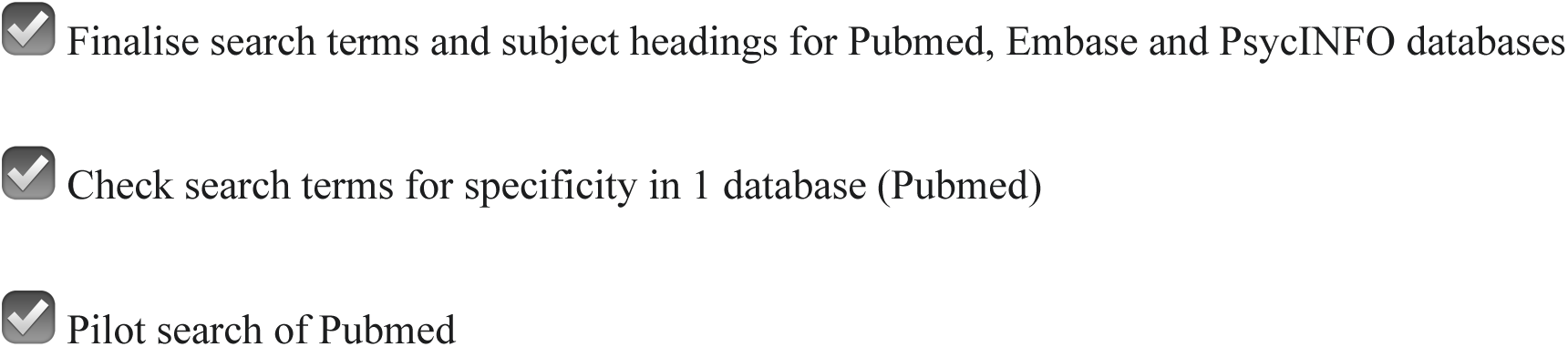
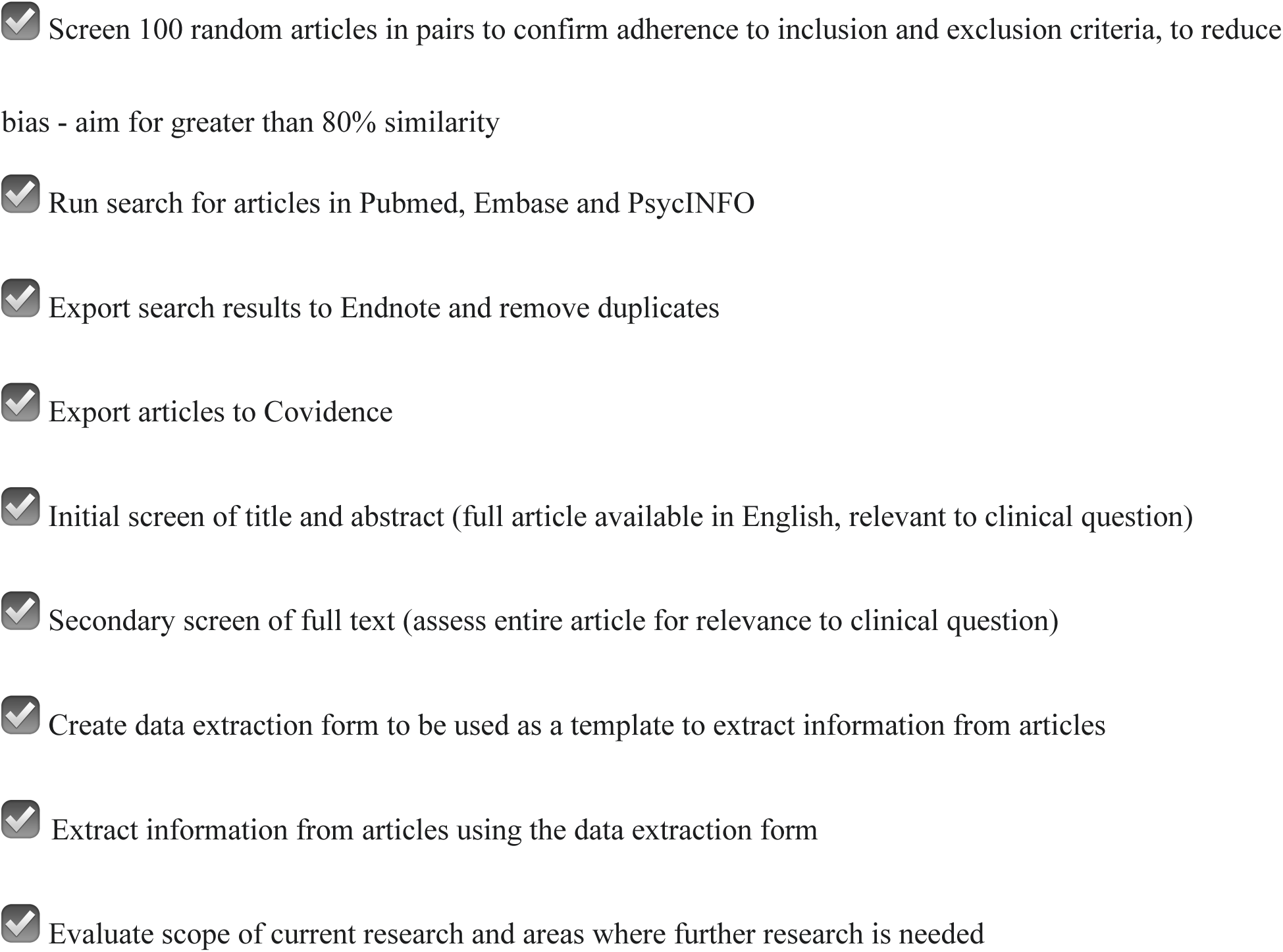
Progress checklist

**Table 5:** Data extraction form template

| General information |  |
| --- | --- |
| Date of data extraction |  |
| Person extracting data | BT ☐ JT ☐ RB ☐ SN ☐ |
| Study number/<br>PMID |  |
| Date of publication |  |
| Author(s) |  |
| Title |  |
| Type of Publication<br>(e.g. journal article, dissertation) |  |
| Location of Study |  |
| **Study Details** |  |
| Study Design |  |
| Aims of Study |  |
| Population |  |
| Concept |  |
| Context |  |
| Results of Study |  |

**Table 6:**
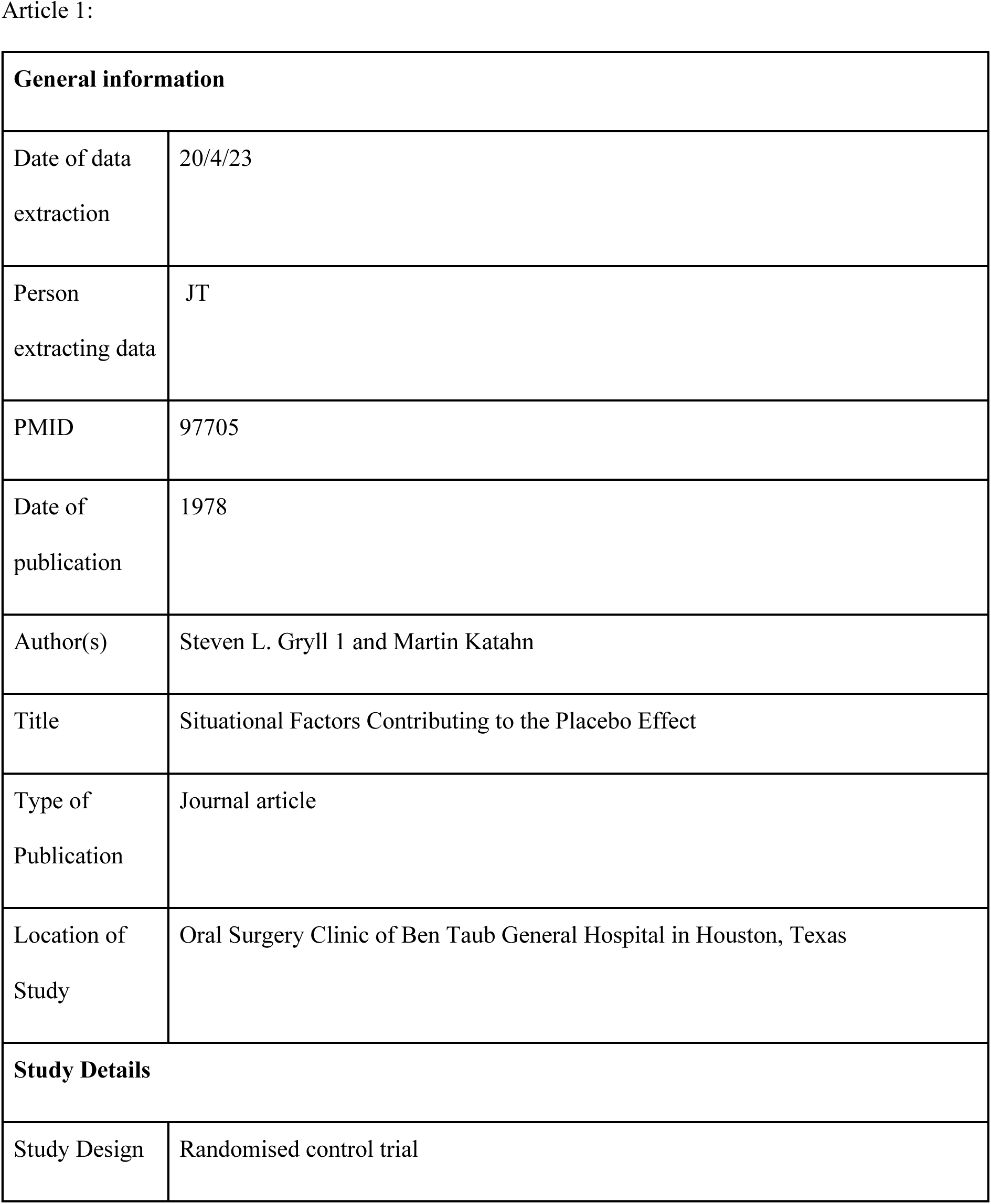
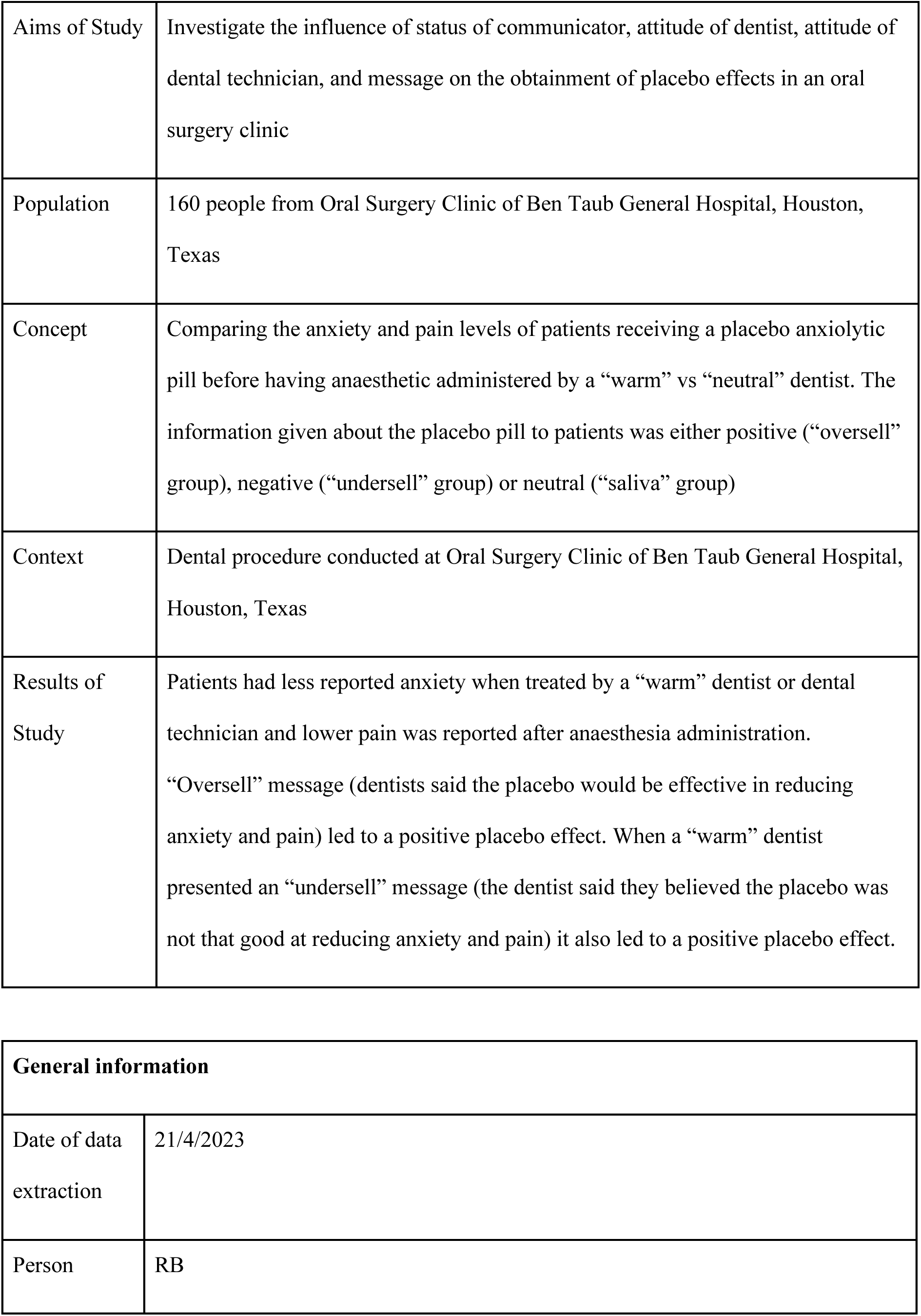
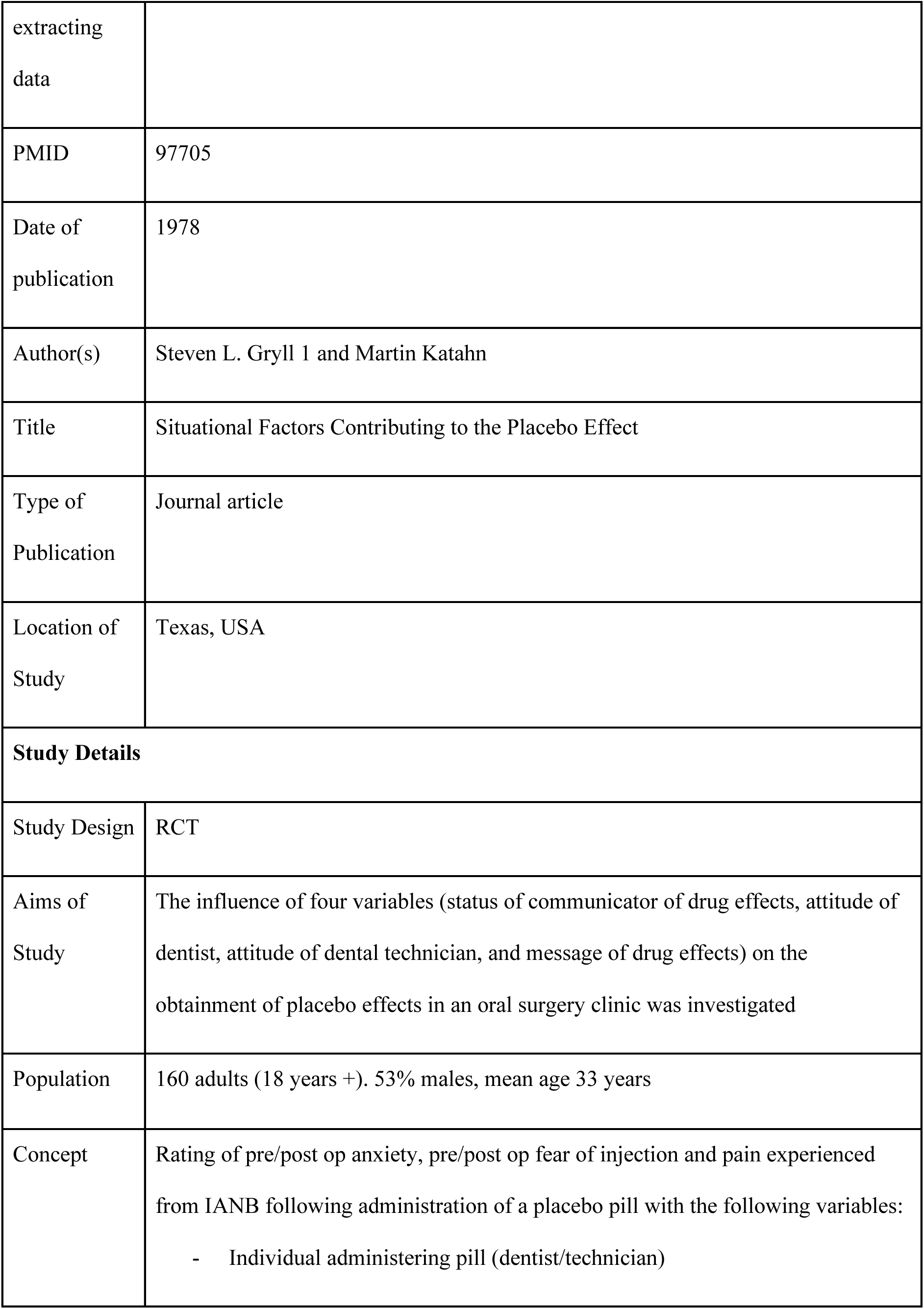
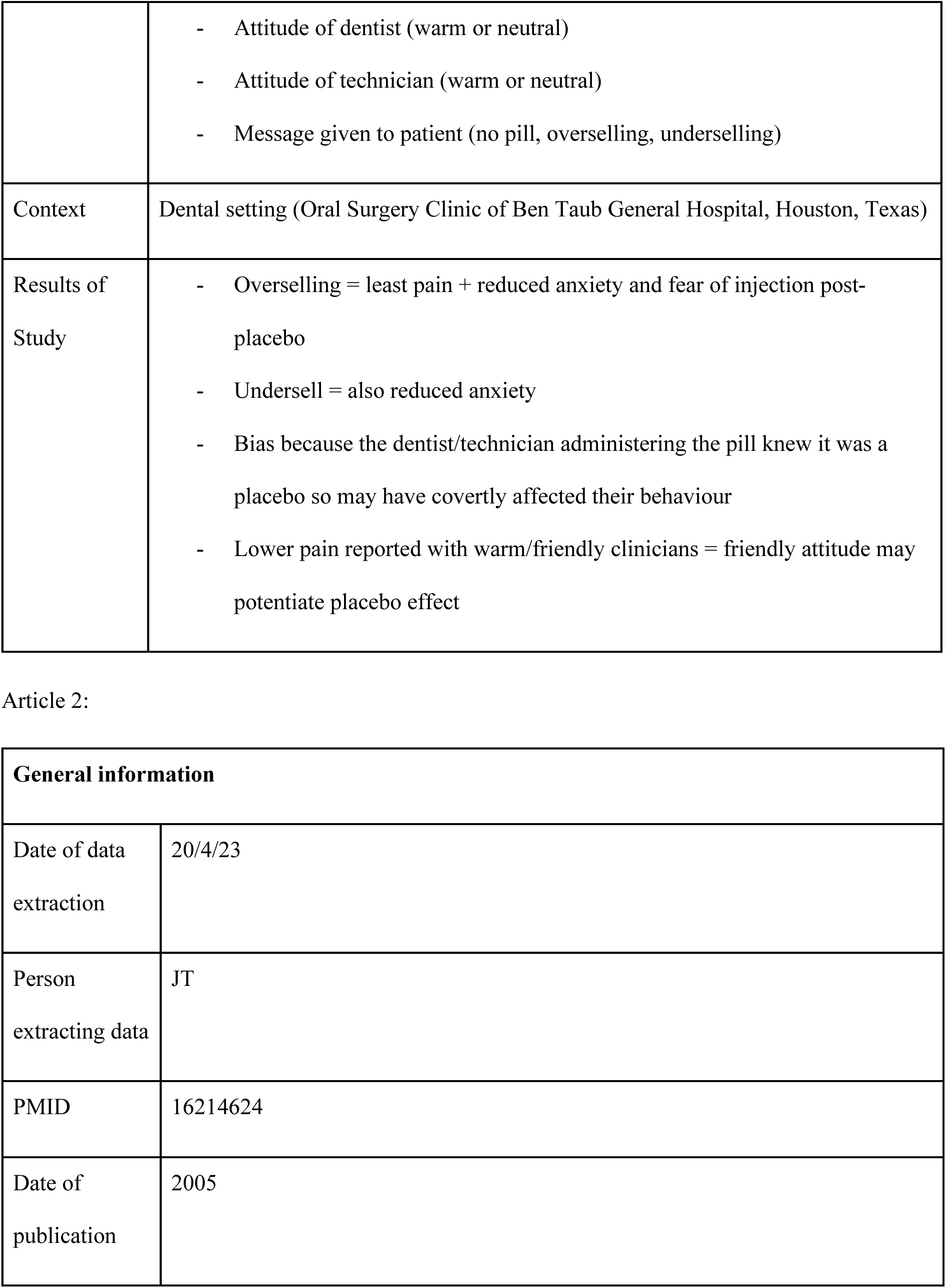
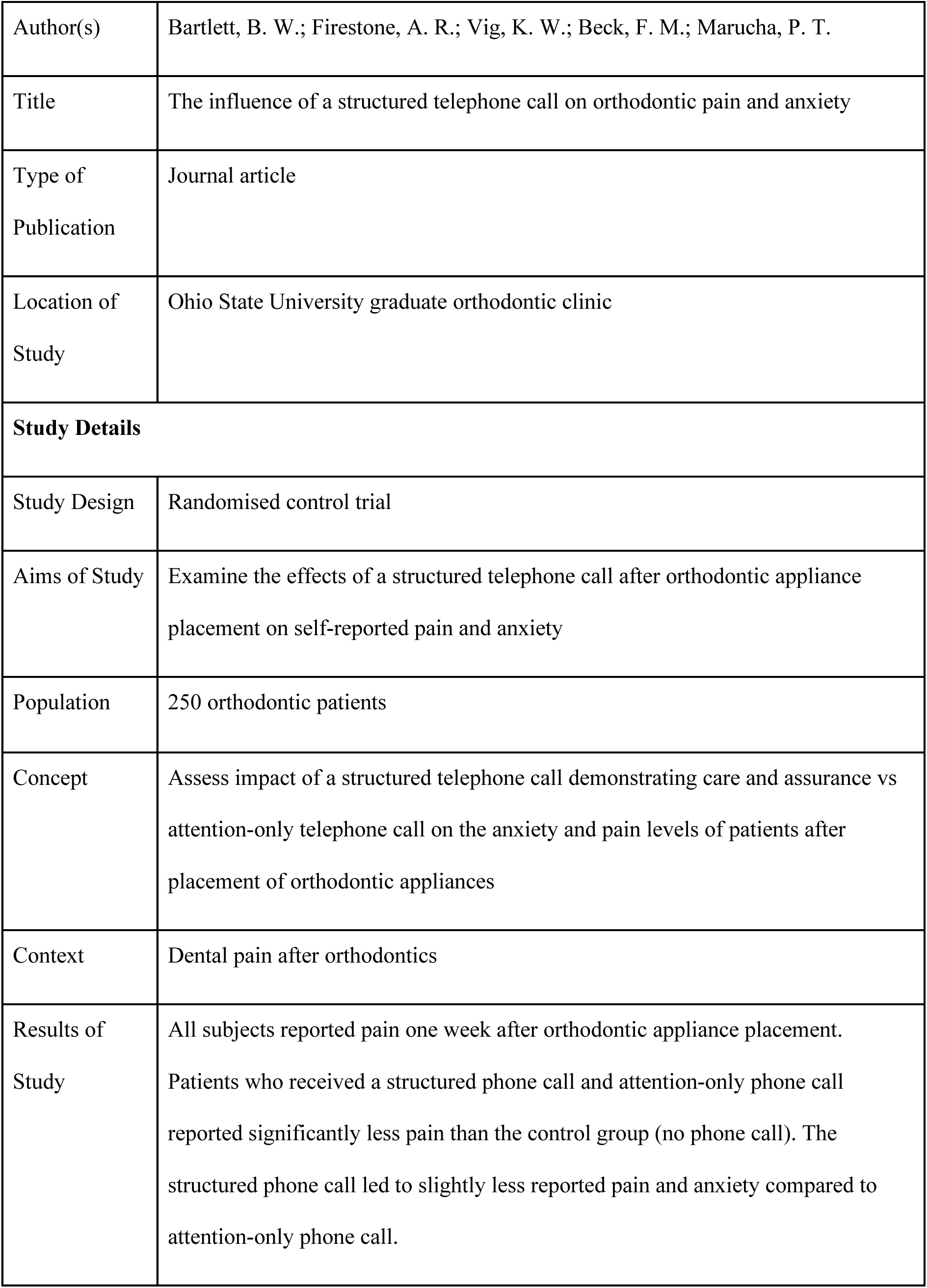
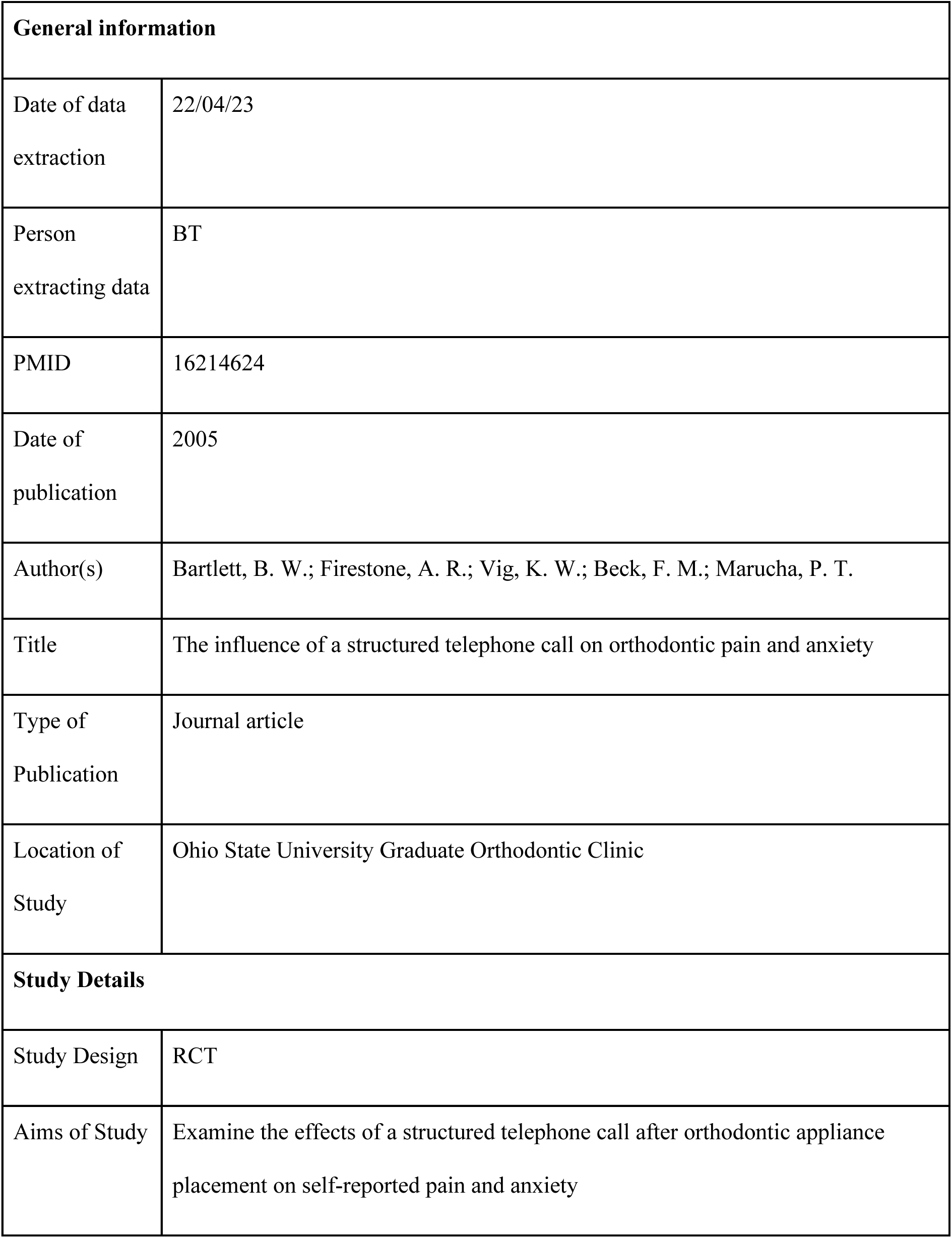
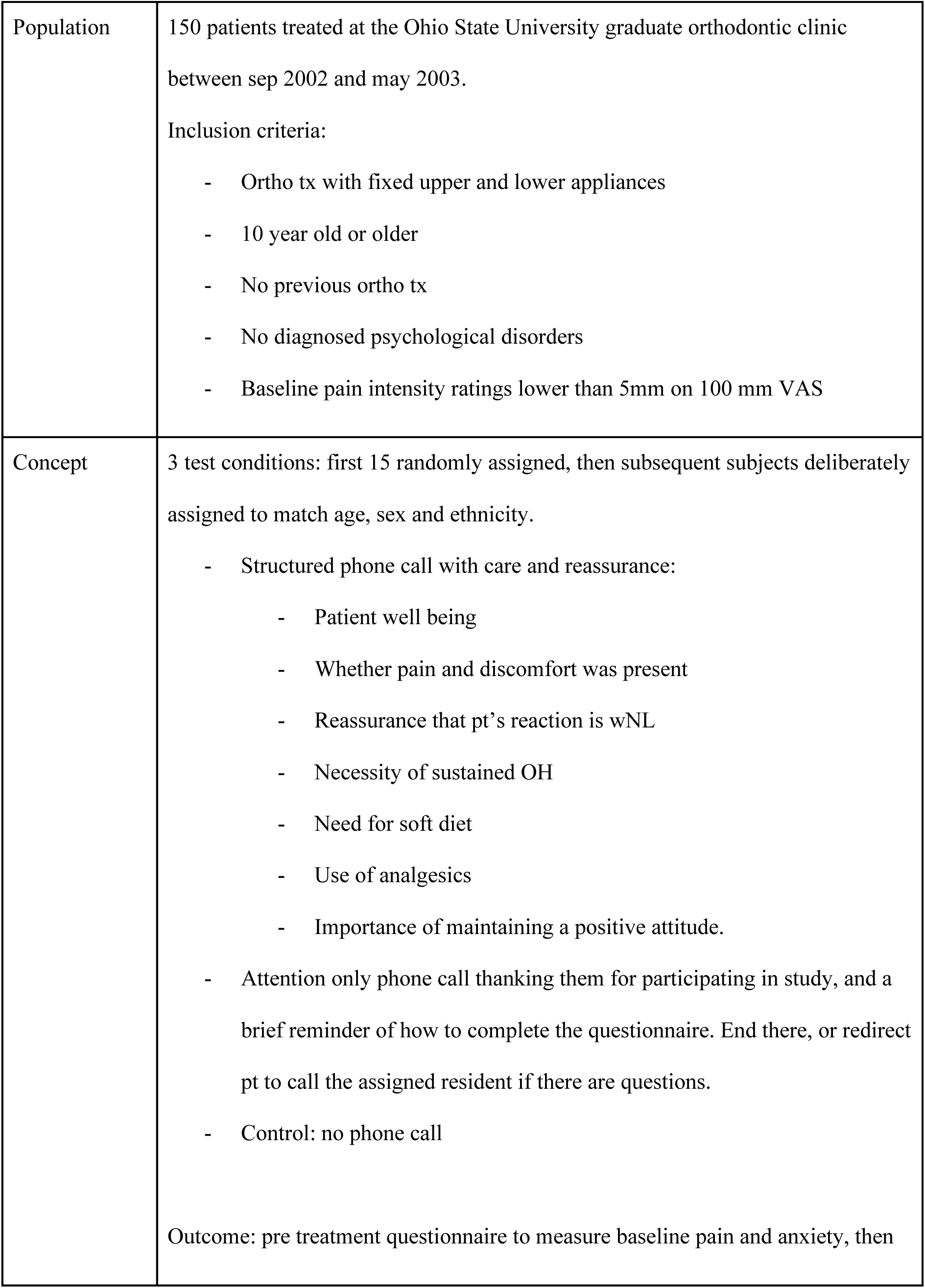
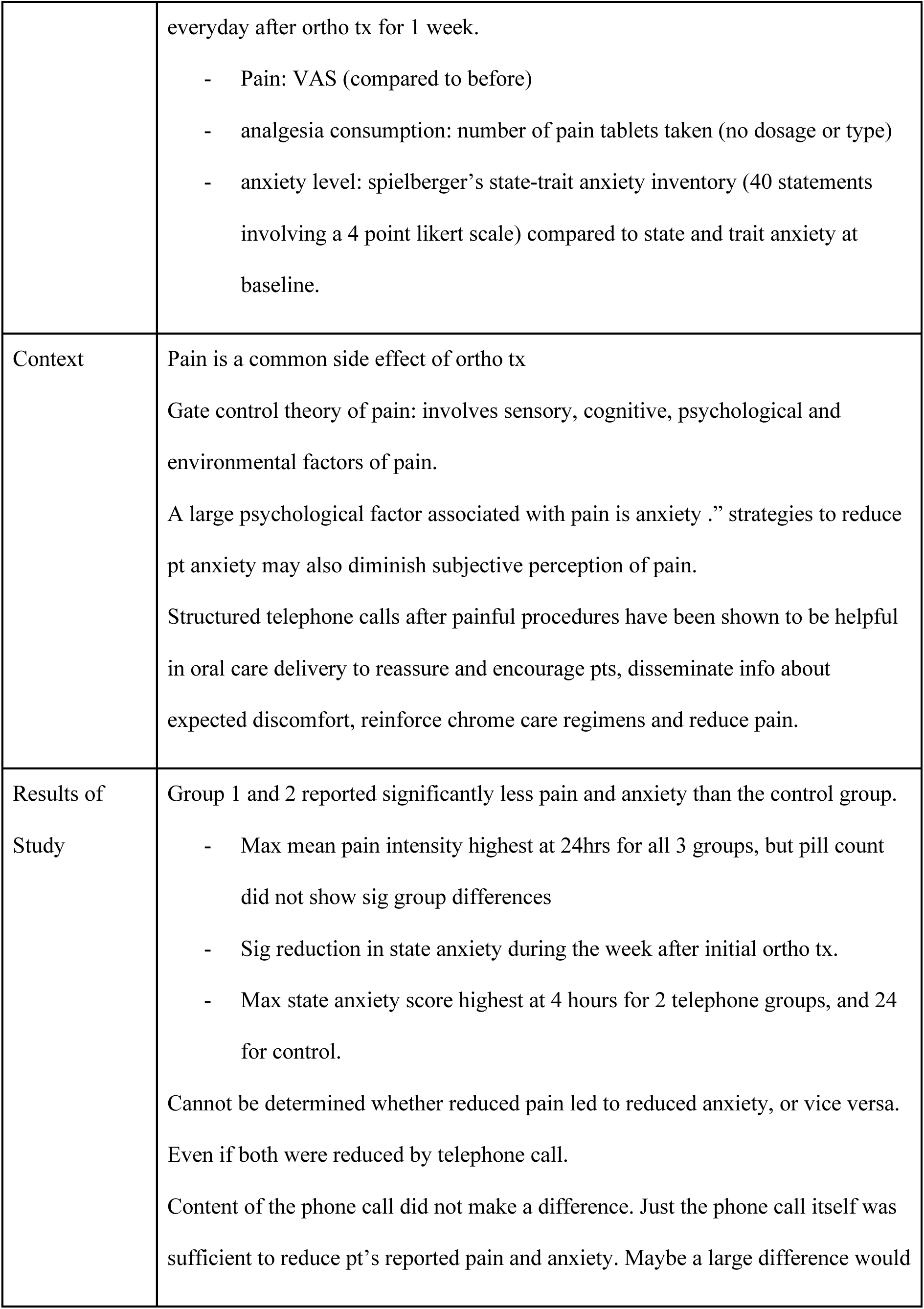
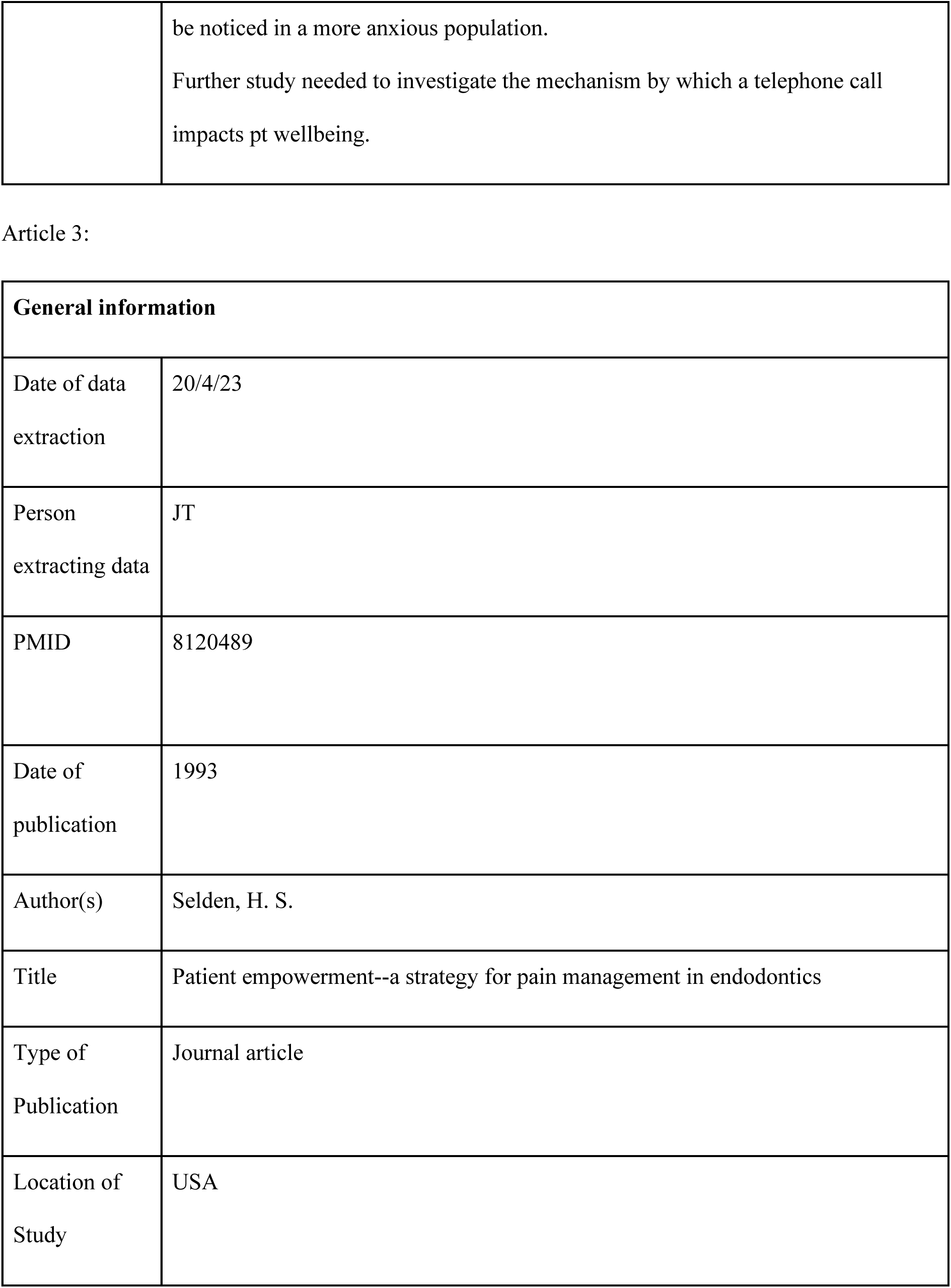
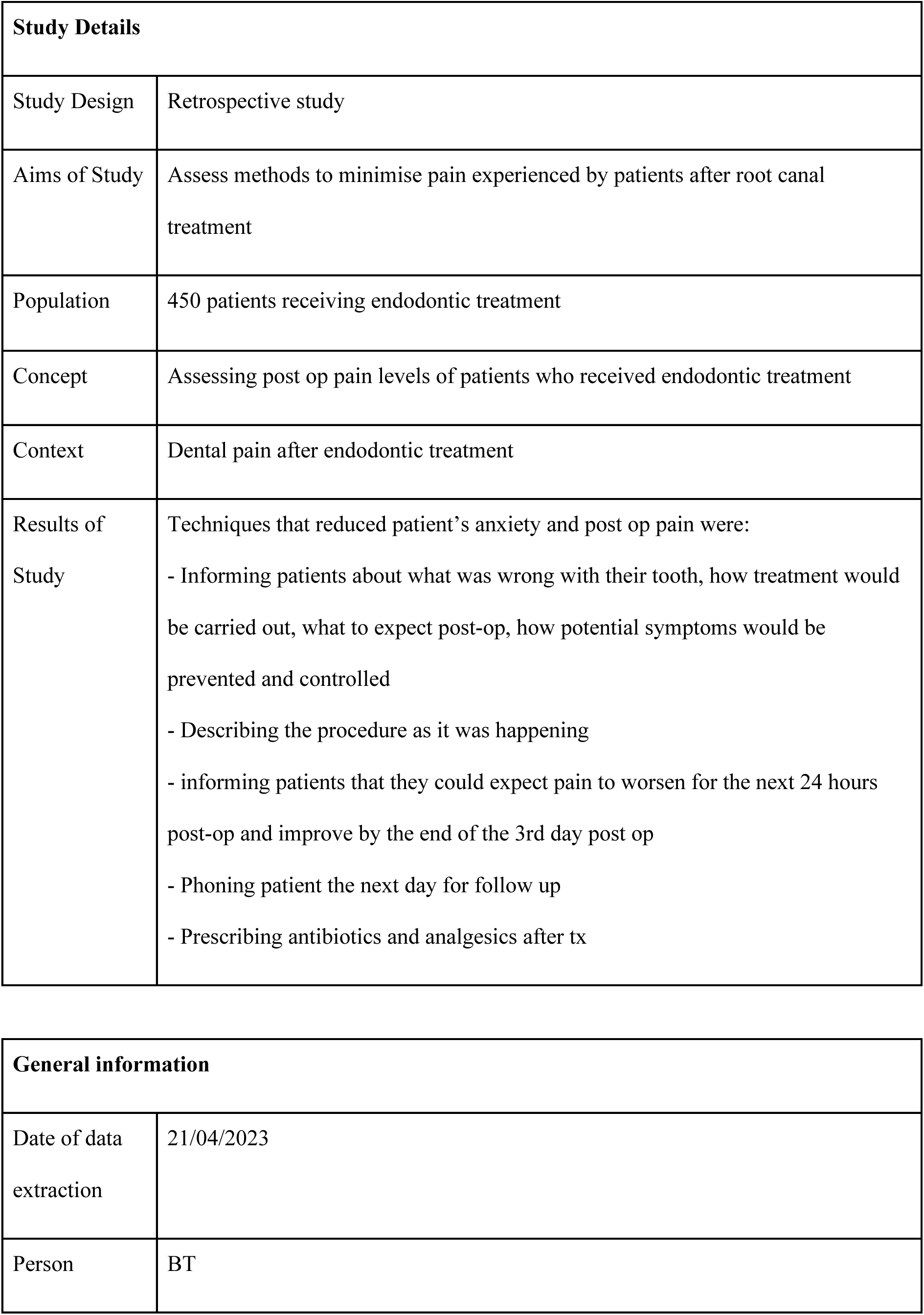
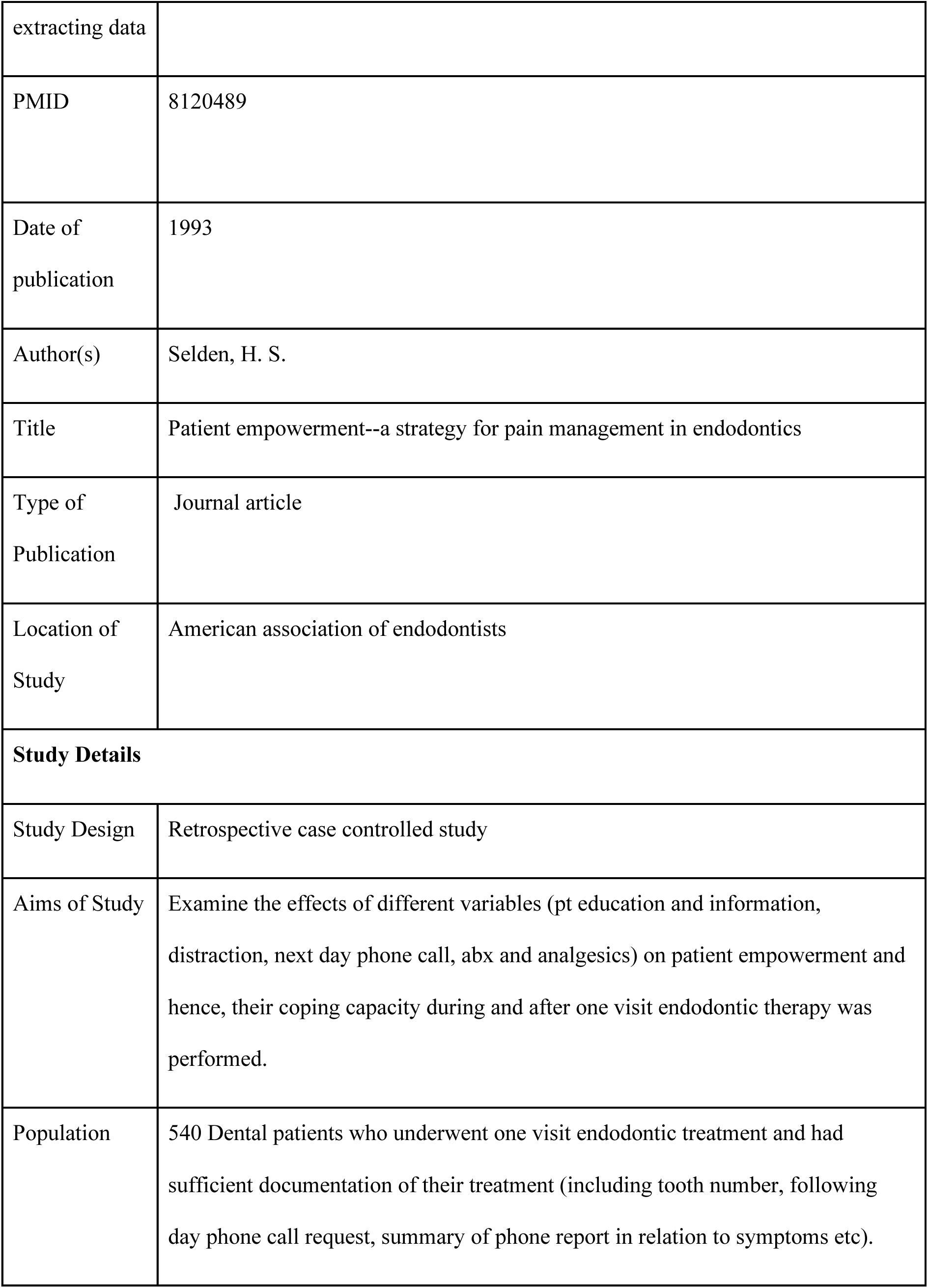
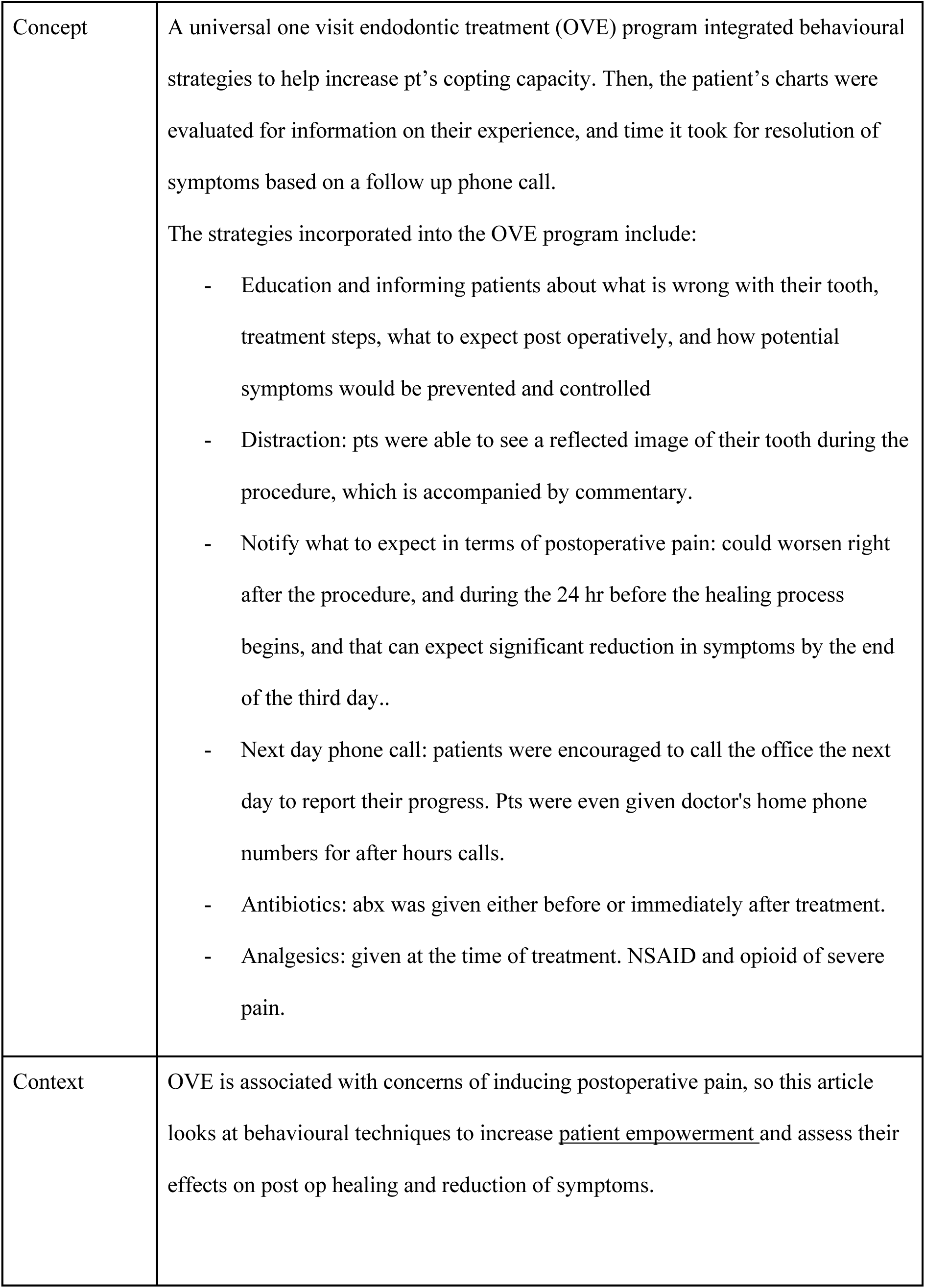
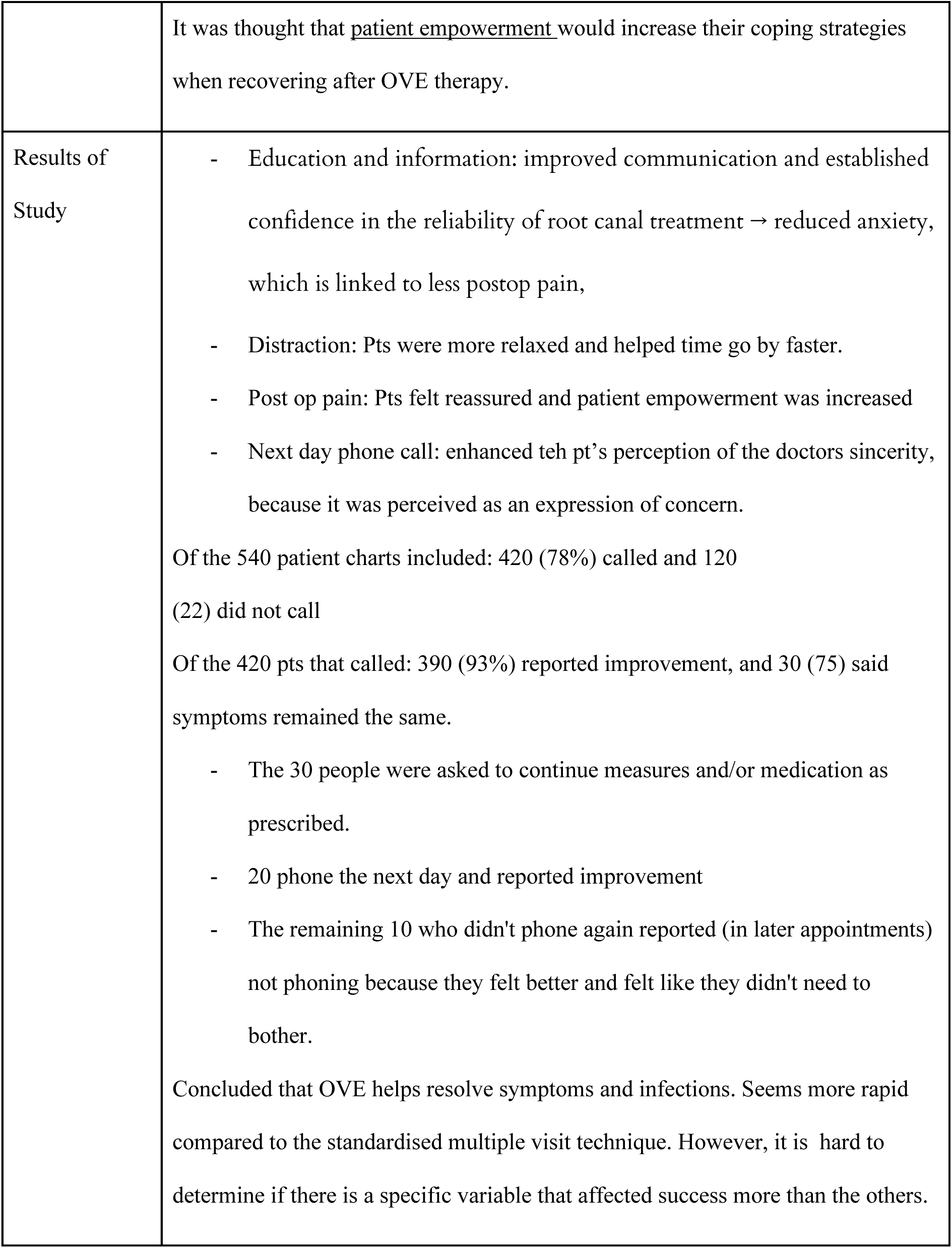
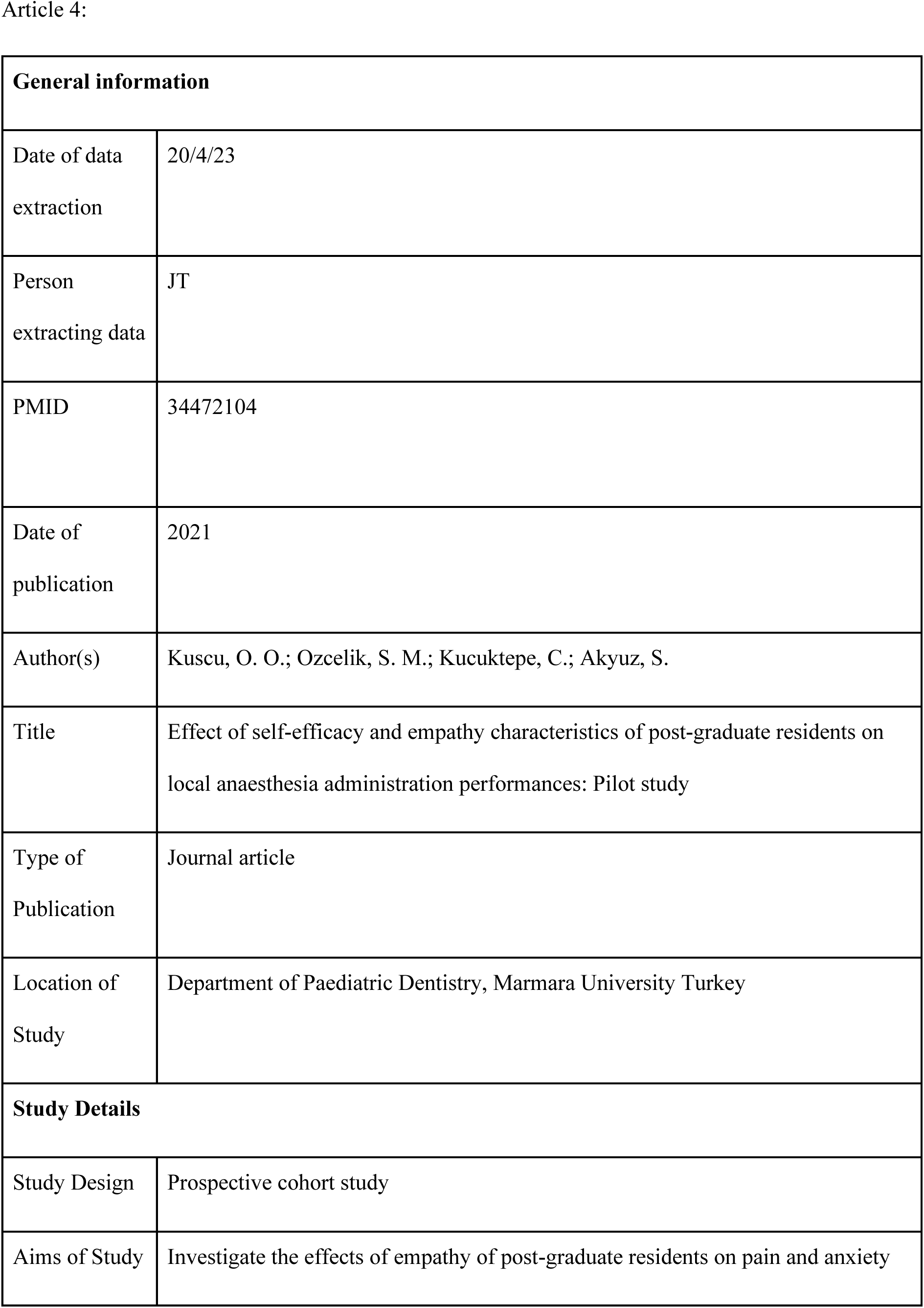
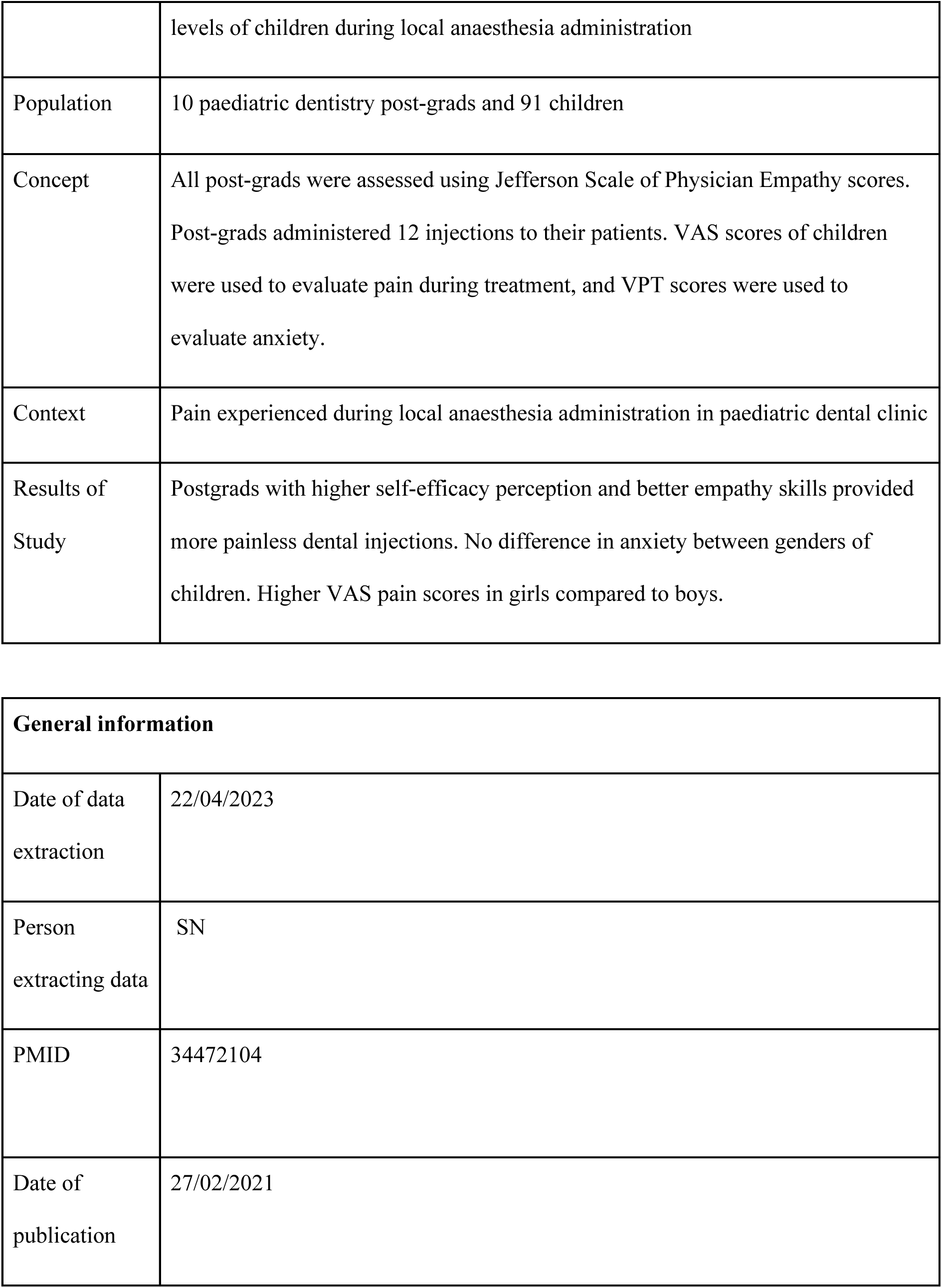
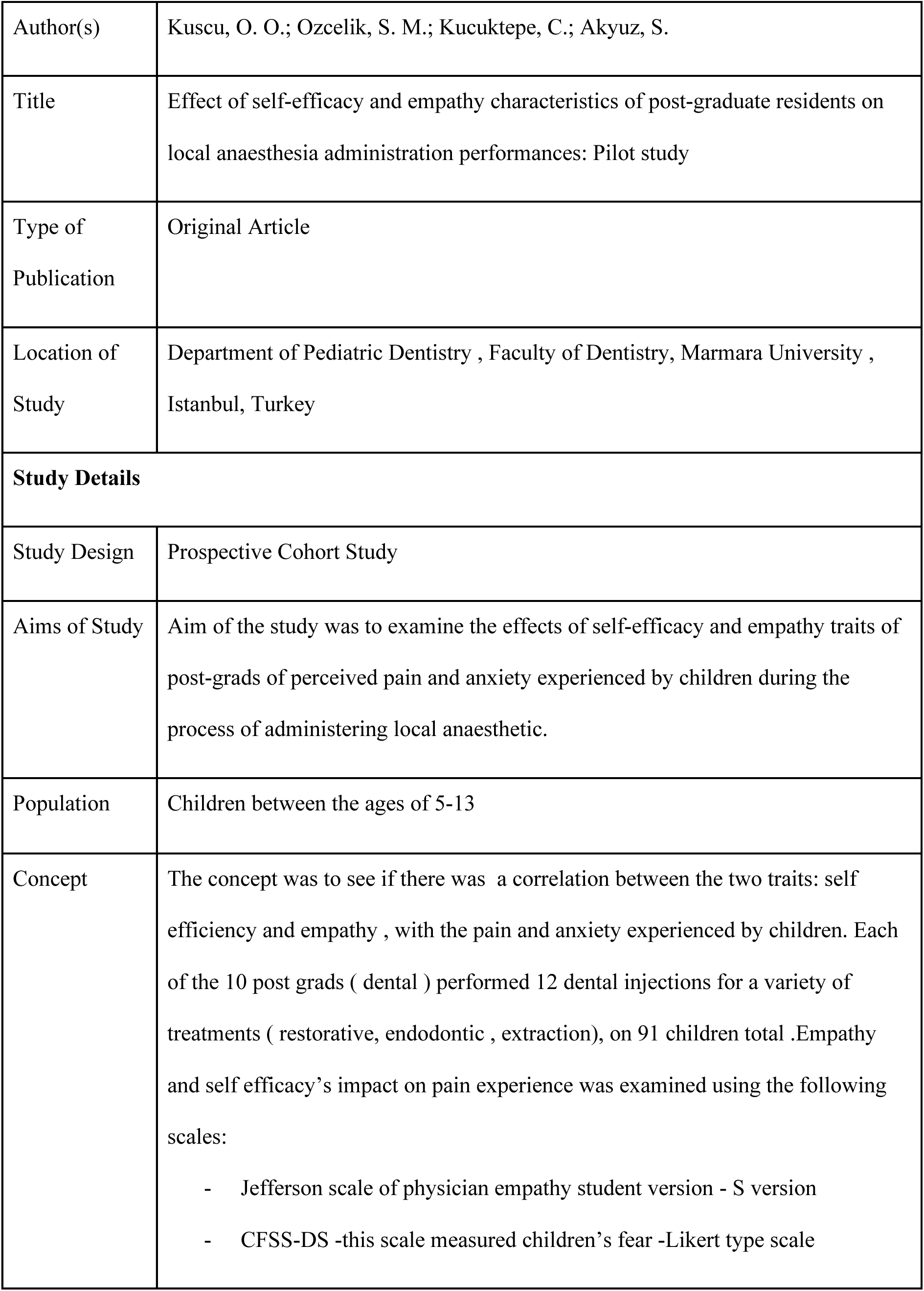
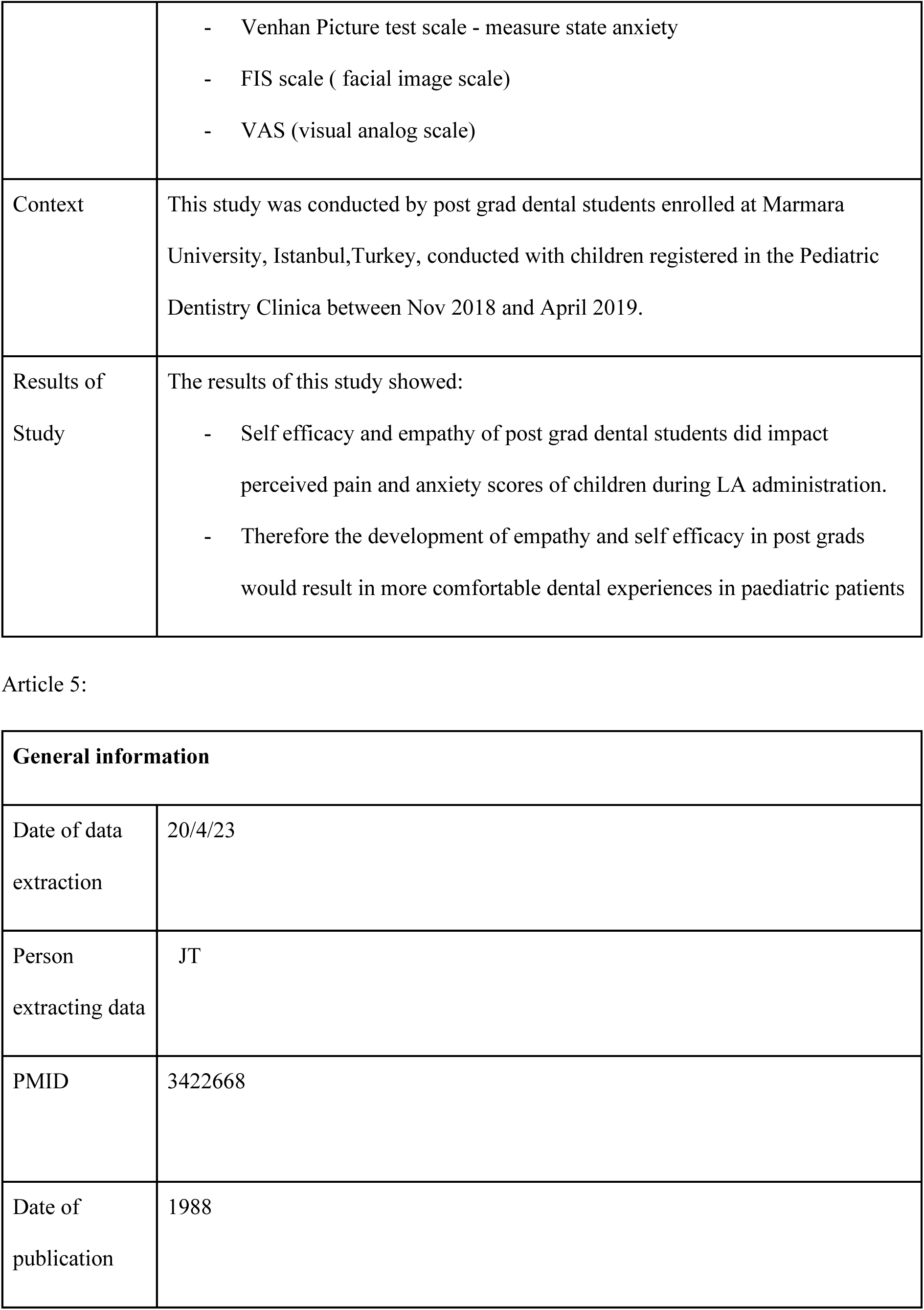
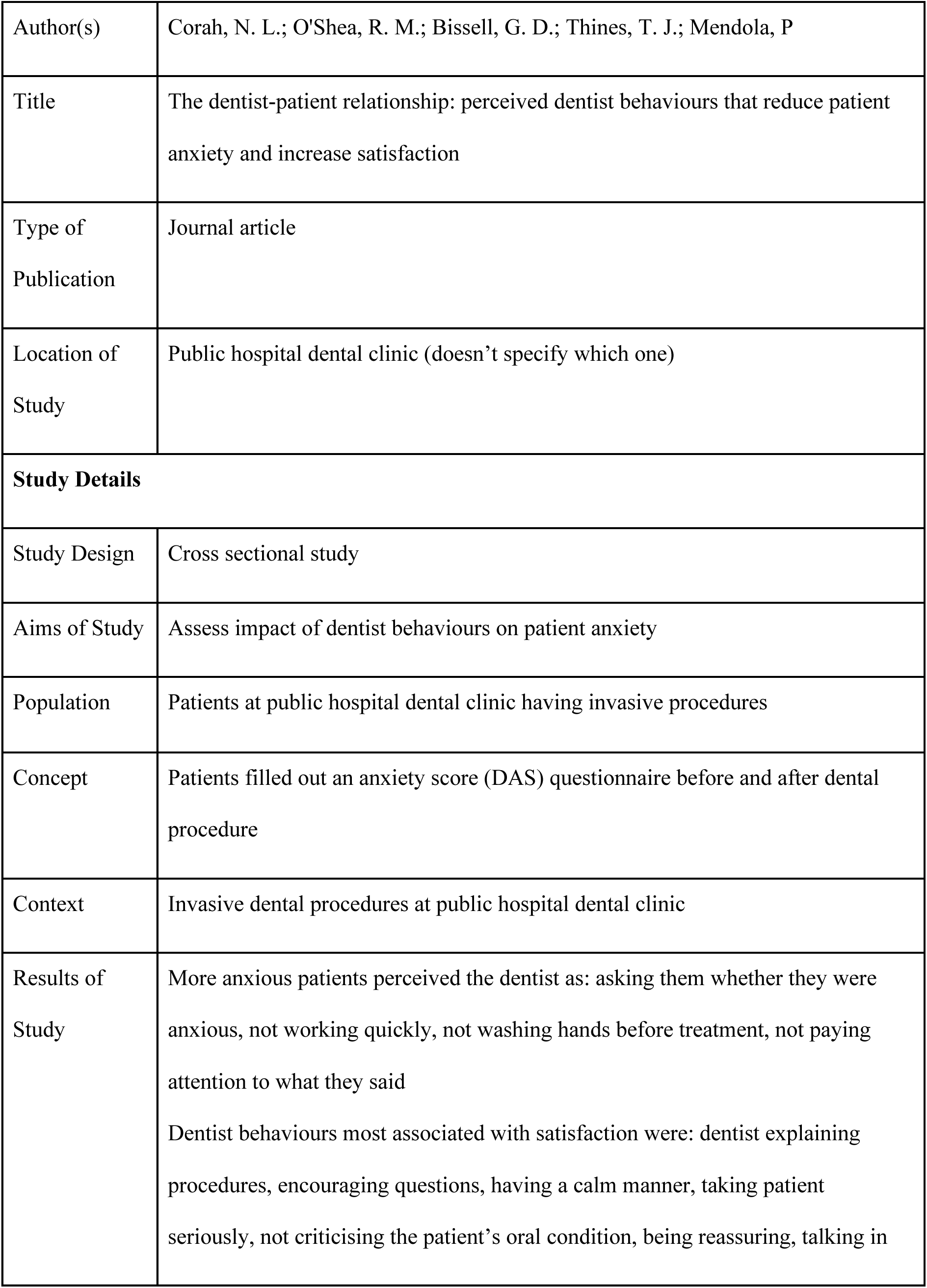
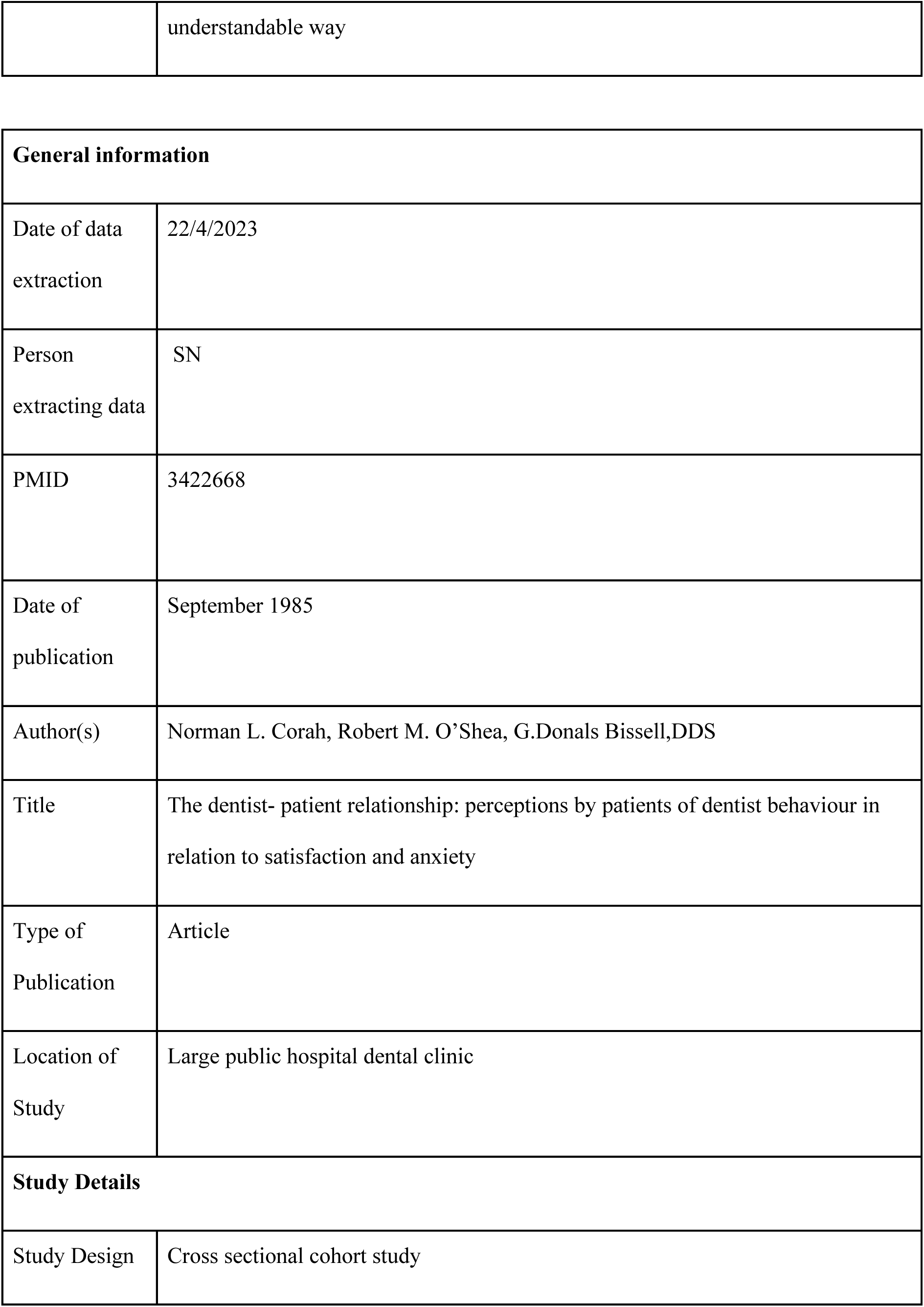
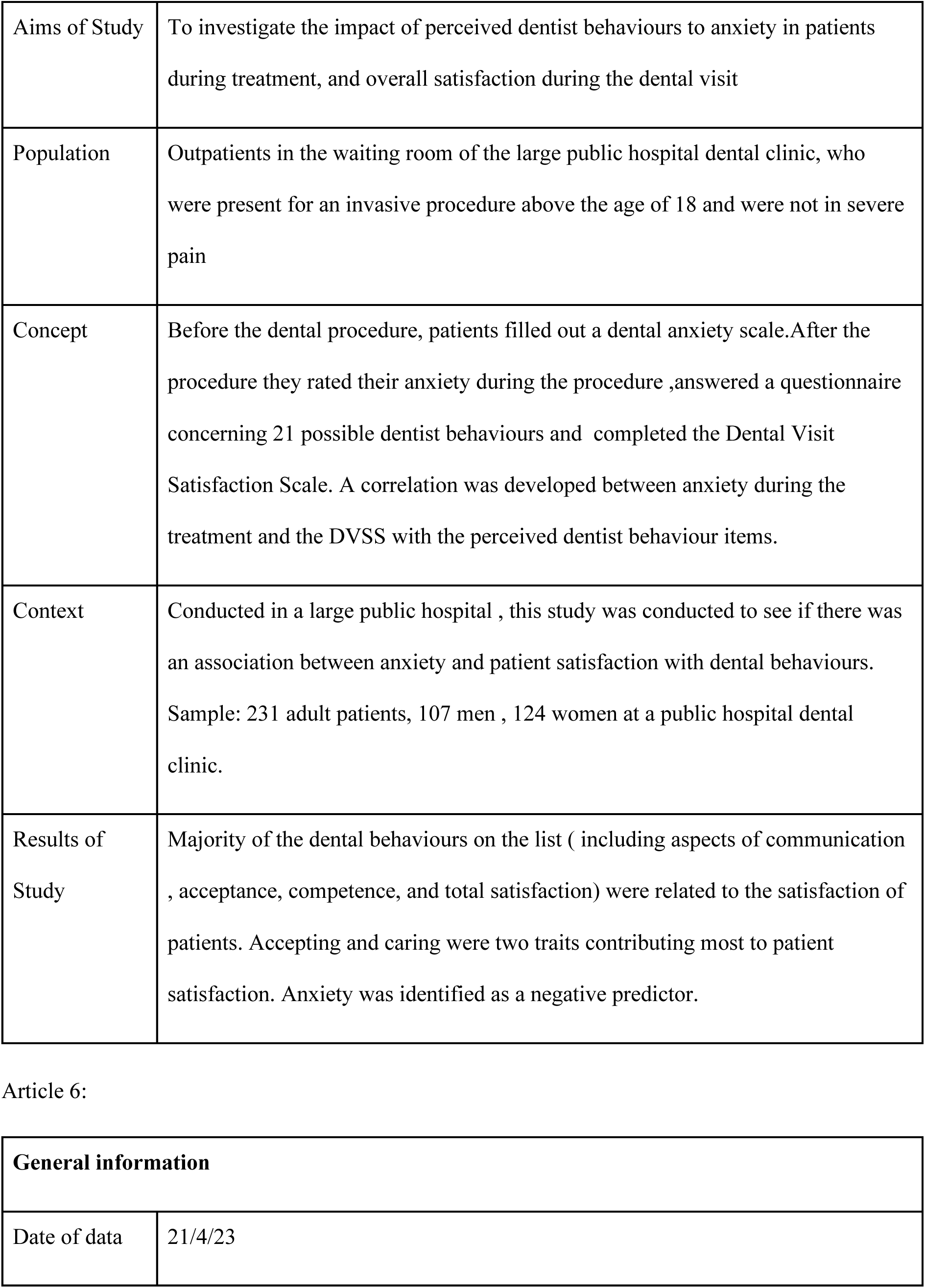
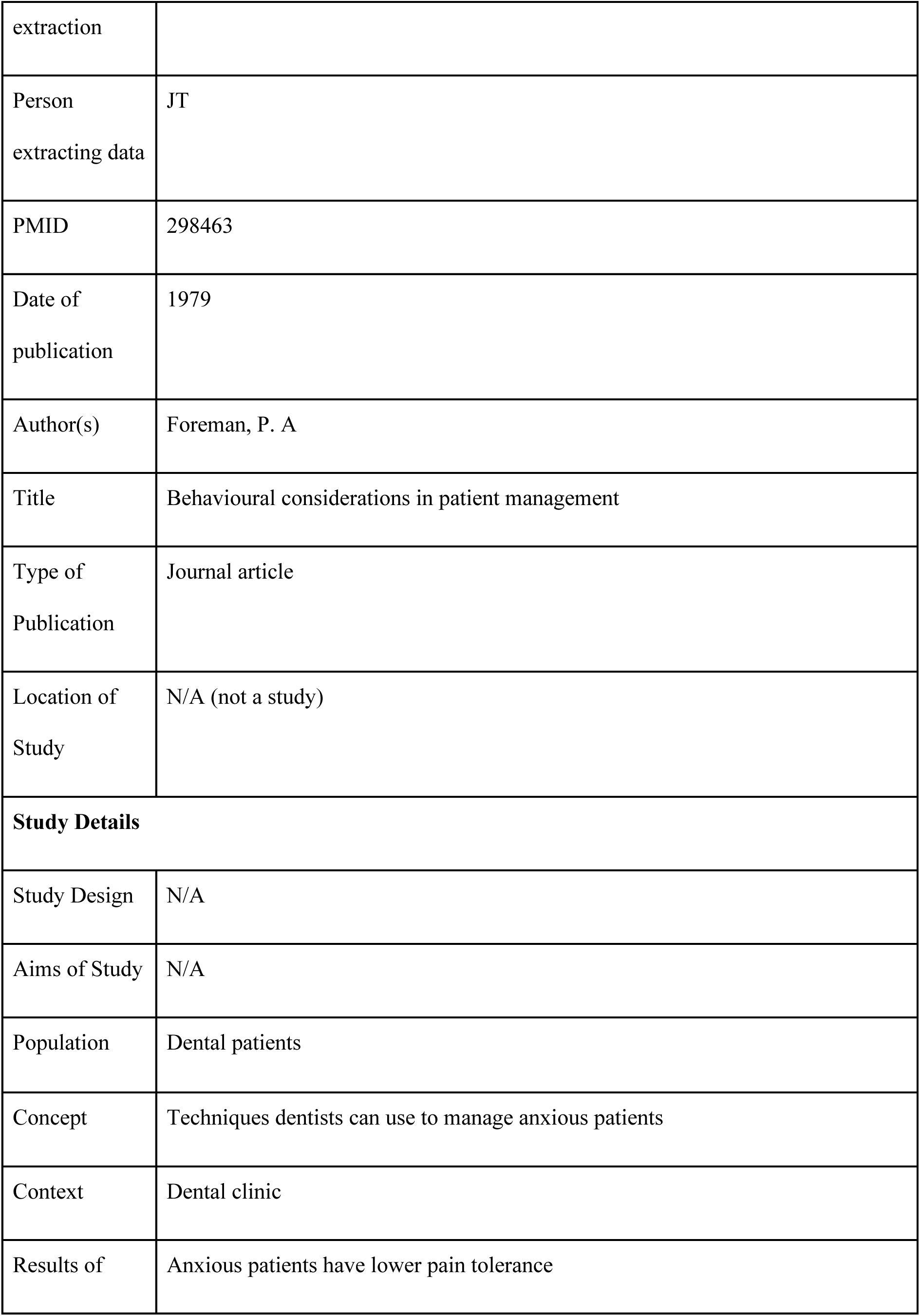
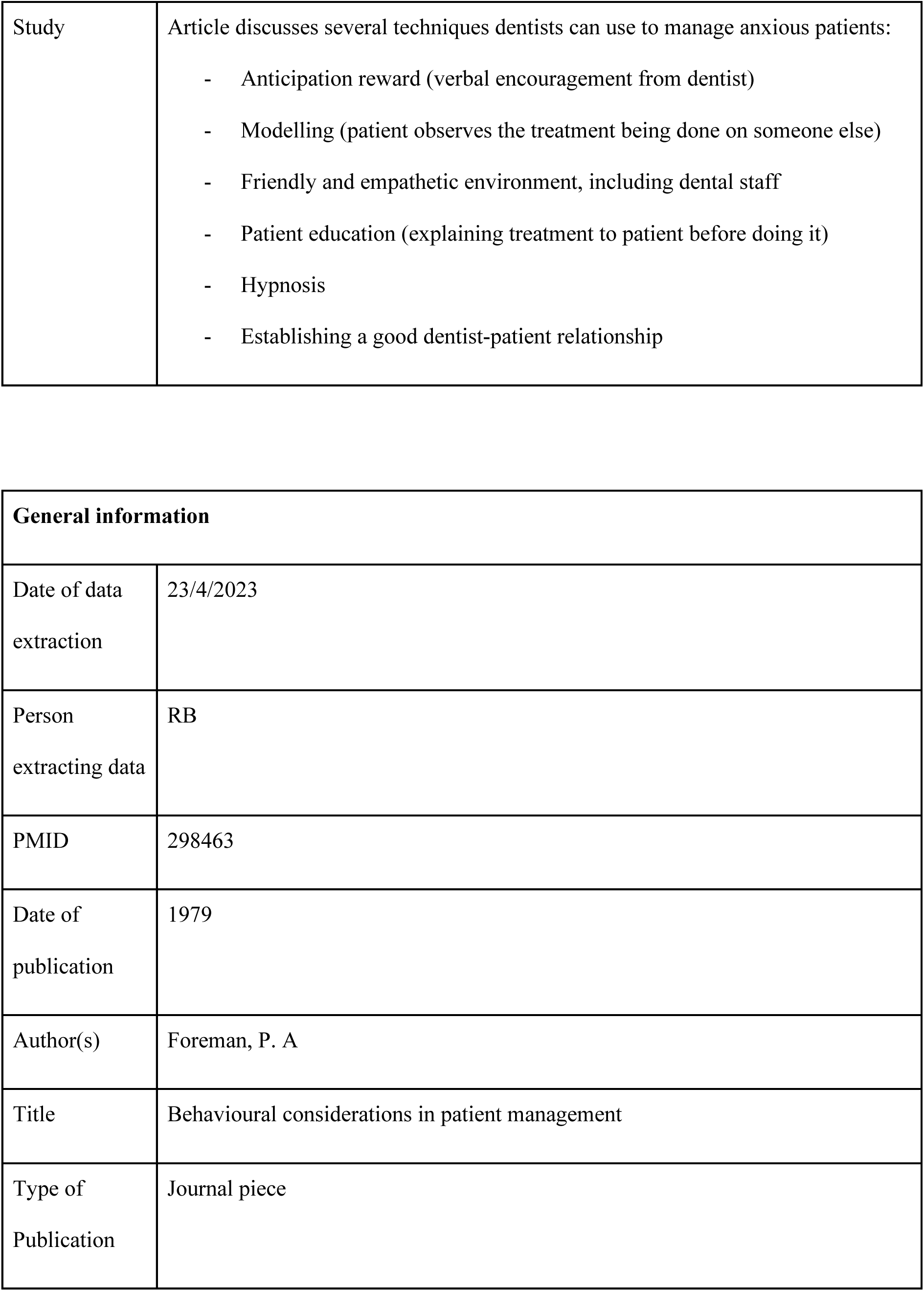
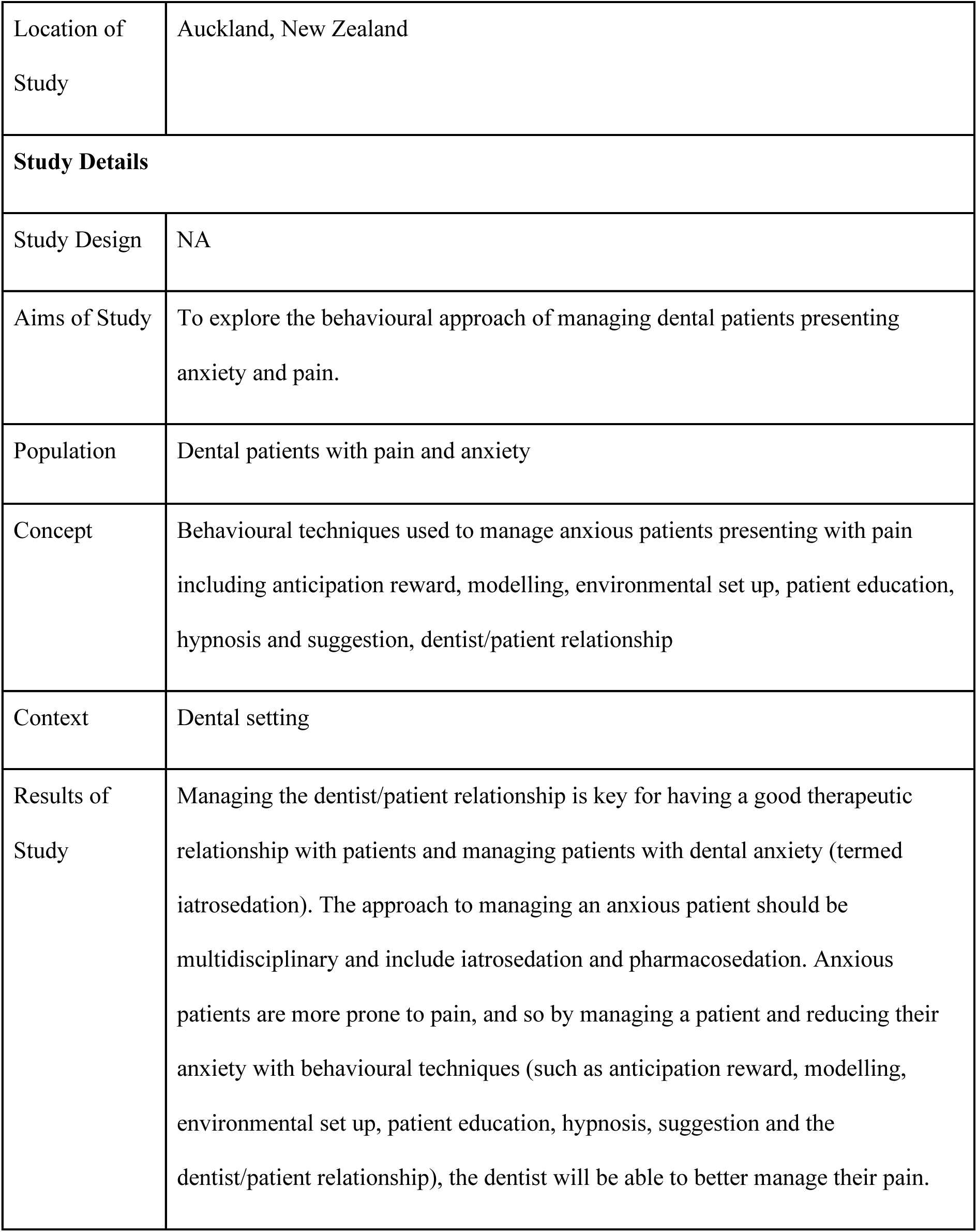
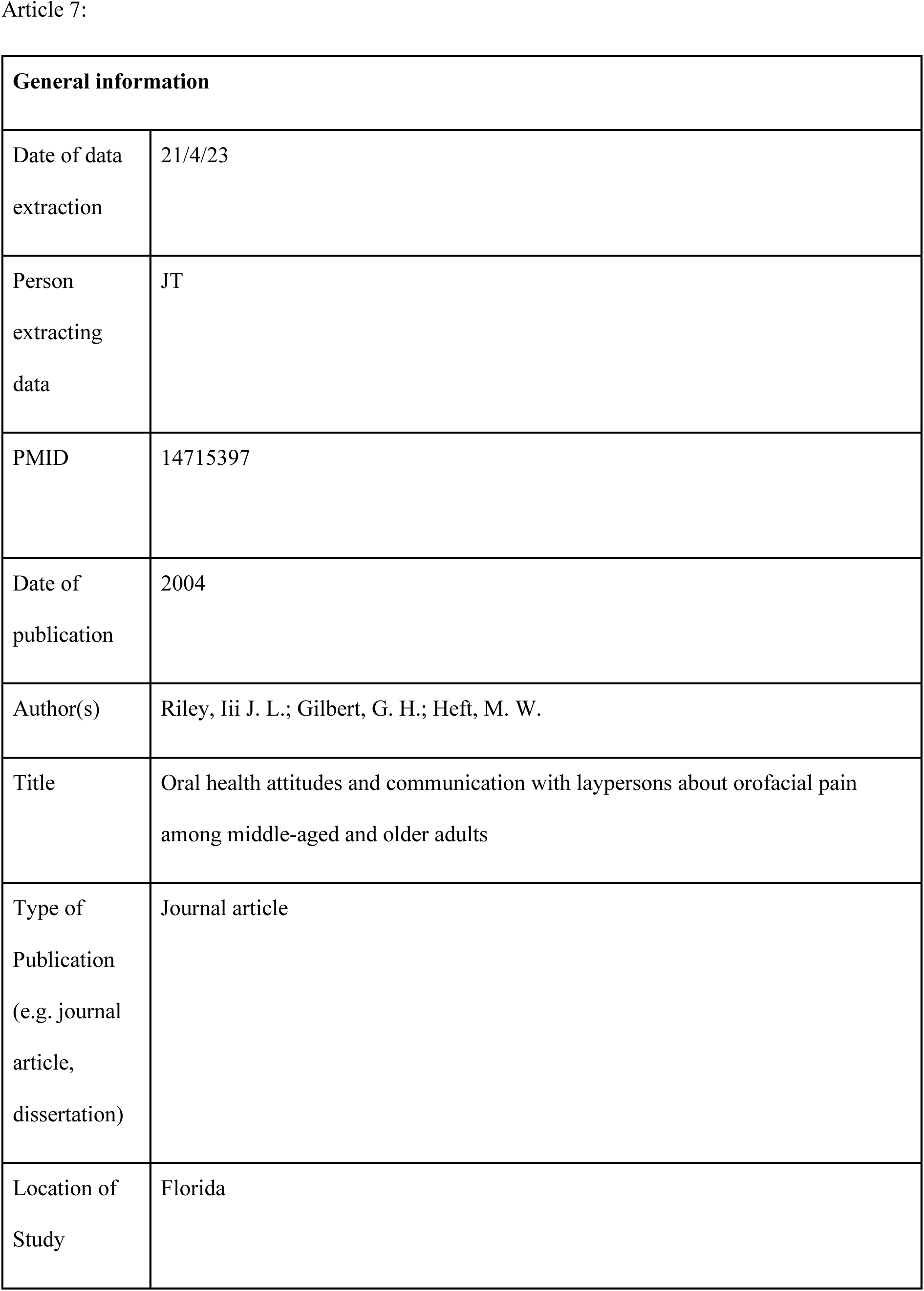
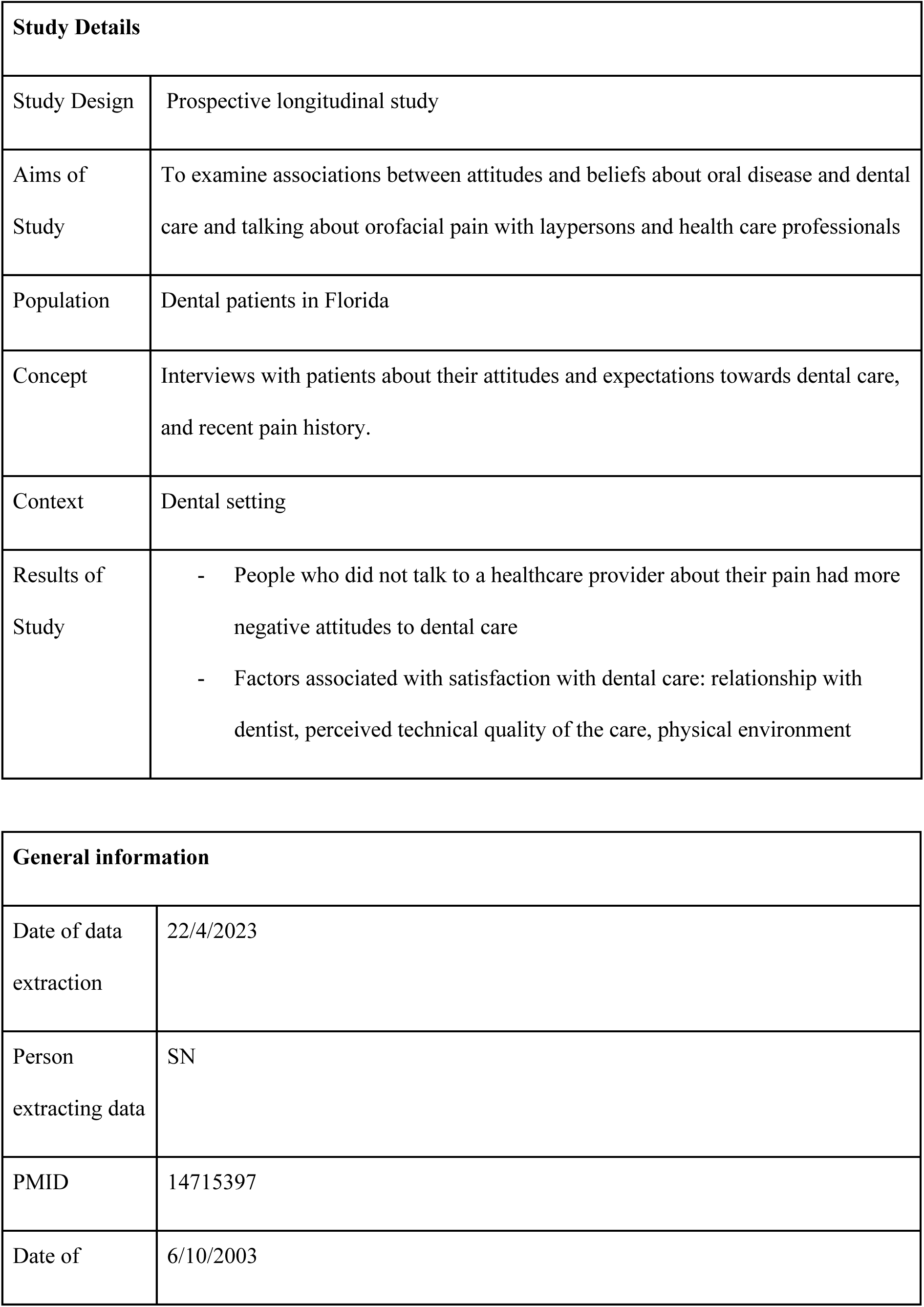
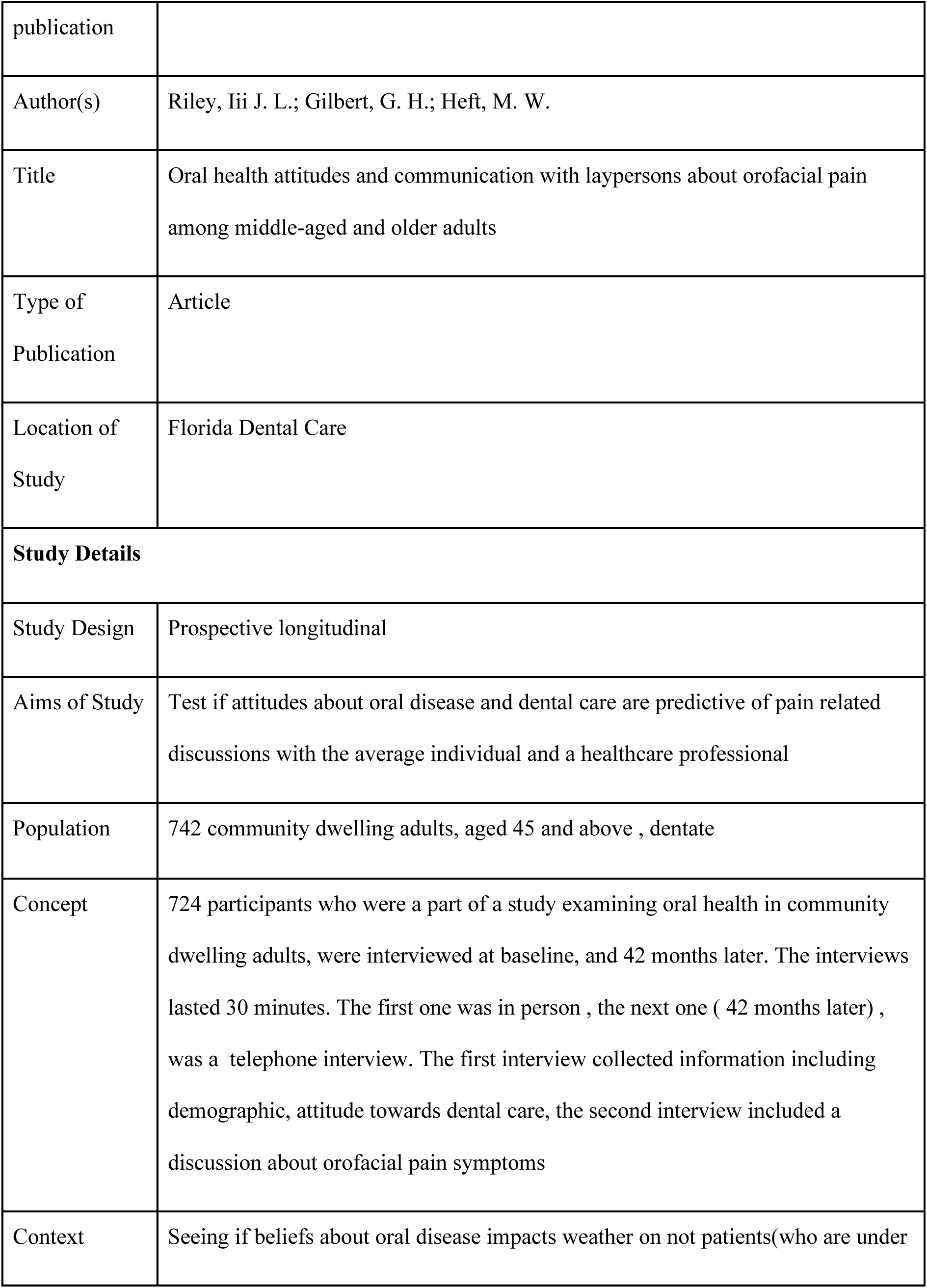
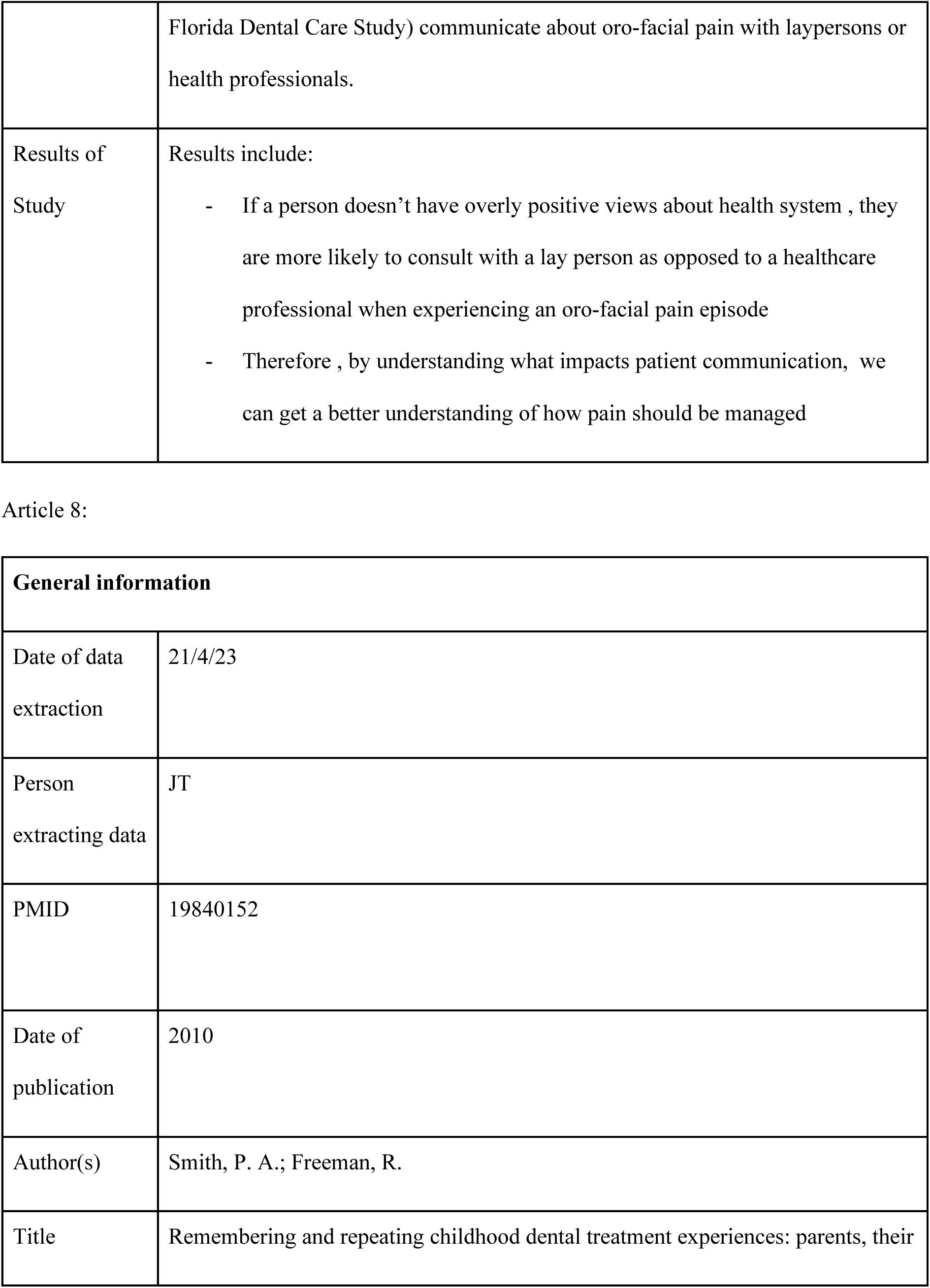
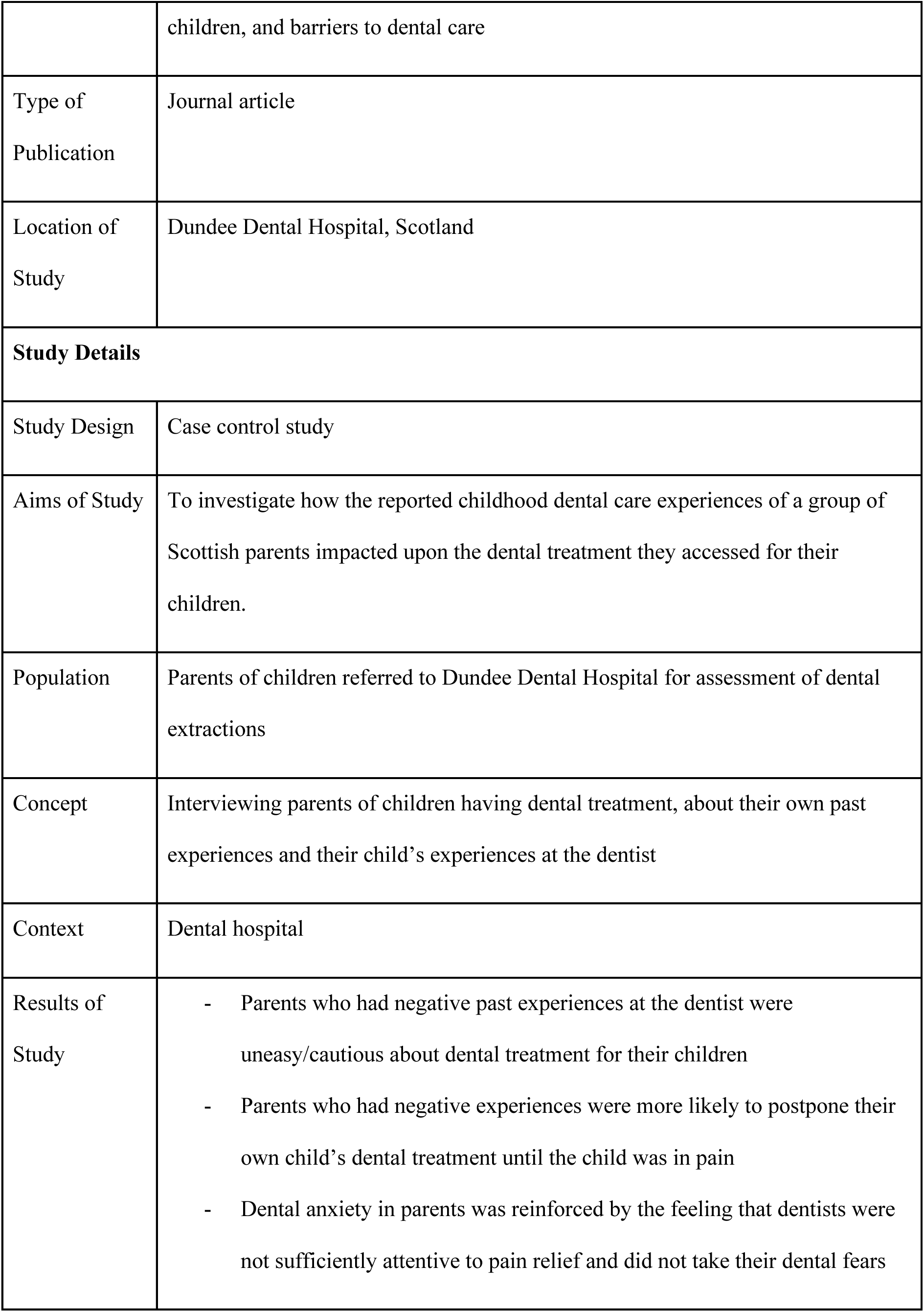
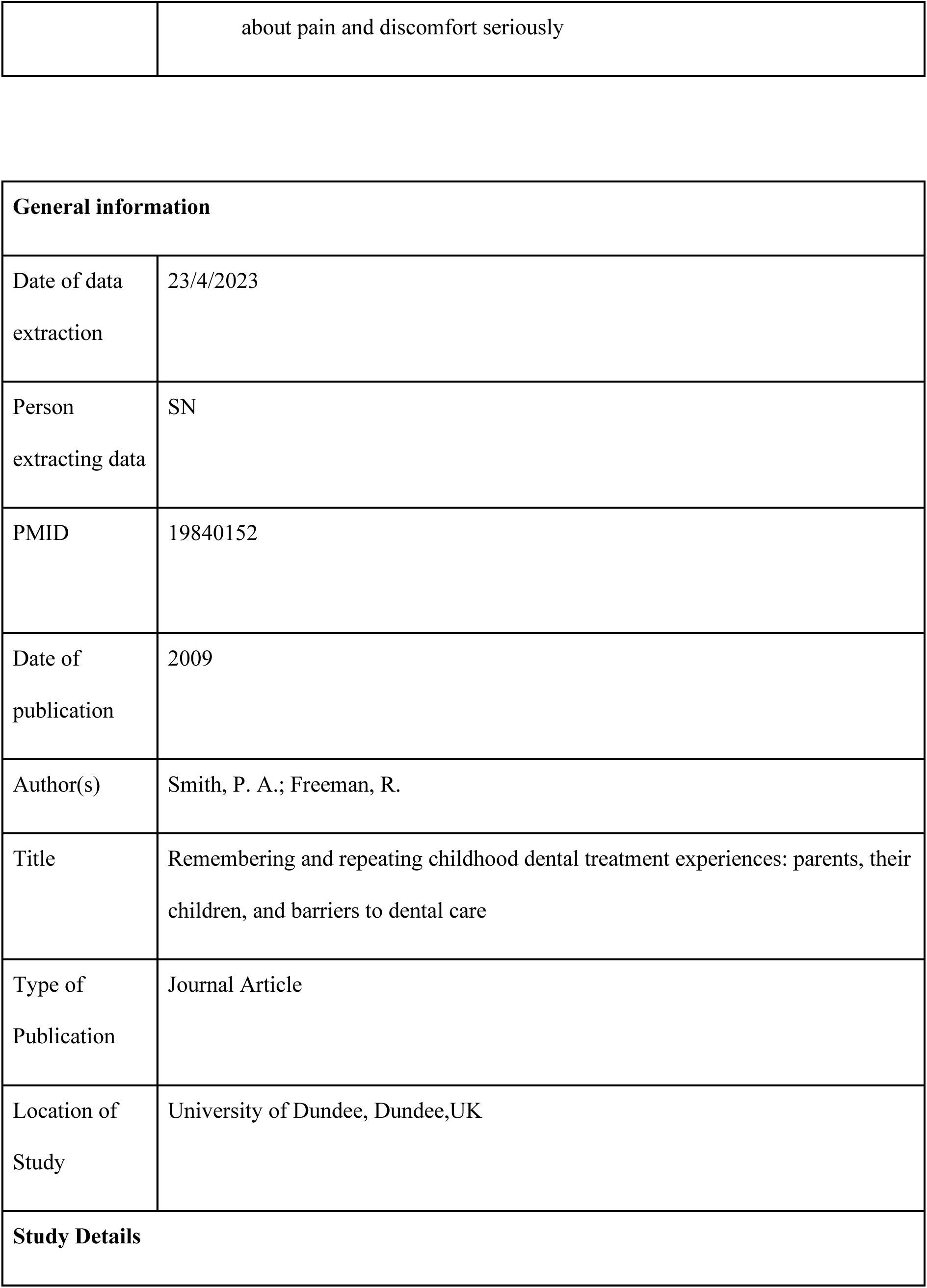
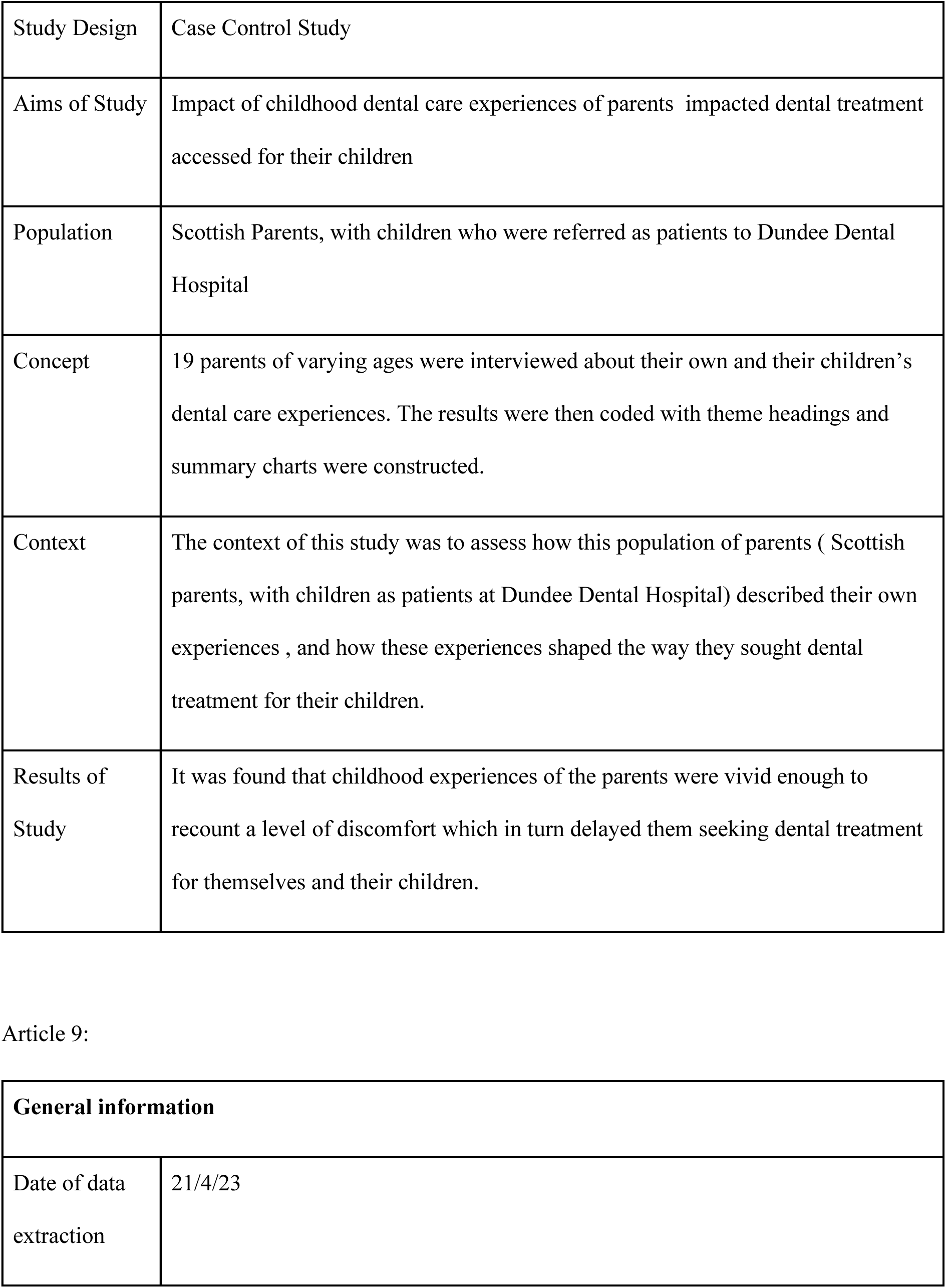
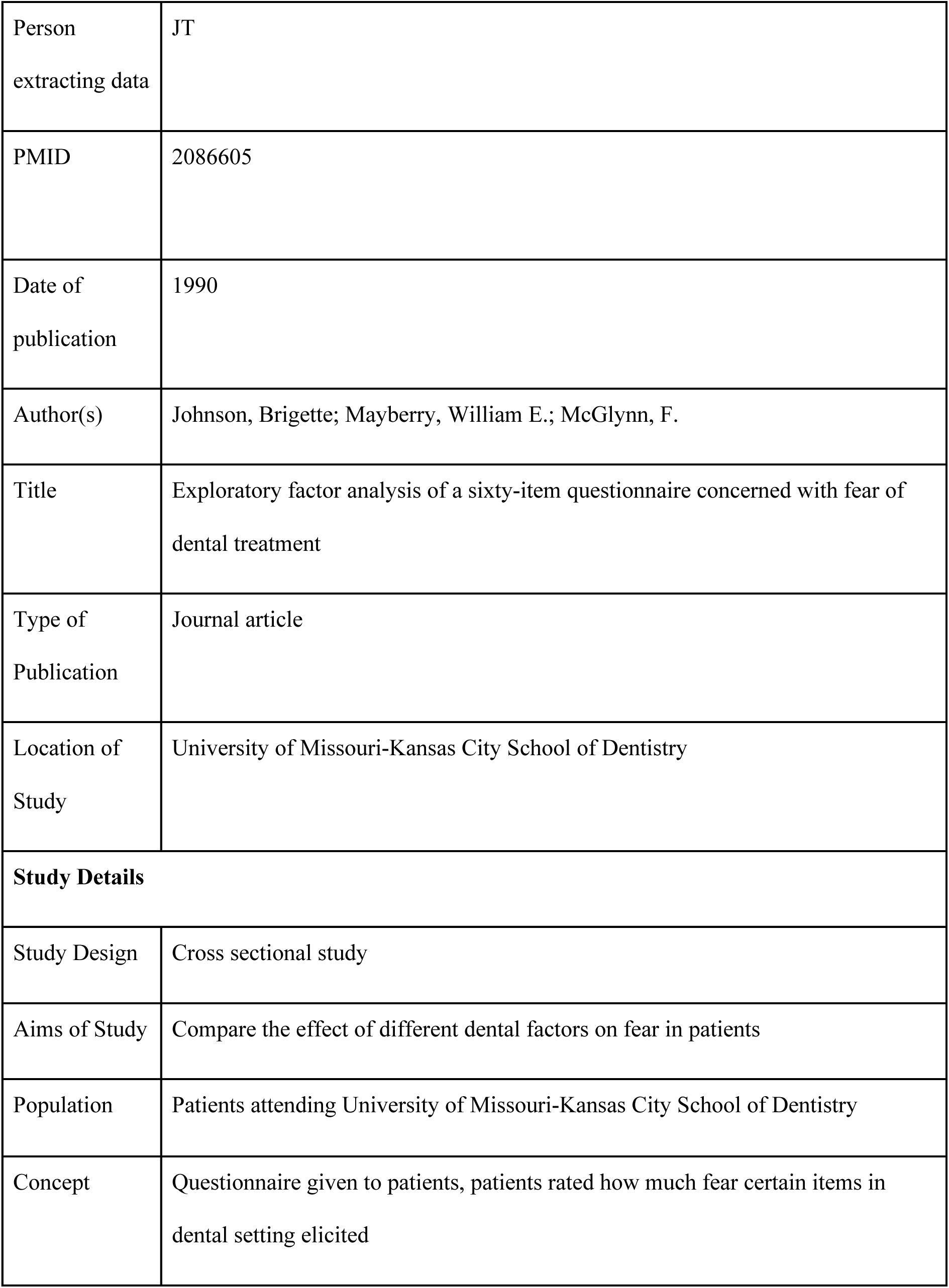
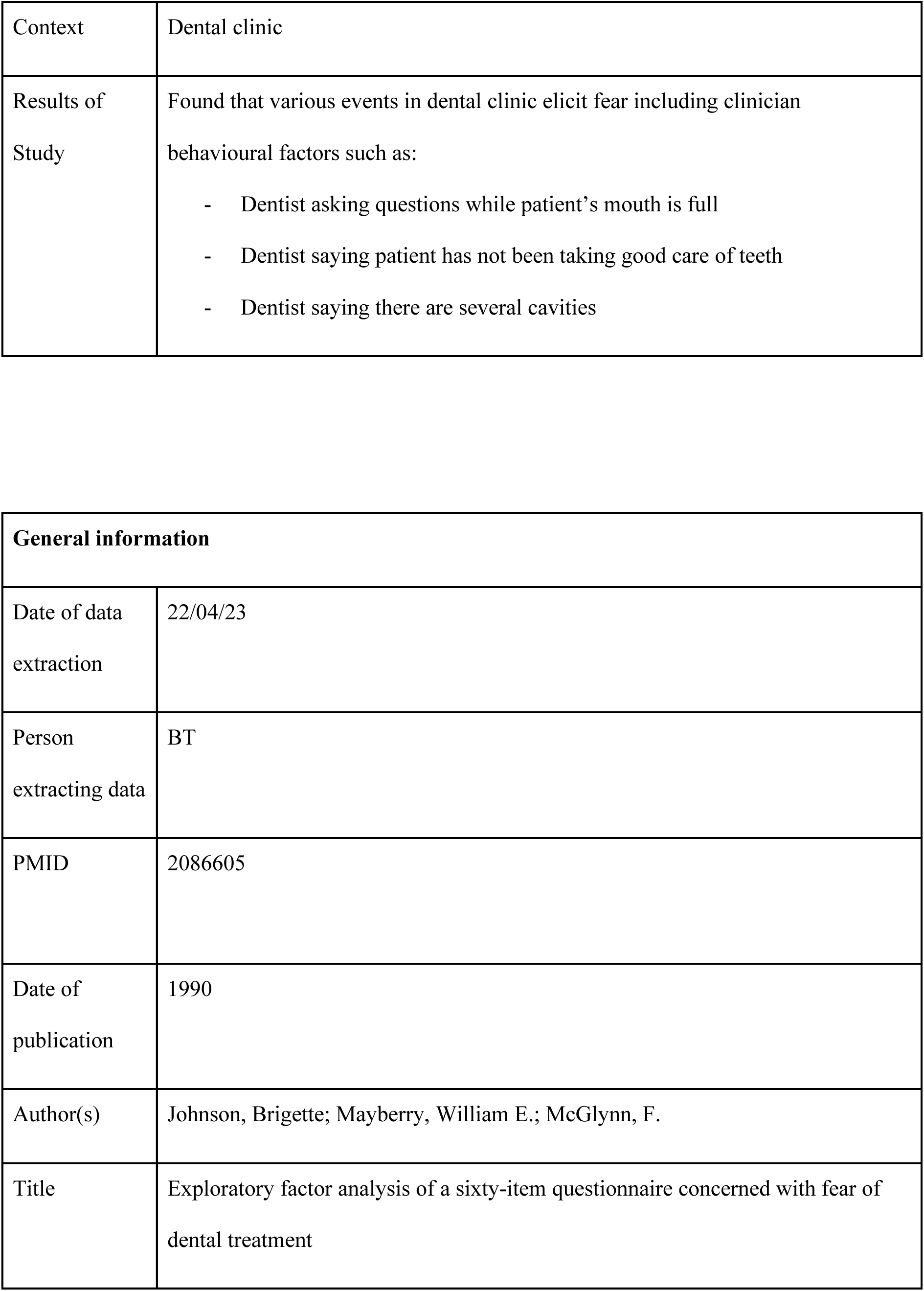
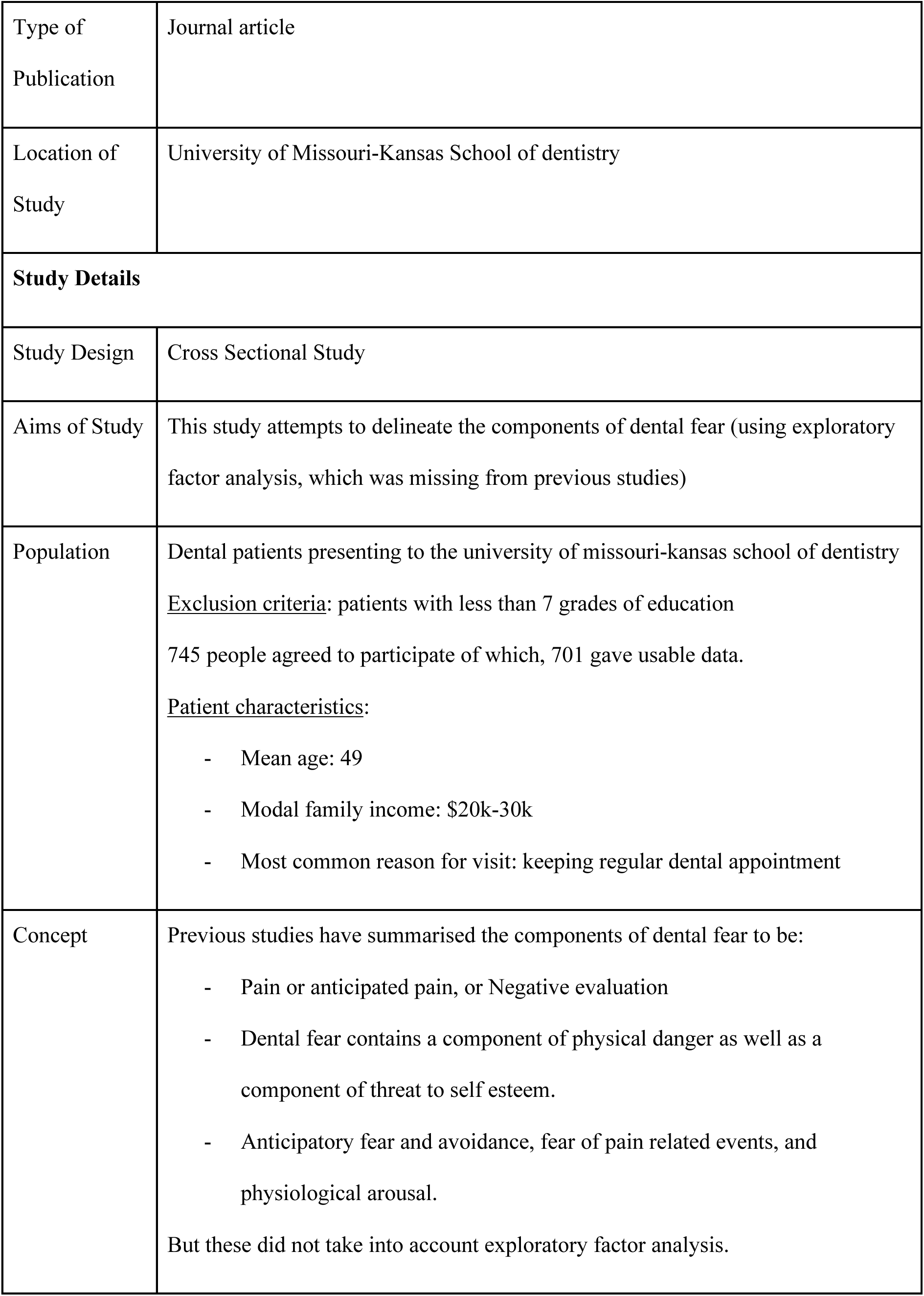
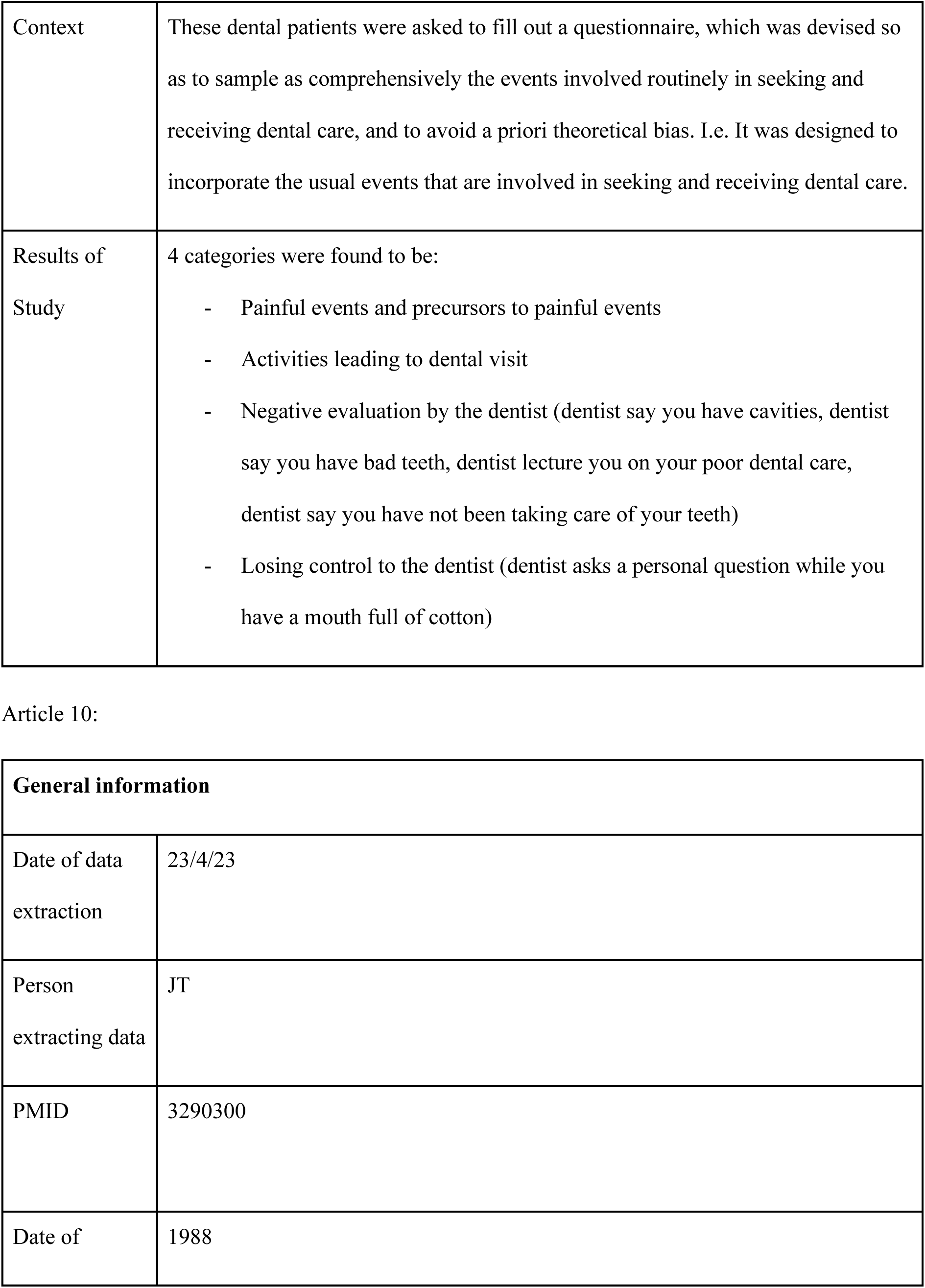
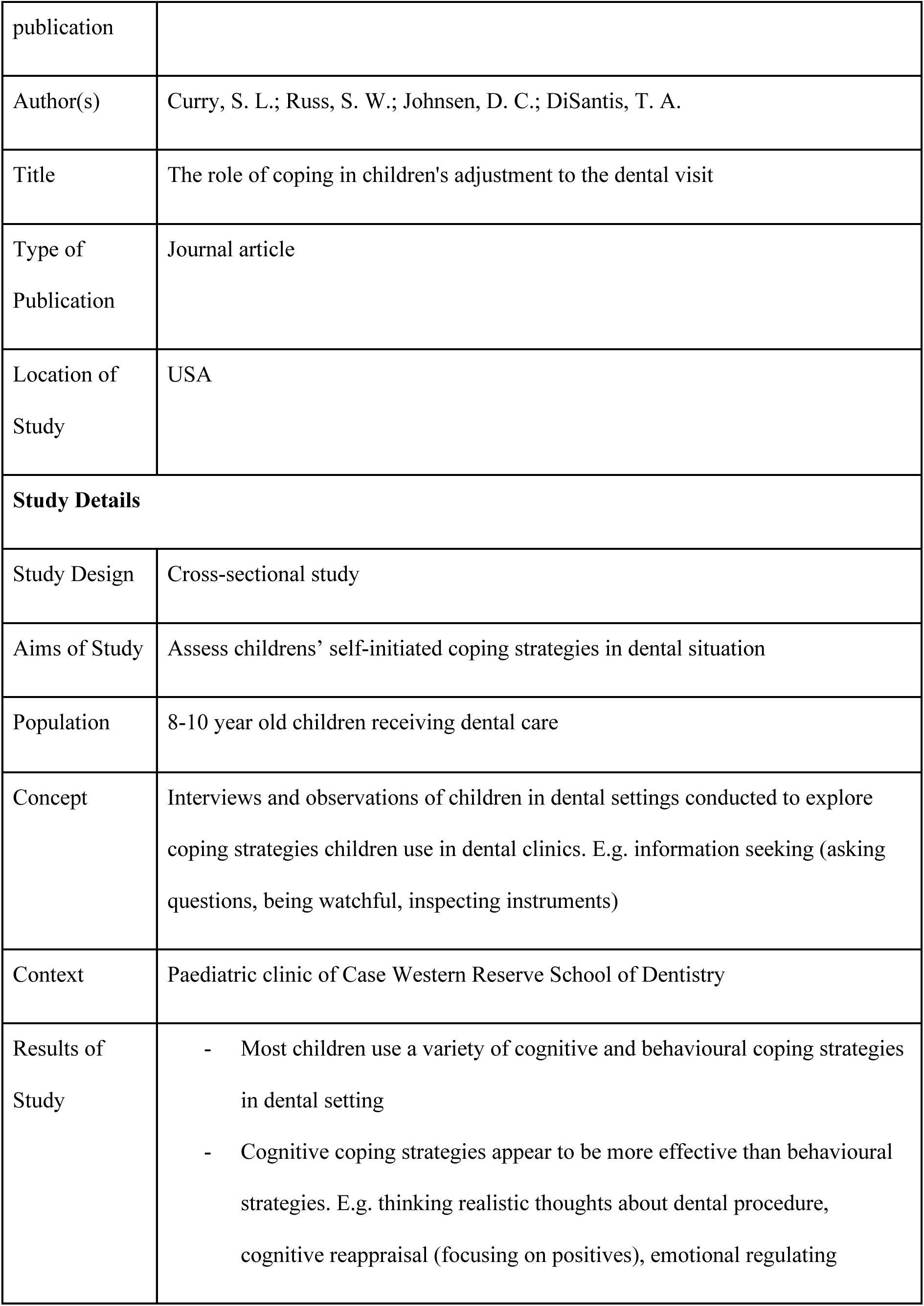
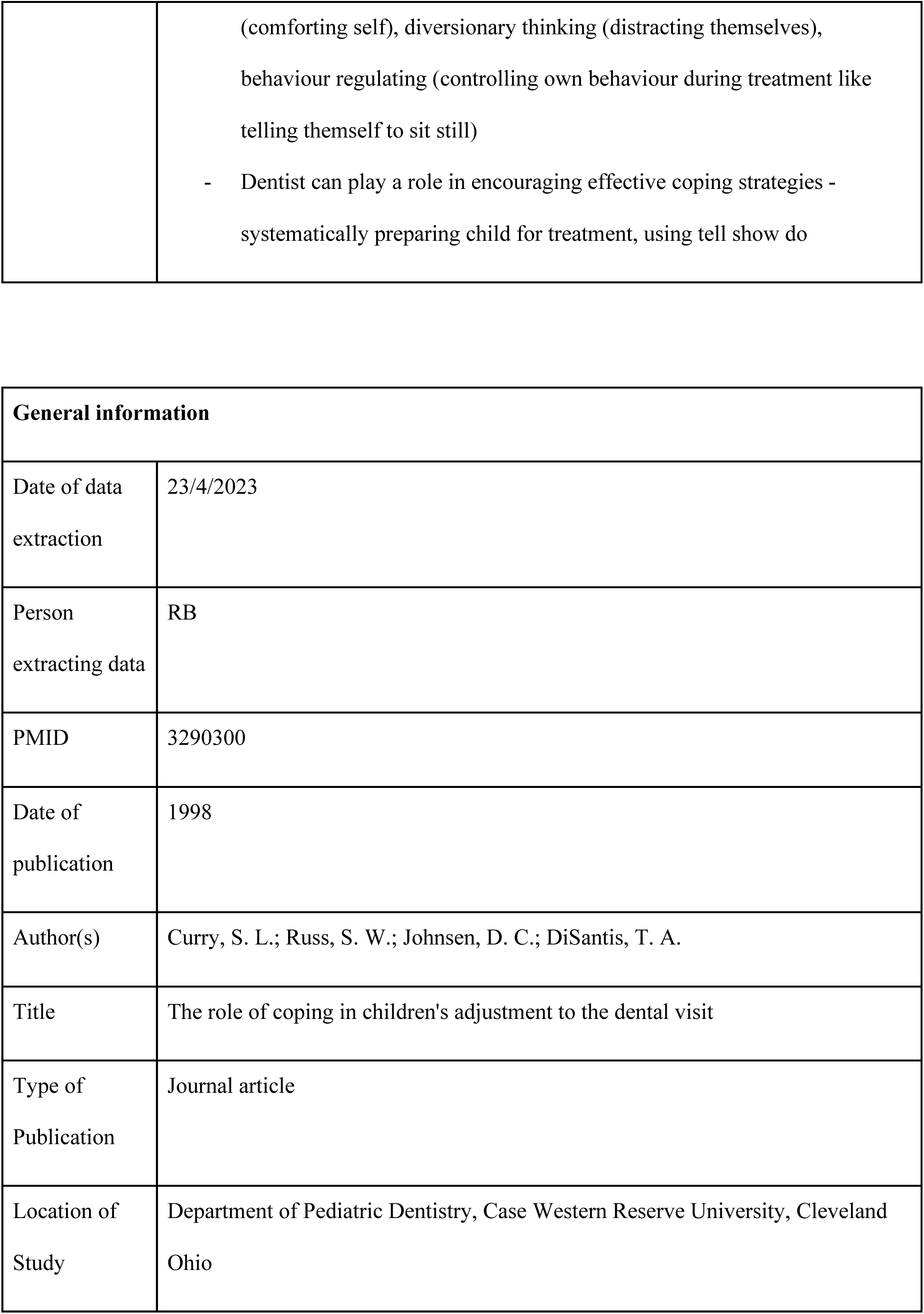
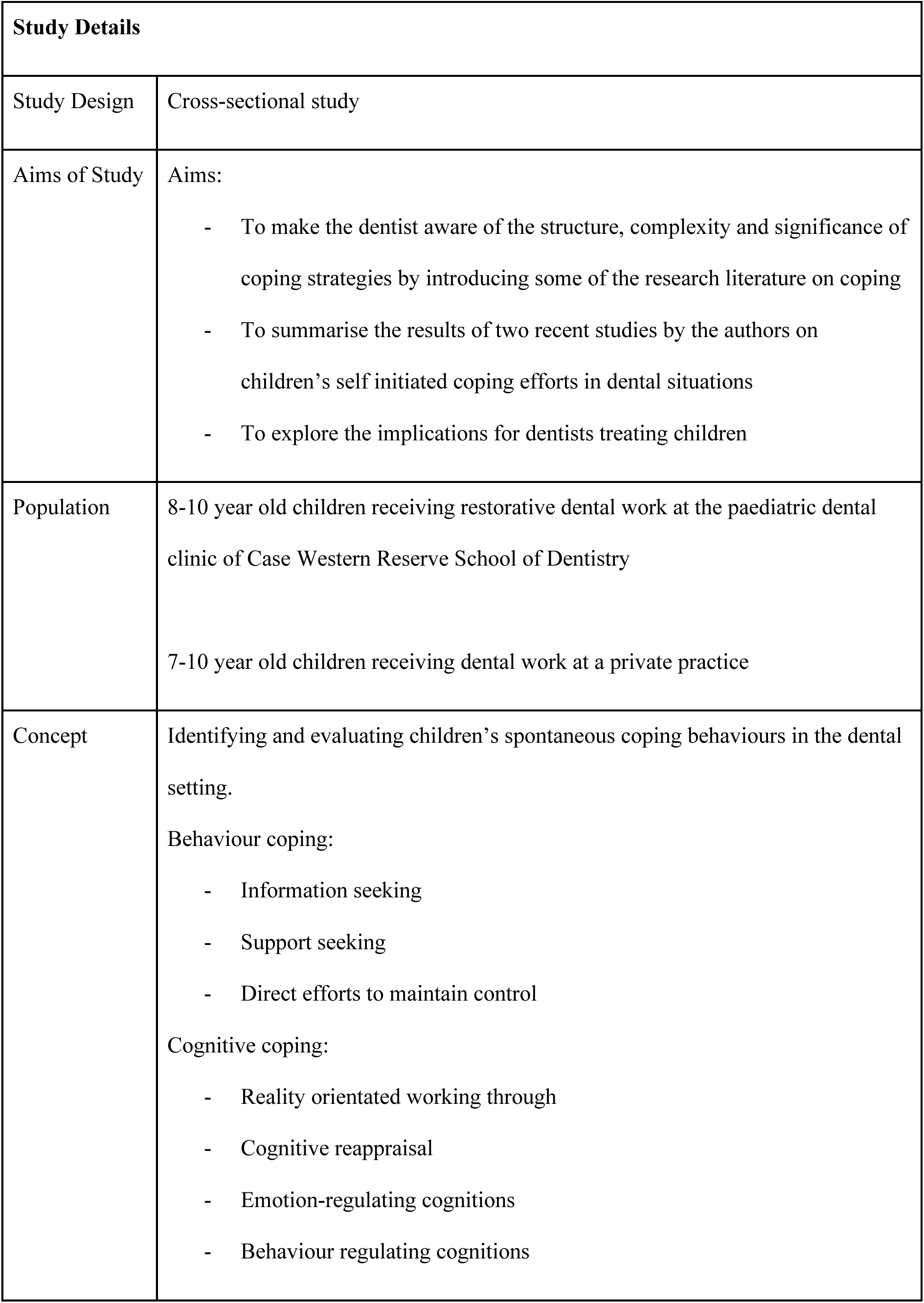
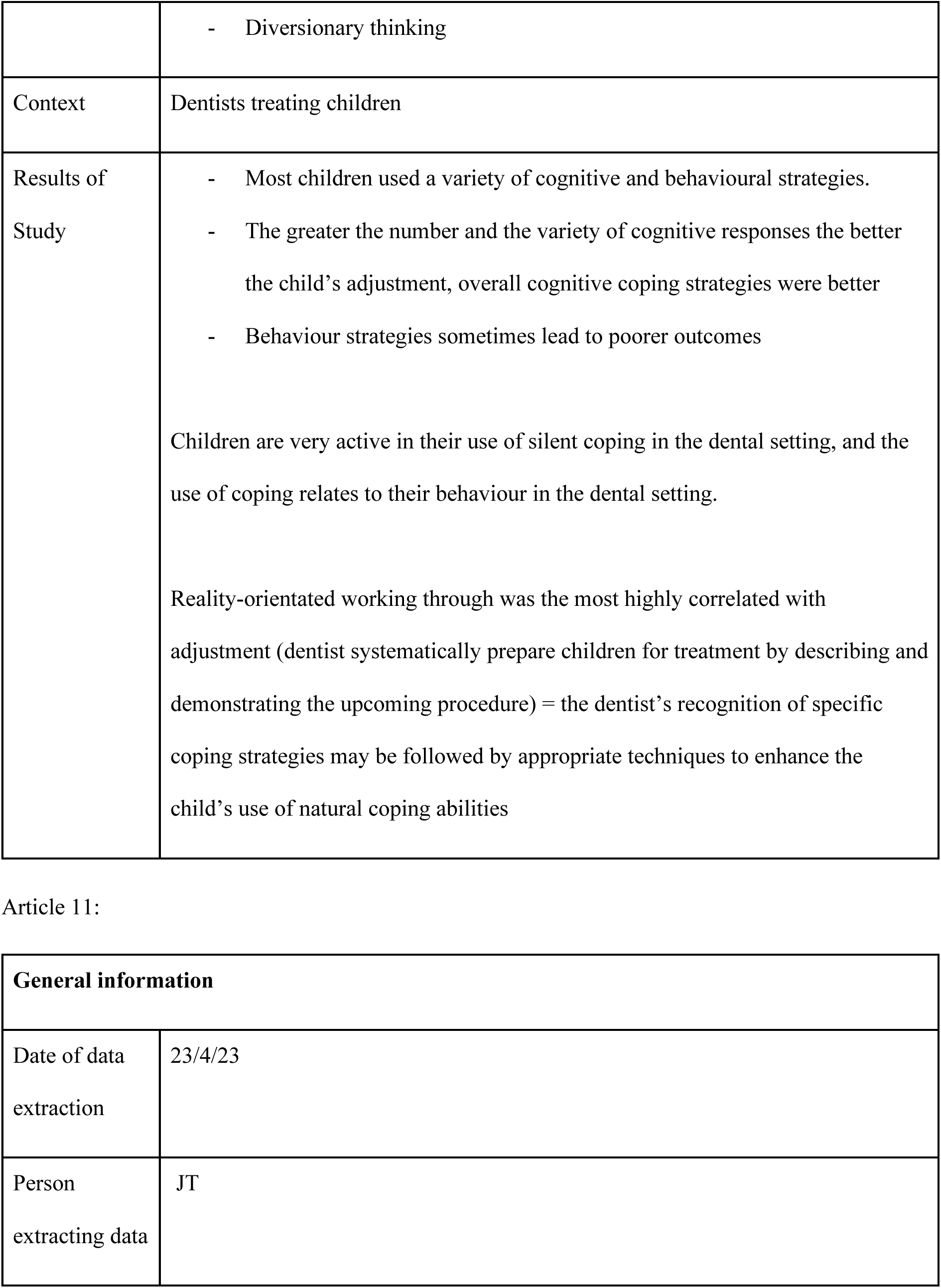
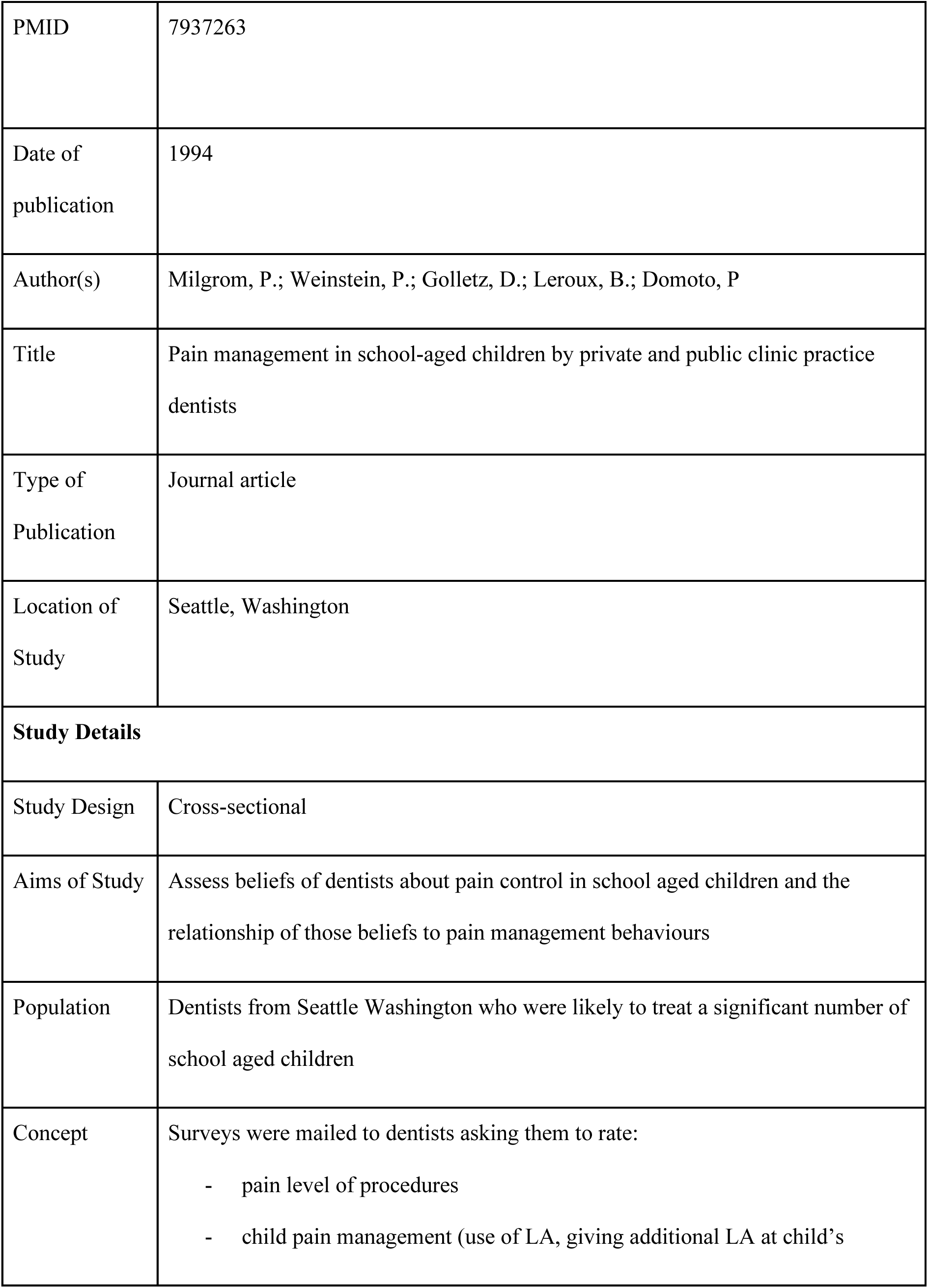
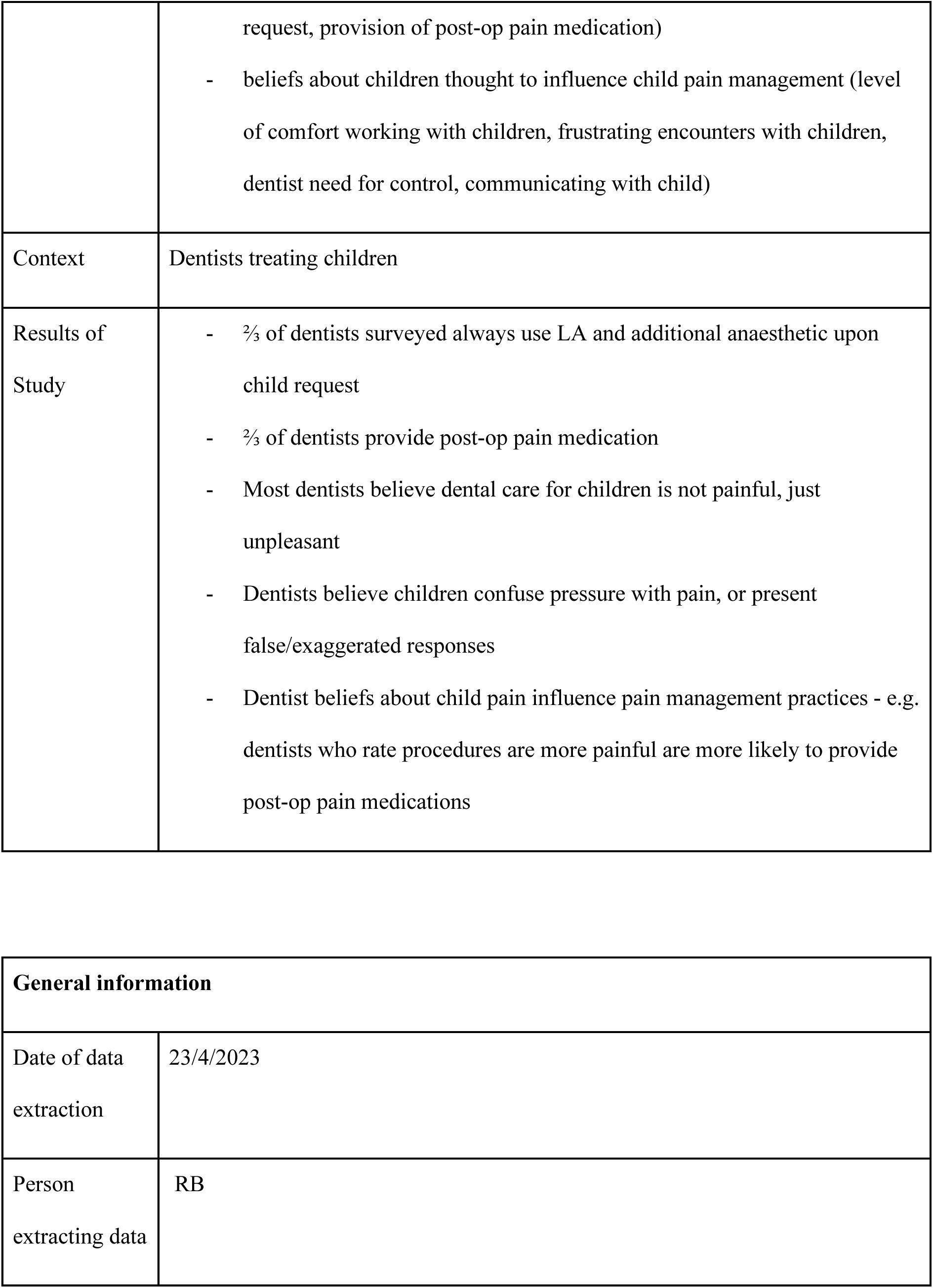
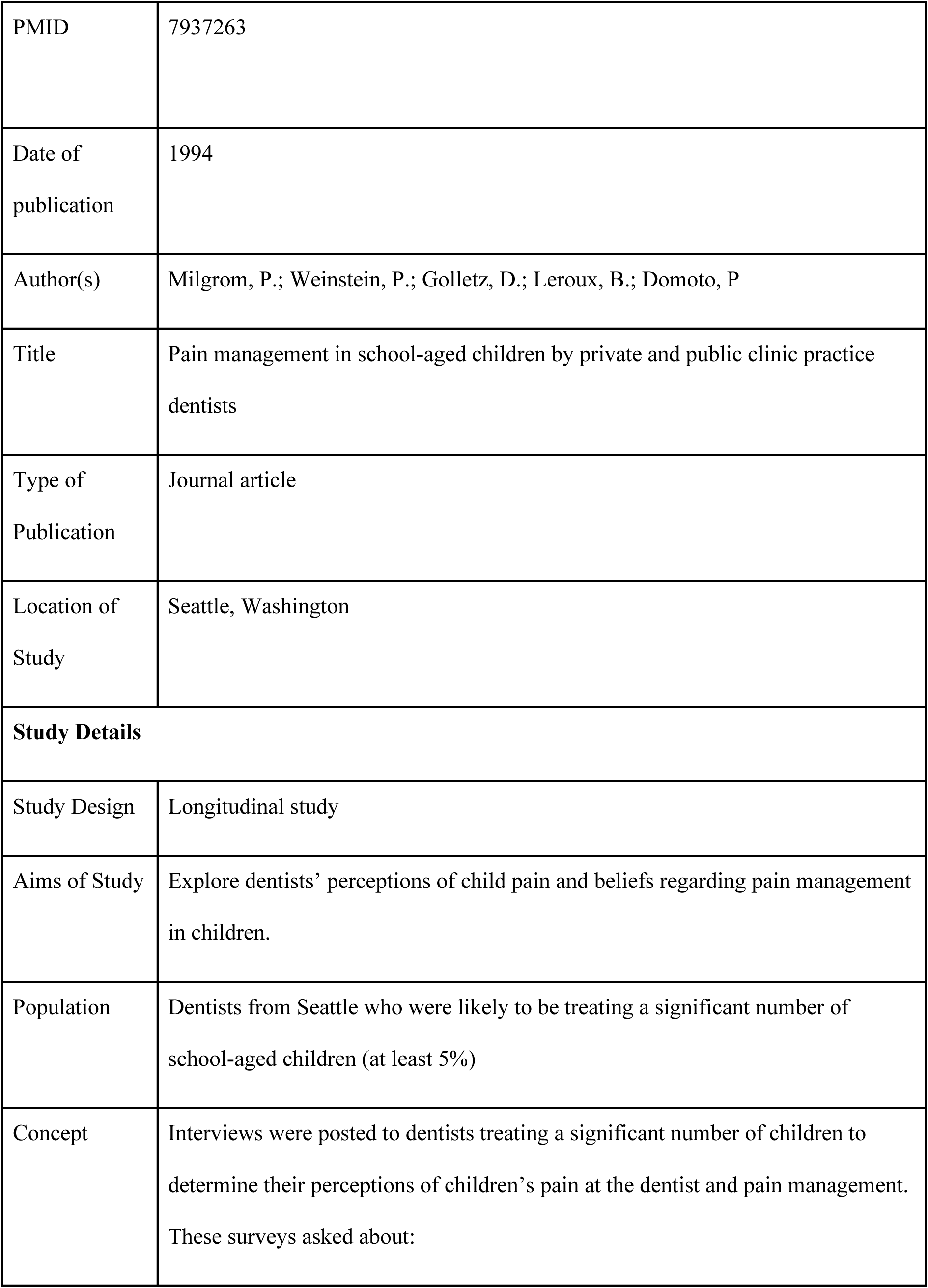
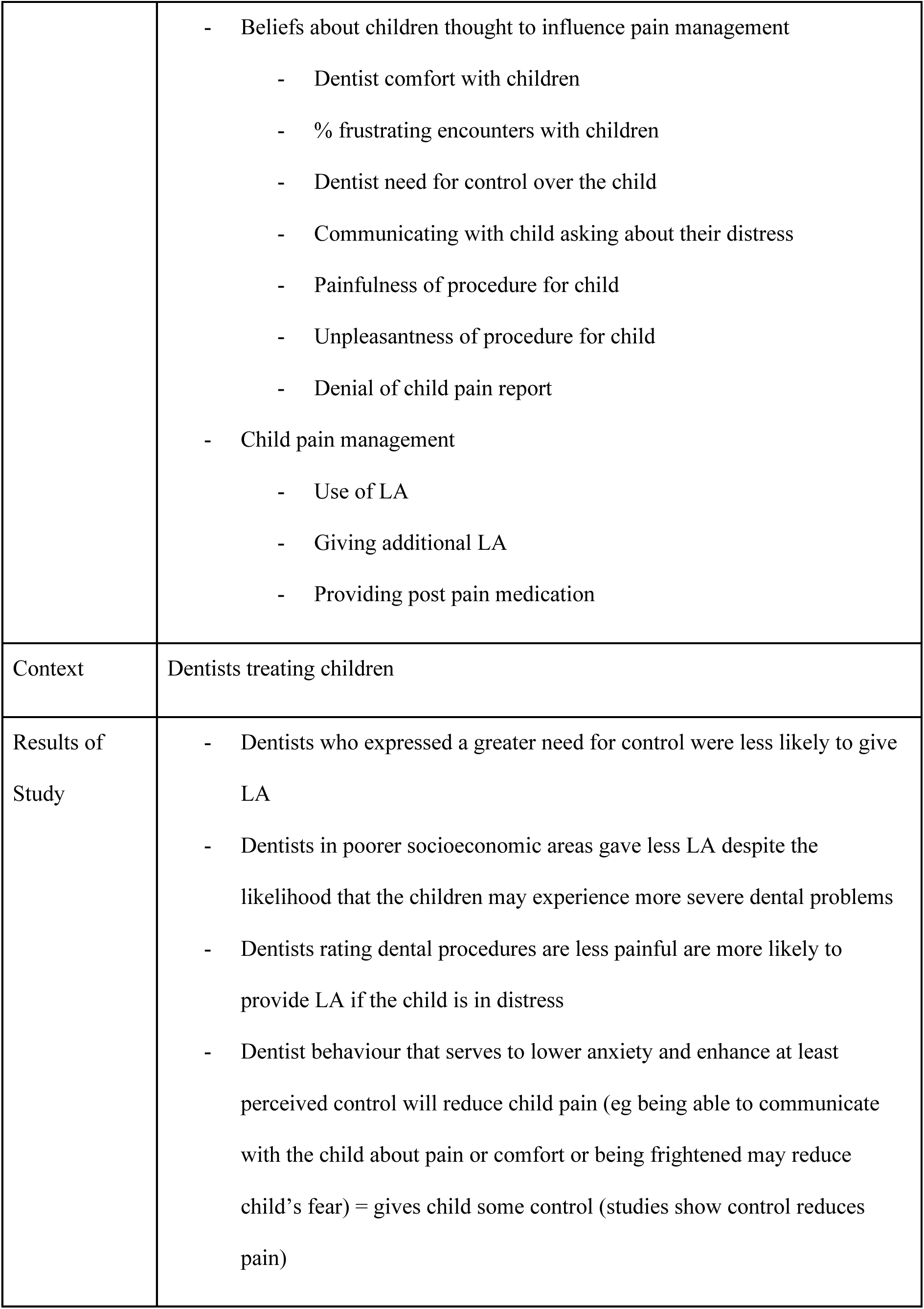
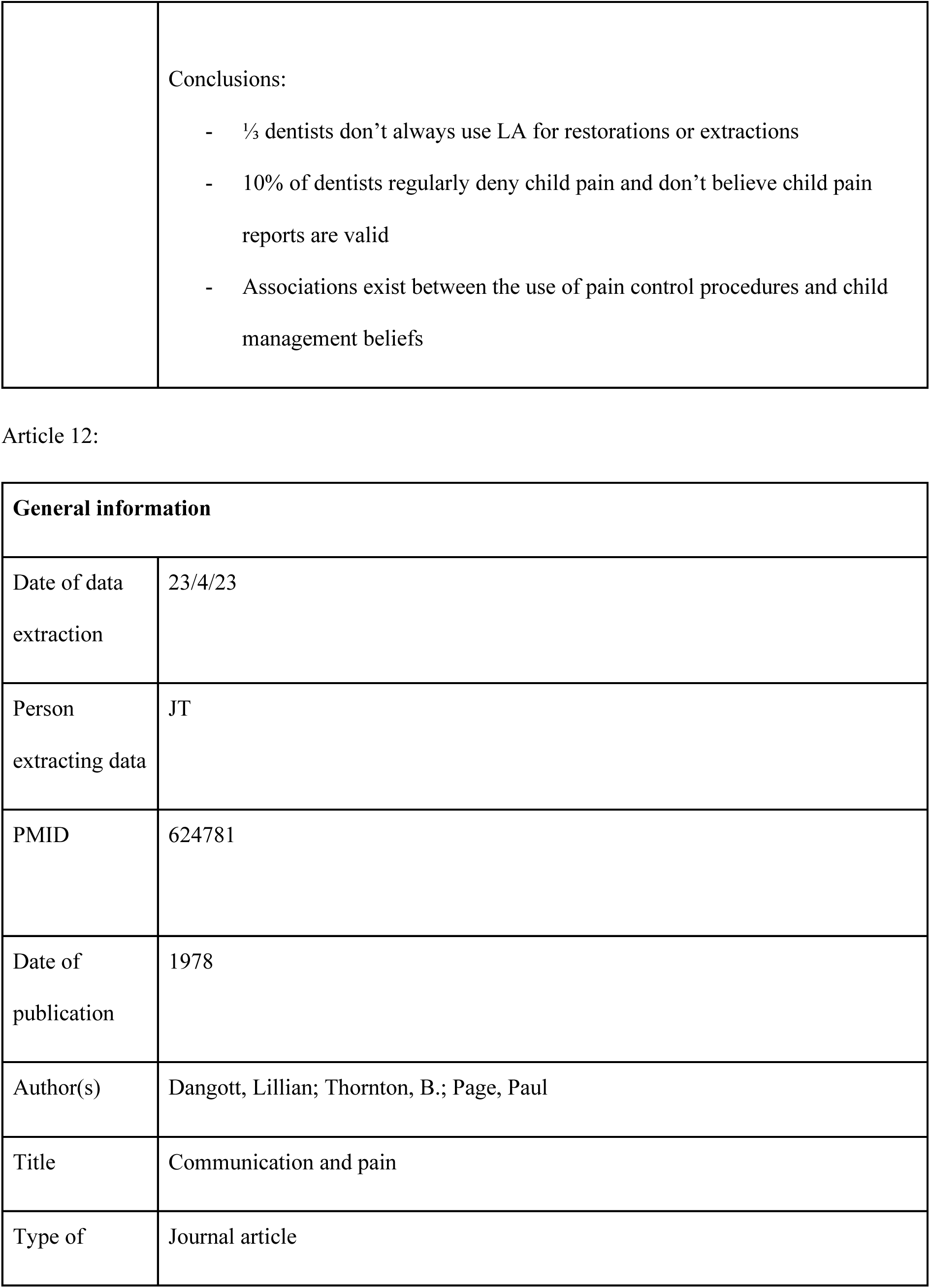
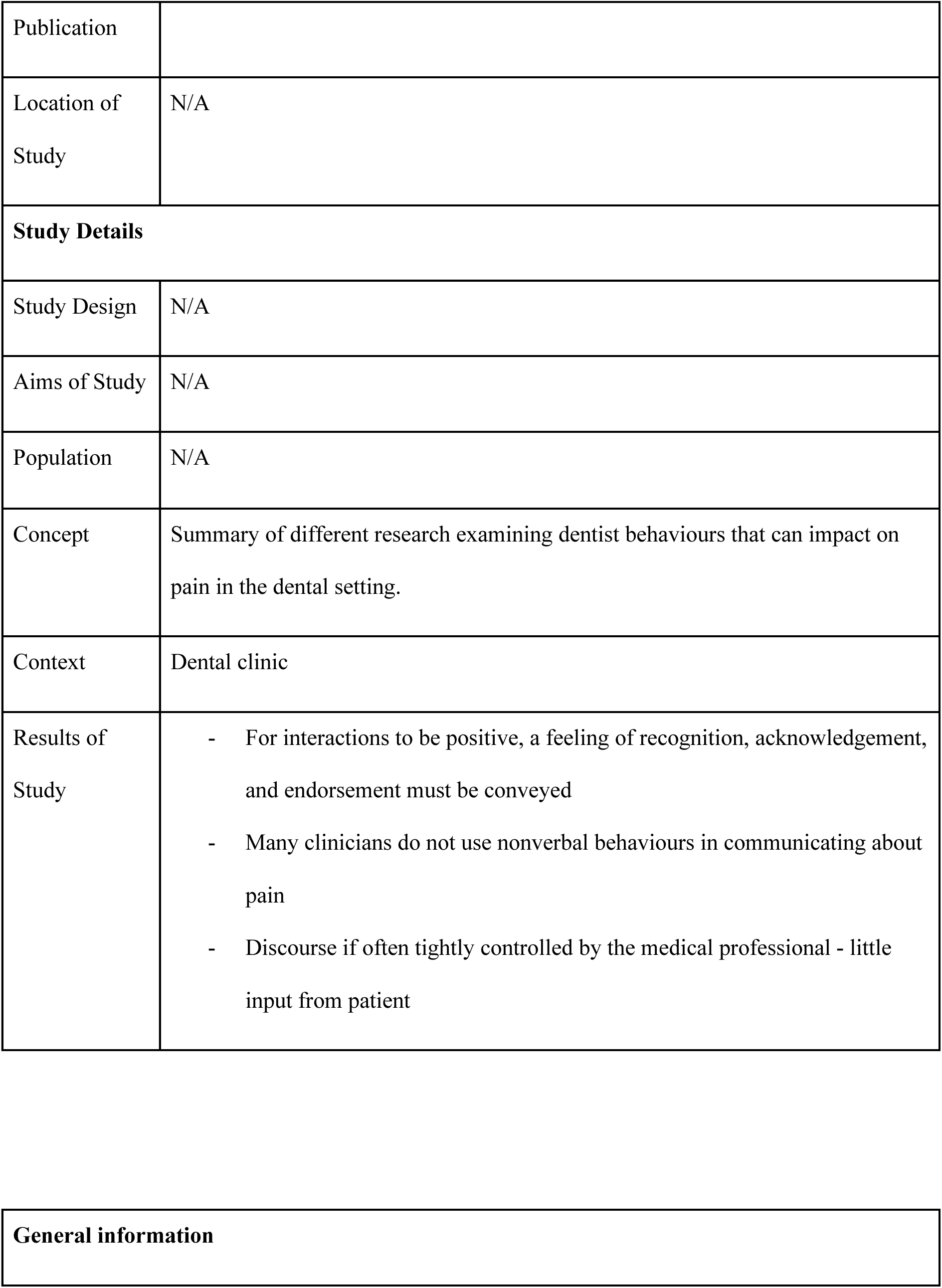
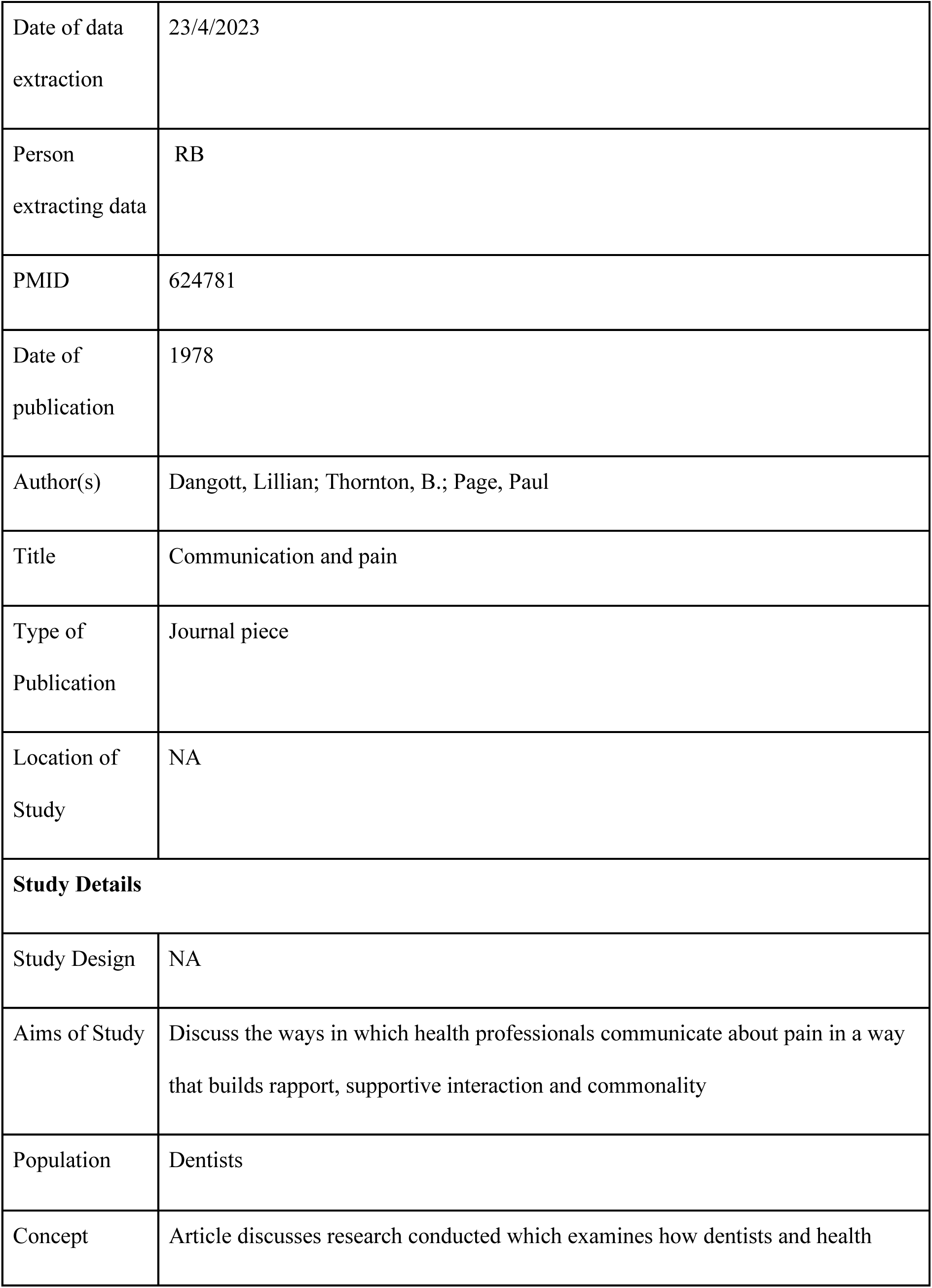
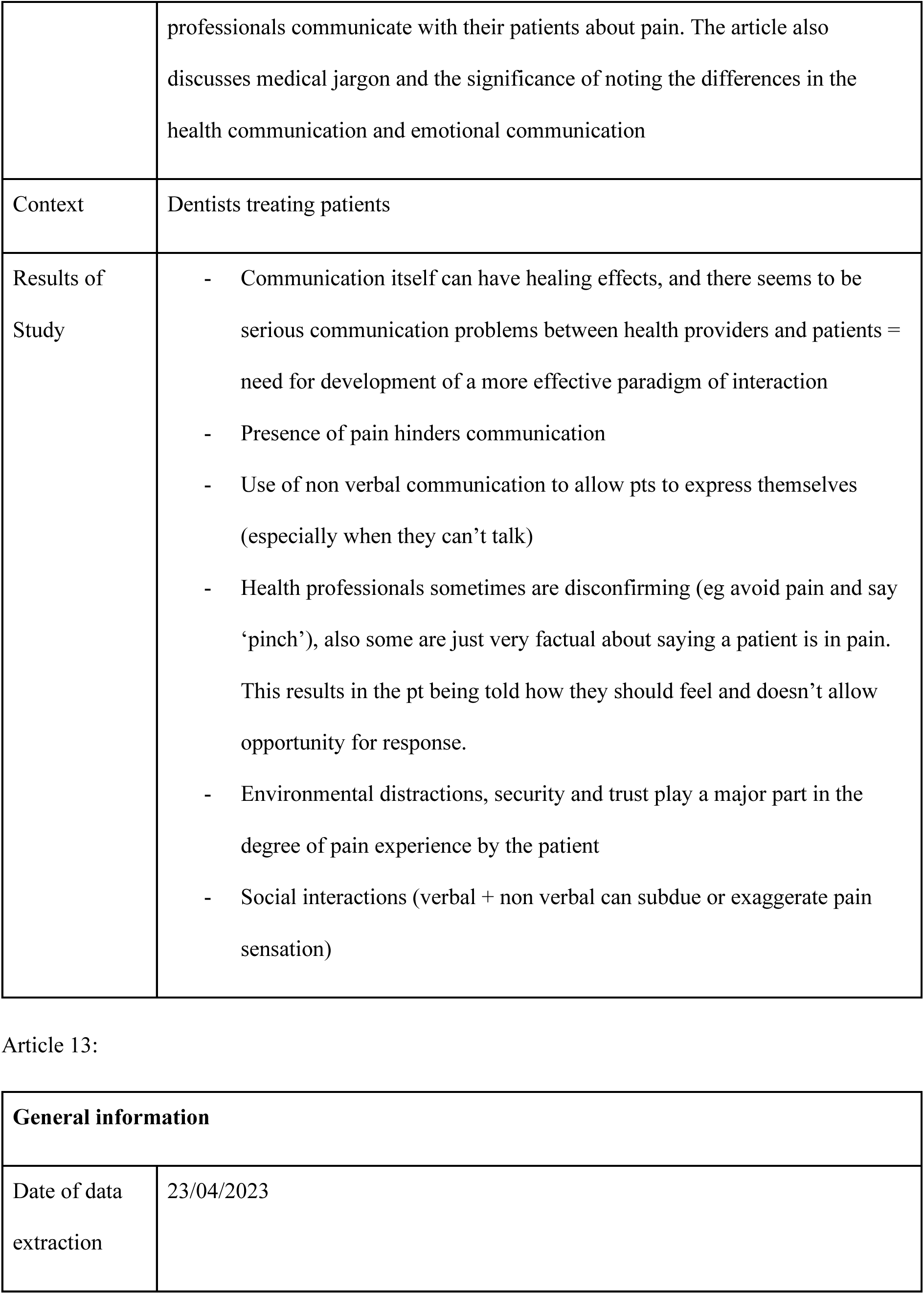
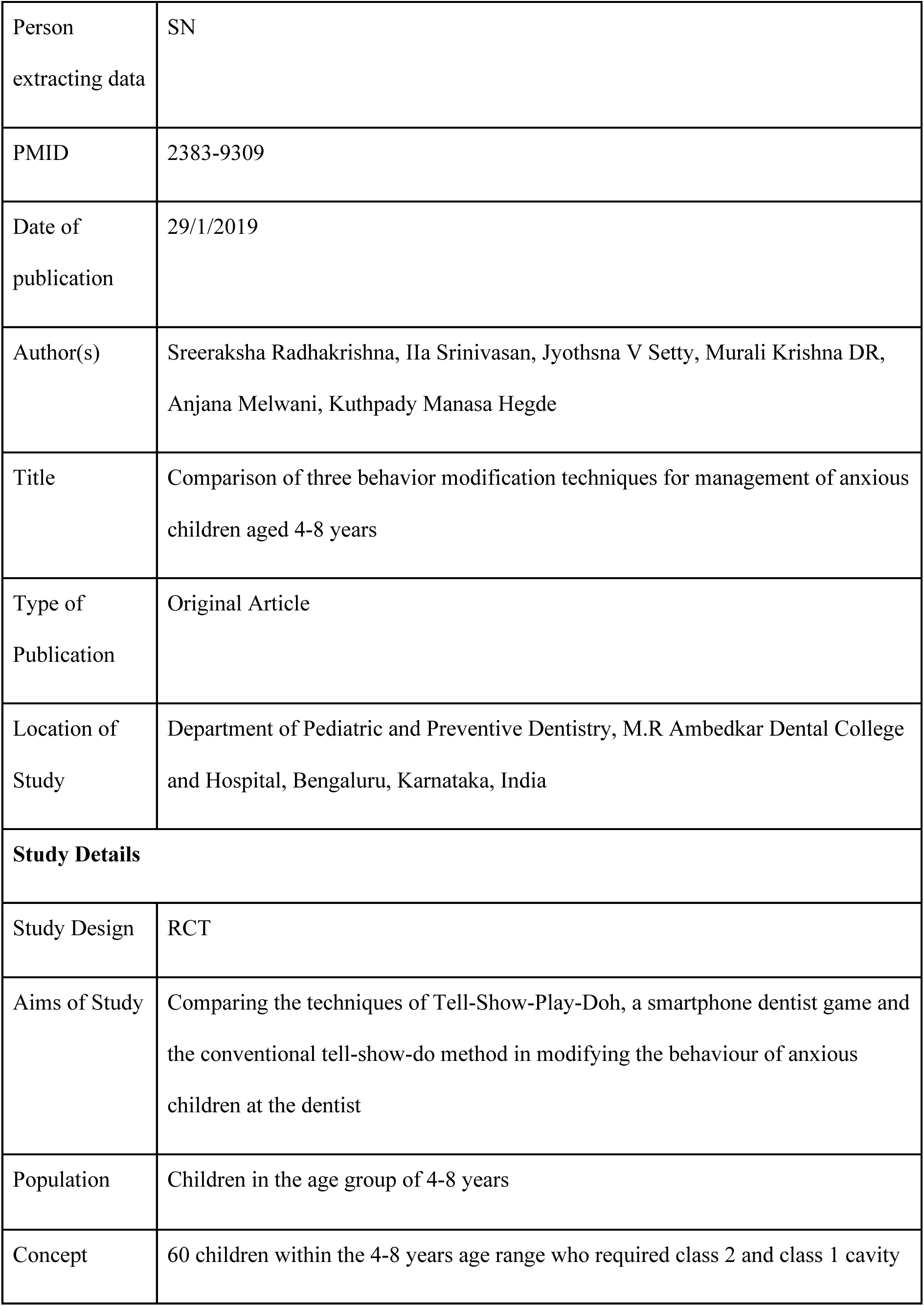
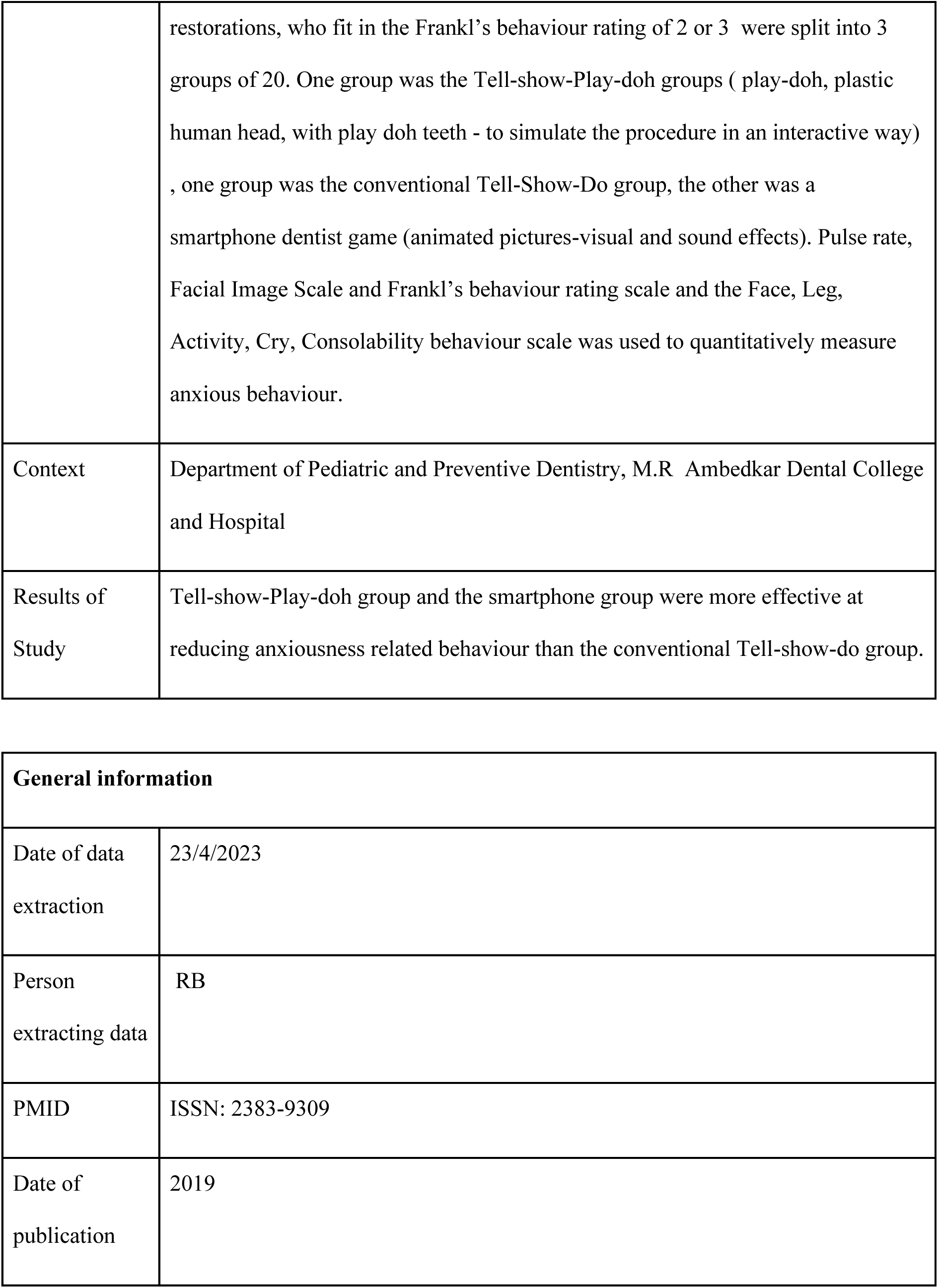
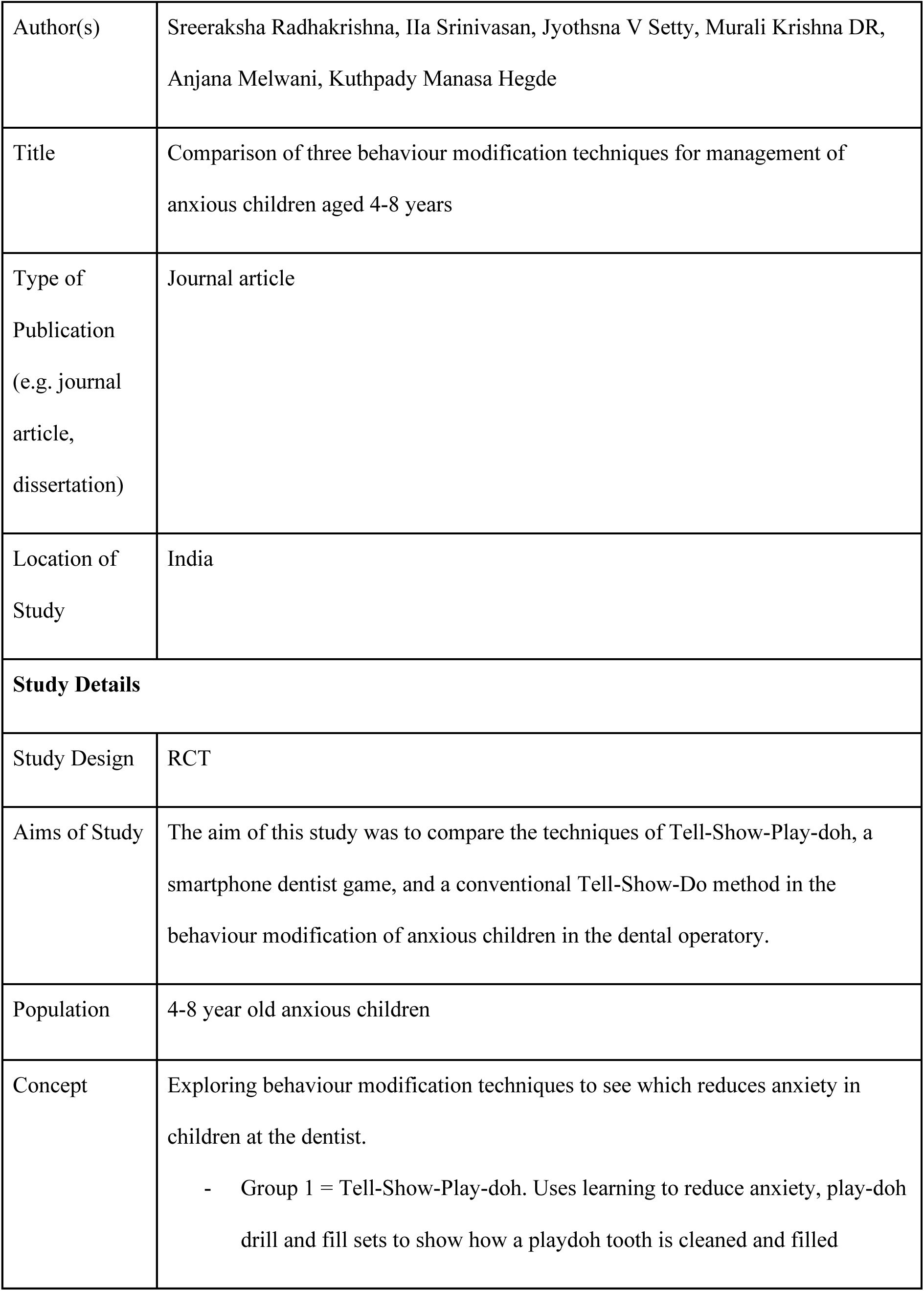
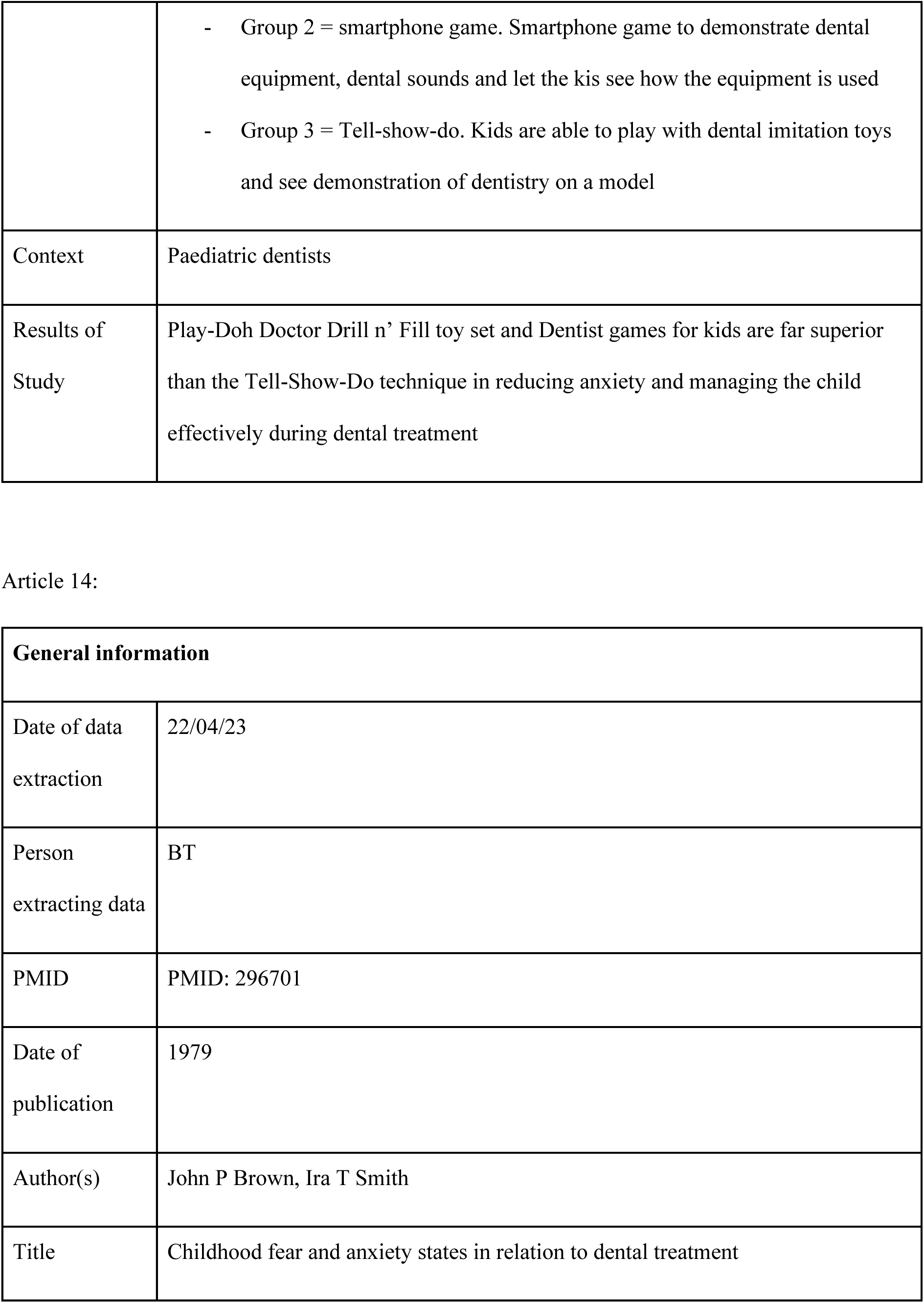
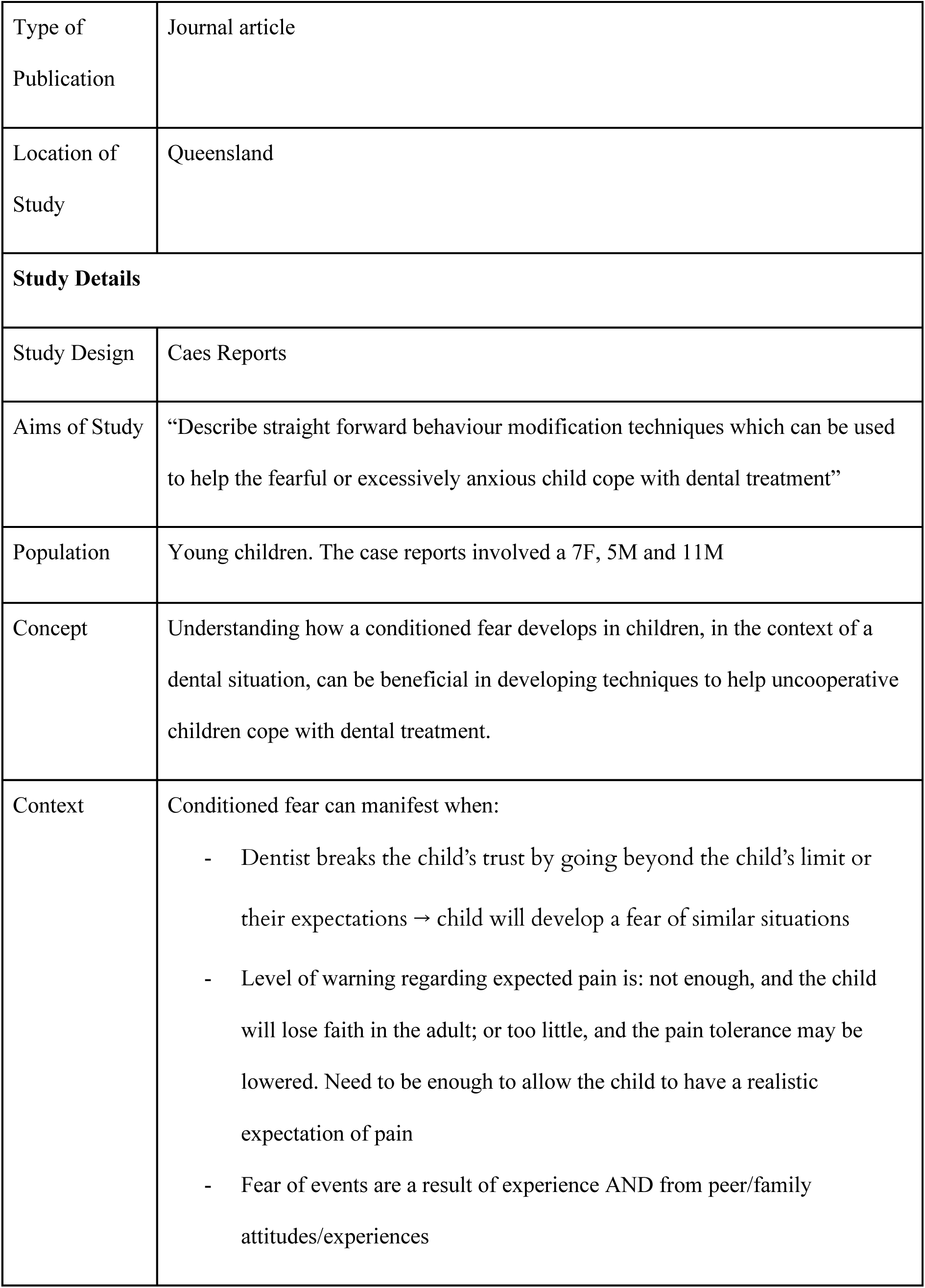
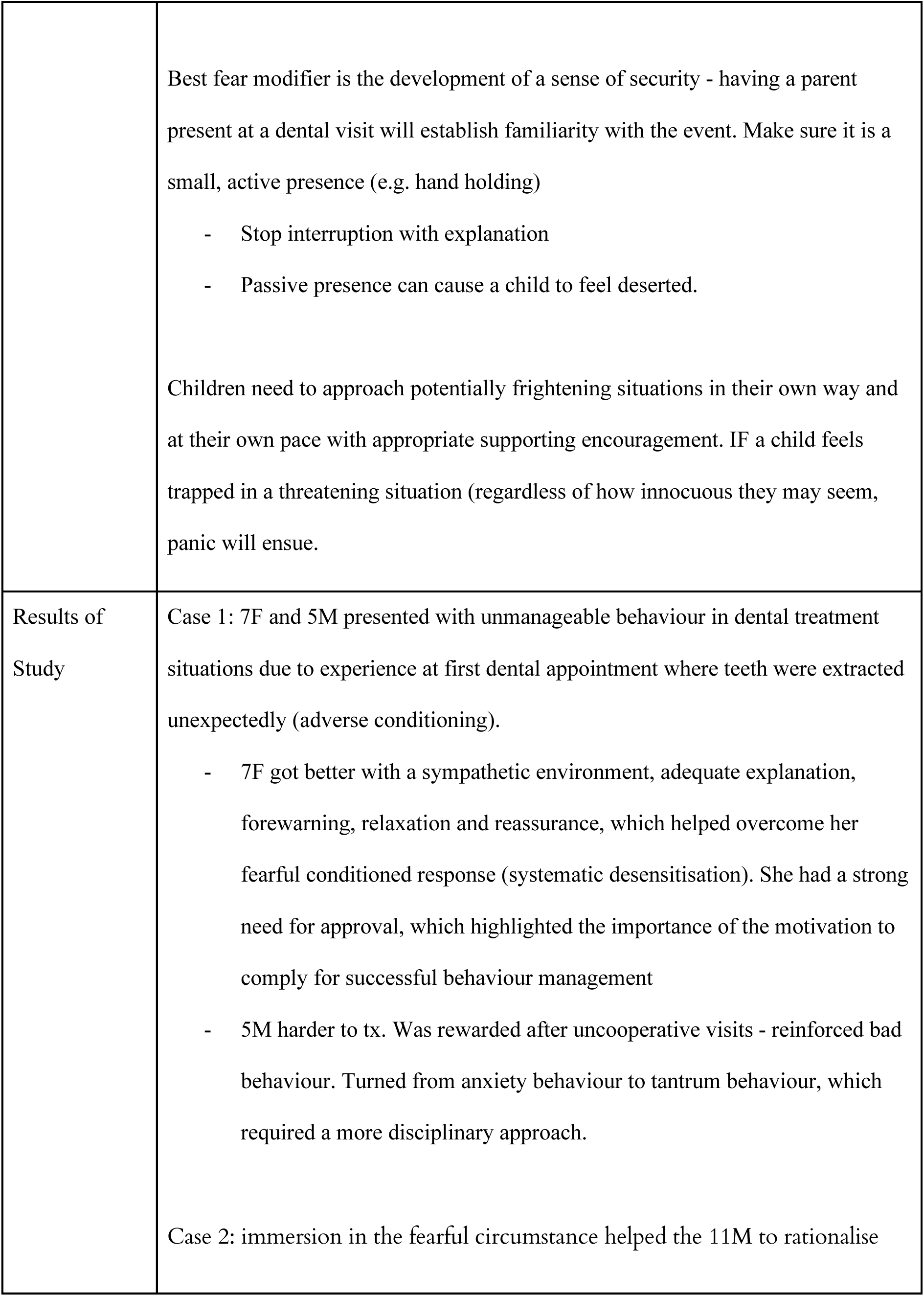
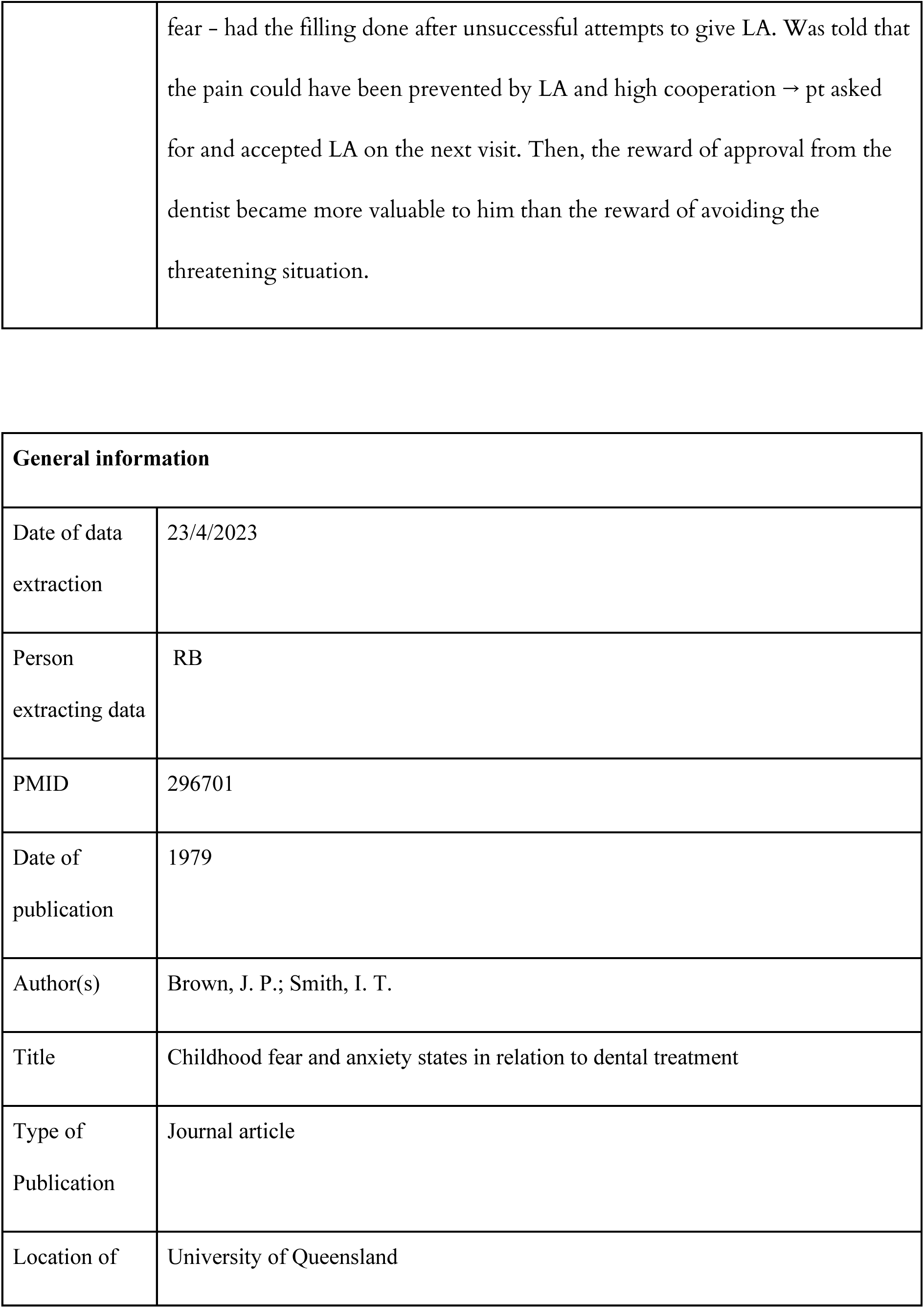
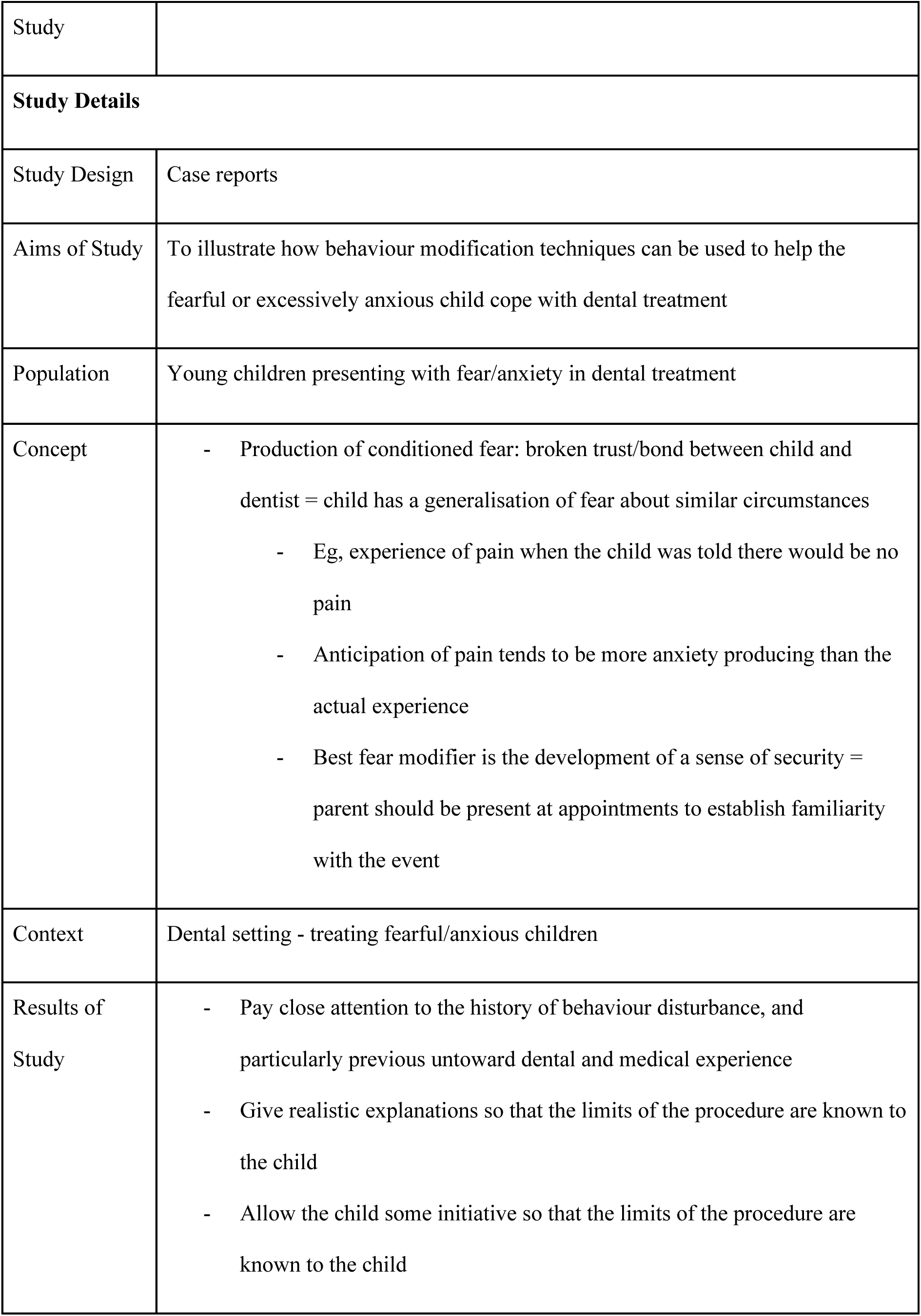
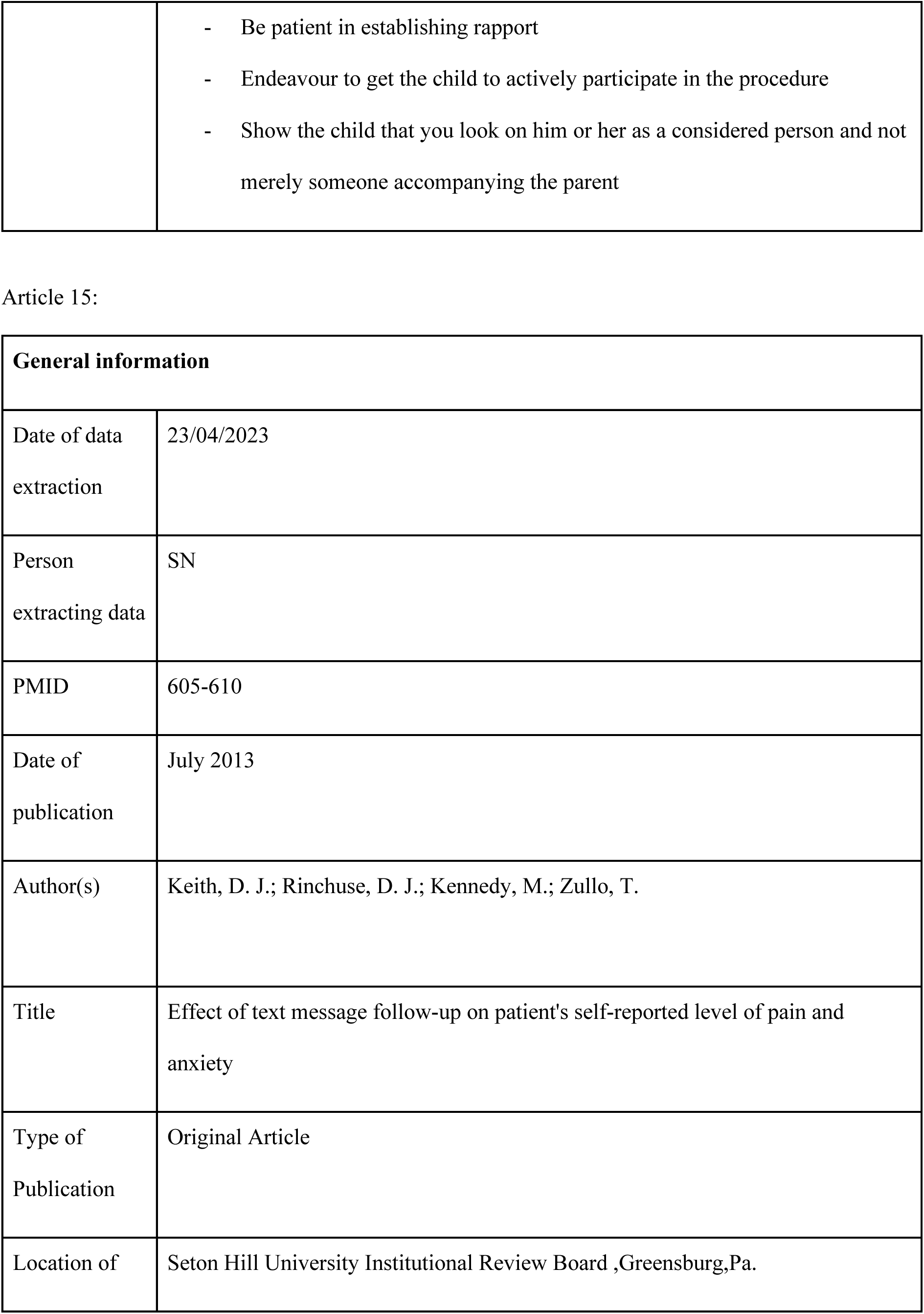
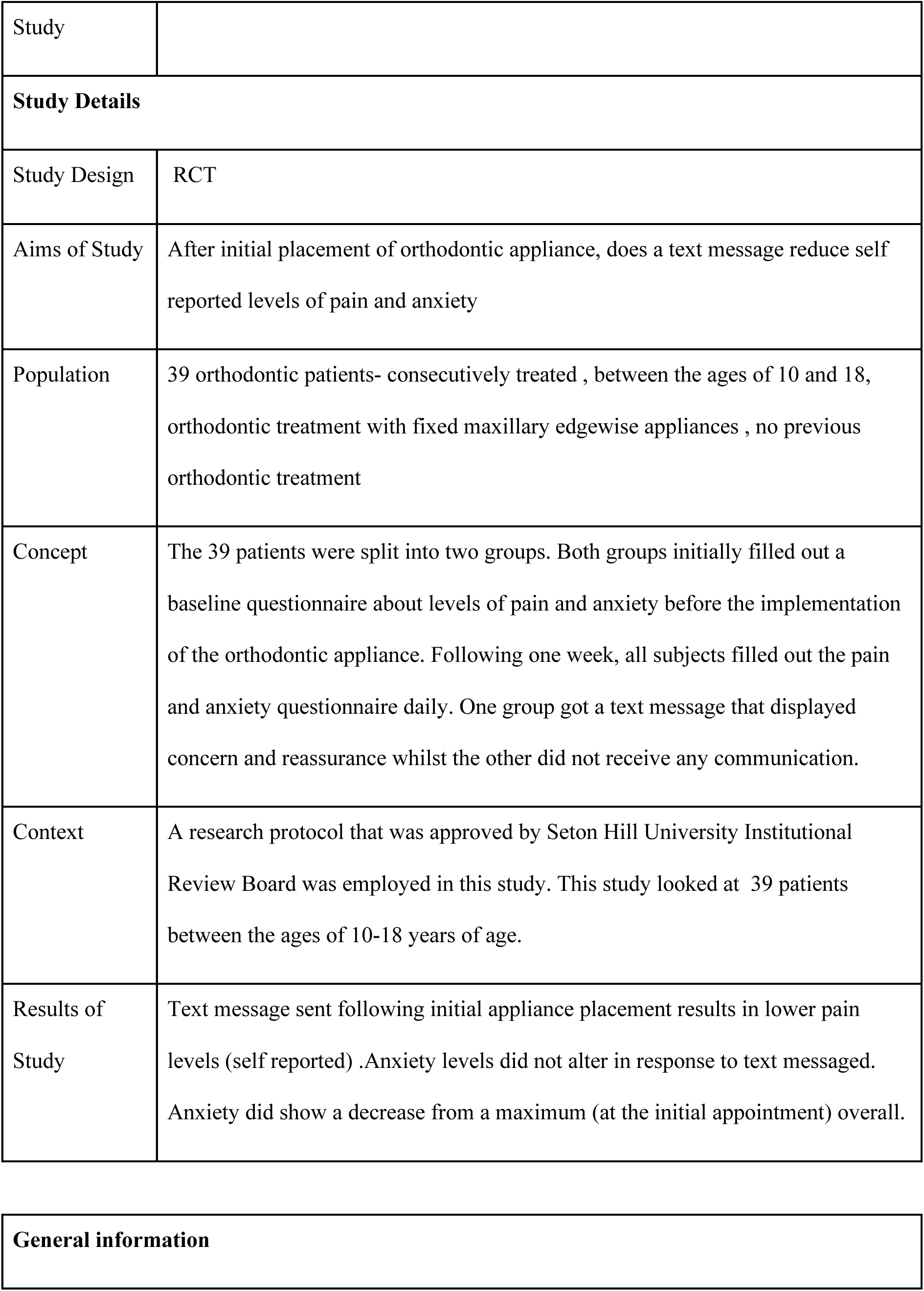
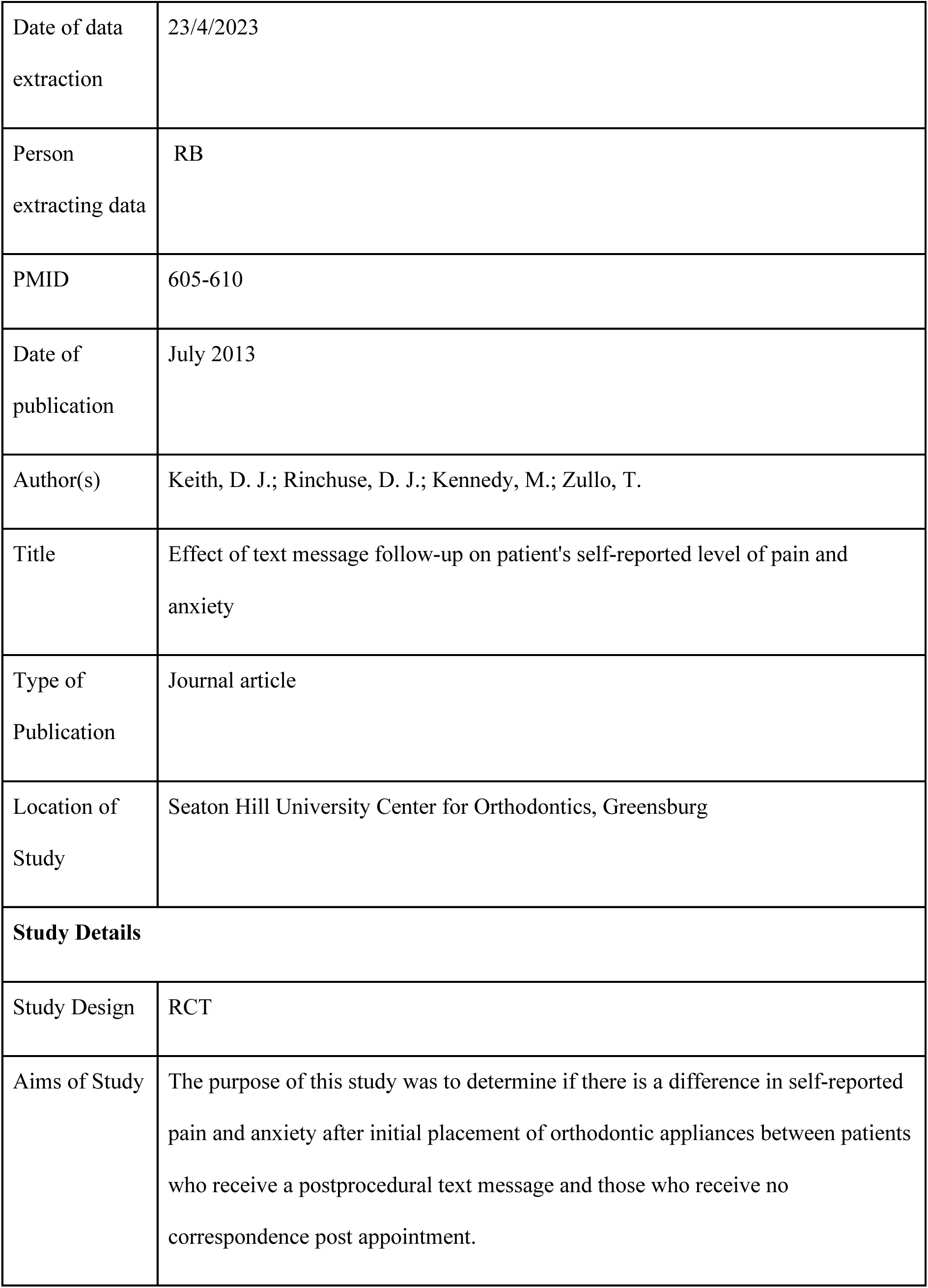
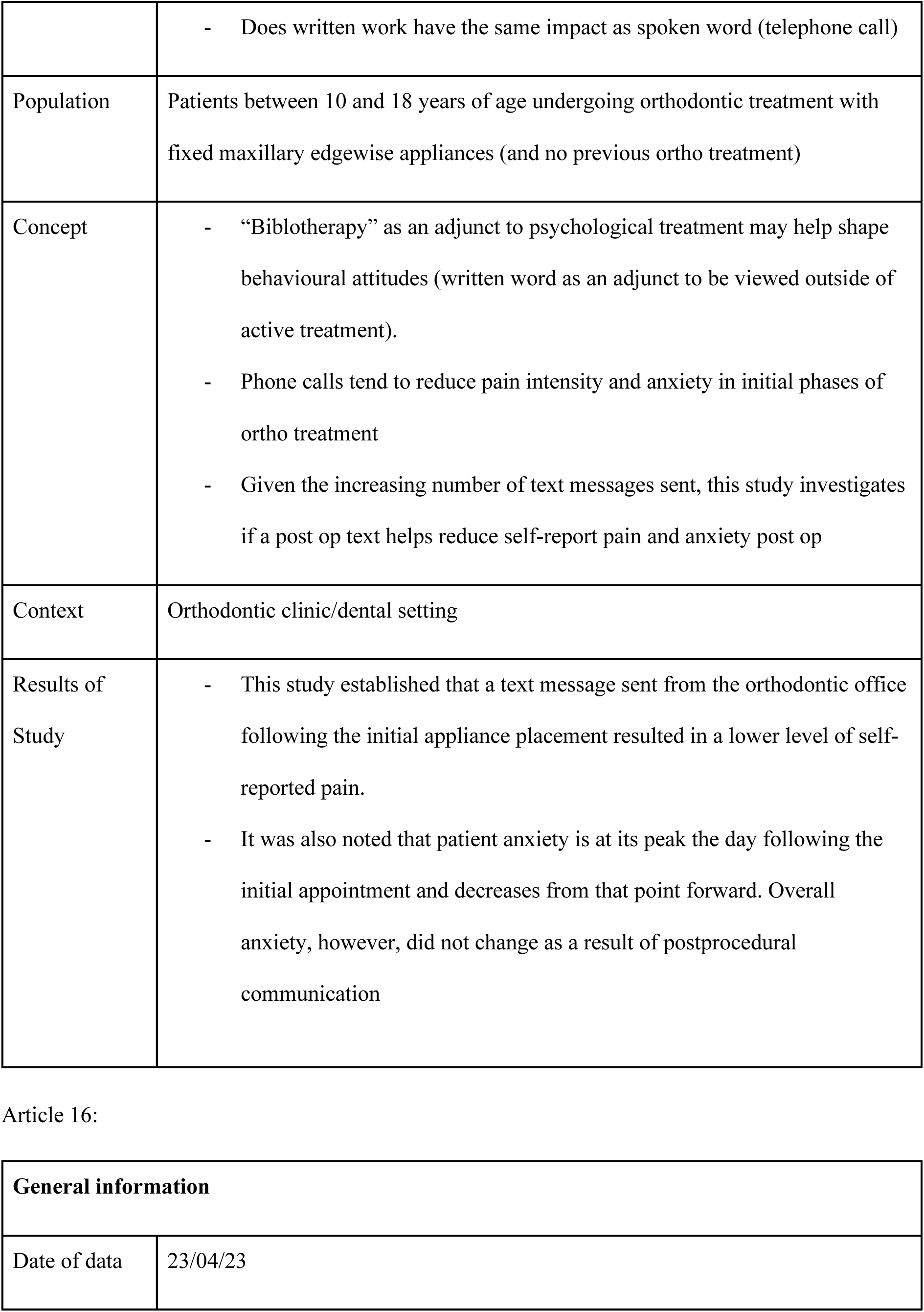
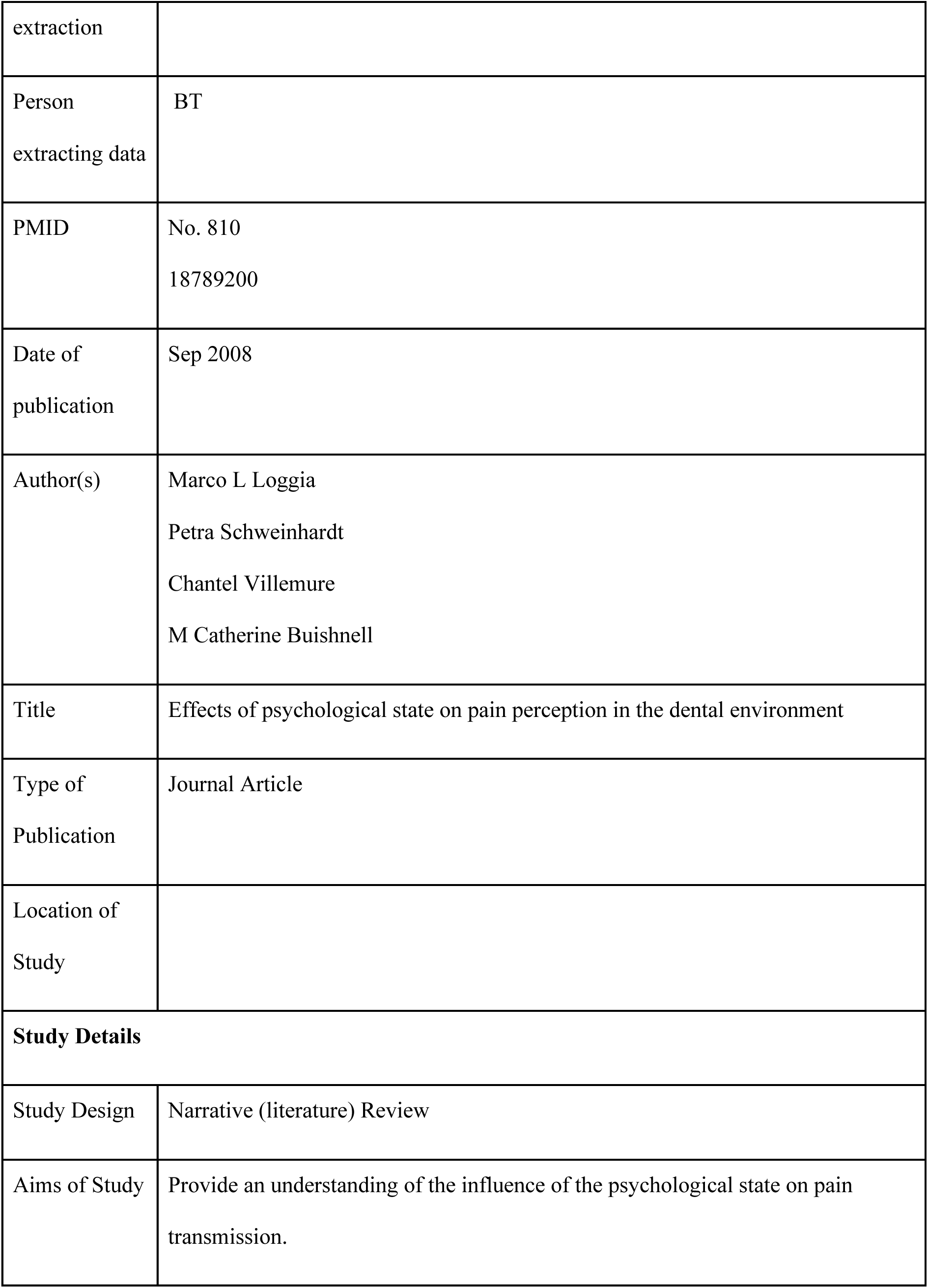
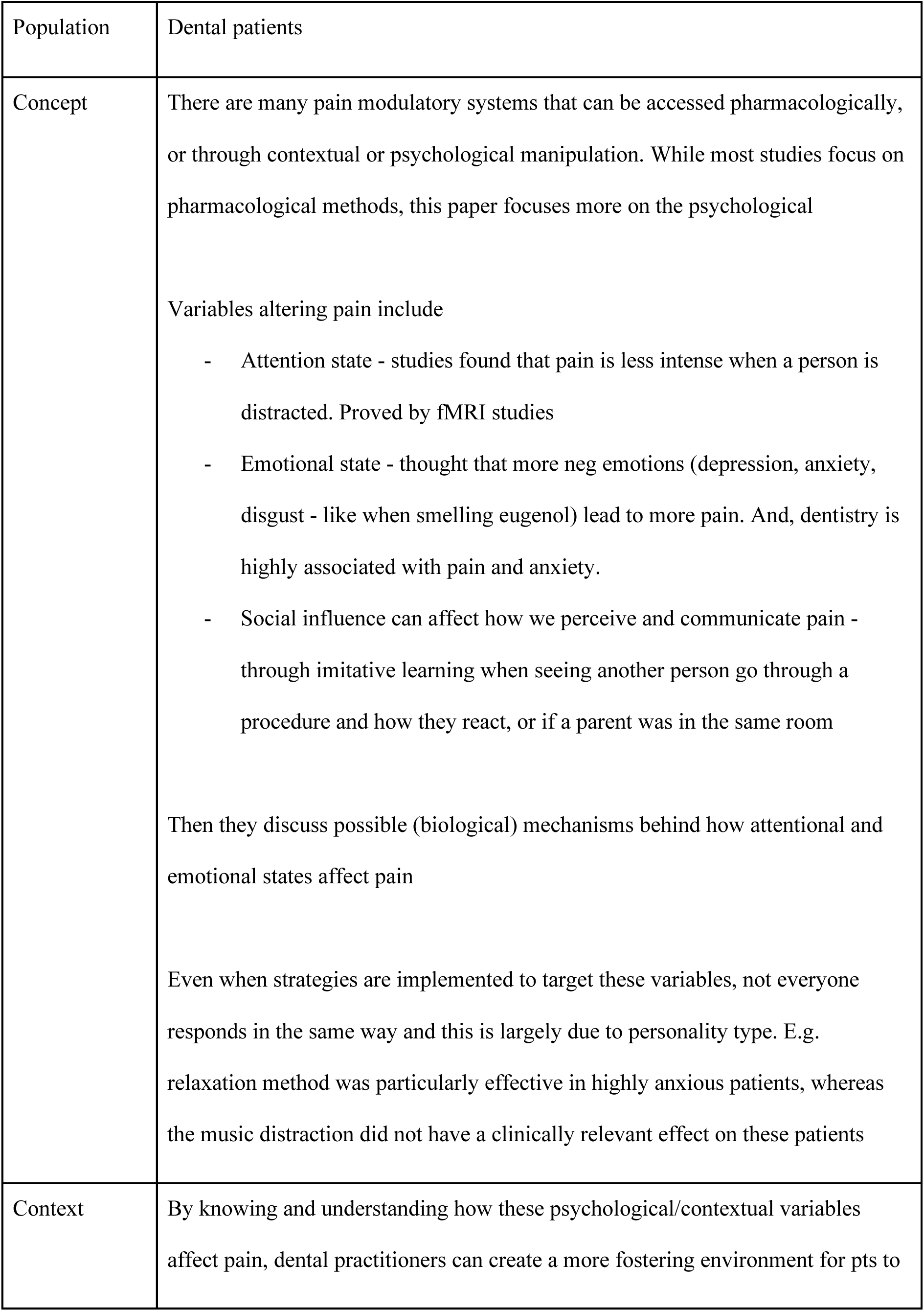
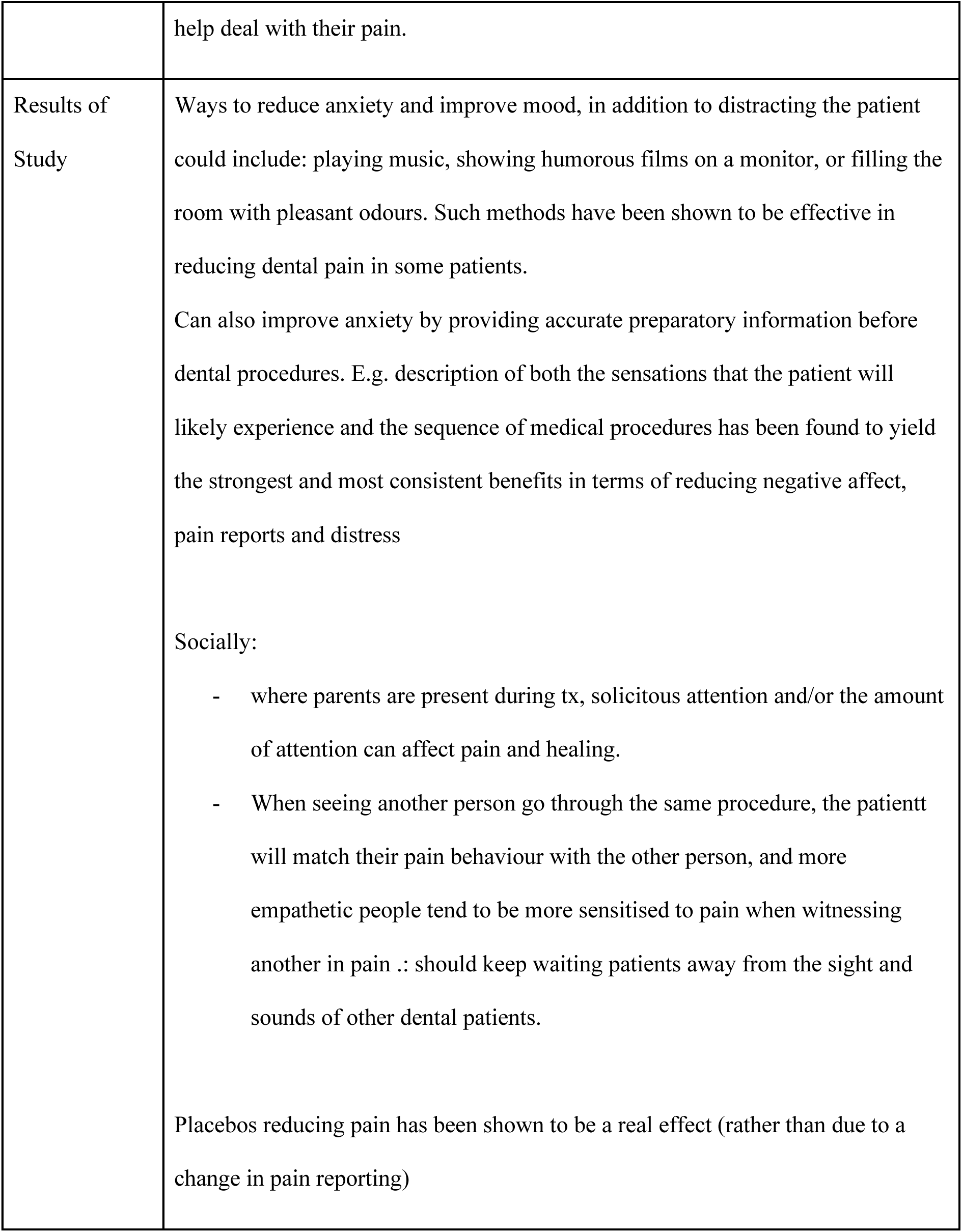
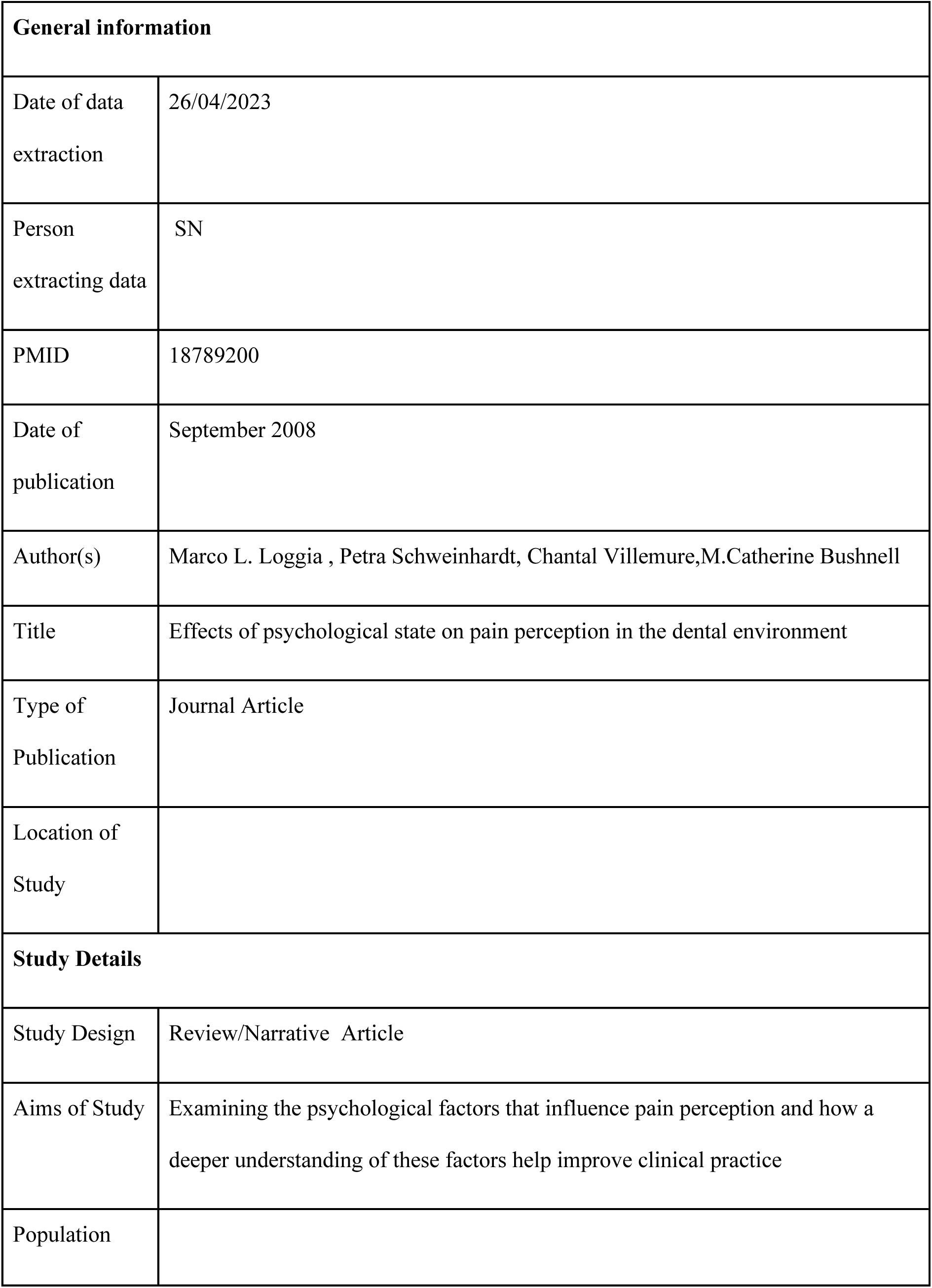
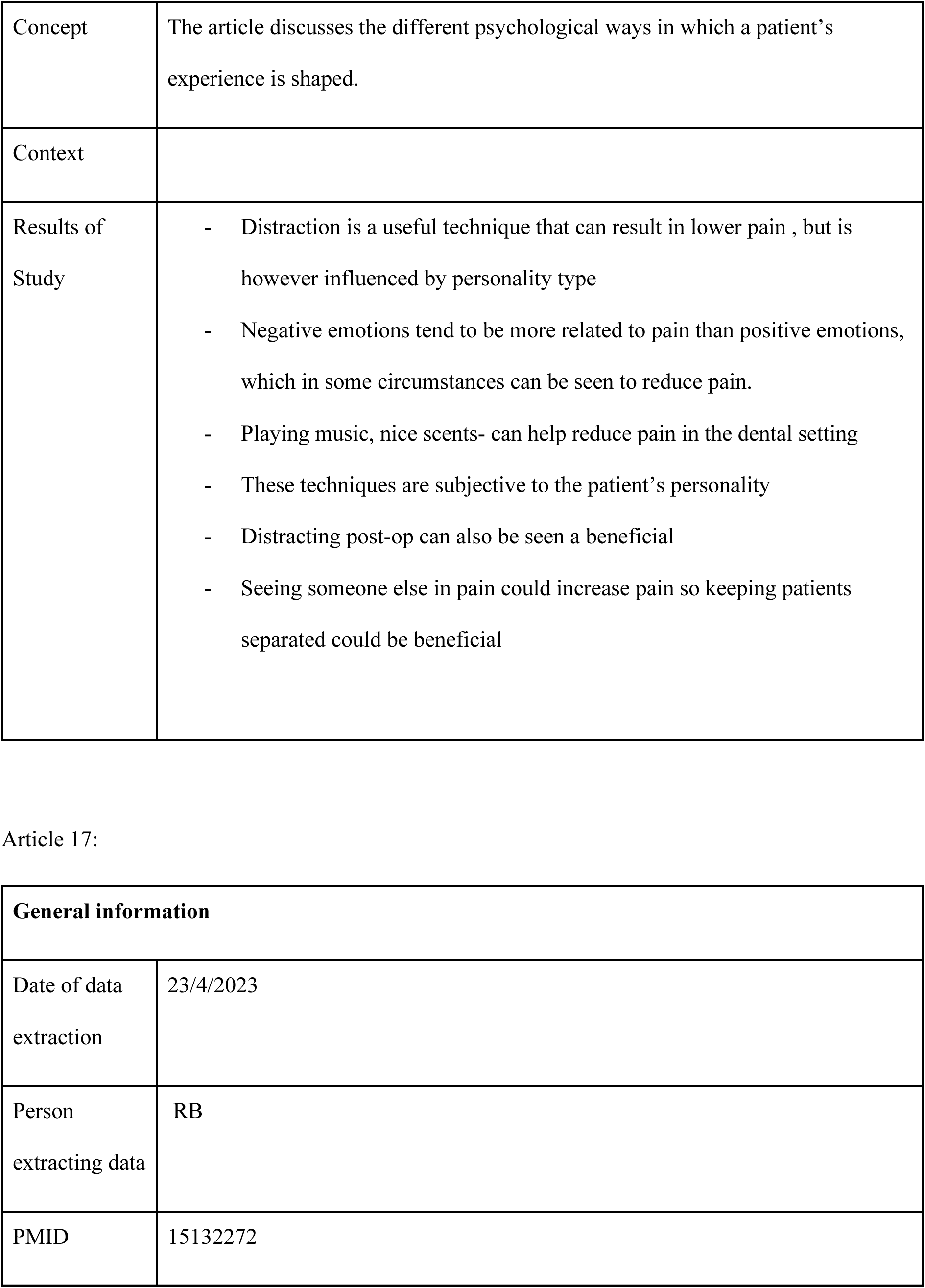
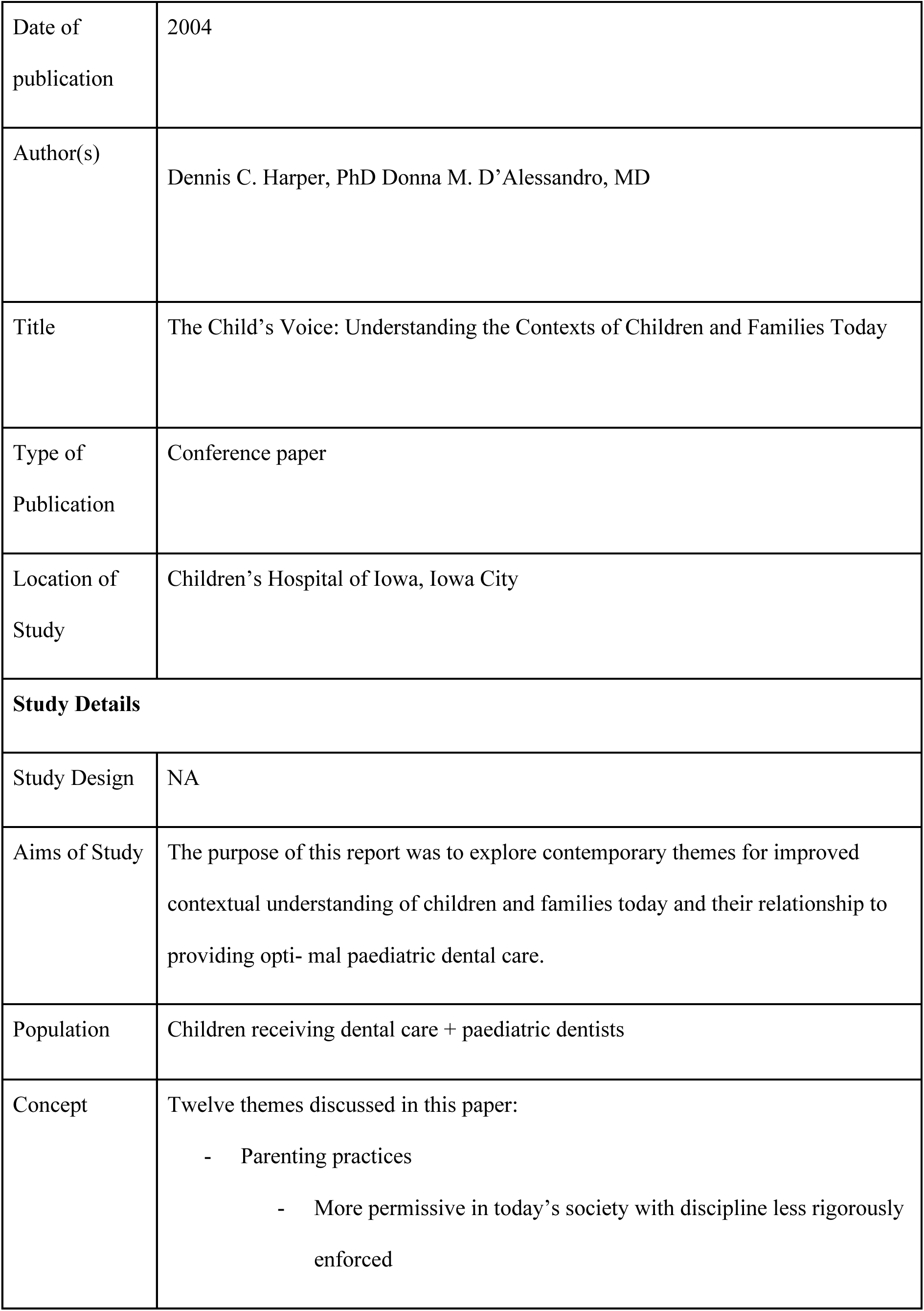
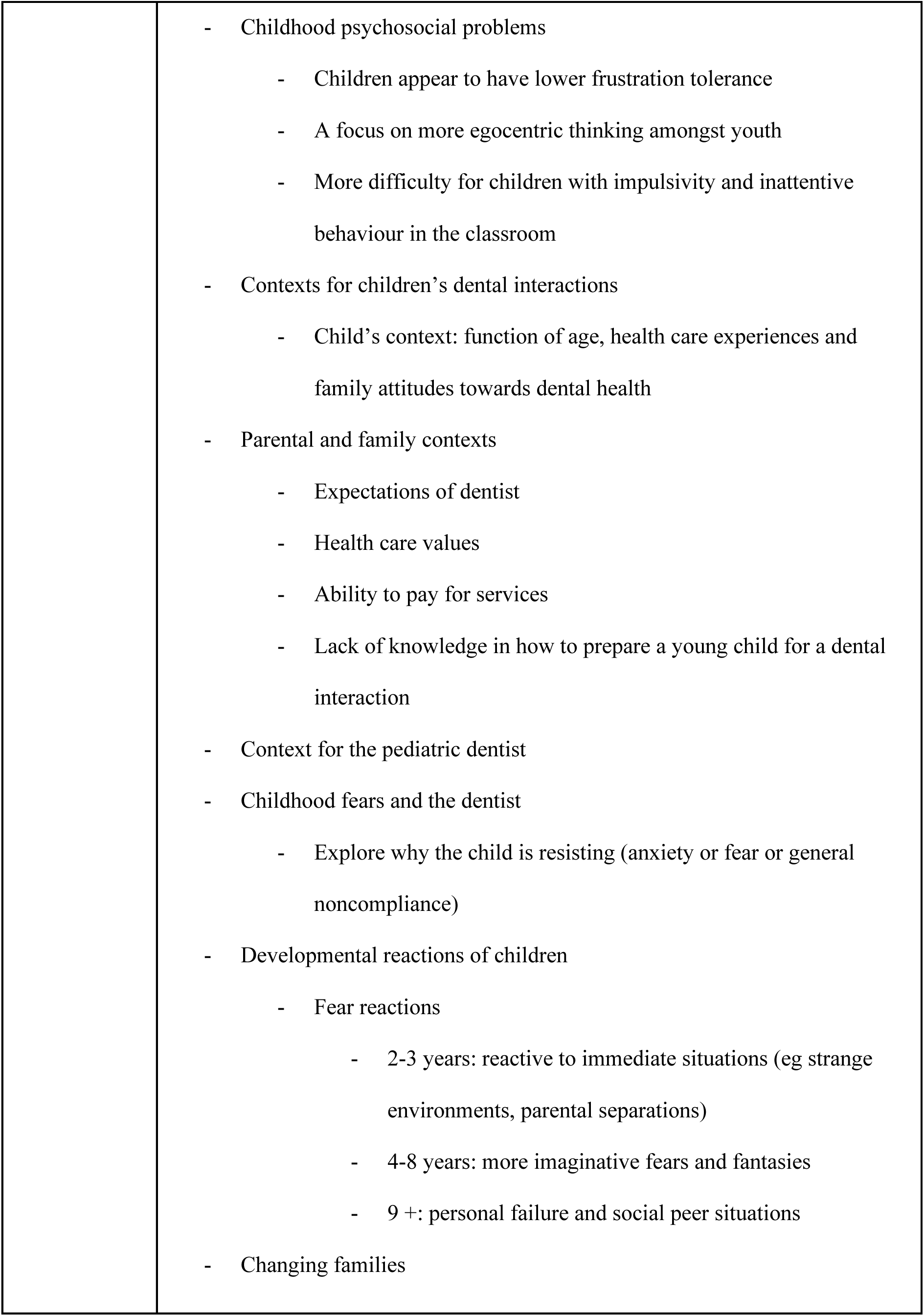
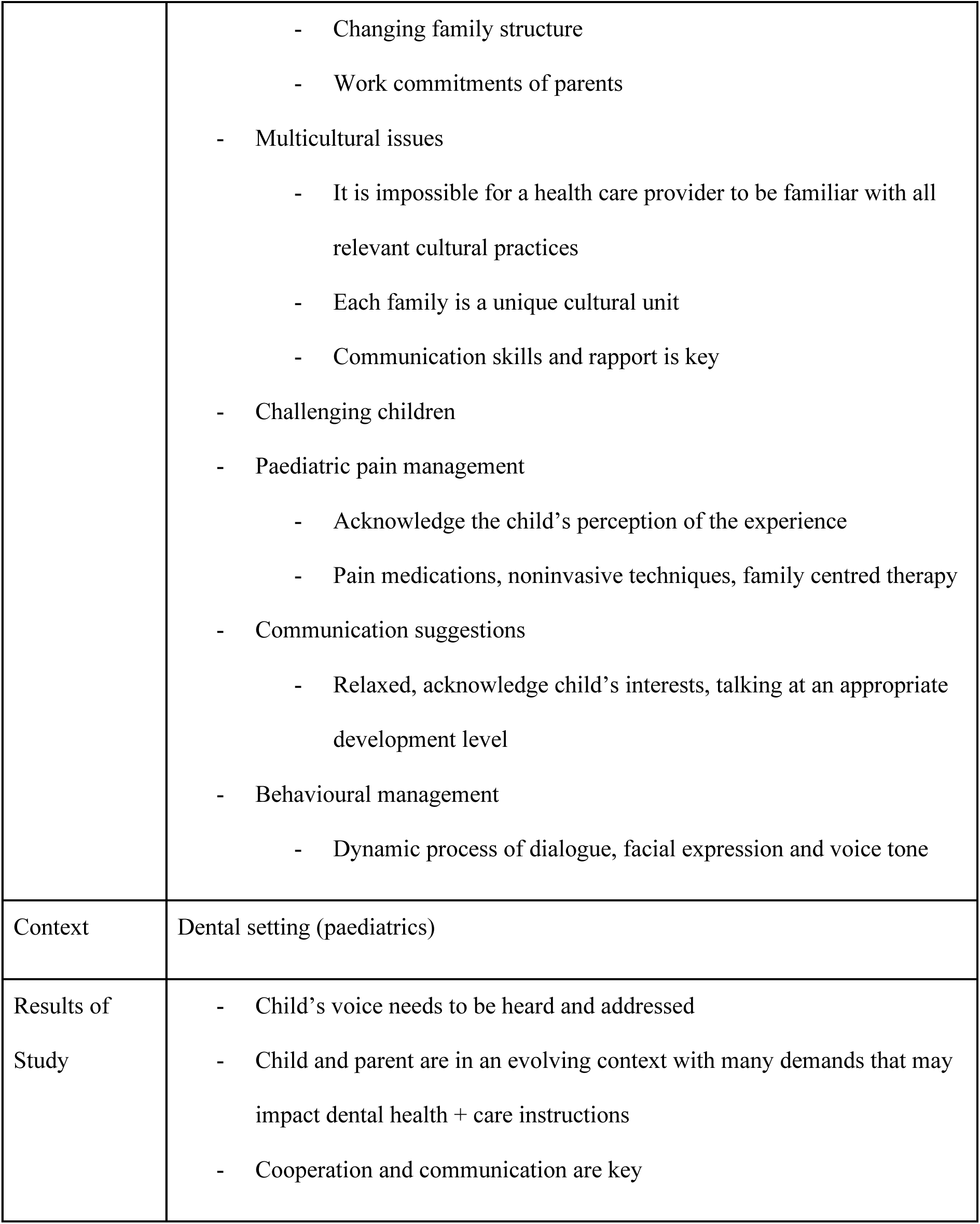
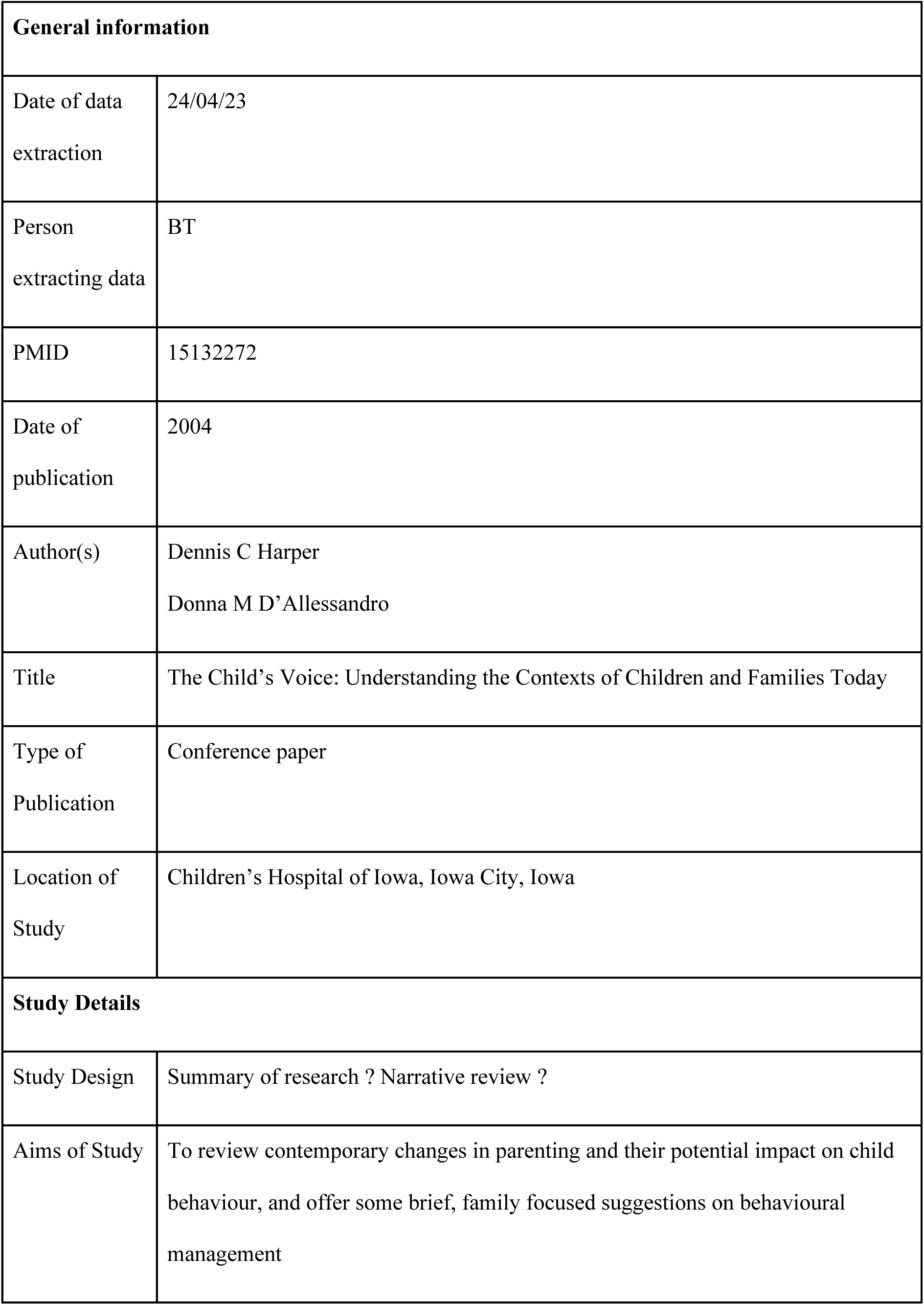
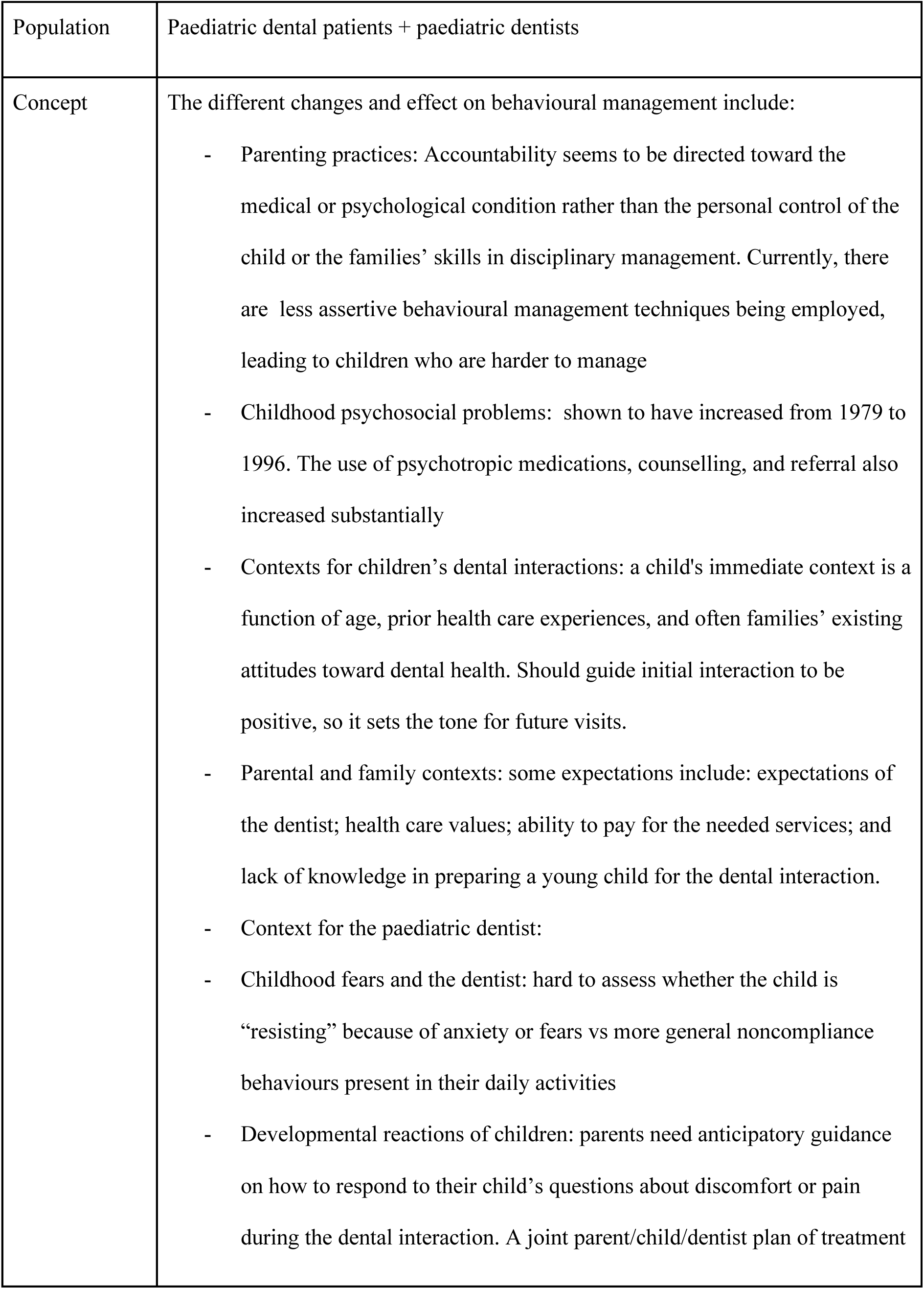
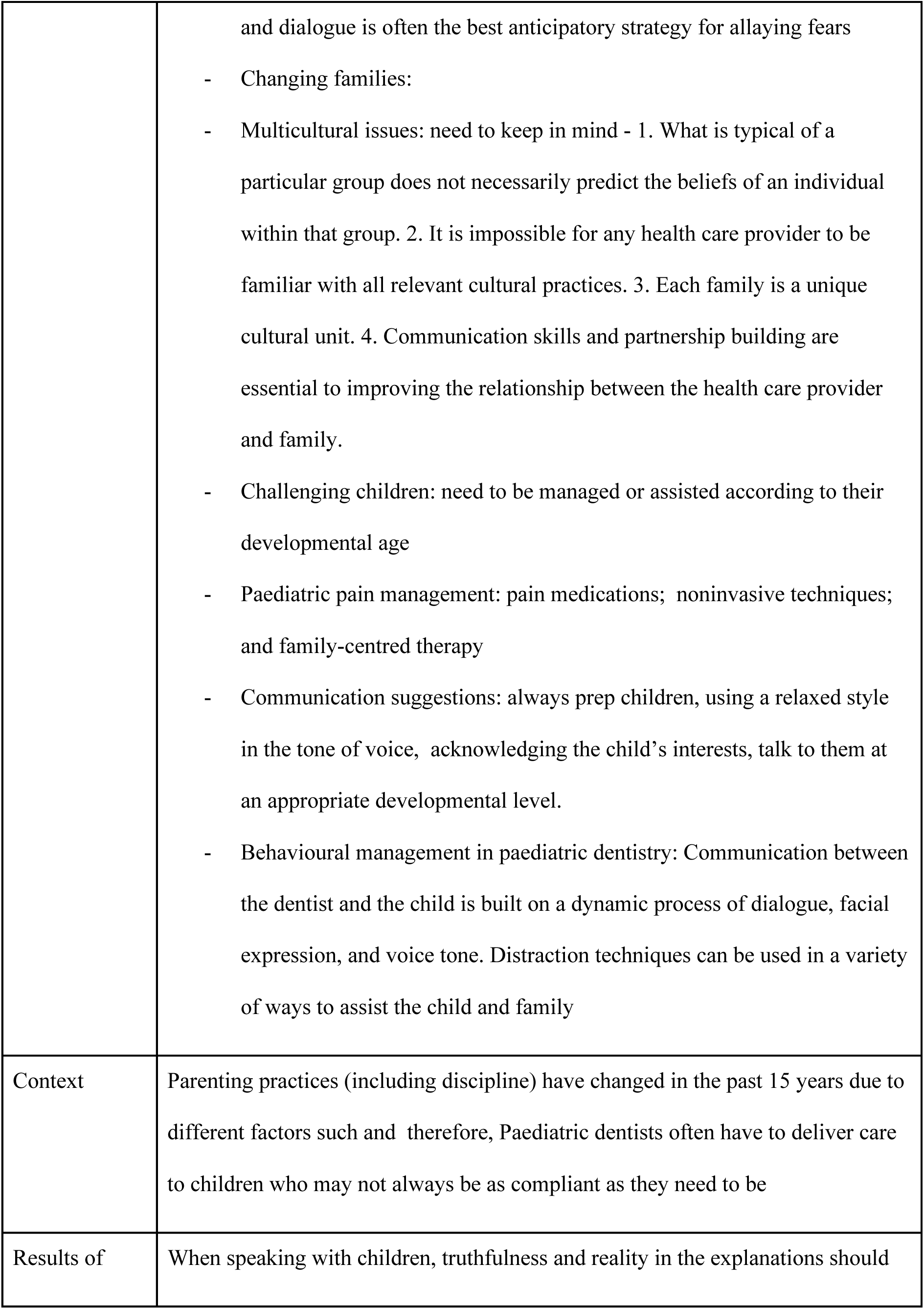
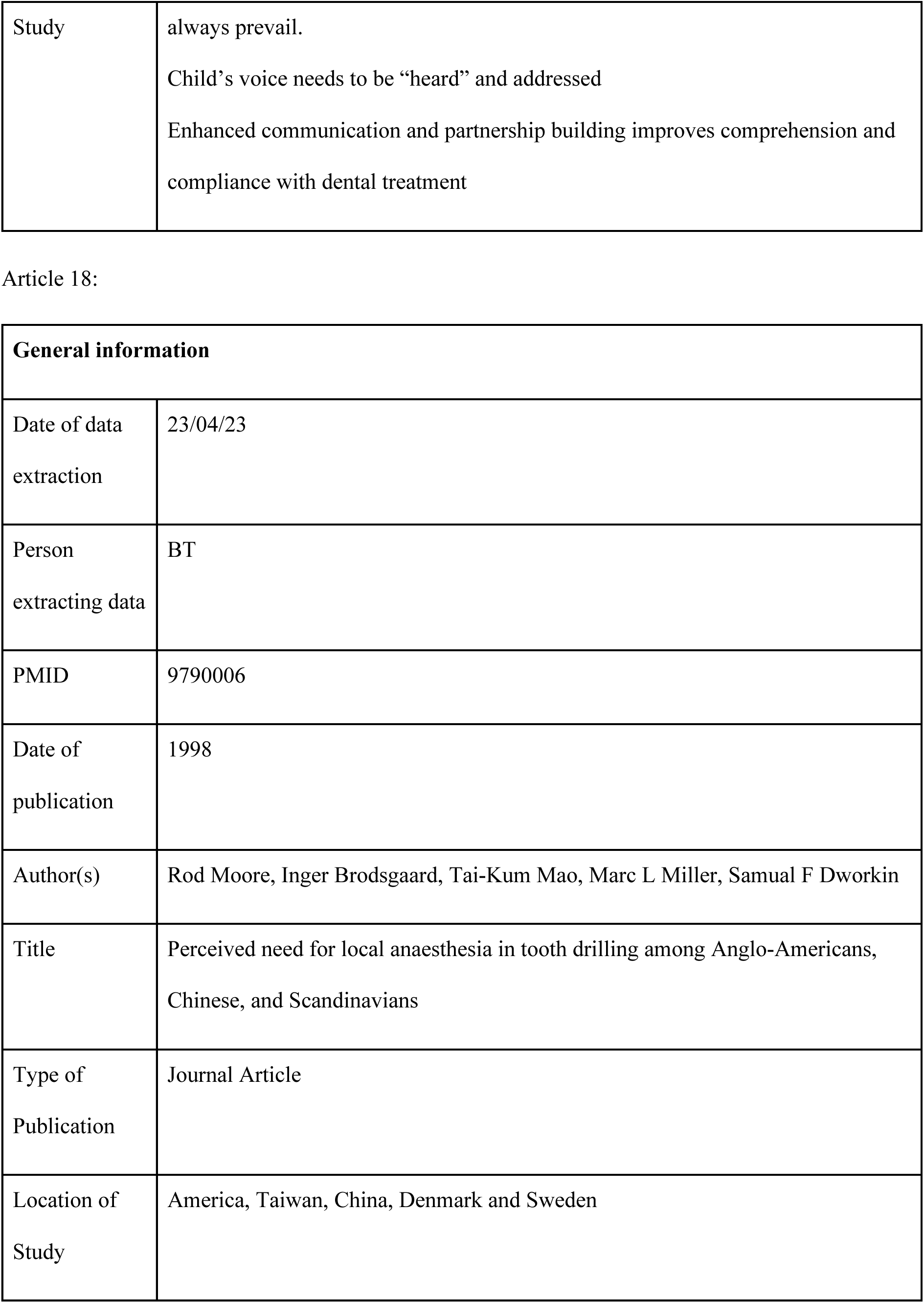
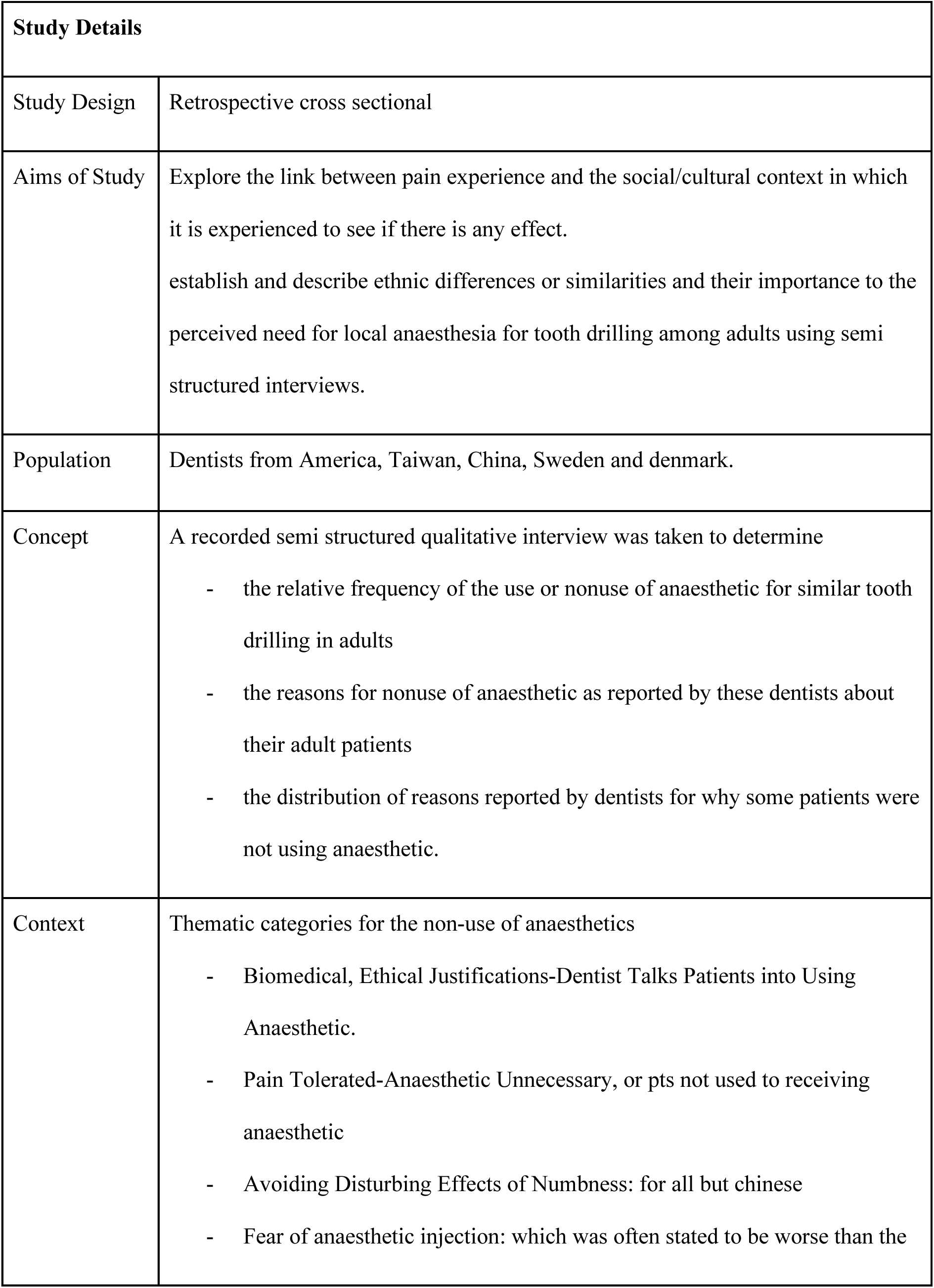
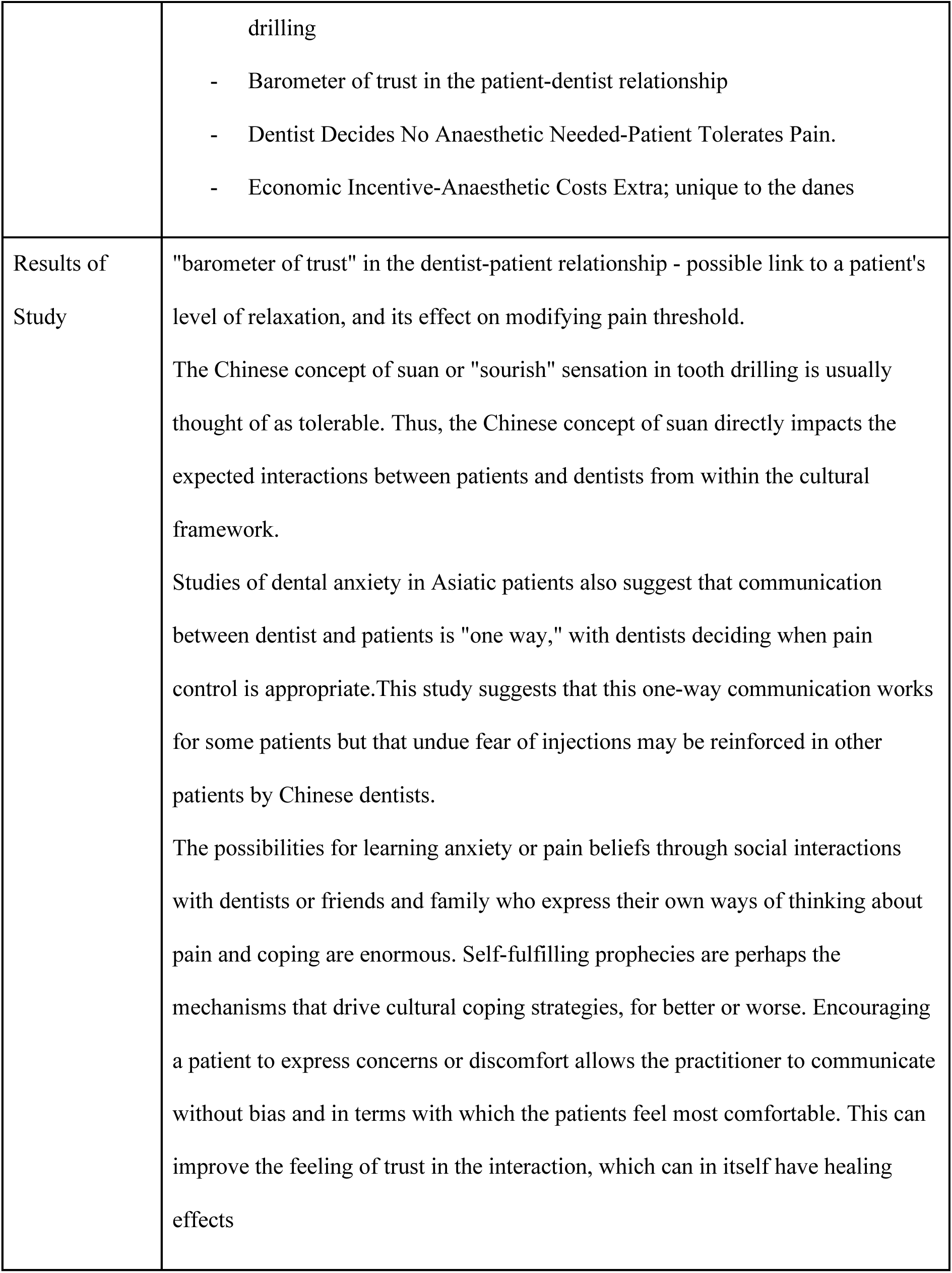
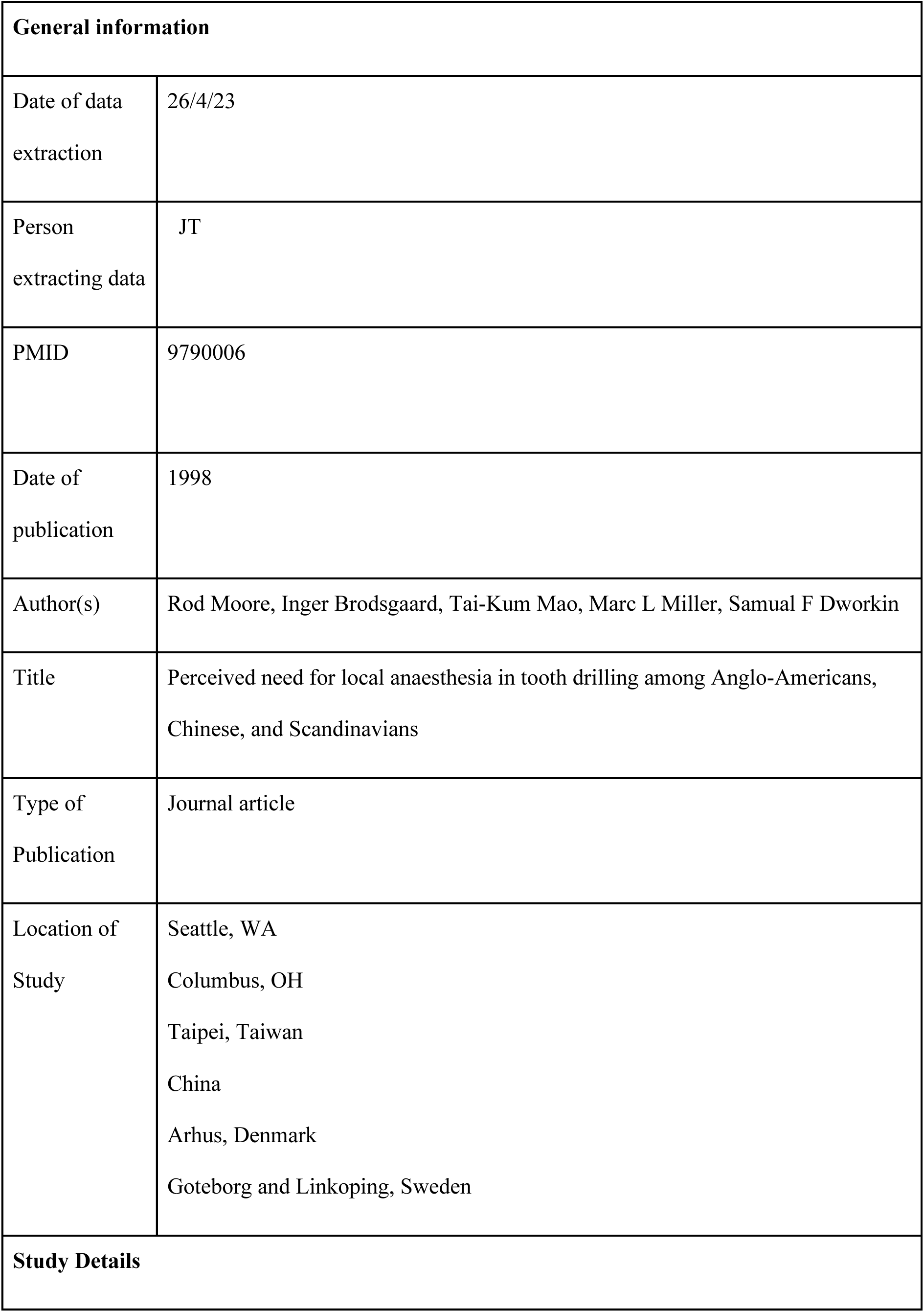
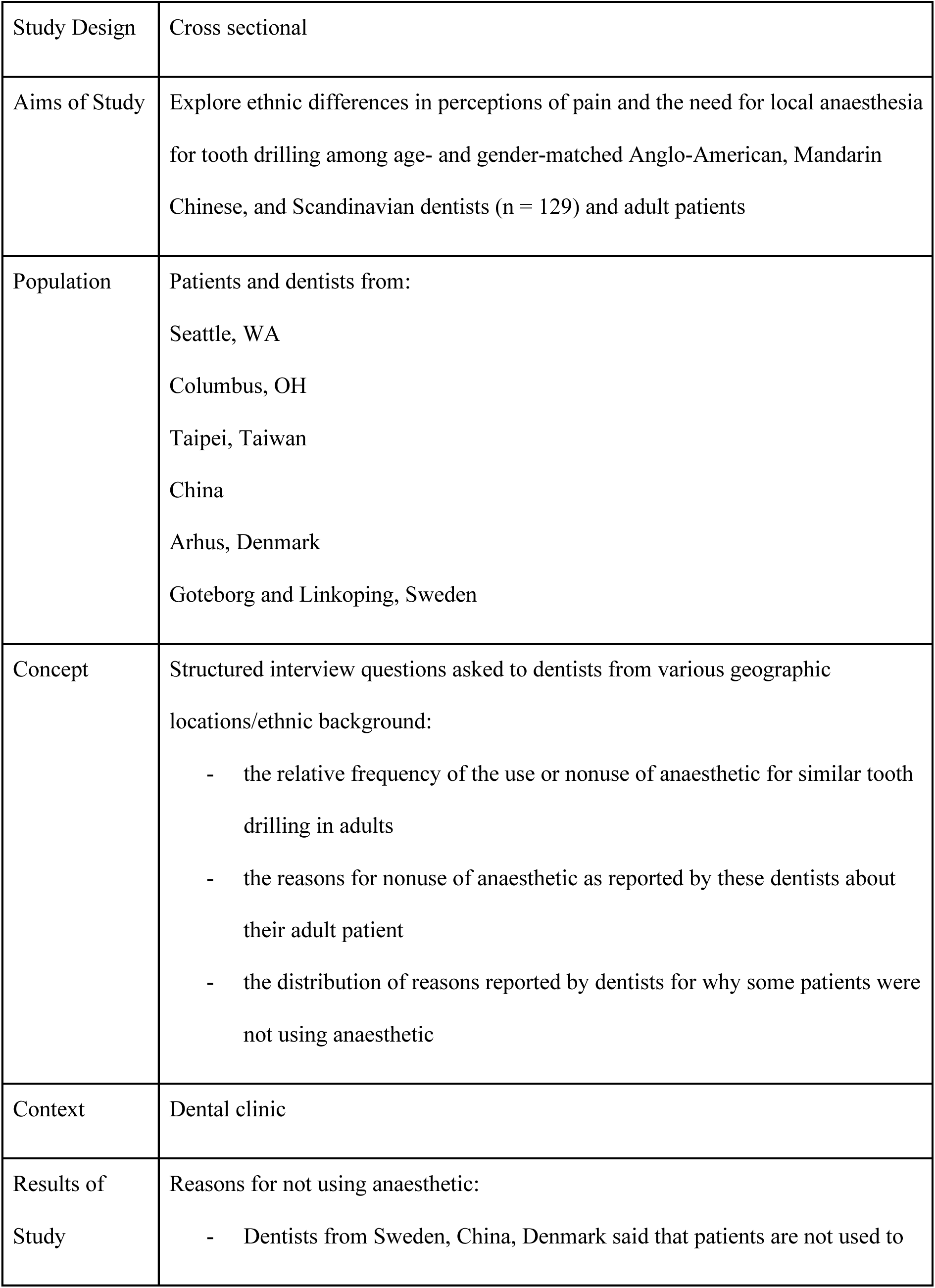
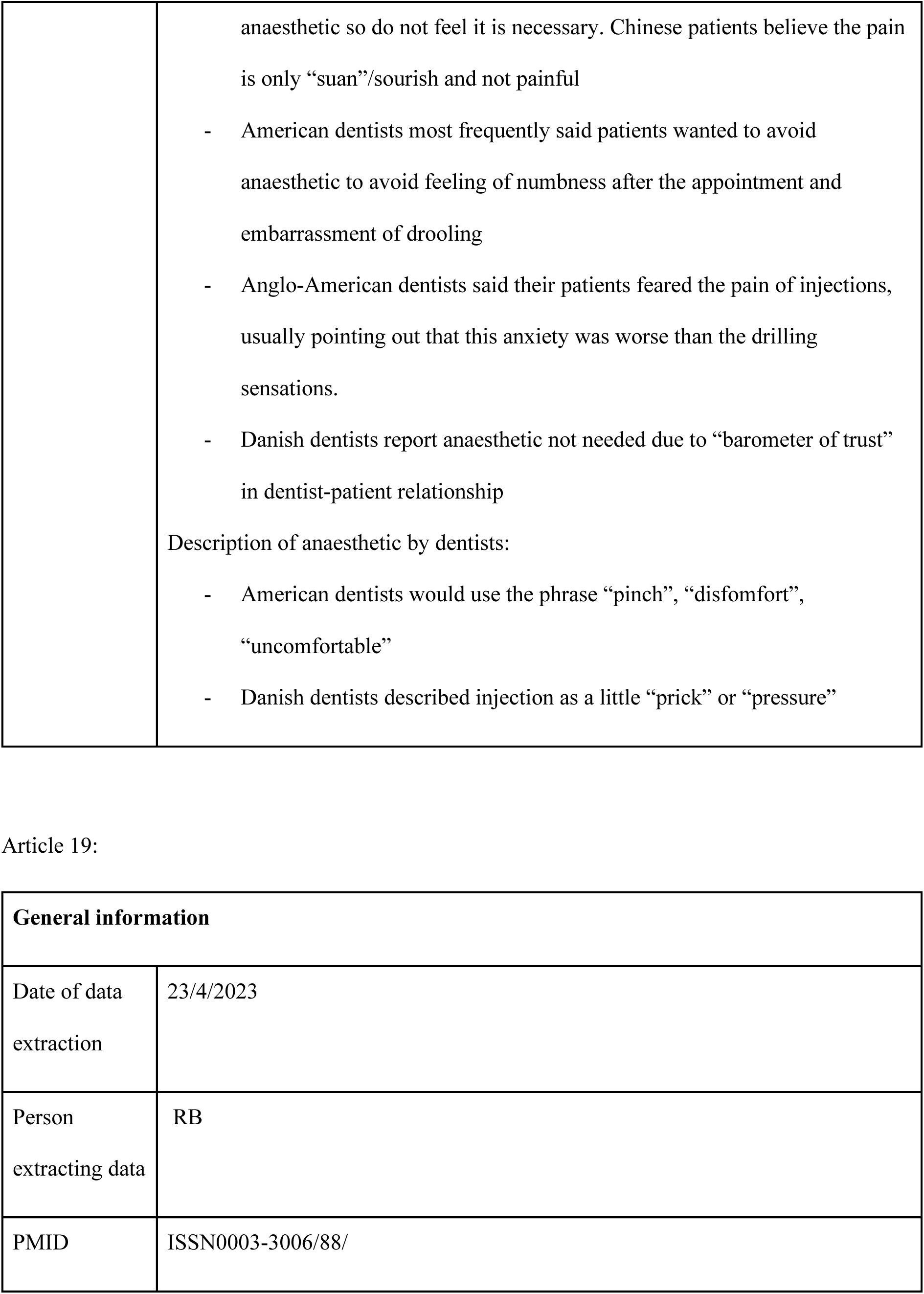
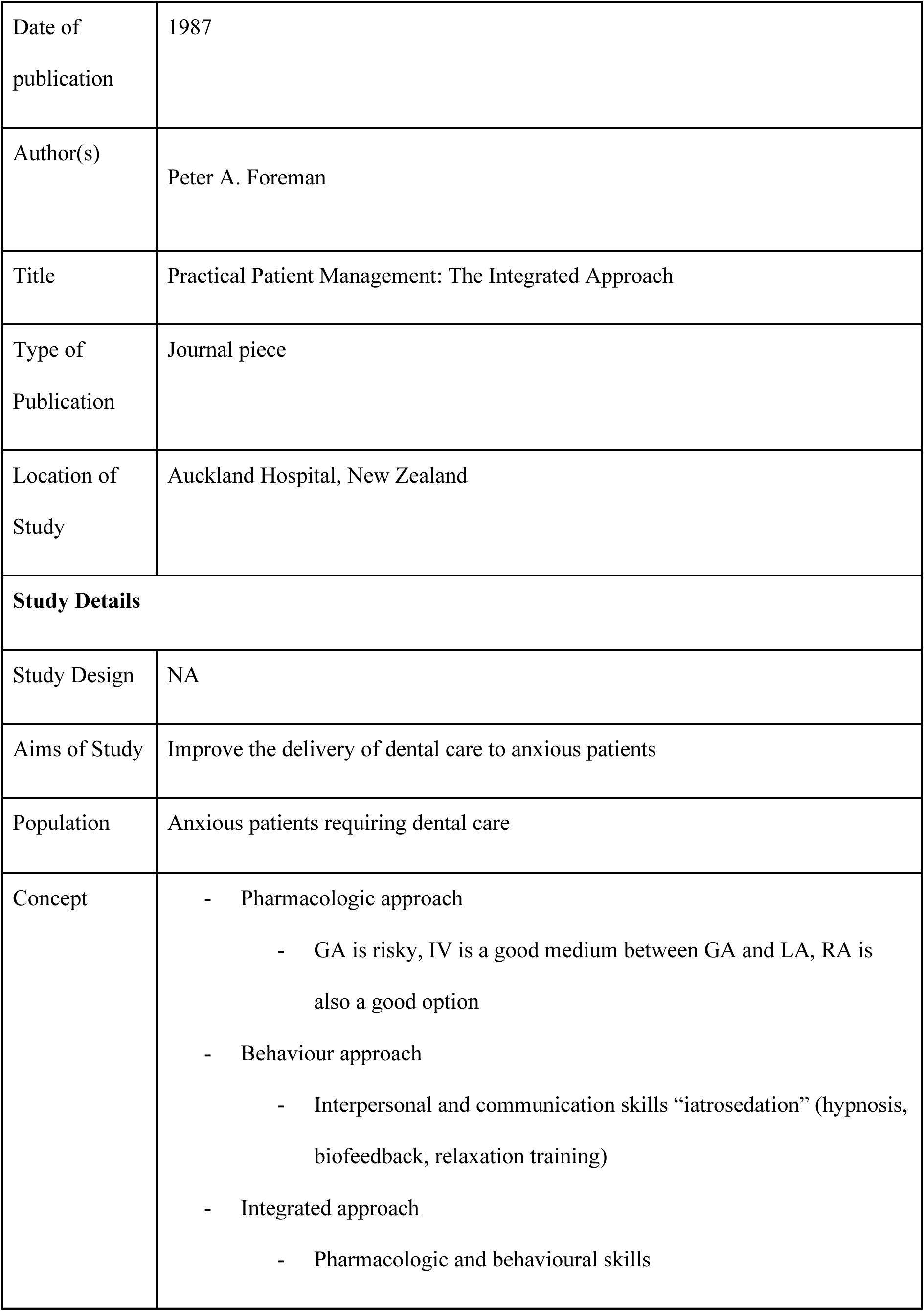
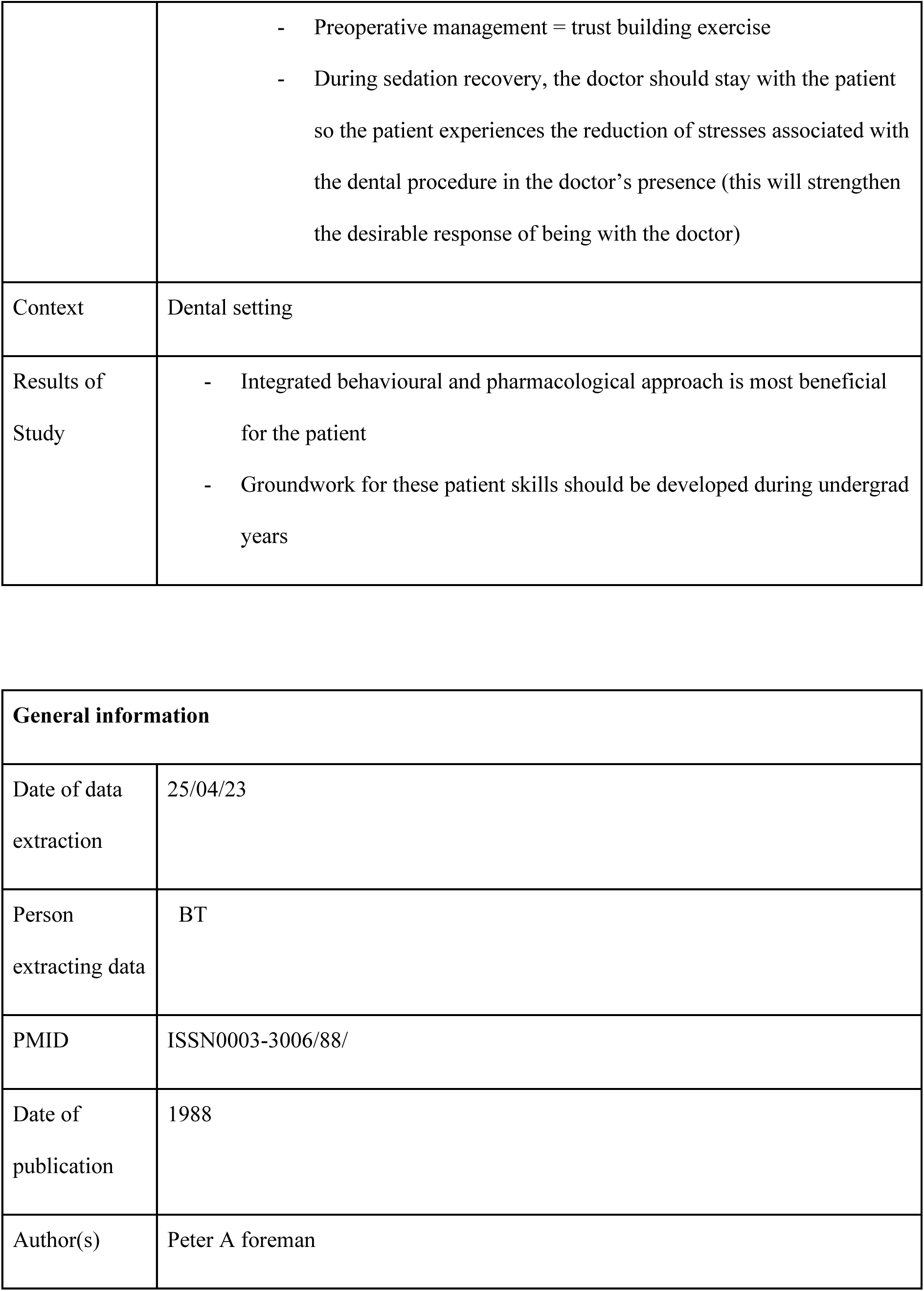
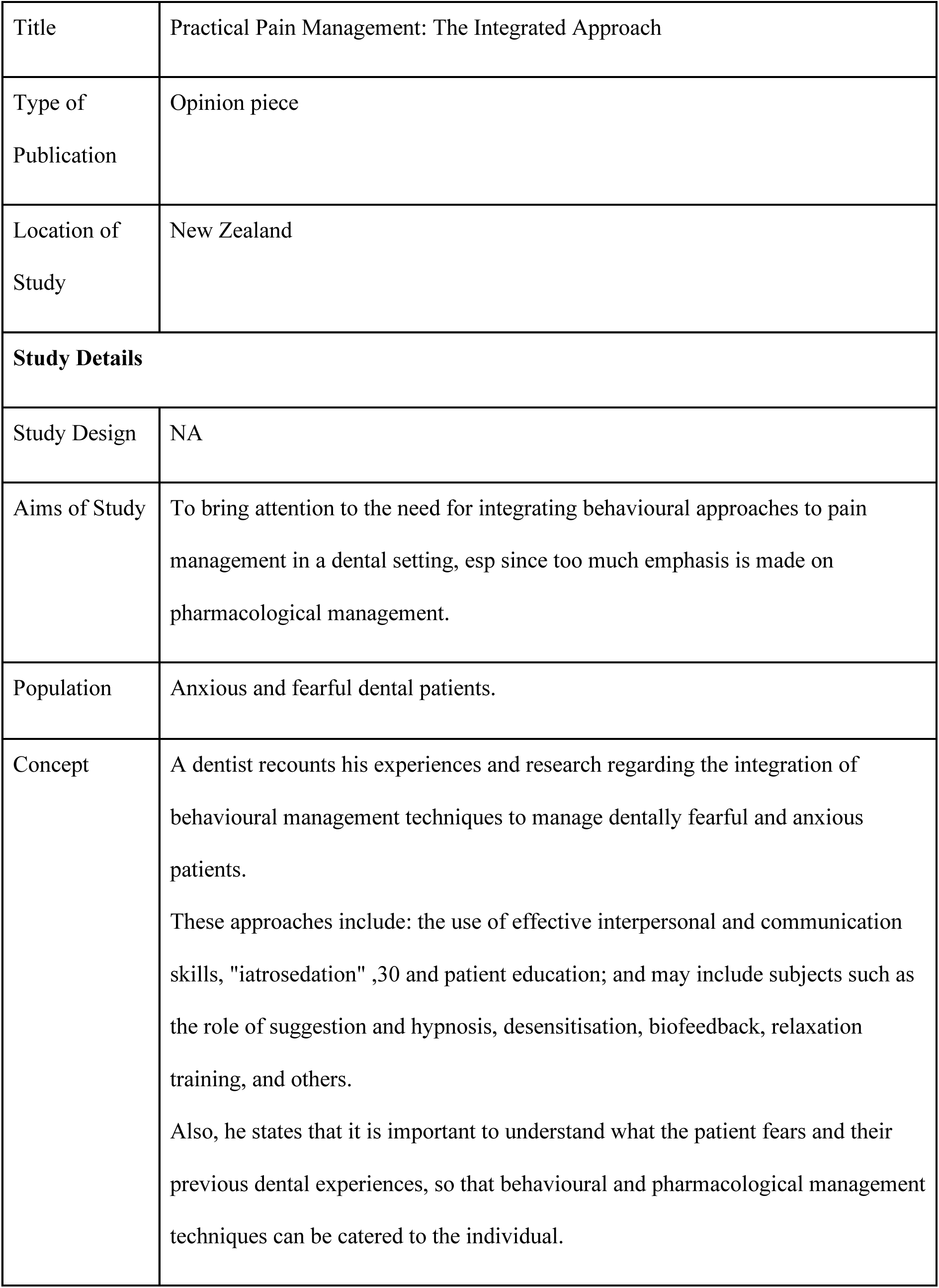
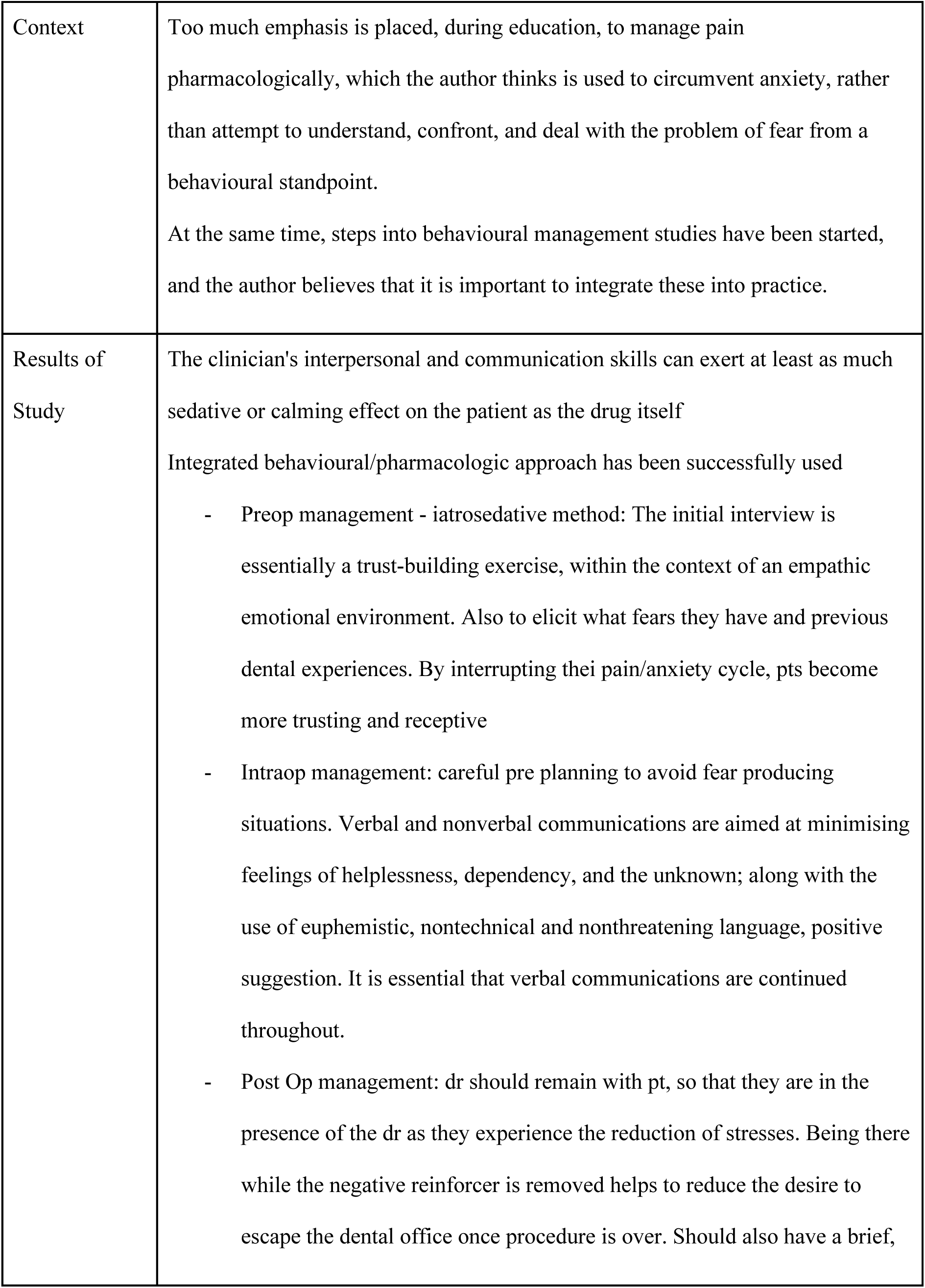
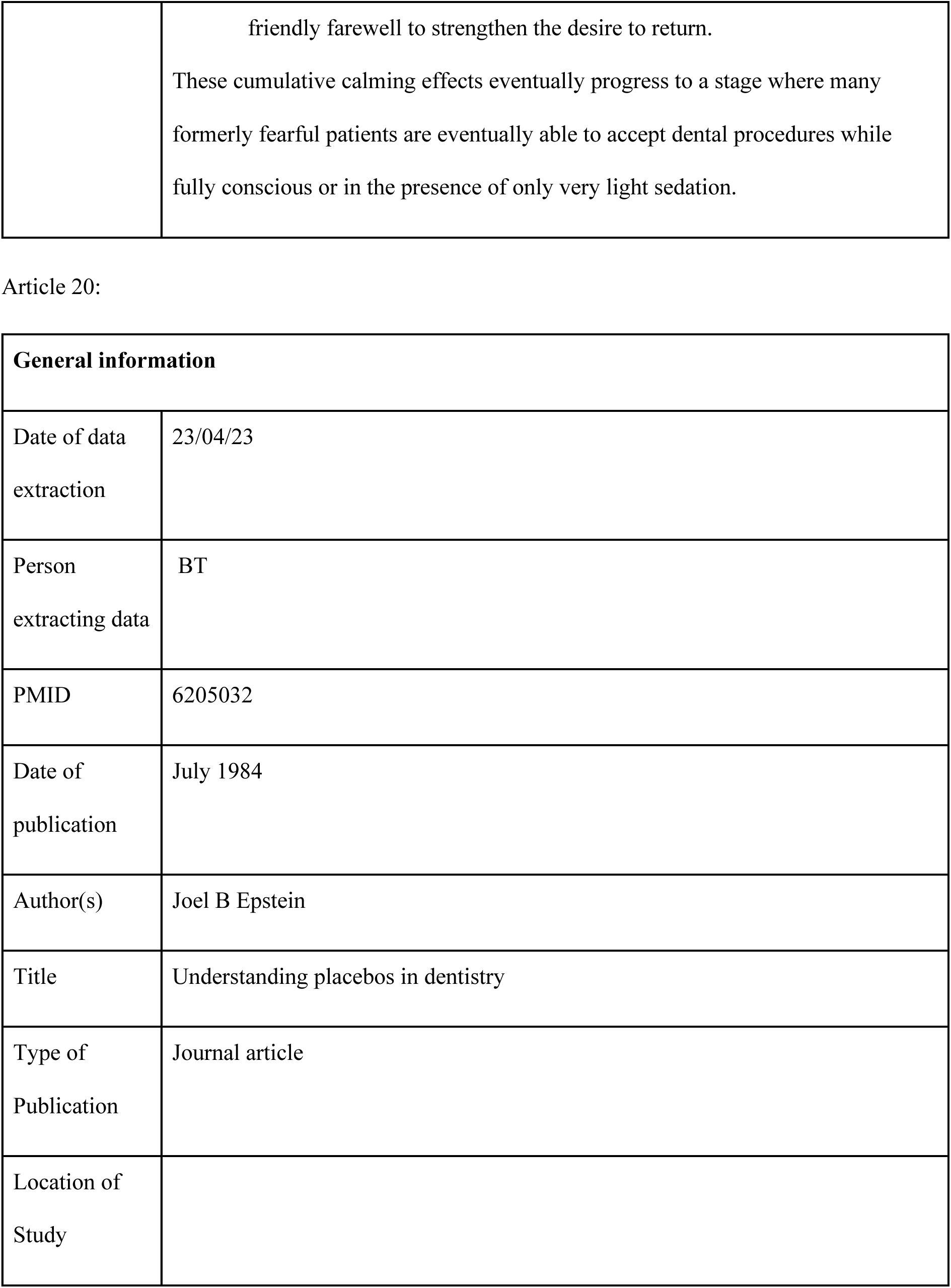
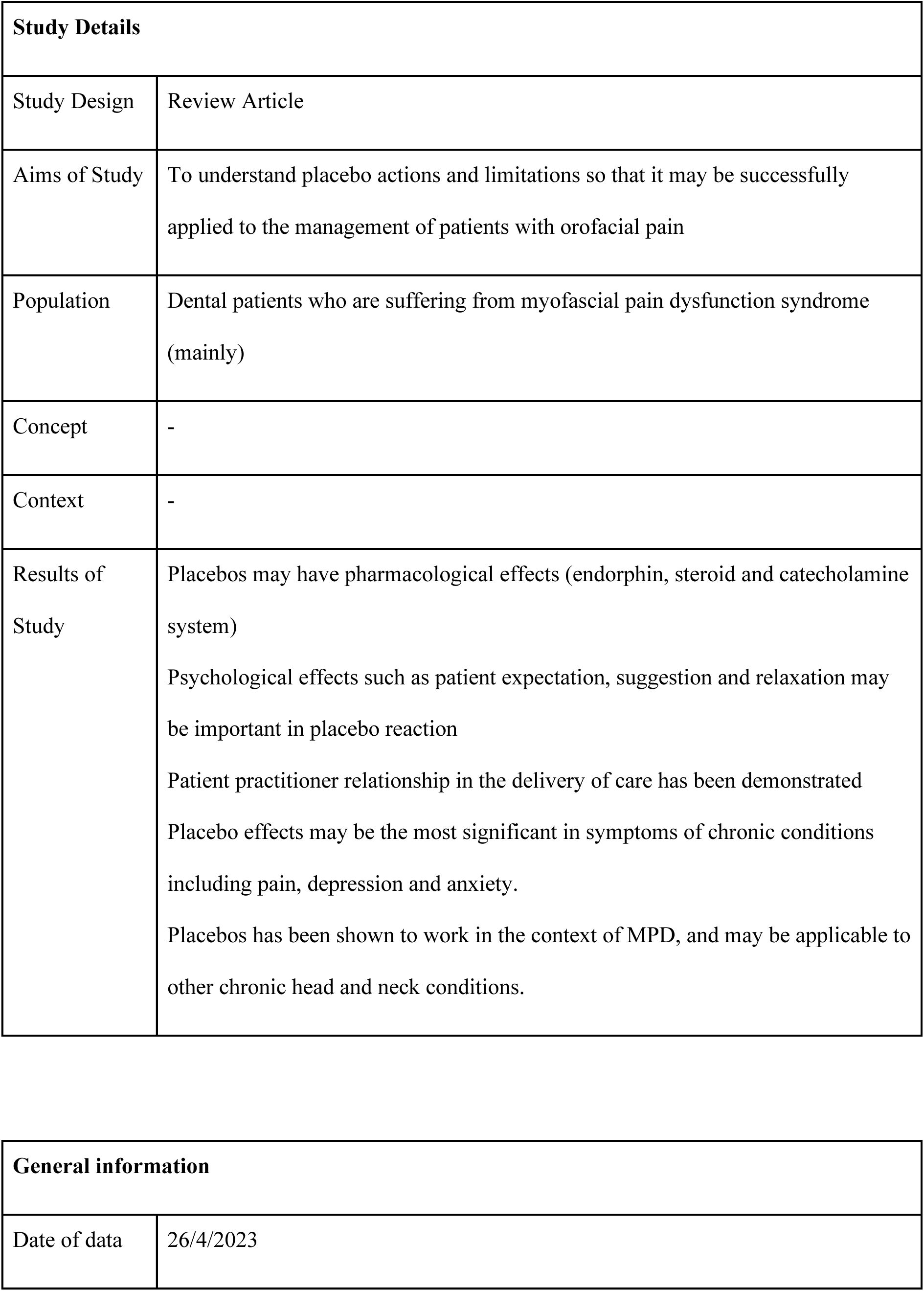
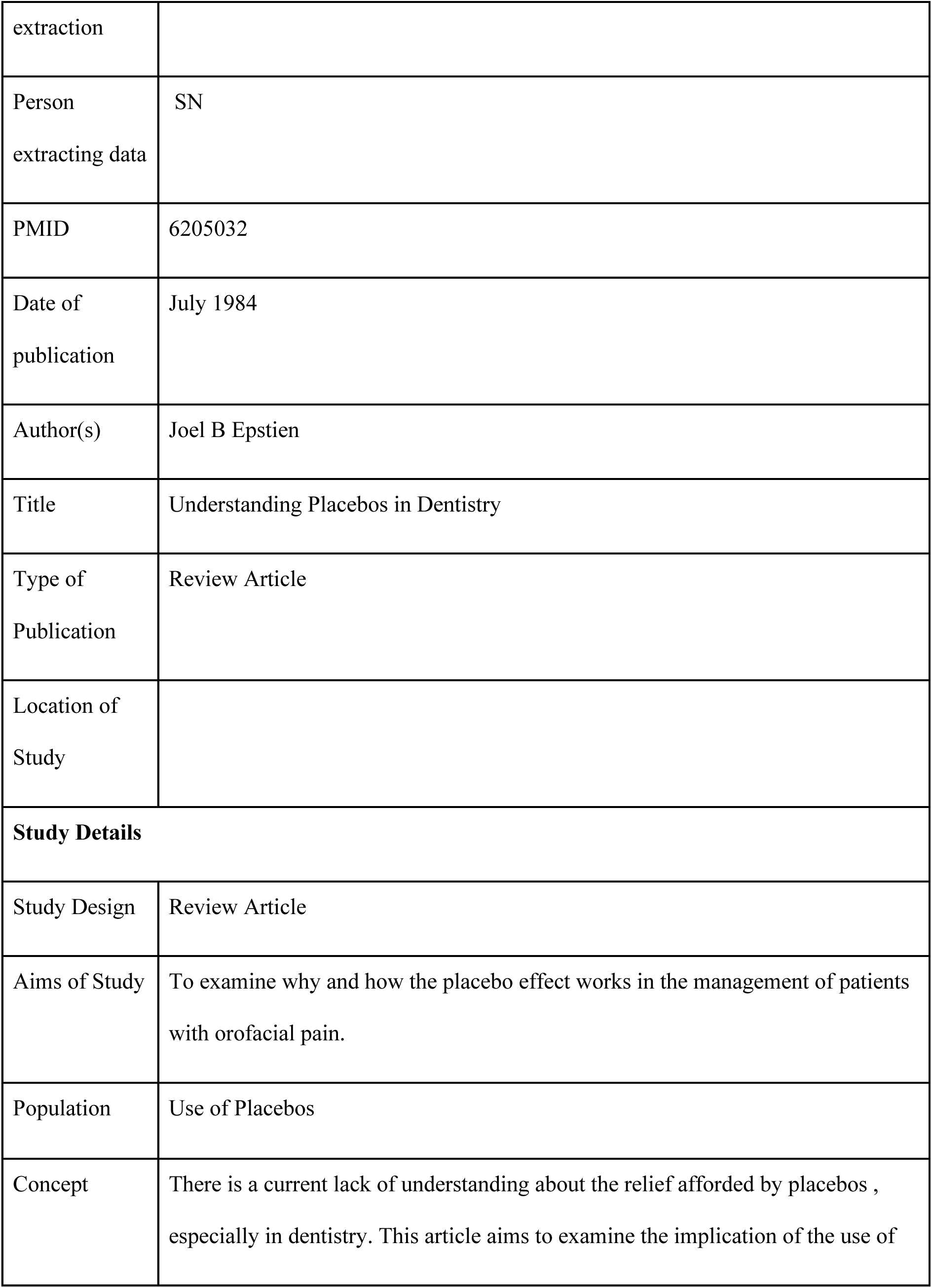
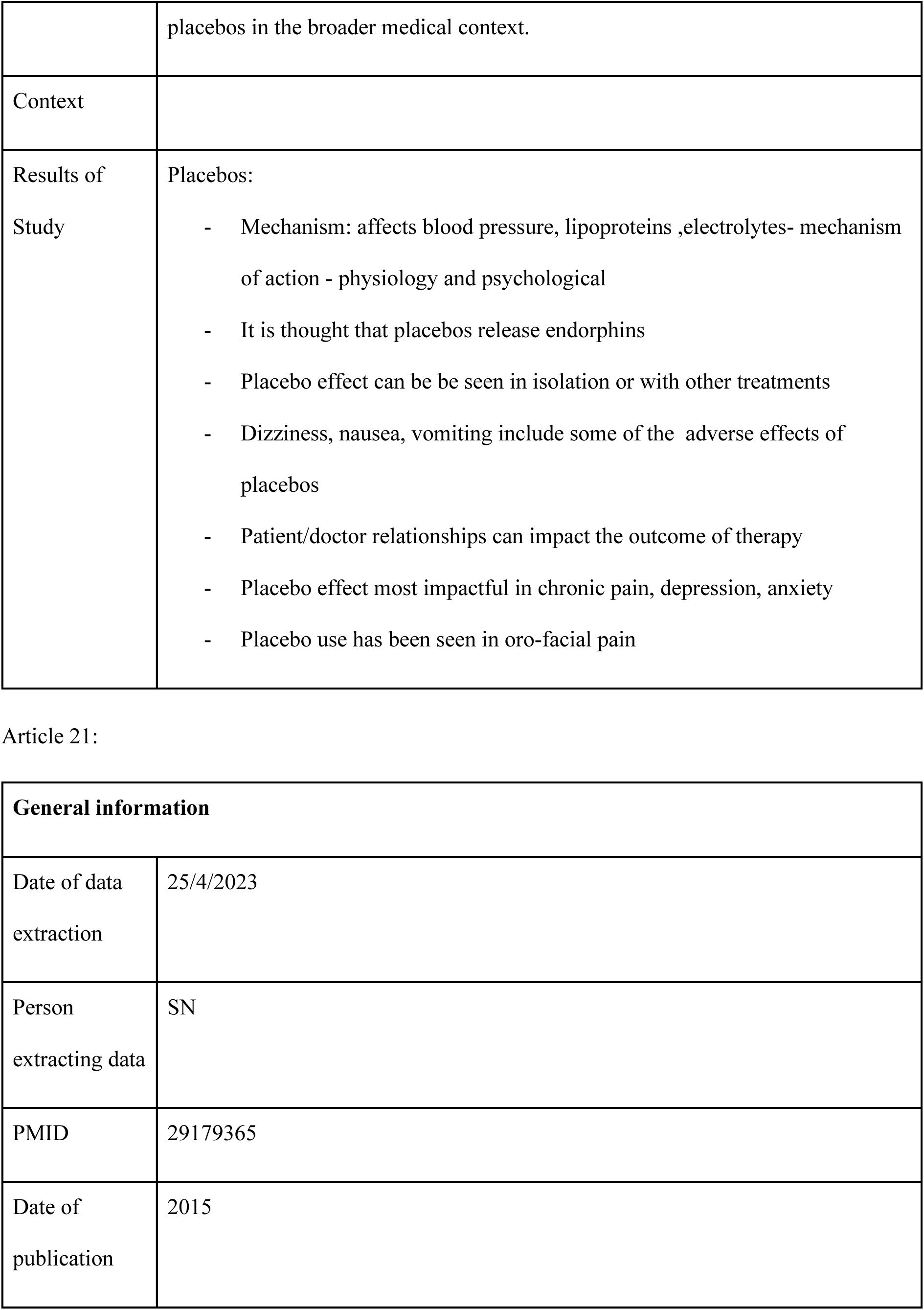
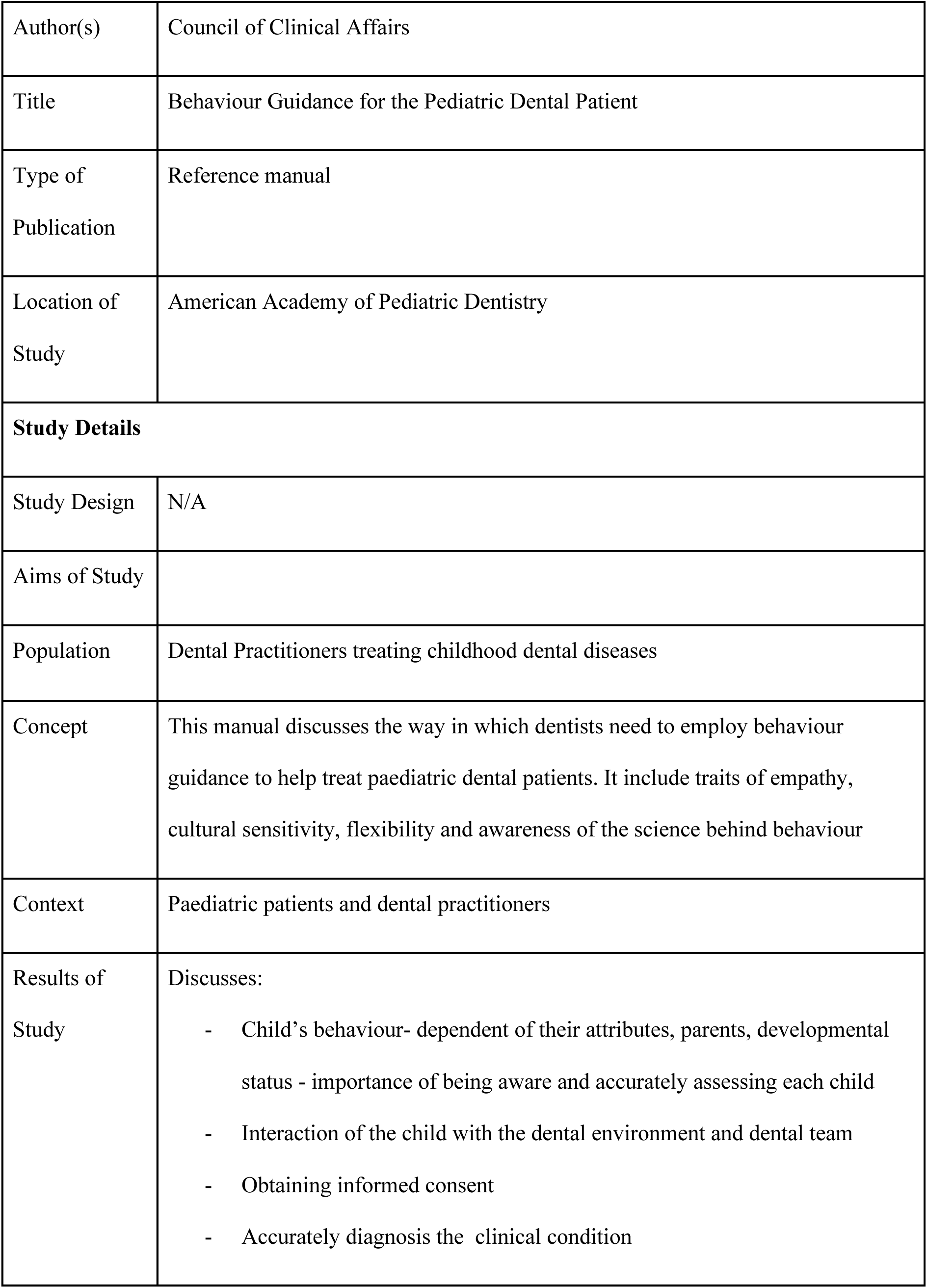
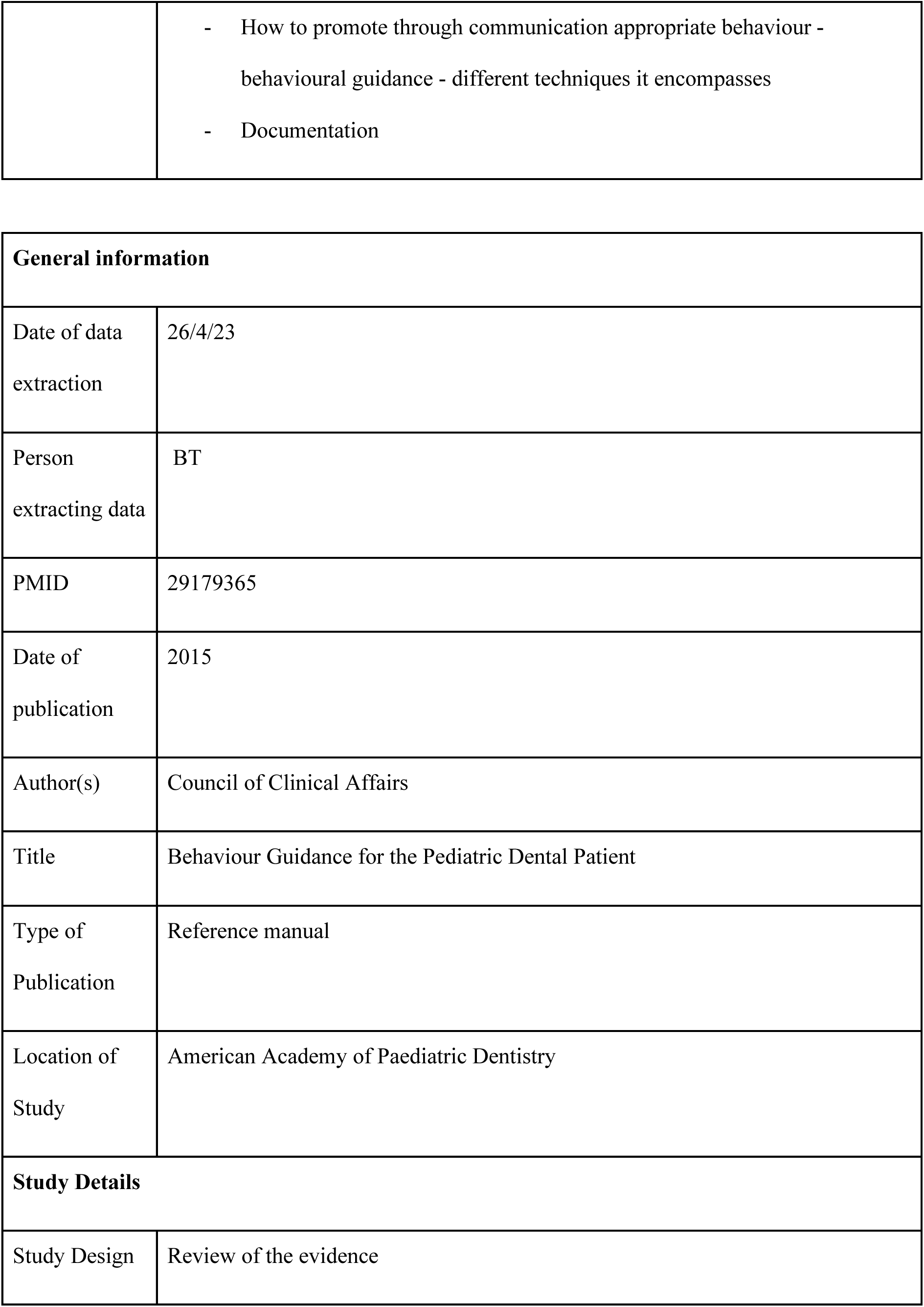
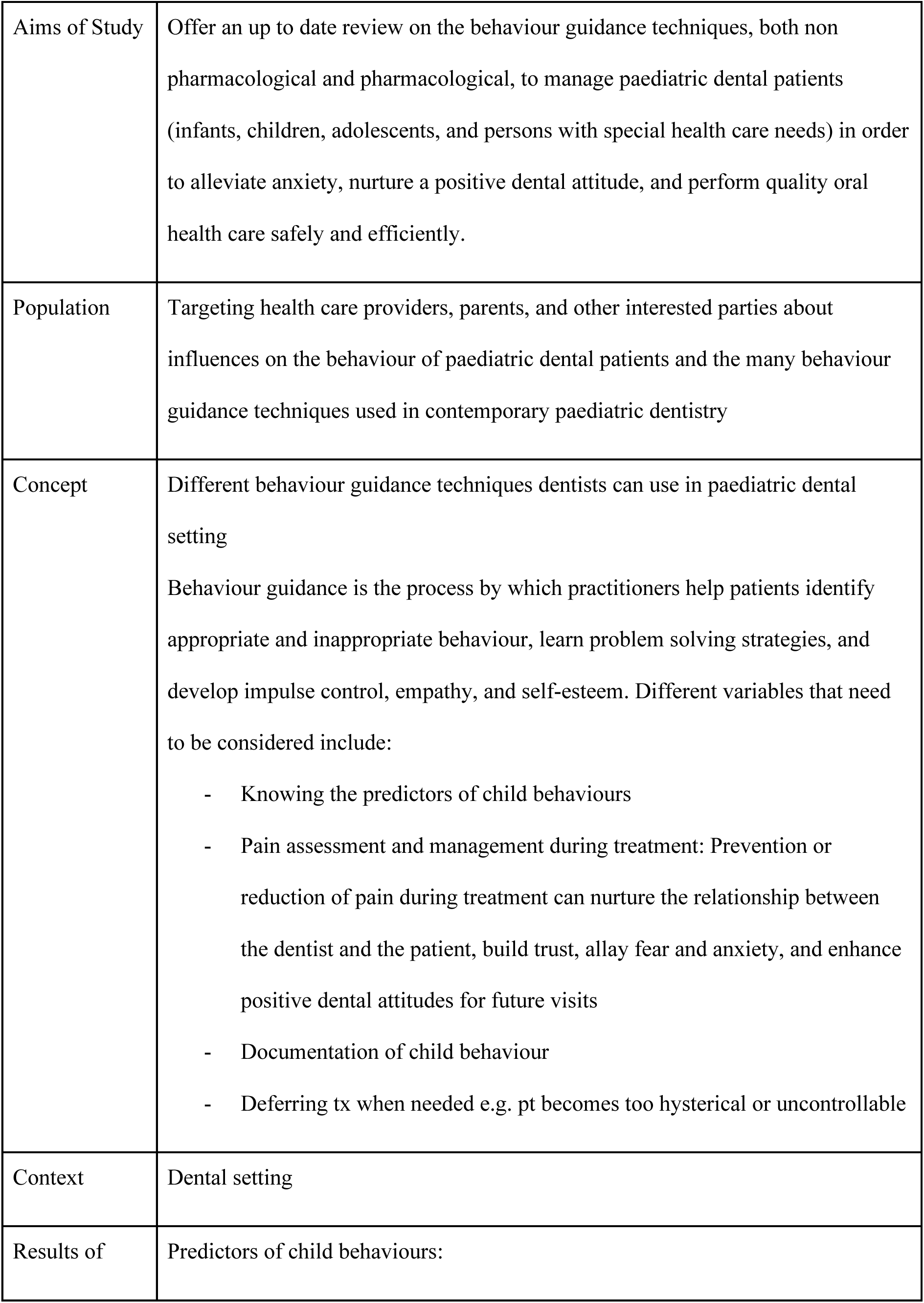
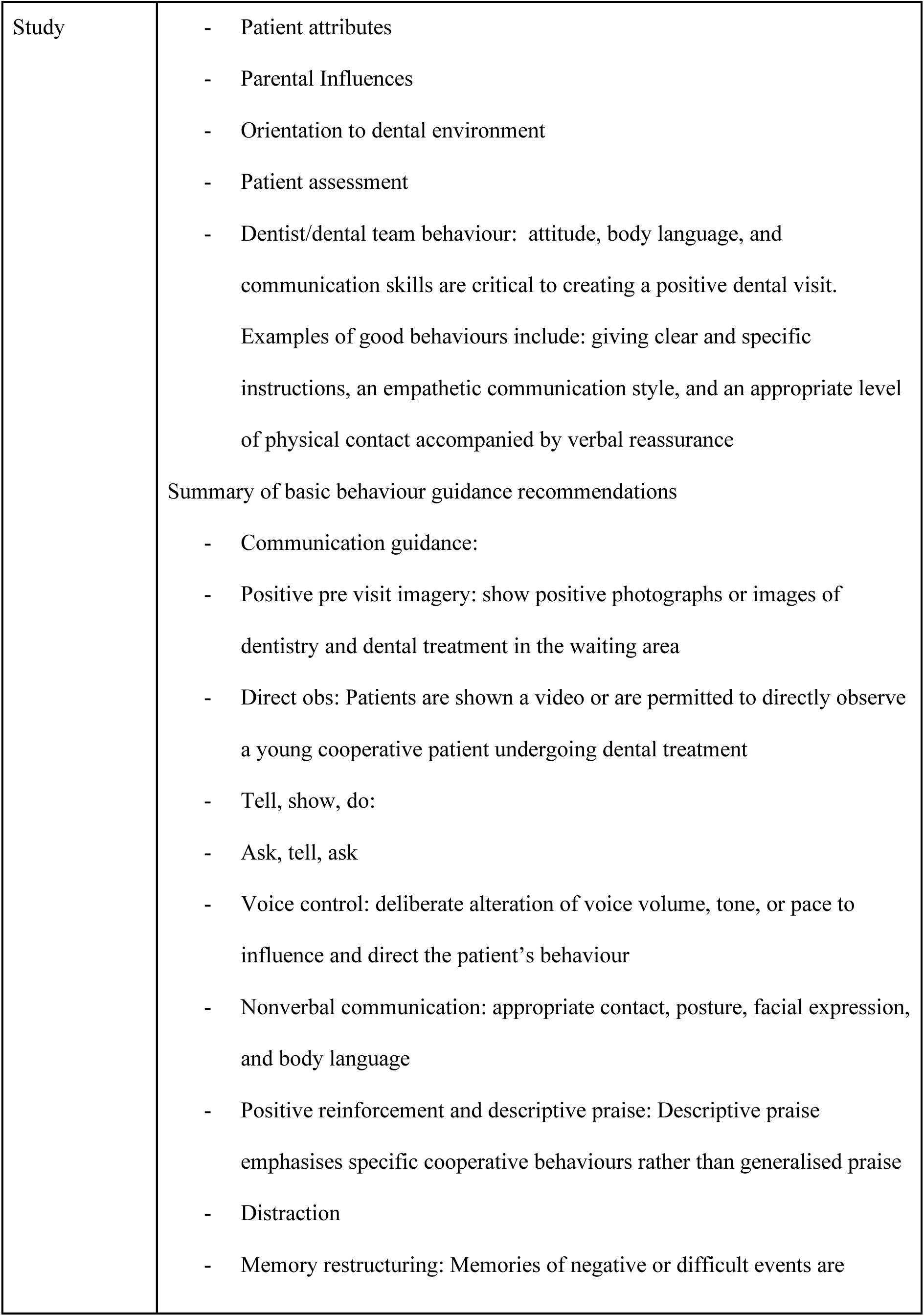
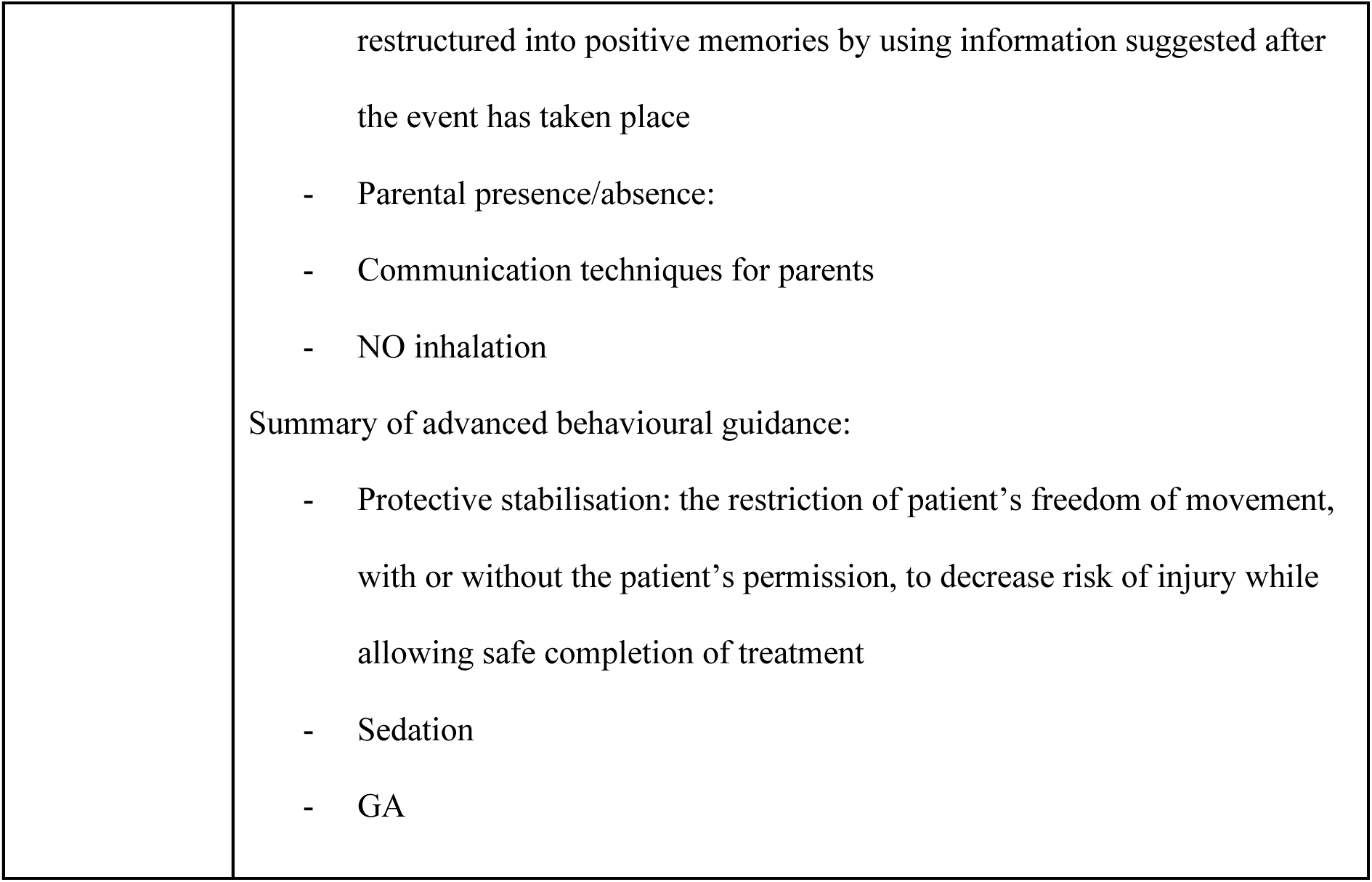
Detailed data extraction

| **General information** |  |
| --- | --- |
| Date of data extraction | 20/4/23 |
| Person extracting data | JT |
| PMID | 97705 |
| Date of publication | 1978 |
| Author(s) | Steven L. Gryll 1 and Martin Katahn |
| Title | Situational Factors Contributing to the Placebo Effect |
| Type of Publication | Journal article |
| Location of Study | Oral Surgery Clinic of Ben Taub General Hospital in Houston, Texas |
| **Study Details** |  |
| Study Design | Randomised control trial |
| Aims of Study | Investigate the influence of status of communicator, attitude of dentist, attitude of dental technician, and message on the obtainment of placebo effects in an oral surgery clinic |
| Population | 160 people from Oral Surgery Clinic of Ben Taub General Hospital, Houston, Texas |
| Concept | Comparing the anxiety and pain levels of patients receiving a placebo anxiolytic pill before having anaesthetic administered by a “warm” vs “neutral” dentist. The information given about the placebo pill to patients was either positive (“oversell” group), negative (“undersell” group) or neutral (“saliva” group) |
| Context | Dental procedure conducted at Oral Surgery Clinic of Ben Taub General Hospital, Houston, Texas |
| Results of Study | Patients had less reported anxiety when treated by a “warm” dentist or dental technician and lower pain was reported after anaesthesia administration.<br>“Oversell” message (dentists said the placebo would be effective in reducing anxiety and pain) led to a positive placebo effect. When a “warm” dentist presented an “undersell” message (the dentist said they believed the placebo was not that good at reducing anxiety and pain) it also led to a positive placebo effect. |

| General information |  |
| --- | --- |
| Date of data extraction | 21/4/2023 |
| Person | RB |
| extracting data |  |
| PMID | 97705 |
| Date of publication | 1978 |
| Author(s) | Steven L. Gryll 1 and Martin Katahn |
| Title | Situational Factors Contributing to the Placebo Effect |
| Type of Publication | Journal article |
| Location of Study | Texas, USA |
| **Study Details** |  |
| Study Design | RCT |
| Aims of Study | The influence of four variables (status of communicator of drug effects, attitude of dentist, attitude of dental technician, and message of drug effects) on the obtainment of placebo effects in an oral surgery clinic was investigated |
| Population | 160 adults (18 years +). 53% males, mean age 33 years |
| Concept | Rating of pre/post op anxiety, pre/post op fear of injection and pain experienced from IANB following administration of a placebo pill with the following variables:<br>- Individual administering pill (dentist/technician) |
|  | - Attitude of dentist (warm or neutral)<br>- Attitude of technician (warm or neutral)<br>- Message given to patient (no pill, overselling, underselling) |
| Context | Dental setting (Oral Surgery Clinic of Ben Taub General Hospital, Houston, Texas) |
| Results of Study | - Overselling = least pain + reduced anxiety and fear of injection post-placebo<br>- Undersell = also reduced anxiety<br>- Bias because the dentist/technician administering the pill knew it was a placebo so may have covertly affected their behaviour<br>- Lower pain reported with warm/friendly clinicians = friendly attitude may potentiate placebo effect |

Article 2:
| General information |  |
| --- | --- |
| Date of data extraction | 20/4/23 |
| Person extracting data | JT |
| PMID | 16214624 |
| Date of publication | 2005 |
| Author(s) | Bartlett, B. W.; Firestone, A. R.; Vig, K. W.; Beck, F. M.; Marucha, P. T. |
| Title | The influence of a structured telephone call on orthodontic pain and anxiety |
| Type of Publication | Journal article |
| Location of Study | Ohio State University graduate orthodontic clinic |
| **Study Details** |  |
| Study Design | Randomised control trial |
| Aims of Study | Examine the effects of a structured telephone call after orthodontic appliance placement on self-reported pain and anxiety |
| Population | 250 orthodontic patients |
| Concept | Assess impact of a structured telephone call demonstrating care and assurance vs attention-only telephone call on the anxiety and pain levels of patients after placement of orthodontic appliances |
| Context | Dental pain after orthodontics |
| Results of Study | All subjects reported pain one week after orthodontic appliance placement.<br>Patients who received a structured phone call and attention-only phone call reported significantly less pain than the control group (no phone call). The structured phone call led to slightly less reported pain and anxiety compared to attention-only phone call. |
| Date of data extraction | 22/04/23 |
| Person extracting data | BT |
| PMID | 16214624 |
| Date of publication | 2005 |
| Author(s) | Bartlett, B. W.; Firestone, A. R.; Vig, K. W.; Beck, F. M.; Marucha, P. T. |
| Title | The influence of a structured telephone call on orthodontic pain and anxiety |
| Type of Publication | Journal article |
| Location of Study | Ohio State University Graduate Orthodontic Clinic |
| Study Details |  |
| Study Design | RCT |
| Aims of Study | Examine the effects of a structured telephone call after orthodontic appliance placement on self-reported pain and anxiety |
| Population | 150 patients treated at the Ohio State University graduate orthodontic clinic between sep 2002 and may 2003.<br>Inclusion criteria:<br>- Ortho tx with fixed upper and lower appliances<br>- 10 year old or older<br>- No previous ortho tx<br>- No diagnosed psychological disorders<br>- Baseline pain intensity ratings lower than 5mm on 100 mm VAS |
| Concept | 3 test conditions: first 15 randomly assigned, then subsequent subjects deliberately assigned to match age, sex and ethnicity.<br>- Structured phone call with care and reassurance:<br>- Patient well being<br>- Whether pain and discomfort was present<br>- Reassurance that pt's reaction is wNL<br>- Necessity of sustained OH<br>- Need for soft diet<br>- Use of analgesics<br>- Importance of maintaining a positive attitude.<br>- Attention only phone call thanking them for participating in study, and a brief reminder of how to complete the questionnaire. End there, or redirect pt to call the assigned resident if there are questions.<br>- Control: no phone call<br>Outcome: pre treatment questionnaire to measure baseline pain and anxiety, then |
|  | everyday after ortho tx for 1 week.<br>- Pain: VAS (compared to before)<br>- analgesia consumption: number of pain tablets taken (no dosage or type)<br>- anxiety level: spielberger's state-trait anxiety inventory (40 statements involving a 4 point likert scale) compared to state and trait anxiety at baseline. |
| Context | Pain is a common side effect of ortho tx<br>Gate control theory of pain: involves sensory, cognitive, psychological and environmental factors of pain.<br>A large psychological factor associated with pain is anxiety ." strategies to reduce pt anxiety may also diminish subjective perception of pain.<br>Structured telephone calls after painful procedures have been shown to be helpful in oral care delivery to reassure and encourage pts, disseminate info about expected discomfort, reinforce chrome care regimens and reduce pain. |
| Results of Study | Group 1 and 2 reported significantly less pain and anxiety than the control group.<br>- Max mean pain intensity highest at 24hrs for all 3 groups, but pill count did not show sig group differences<br>- Sig reduction in state anxiety during the week after initial ortho tx.<br>- Max state anxiety score highest at 4 hours for 2 telephone groups, and 24 for control.<br>Cannot be determined whether reduced pain led to reduced anxiety, or vice versa.<br>Even if both were reduced by telephone call.<br>Content of the phone call did not make a difference. Just the phone call itself was sufficient to reduce pt's reported pain and anxiety. Maybe a large difference would |
|  | be noticed in a more anxious population.<br>Further study needed to investigate the mechanism by which a telephone call impacts pt wellbeing. |

Article 3:
| General information |  |
| --- | --- |
| Date of data extraction | 20/4/23 |
| Person extracting data | JT |
| PMID | 8120489 |
| Date of publication | 1993 |
| Author(s) | Selden, H. S. |
| Title | Patient empowerment--a strategy for pain management in endodontics |
| Type of Publication | Journal article |
| Location of Study | USA |

| Study Details |  |
| --- | --- |
| Study Design | Retrospective study |
| Aims of Study | Assess methods to minimise pain experienced by patients after root canal treatment |
| Population | 450 patients receiving endodontic treatment |
| Concept | Assessing post op pain levels of patients who received endodontic treatment |
| Context | Dental pain after endodontic treatment |
| Results of Study | Techniques that reduced patient's anxiety and post op pain were:<br>- Informing patients about what was wrong with their tooth, how treatment would be carried out, what to expect post-op, how potential symptoms would be prevented and controlled<br>- Describing the procedure as it was happening<br>- informing patients that they could expect pain to worsen for the next 24 hours post-op and improve by the end of the 3rd day post op<br>- Phoning patient the next day for follow up<br>- Prescribing antibiotics and analgesics after tx |

| General information |  |
| --- | --- |
| Date of data extraction | 21/04/2023 |
| Person | BT |
| extracting data |  |
| PMID | 8120489 |
| Date of publication | 1993 |
| Author(s) | Selden, H. S. |
| Title | Patient empowerment--a strategy for pain management in endodontics |
| Type of Publication | Journal article |
| Location of Study | American association of endodontists |
| **Study Details** |  |
| Study Design | Retrospective case controlled study |
| Aims of Study | Examine the effects of different variables (pt education and information, distraction, next day phone call, abx and analgesics) on patient empowerment and hence, their coping capacity during and after one visit endodontic therapy was performed. |
| Population | 540 Dental patients who underwent one visit endodontic treatment and had sufficient documentation of their treatment (including tooth number, following day phone call request, summary of phone report in relation to symptoms etc). |
| Concept | A universal one visit endodontic treatment (OVE) program integrated behavioural strategies to help increase pt's coping capacity. Then, the patient's charts were evaluated for information on their experience, and time it took for resolution of symptoms based on a follow up phone call.<br>The strategies incorporated into the OVE program include:<br>- Education and informing patients about what is wrong with their tooth, treatment steps, what to expect post operatively, and how potential symptoms would be prevented and controlled<br>- Distraction: pts were able to see a reflected image of their tooth during the procedure, which is accompanied by commentary.<br>- Notify what to expect in terms of postoperative pain: could worsen right after the procedure, and during the 24 hr before the healing process begins, and that can expect significant reduction in symptoms by the end of the third day..<br>- Next day phone call: patients were encouraged to call the office the next day to report their progress. Pts were even given doctor's home phone numbers for after hours calls.<br>- Antibiotics: abx was given either before or immediately after treatment.<br>- Analgesics: given at the time of treatment. NSAID and opioid of severe pain. |
| Context | OVE is associated with concerns of inducing postoperative pain, so this article looks at behavioural techniques to increase <u>patient empowerment</u> and assess their effects on post op healing and reduction of symptoms. |
|  | It was thought that <u>patient empowerment</u> would increase their coping strategies when recovering after OVE therapy. |
| Results of Study | - Education and information: improved communication and established confidence in the reliability of root canal treatment → reduced anxiety, which is linked to less postop pain,<br>- Distraction: Pts were more relaxed and helped time go by faster.<br>- Post op pain: Pts felt reassured and patient empowerment was increased<br>- Next day phone call: enhanced the pt's perception of the doctors sincerity, because it was perceived as an expression of concern.<br>Of the 540 patient charts included: 420 (78%) called and 120 (22) did not call<br>Of the 420 pts that called: 390 (93%) reported improvement, and 30 (7%) said symptoms remained the same.<br>- The 30 people were asked to continue measures and/or medication as prescribed.<br>- 20 phoned the next day and reported improvement<br>- The remaining 10 who didn't phone again reported (in later appointments) not phoning because they felt better and felt like they didn't need to bother.<br>Concluded that OVE helps resolve symptoms and infections. Seems more rapid compared to the standardised multiple visit technique. However, it is hard to determine if there is a specific variable that affected success more than the others. |

Article 4:
| General information |  |
| --- | --- |
| Date of data extraction | 20/4/23 |
| Person extracting data | JT |
| PMID | 34472104 |
| Date of publication | 2021 |
| Author(s) | Kuscu, O. O.; Ozcelik, S. M.; Kucuktepe, C.; Akyuz, S. |
| Title | Effect of self-efficacy and empathy characteristics of post-graduate residents on local anaesthesia administration performances: Pilot study |
| Type of Publication | Journal article |
| Location of Study | Department of Paediatric Dentistry, Marmara University Turkey |
| Study Details |  |
| Study Design | Prospective cohort study |
| Aims of Study | Investigate the effects of empathy of post-graduate residents on pain and anxiety |
|  | levels of children during local anaesthesia administration |
| Population | 10 paediatric dentistry post-grads and 91 children |
| Concept | All post-grads were assessed using Jefferson Scale of Physician Empathy scores. Post-grads administered 12 injections to their patients. VAS scores of children were used to evaluate pain during treatment, and VPT scores were used to evaluate anxiety. |
| Context | Pain experienced during local anaesthesia administration in paediatric dental clinic |
| Results of Study | Postgrads with higher self-efficacy perception and better empathy skills provided more painless dental injections. No difference in anxiety between genders of children. Higher VAS pain scores in girls compared to boys. |
| Date of data extraction | 22/04/2023 |
| Person extracting data | SN |
| PMID | 34472104 |
| Date of publication | 27/02/2021 |
| Author(s) | Kuscu, O. O.; Ozcelik, S. M.; Kucuktepe, C.; Akyuz, S. |
| Title | Effect of self-efficacy and empathy characteristics of post-graduate residents on local anaesthesia administration performances: Pilot study |
| Type of Publication | Original Article |
| Location of Study | Department of Pediatric Dentistry , Faculty of Dentistry, Marmara University , Istanbul, Turkey |
| **Study Details** |  |
| Study Design | Prospective Cohort Study |
| Aims of Study | Aim of the study was to examine the effects of self-efficacy and empathy traits of post-grads of perceived pain and anxiety experienced by children during the process of administering local anaesthetic. |
| Population | Children between the ages of 5-13 |
| Concept | The concept was to see if there was a correlation between the two traits: self efficiency and empathy , with the pain and anxiety experienced by children. Each of the 10 post grads ( dental ) performed 12 dental injections for a variety of treatments ( restorative, endodontic , extraction), on 91 children total .Empathy and self efficacy's impact on pain experience was examined using the following scales:<br>- Jefferson scale of physician empathy student version - S version<br>- CFSS-DS -this scale measured children's fear -Likert type scale |
|  | - Venhan Picture test scale - measure state anxiety<br>- FIS scale ( facial image scale)<br>- VAS (visual analog scale) |
| Context | This study was conducted by post grad dental students enrolled at Marmara University, Istanbul,Turkey, conducted with children registered in the Pediatric Dentistry Clinica between Nov 2018 and April 2019. |
| Results of Study | The results of this study showed:<br>- Self efficacy and empathy of post grad dental students did impact perceived pain and anxiety scores of children during LA administration.<br>- Therefore the development of empathy and self efficacy in post grads would result in more comfortable dental experiences in paediatric patients |

Article 5:
| General information |  |
| --- | --- |
| Date of data extraction | 20/4/23 |
| Person extracting data | JT |
| PMID | 3422668 |
| Date of publication | 1988 |
| Author(s) | Corah, N. L.; O'Shea, R. M.; Bissell, G. D.; Thines, T. J.; Mendola, P |
| Title | The dentist-patient relationship: perceived dentist behaviours that reduce patient anxiety and increase satisfaction |
| Type of Publication | Journal article |
| Location of Study | Public hospital dental clinic (doesn't specify which one) |
| **Study Details** |  |
| Study Design | Cross sectional study |
| Aims of Study | Assess impact of dentist behaviours on patient anxiety |
| Population | Patients at public hospital dental clinic having invasive procedures |
| Concept | Patients filled out an anxiety score (DAS) questionnaire before and after dental procedure |
| Context | Invasive dental procedures at public hospital dental clinic |
| Results of Study | More anxious patients perceived the dentist as: asking them whether they were anxious, not working quickly, not washing hands before treatment, not paying attention to what they said<br>Dentist behaviours most associated with satisfaction were: dentist explaining procedures, encouraging questions, having a calm manner, taking patient seriously, not criticising the patient's oral condition, being reassuring, talking in |
|  | understandable way |
| Date of data extraction | 22/4/2023 |
| Person extracting data | SN |
| PMID | 3422668 |
| Date of publication | September 1985 |
| Author(s) | Norman L. Corah, Robert M. O'Shea, G.Donals Bissell,DDS |
| Title | The dentist- patient relationship: perceptions by patients of dentist behaviour in relation to satisfaction and anxiety |
| Type of Publication | Article |
| Location of Study | Large public hospital dental clinic |
| Study Details |  |
| Study Design | Cross sectional cohort study |
| Aims of Study | To investigate the impact of perceived dentist behaviours to anxiety in patients during treatment, and overall satisfaction during the dental visit |
| Population | Outpatients in the waiting room of the large public hospital dental clinic, who were present for an invasive procedure above the age of 18 and were not in severe pain |
| Concept | Before the dental procedure, patients filled out a dental anxiety scale. After the procedure they rated their anxiety during the procedure, answered a questionnaire concerning 21 possible dentist behaviours and completed the Dental Visit Satisfaction Scale. A correlation was developed between anxiety during the treatment and the DVSS with the perceived dentist behaviour items. |
| Context | Conducted in a large public hospital, this study was conducted to see if there was an association between anxiety and patient satisfaction with dental behaviours.<br>Sample: 231 adult patients, 107 men, 124 women at a public hospital dental clinic. |
| Results of Study | Majority of the dental behaviours on the list (including aspects of communication, acceptance, competence, and total satisfaction) were related to the satisfaction of patients. Accepting and caring were two traits contributing most to patient satisfaction. Anxiety was identified as a negative predictor. |

Article 6:
| General information |  |
| --- | --- |
| Date of data | 21/4/23 |
| extraction |  |
| Person<br>extracting data | JT |
| PMID | 298463 |
| Date of<br>publication | 1979 |
| Author(s) | Foreman, P. A |
| Title | Behavioural considerations in patient management |
| Type of<br>Publication | Journal article |
| Location of<br>Study | N/A (not a study) |
| **Study Details** |  |
| Study Design | N/A |
| Aims of Study | N/A |
| Population | Dental patients |
| Concept | Techniques dentists can use to manage anxious patients |
| Context | Dental clinic |
| Results of | Anxious patients have lower pain tolerance |
| Study | Article discusses several techniques dentists can use to manage anxious patients:<br>- Anticipation reward (verbal encouragement from dentist)<br>- Modelling (patient observes the treatment being done on someone else)<br>- Friendly and empathetic environment, including dental staff<br>- Patient education (explaining treatment to patient before doing it)<br>- Hypnosis<br>- Establishing a good dentist-patient relationship |
| Date of data extraction | 23/4/2023 |
| Person extracting data | RB |
| PMID | 298463 |
| Date of publication | 1979 |
| Author(s) | Foreman, P. A |
| Title | Behavioural considerations in patient management |
| Type of Publication | Journal piece |
| Location of Study | Auckland, New Zealand |
| **Study Details** |  |
| Study Design | NA |
| Aims of Study | To explore the behavioural approach of managing dental patients presenting anxiety and pain. |
| Population | Dental patients with pain and anxiety |
| Concept | Behavioural techniques used to manage anxious patients presenting with pain including anticipation reward, modelling, environmental set up, patient education, hypnosis and suggestion, dentist/patient relationship |
| Context | Dental setting |
| Results of Study | Managing the dentist/patient relationship is key for having a good therapeutic relationship with patients and managing patients with dental anxiety (termed iatrosedation). The approach to managing an anxious patient should be multidisciplinary and include iatrosedation and pharmacosedation. Anxious patients are more prone to pain, and so by managing a patient and reducing their anxiety with behavioural techniques (such as anticipation reward, modelling, environmental set up, patient education, hypnosis, suggestion and the dentist/patient relationship), the dentist will be able to better manage their pain. |

Article 7:
| General information |  |
| --- | --- |
| Date of data extraction | 21/4/23 |
| Person extracting data | JT |
| PMID | 14715397 |
| Date of publication | 2004 |
| Author(s) | Riley, Iii J. L.; Gilbert, G. H.; Heft, M. W. |
| Title | Oral health attitudes and communication with laypersons about orofacial pain among middle-aged and older adults |
| Type of Publication (e.g. journal article, dissertation) | Journal article |
| Location of Study | Florida |

| Study Details |  |
| --- | --- |
| Study Design | Prospective longitudinal study |
| Aims of Study | To examine associations between attitudes and beliefs about oral disease and dental care and talking about orofacial pain with laypersons and health care professionals |
| Population | Dental patients in Florida |
| Concept | Interviews with patients about their attitudes and expectations towards dental care, and recent pain history. |
| Context | Dental setting |
| Results of Study | - People who did not talk to a healthcare provider about their pain had more negative attitudes to dental care<br>- Factors associated with satisfaction with dental care: relationship with dentist, perceived technical quality of the care, physical environment |

| General information |  |
| --- | --- |
| Date of data extraction | 22/4/2023 |
| Person extracting data | SN |
| PMID | 14715397 |
| Date of | 6/10/2003 |
| publication |  |
| Author(s) | Riley, Iii J. L.; Gilbert, G. H.; Heft, M. W. |
| Title | Oral health attitudes and communication with laypersons about orofacial pain among middle-aged and older adults |
| Type of Publication | Article |
| Location of Study | Florida Dental Care |
| **Study Details** |  |
| Study Design | Prospective longitudinal |
| Aims of Study | Test if attitudes about oral disease and dental care are predictive of pain related discussions with the average individual and a healthcare professional |
| Population | 742 community dwelling adults, aged 45 and above , dentate |
| Concept | 724 participants who were a part of a study examining oral health in community dwelling adults, were interviewed at baseline, and 42 months later. The interviews lasted 30 minutes. The first one was in person , the next one ( 42 months later) , was a telephone interview. The first interview collected information including demographic, attitude towards dental care, the second interview included a discussion about orofacial pain symptoms |
| Context | Seeing if beliefs about oral disease impacts whether or not patients(who are under |
|  | Florida Dental Care Study) communicate about oro-facial pain with laypersons or health professionals. |
| Results of Study | Results include:<br>- If a person doesn't have overly positive views about health system , they are more likely to consult with a lay person as opposed to a healthcare professional when experiencing an oro-facial pain episode<br>- Therefore , by understanding what impacts patient communication, we can get a better understanding of how pain should be managed |

Article 8:
| General information |  |
| --- | --- |
| Date of data extraction | 21/4/23 |
| Person extracting data | JT |
| PMID | 19840152 |
| Date of publication | 2010 |
| Author(s) | Smith, P. A.; Freeman, R. |
| Title | Remembering and repeating childhood dental treatment experiences: parents, their |
|  | children, and barriers to dental care |
| Type of Publication | Journal article |
| Location of Study | Dundee Dental Hospital, Scotland |
| **Study Details** |  |
| Study Design | Case control study |
| Aims of Study | To investigate how the reported childhood dental care experiences of a group of Scottish parents impacted upon the dental treatment they accessed for their children. |
| Population | Parents of children referred to Dundee Dental Hospital for assessment of dental extractions |
| Concept | Interviewing parents of children having dental treatment, about their own past experiences and their child's experiences at the dentist |
| Context | Dental hospital |
| Results of Study | - Parents who had negative past experiences at the dentist were uneasy/cautious about dental treatment for their children<br>- Parents who had negative experiences were more likely to postpone their own child's dental treatment until the child was in pain<br>- Dental anxiety in parents was reinforced by the feeling that dentists were not sufficiently attentive to pain relief and did not take their dental fears |
|  | about pain and discomfort seriously |
| Date of data extraction | 23/4/2023 |
| Person extracting data | SN |
| PMID | 19840152 |
| Date of publication | 2009 |
| Author(s) | Smith, P. A.; Freeman, R. |
| Title | Remembering and repeating childhood dental treatment experiences: parents, their children, and barriers to dental care |
| Type of Publication | Journal Article |
| Location of Study | University of Dundee, Dundee,UK |
| Study Details |  |
| Study Design | Case Control Study |
| Aims of Study | Impact of childhood dental care experiences of parents impacted dental treatment accessed for their children |
| Population | Scottish Parents, with children who were referred as patients to Dundee Dental Hospital |
| Concept | 19 parents of varying ages were interviewed about their own and their children's dental care experiences. The results were then coded with theme headings and summary charts were constructed. |
| Context | The context of this study was to assess how this population of parents ( Scottish parents, with children as patients at Dundee Dental Hospital) described their own experiences , and how these experiences shaped the way they sought dental treatment for their children. |
| Results of Study | It was found that childhood experiences of the parents were vivid enough to recount a level of discomfort which in turn delayed them seeking dental treatment for themselves and their children. |

Article 9:
| General information |  |
| --- | --- |
| Date of data extraction | 21/4/23 |
| Person extracting data | JT |
| PMID | 2086605 |
| Date of publication | 1990 |
| Author(s) | Johnson, Brigitte; Mayberry, William E.; McGlynn, F. |
| Title | Exploratory factor analysis of a sixty-item questionnaire concerned with fear of dental treatment |
| Type of Publication | Journal article |
| Location of Study | University of Missouri-Kansas City School of Dentistry |
| **Study Details** |  |
| Study Design | Cross sectional study |
| Aims of Study | Compare the effect of different dental factors on fear in patients |
| Population | Patients attending University of Missouri-Kansas City School of Dentistry |
| Concept | Questionnaire given to patients, patients rated how much fear certain items in dental setting elicited |
| Context | Dental clinic |
| Results of Study | Found that various events in dental clinic elicit fear including clinician behavioural factors such as:<br>- Dentist asking questions while patient's mouth is full<br>- Dentist saying patient has not been taking good care of teeth<br>- Dentist saying there are several cavities |
| Date of data extraction | 22/04/23 |
| Person extracting data | BT |
| PMID | 2086605 |
| Date of publication | 1990 |
| Author(s) | Johnson, Brigitte; Mayberry, William E.; McGlynn, F. |
| Title | Exploratory factor analysis of a sixty-item questionnaire concerned with fear of dental treatment |
| Type of Publication | Journal article |
| Location of Study | University of Missouri-Kansas School of dentistry |
| **Study Details** |  |
| Study Design | Cross Sectional Study |
| Aims of Study | This study attempts to delineate the components of dental fear (using exploratory factor analysis, which was missing from previous studies) |
| Population | Dental patients presenting to the university of missouri-kansas school of dentistry<br><u>Exclusion criteria:</u> patients with less than 7 grades of education<br>745 people agreed to participate of which, 701 gave usable data.<br><u>Patient characteristics:</u><br>- Mean age: 49<br>- Modal family income: \$20k-30k<br>- Most common reason for visit: keeping regular dental appointment |
| Concept | Previous studies have summarised the components of dental fear to be:<br>- Pain or anticipated pain, or Negative evaluation<br>- Dental fear contains a component of physical danger as well as a component of threat to self esteem.<br>- Anticipatory fear and avoidance, fear of pain related events, and physiological arousal.<br>But these did not take into account exploratory factor analysis. |
| Context | These dental patients were asked to fill out a questionnaire, which was devised so as to sample as comprehensively the events involved routinely in seeking and receiving dental care, and to avoid a priori theoretical bias. I.e. It was designed to incorporate the usual events that are involved in seeking and receiving dental care. |
| Results of Study | 4 categories were found to be:<br>- Painful events and precursors to painful events<br>- Activities leading to dental visit<br>- Negative evaluation by the dentist (dentist say you have cavities, dentist say you have bad teeth, dentist lecture you on your poor dental care, dentist say you have not been taking care of your teeth)<br>- Losing control to the dentist (dentist asks a personal question while you have a mouth full of cotton) |

Article 10:
| General information |  |
| --- | --- |
| Date of data extraction | 23/4/23 |
| Person extracting data | JT |
| PMID | 3290300 |
| Date of | 1988 |
| publication |  |
| Author(s) | Curry, S. L.; Russ, S. W.; Johnsen, D. C.; DiSantis, T. A. |
| Title | The role of coping in children's adjustment to the dental visit |
| Type of Publication | Journal article |
| Location of Study | USA |
| **Study Details** |  |
| Study Design | Cross-sectional study |
| Aims of Study | Assess childrens' self-initiated coping strategies in dental situation |
| Population | 8-10 year old children receiving dental care |
| Concept | Interviews and observations of children in dental settings conducted to explore coping strategies children use in dental clinics. E.g. information seeking (asking questions, being watchful, inspecting instruments) |
| Context | Paediatric clinic of Case Western Reserve School of Dentistry |
| Results of Study | - Most children use a variety of cognitive and behavioural coping strategies in dental setting<br>- Cognitive coping strategies appear to be more effective than behavioural strategies. E.g. thinking realistic thoughts about dental procedure, cognitive reappraisal (focusing on positives), emotional regulating |
|  | (comforting self), diversionary thinking (distracting themselves),<br>behaviour regulating (controlling own behaviour during treatment like<br>telling themselves to sit still)<br>- Dentist can play a role in encouraging effective coping strategies -<br>systematically preparing child for treatment, using tell show do |
| Date of data<br>extraction | 23/4/2023 |
| Person<br>extracting data | RB |
| PMID | 3290300 |
| Date of<br>publication | 1998 |
| Author(s) | Curry, S. L.; Russ, S. W.; Johnsen, D. C.; DiSantis, T. A. |
| Title | The role of coping in children's adjustment to the dental visit |
| Type of<br>Publication | Journal article |
| Location of<br>Study | Department of Pediatric Dentistry, Case Western Reserve University, Cleveland<br>Ohio |

| Study Details |  |
| --- | --- |
| Study Design | Cross-sectional study |
| Aims of Study | Aims:<br>- To make the dentist aware of the structure, complexity and significance of coping strategies by introducing some of the research literature on coping<br>- To summarise the results of two recent studies by the authors on children's self initiated coping efforts in dental situations<br>- To explore the implications for dentists treating children |
| Population | 8-10 year old children receiving restorative dental work at the paediatric dental clinic of Case Western Reserve School of Dentistry<br>7-10 year old children receiving dental work at a private practice |
| Concept | Identifying and evaluating children's spontaneous coping behaviours in the dental setting.<br>Behaviour coping:<br>- Information seeking<br>- Support seeking<br>- Direct efforts to maintain control<br>Cognitive coping:<br>- Reality orientated working through<br>- Cognitive reappraisal<br>- Emotion-regulating cognitions<br>- Behaviour regulating cognitions |
|  | - Diversionary thinking |
| Context | Dentists treating children |
| Results of Study | - Most children used a variety of cognitive and behavioural strategies.<br>- The greater the number and the variety of cognitive responses the better the child's adjustment, overall cognitive coping strategies were better<br>- Behaviour strategies sometimes lead to poorer outcomes<br>Children are very active in their use of silent coping in the dental setting, and the use of coping relates to their behaviour in the dental setting.<br>Reality-orientated working through was the most highly correlated with adjustment (dentist systematically prepare children for treatment by describing and demonstrating the upcoming procedure) = the dentist's recognition of specific coping strategies may be followed by appropriate techniques to enhance the child's use of natural coping abilities |

Article 11:
| General information |  |
| --- | --- |
| Date of data extraction | 23/4/23 |
| Person extracting data | JT |
| PMID | 7937263 |
| Date of publication | 1994 |
| Author(s) | Milgrom, P.; Weinstein, P.; Golletz, D.; Leroux, B.; Domoto, P |
| Title | Pain management in school-aged children by private and public clinic practice dentists |
| Type of Publication | Journal article |
| Location of Study | Seattle, Washington |
| **Study Details** |  |
| Study Design | Cross-sectional |
| Aims of Study | Assess beliefs of dentists about pain control in school aged children and the relationship of those beliefs to pain management behaviours |
| Population | Dentists from Seattle Washington who were likely to treat a significant number of school aged children |
| Concept | Surveys were mailed to dentists asking them to rate:<br>- pain level of procedures<br>- child pain management (use of LA, giving additional LA at child's |
|  | request, provision of post-op pain medication)<br>- beliefs about children thought to influence child pain management (level of comfort working with children, frustrating encounters with children, dentist need for control, communicating with child) |
| Context | Dentists treating children |
| Results of Study | - 2/3 of dentists surveyed always use LA and additional anaesthetic upon child request<br>- 2/3 of dentists provide post-op pain medication<br>- Most dentists believe dental care for children is not painful, just unpleasant<br>- Dentists believe children confuse pressure with pain, or present false/exaggerated responses<br>- Dentist beliefs about child pain influence pain management practices - e.g. dentists who rate procedures are more painful are more likely to provide post-op pain medications |
| Date of data extraction | 23/4/2023 |
| Person extracting data | RB |
| PMID | 7937263 |
| Date of publication | 1994 |
| Author(s) | Milgrom, P.; Weinstein, P.; Golletz, D.; Leroux, B.; Domoto, P |
| Title | Pain management in school-aged children by private and public clinic practice dentists |
| Type of Publication | Journal article |
| Location of Study | Seattle, Washington |
| **Study Details** |  |
| Study Design | Longitudinal study |
| Aims of Study | Explore dentists' perceptions of child pain and beliefs regarding pain management in children. |
| Population | Dentists from Seattle who were likely to be treating a significant number of school-aged children (at least 5%) |
| Concept | Interviews were posted to dentists treating a significant number of children to determine their perceptions of children's pain at the dentist and pain management. These surveys asked about: |
|  | - Beliefs about children thought to influence pain management<br>- Dentist comfort with children<br>- % frustrating encounters with children<br>- Dentist need for control over the child<br>- Communicating with child asking about their distress<br>- Painfulness of procedure for child<br>- Unpleasantness of procedure for child<br>- Denial of child pain report<br>- Child pain management<br>- Use of LA<br>- Giving additional LA<br>- Providing post pain medication |
| Context | Dentists treating children |
| Results of Study | - Dentists who expressed a greater need for control were less likely to give LA<br>- Dentists in poorer socioeconomic areas gave less LA despite the likelihood that the children may experience more severe dental problems<br>- Dentists rating dental procedures are less painful are more likely to provide LA if the child is in distress<br>- Dentist behaviour that serves to lower anxiety and enhance at least perceived control will reduce child pain (eg being able to communicate with the child about pain or comfort or being frightened may reduce child's fear) = gives child some control (studies show control reduces pain) |
|  | Conclusions:<br>- 1/3 dentists don't always use LA for restorations or extractions<br>- 10% of dentists regularly deny child pain and don't believe child pain reports are valid<br>- Associations exist between the use of pain control procedures and child management beliefs |

Article 12:
| General information |  |
| --- | --- |
| Date of data extraction | 23/4/23 |
| Person extracting data | JT |
| PMID | 624781 |
| Date of publication | 1978 |
| Author(s) | Dangott, Lillian; Thornton, B.; Page, Paul |
| Title | Communication and pain |
| Type of | Journal article |
| Publication |  |
| Location of Study | N/A |
| **Study Details** |  |
| Study Design | N/A |
| Aims of Study | N/A |
| Population | N/A |
| Concept | Summary of different research examining dentist behaviours that can impact on pain in the dental setting. |
| Context | Dental clinic |
| Results of Study | - For interactions to be positive, a feeling of recognition, acknowledgement, and endorsement must be conveyed<br>- Many clinicians do not use nonverbal behaviours in communicating about pain<br>- Discourse is often tightly controlled by the medical professional - little input from patient |

|  |
| --- |
| **General information** |
| Date of data extraction | 23/4/2023 |
| Person extracting data | RB |
| PMID | 624781 |
| Date of publication | 1978 |
| Author(s) | Dangott, Lillian; Thornton, B.; Page, Paul |
| Title | Communication and pain |
| Type of Publication | Journal piece |
| Location of Study | NA |
| **Study Details** |  |
| Study Design | NA |
| Aims of Study | Discuss the ways in which health professionals communicate about pain in a way that builds rapport, supportive interaction and commonality |
| Population | Dentists |
| Concept | Article discusses research conducted which examines how dentists and health |
|  | professionals communicate with their patients about pain. The article also discusses medical jargon and the significance of noting the differences in the health communication and emotional communication |
| Context | Dentists treating patients |
| Results of Study | - Communication itself can have healing effects, and there seems to be serious communication problems between health providers and patients = need for development of a more effective paradigm of interaction<br>- Presence of pain hinders communication<br>- Use of non verbal communication to allow pts to express themselves (especially when they can't talk)<br>- Health professionals sometimes are disconfirming (eg avoid pain and say 'pinch'), also some are just very factual about saying a patient is in pain. This results in the pt being told how they should feel and doesn't allow opportunity for response.<br>- Environmental distractions, security and trust play a major part in the degree of pain experience by the patient<br>- Social interactions (verbal + non verbal can subdue or exaggerate pain sensation) |

Article 13:
| General information |  |
| --- | --- |
| Date of data extraction | 23/04/2023 |
| Person extracting data | SN |
| PMID | 2383-9309 |
| Date of publication | 29/1/2019 |
| Author(s) | Sreeraksha Radhakrishna, Ila Srinivasan, Jyothsna V Setty, Murali Krishna DR, Anjana Melwani, Kuthpady Manasa Hegde |
| Title | Comparison of three behavior modification techniques for management of anxious children aged 4-8 years |
| Type of Publication | Original Article |
| Location of Study | Department of Pediatric and Preventive Dentistry, M.R Ambedkar Dental College and Hospital, Bengaluru, Karnataka, India |
| **Study Details** |  |
| Study Design | RCT |
| Aims of Study | Comparing the techniques of Tell-Show-Play-Doh, a smartphone dentist game and the conventional tell-show-do method in modifying the behaviour of anxious children at the dentist |
| Population | Children in the age group of 4-8 years |
| Concept | 60 children within the 4-8 years age range who required class 2 and class 1 cavity |
|  | restorations, who fit in the Frankl's behaviour rating of 2 or 3 were split into 3 groups of 20. One group was the Tell-show-Play-doh groups ( play-doh, plastic human head, with play doh teeth - to simulate the procedure in an interactive way) , one group was the conventional Tell-Show-Do group, the other was a smartphone dentist game (animated pictures-visual and sound effects). Pulse rate, Facial Image Scale and Frankl's behaviour rating scale and the Face, Leg, Activity, Cry, Consolability behaviour scale was used to quantitatively measure anxious behaviour. |
| Context | Department of Pediatric and Preventive Dentistry, M.R Ambedkar Dental College and Hospital |
| Results of Study | Tell-show-Play-doh group and the smartphone group were more effective at reducing anxiousness related behaviour than the conventional Tell-show-do group. |
| Date of data extraction | 23/4/2023 |
| Person extracting data | RB |
| PMID | ISSN: 2383-9309 |
| Date of publication | 2019 |
| Author(s) | Sreeraksha Radhakrishna, Ila Srinivasan, Jyothsna V Setty, Murali Krishna DR, Anjana Melwani, Kuthpady Manasa Hegde |
| Title | Comparison of three behaviour modification techniques for management of anxious children aged 4-8 years |
| Type of Publication<br>(e.g. journal article, dissertation) | Journal article |
| Location of Study | India |
| **Study Details** |  |
| Study Design | RCT |
| Aims of Study | The aim of this study was to compare the techniques of Tell-Show-Play-doh, a smartphone dentist game, and a conventional Tell-Show-Do method in the behaviour modification of anxious children in the dental operatory. |
| Population | 4-8 year old anxious children |
| Concept | Exploring behaviour modification techniques to see which reduces anxiety in children at the dentist.<br>- Group 1 = Tell-Show-Play-doh. Uses learning to reduce anxiety, play-doh drill and fill sets to show how a playdoh tooth is cleaned and filled |
|  | - Group 2 = smartphone game. Smartphone game to demonstrate dental equipment, dental sounds and let the kids see how the equipment is used<br>- Group 3 = Tell-show-do. Kids are able to play with dental imitation toys and see demonstration of dentistry on a model |
| Context | Paediatric dentists |
| Results of Study | Play-Doh Doctor Drill n' Fill toy set and Dentist games for kids are far superior than the Tell-Show-Do technique in reducing anxiety and managing the child effectively during dental treatment |

Article 14:
| General information |  |
| --- | --- |
| Date of data extraction | 22/04/23 |
| Person extracting data | BT |
| PMID | PMID: 296701 |
| Date of publication | 1979 |
| Author(s) | John P Brown, Ira T Smith |
| Title | Childhood fear and anxiety states in relation to dental treatment |
| Type of Publication | Journal article |
| Location of Study | Queensland |
| **Study Details** |  |
| Study Design | Caes Reports |
| Aims of Study | “Describe straight forward behaviour modification techniques which can be used to help the fearful or excessively anxious child cope with dental treatment” |
| Population | Young children. The case reports involved a 7F, 5M and 11M |
| Concept | Understanding how a conditioned fear develops in children, in the context of a dental situation, can be beneficial in developing techniques to help uncooperative children cope with dental treatment. |
| Context | Conditioned fear can manifest when:<br>- Dentist breaks the child’s trust by going beyond the child’s limit or their expectations → child will develop a fear of similar situations<br>- Level of warning regarding expected pain is: not enough, and the child will lose faith in the adult; or too little, and the pain tolerance may be lowered. Need to be enough to allow the child to have a realistic expectation of pain<br>- Fear of events are a result of experience AND from peer/family attitudes/experiences |
|  | Best fear modifier is the development of a sense of security - having a parent present at a dental visit will establish familiarity with the event. Make sure it is a small, active presence (e.g. hand holding)<br>- Stop interruption with explanation<br>- Passive presence can cause a child to feel deserted.<br>Children need to approach potentially frightening situations in their own way and at their own pace with appropriate supporting encouragement. IF a child feels trapped in a threatening situation (regardless of how innocuous they may seem, panic will ensue. |
| Results of Study | Case 1: 7F and 5M presented with unmanageable behaviour in dental treatment situations due to experience at first dental appointment where teeth were extracted unexpectedly (adverse conditioning).<br>- 7F got better with a sympathetic environment, adequate explanation, forewarning, relaxation and reassurance, which helped overcome her fearful conditioned response (systematic desensitisation). She had a strong need for approval, which highlighted the importance of the motivation to comply for successful behaviour management<br>- 5M harder to tx. Was rewarded after uncooperative visits - reinforced bad behaviour. Turned from anxiety behaviour to tantrum behaviour, which required a more disciplinary approach.<br>Case 2: immersion in the fearful circumstance helped the 11M to rationalise |
|  | fear – had the filling done after unsuccessful attempts to give LA. Was told that the pain could have been prevented by LA and high cooperation → pt asked for and accepted LA on the next visit. Then, the reward of approval from the dentist became more valuable to him than the reward of avoiding the threatening situation. |
| Date of data extraction | 23/4/2023 |
| Person extracting data | RB |
| PMID | 296701 |
| Date of publication | 1979 |
| Author(s) | Brown, J. P.; Smith, I. T. |
| Title | Childhood fear and anxiety states in relation to dental treatment |
| Type of Publication | Journal article |
| Location of | University of Queensland |
| Study |  |
| **Study Details** |  |
| Study Design | Case reports |
| Aims of Study | To illustrate how behaviour modification techniques can be used to help the fearful or excessively anxious child cope with dental treatment |
| Population | Young children presenting with fear/anxiety in dental treatment |
| Concept | - Production of conditioned fear: broken trust/bond between child and dentist = child has a generalisation of fear about similar circumstances<br>- Eg, experience of pain when the child was told there would be no pain<br>- Anticipation of pain tends to be more anxiety producing than the actual experience<br>- Best fear modifier is the development of a sense of security = parent should be present at appointments to establish familiarity with the event |
| Context | Dental setting - treating fearful/anxious children |
| Results of Study | - Pay close attention to the history of behaviour disturbance, and particularly previous untoward dental and medical experience<br>- Give realistic explanations so that the limits of the procedure are known to the child<br>- Allow the child some initiative so that the limits of the procedure are known to the child |
|  | - Be patient in establishing rapport<br>- Endeavour to get the child to actively participate in the procedure<br>- Show the child that you look on him or her as a considered person and not merely someone accompanying the parent |

Article 15:
| General information |  |
| --- | --- |
| Date of data extraction | 23/04/2023 |
| Person extracting data | SN |
| PMID | 605-610 |
| Date of publication | July 2013 |
| Author(s) | Keith, D. J.; Rinchuse, D. J.; Kennedy, M.; Zullo, T. |
| Title | Effect of text message follow-up on patient's self-reported level of pain and anxiety |
| Type of Publication | Original Article |
| Location of | Seton Hill University Institutional Review Board ,Greensburg,Pa. |
| Study |  |
| **Study Details** |  |
| Study Design | RCT |
| Aims of Study | After initial placement of orthodontic appliance, does a text message reduce self reported levels of pain and anxiety |
| Population | 39 orthodontic patients- consecutively treated , between the ages of 10 and 18, orthodontic treatment with fixed maxillary edgewise appliances , no previous orthodontic treatment |
| Concept | The 39 patients were split into two groups. Both groups initially filled out a baseline questionnaire about levels of pain and anxiety before the implementation of the orthodontic appliance. Following one week, all subjects filled out the pain and anxiety questionnaire daily. One group got a text message that displayed concern and reassurance whilst the other did not receive any communication. |
| Context | A research protocol that was approved by Seton Hill University Institutional Review Board was employed in this study. This study looked at 39 patients between the ages of 10-18 years of age. |
| Results of Study | Text message sent following initial appliance placement results in lower pain levels (self reported) .Anxiety levels did not alter in response to text messaged. Anxiety did show a decrease from a maximum (at the initial appointment) overall. |

|  |
| --- |
| **General information** |
| Date of data extraction | 23/4/2023 |
| Person extracting data | RB |
| PMID | 605-610 |
| Date of publication | July 2013 |
| Author(s) | Keith, D. J.; Rinchuse, D. J.; Kennedy, M.; Zullo, T. |
| Title | Effect of text message follow-up on patient's self-reported level of pain and anxiety |
| Type of Publication | Journal article |
| Location of Study | Seaton Hill University Center for Orthodontics, Greensburg |
| **Study Details** |  |
| Study Design | RCT |
| Aims of Study | The purpose of this study was to determine if there is a difference in self-reported pain and anxiety after initial placement of orthodontic appliances between patients who receive a postprocedural text message and those who receive no correspondence post appointment. |
|  | - Does written work have the same impact as spoken word (telephone call) |
| Population | Patients between 10 and 18 years of age undergoing orthodontic treatment with fixed maxillary edgewise appliances (and no previous ortho treatment) |
| Concept | - “Bibliotherapy” as an adjunct to psychological treatment may help shape behavioural attitudes (written word as an adjunct to be viewed outside of active treatment).<br>- Phone calls tend to reduce pain intensity and anxiety in initial phases of ortho treatment<br>- Given the increasing number of text messages sent, this study investigates if a post op text helps reduce self-report pain and anxiety post op |
| Context | Orthodontic clinic/dental setting |
| Results of Study | - This study established that a text message sent from the orthodontic office following the initial appliance placement resulted in a lower level of self-reported pain.<br>- It was also noted that patient anxiety is at its peak the day following the initial appointment and decreases from that point forward. Overall anxiety, however, did not change as a result of postprocedural communication |

Article 16:
| General information |  |
| --- | --- |
| Date of data | 23/04/23 |
| extraction |  |
| Person extracting data | BT |
| PMID | No. 810<br>18789200 |
| Date of<br>publication | Sep 2008 |
| Author(s) | Marco L Loggia<br>Petra Schweinhardt<br>Chantel Villemure<br>M Catherine Buishnell |
| Title | Effects of psychological state on pain perception in the dental environment |
| Type of Publication | Journal Article |
| Location of<br>Study |  |
| **Study Details** |  |
| Study Design | Narrative (literature) Review |
| Aims of Study | Provide an understanding of the influence of the psychological state on pain transmission. |
| Population | Dental patients |
| Concept | There are many pain modulatory systems that can be accessed pharmacologically, or through contextual or psychological manipulation. While most studies focus on pharmacological methods, this paper focuses more on the psychological<br>Variables altering pain include<br>- Attention state - studies found that pain is less intense when a person is distracted. Proved by fMRI studies<br>- Emotional state - thought that more neg emotions (depression, anxiety, disgust - like when smelling eugenol) lead to more pain. And, dentistry is highly associated with pain and anxiety.<br>- Social influence can affect how we perceive and communicate pain - through imitative learning when seeing another person go through a procedure and how they react, or if a parent was in the same room<br>Then they discuss possible (biological) mechanisms behind how attentional and emotional states affect pain<br>Even when strategies are implemented to target these variables, not everyone responds in the same way and this is largely due to personality type. E.g. relaxation method was particularly effective in highly anxious patients, whereas the music distraction did not have a clinically relevant effect on these patients |
| Context | By knowing and understanding how these psychological/contextual variables affect pain, dental practitioners can create a more fostering environment for pts to |
|  | help deal with their pain. |
| Results of Study | Ways to reduce anxiety and improve mood, in addition to distracting the patient could include: playing music, showing humorous films on a monitor, or filling the room with pleasant odours. Such methods have been shown to be effective in reducing dental pain in some patients.<br>Can also improve anxiety by providing accurate preparatory information before dental procedures. E.g. description of both the sensations that the patient will likely experience and the sequence of medical procedures has been found to yield the strongest and most consistent benefits in terms of reducing negative affect, pain reports and distress<br>Socially:<br>- where parents are present during tx, solicitous attention and/or the amount of attention can affect pain and healing.<br>- When seeing another person go through the same procedure, the patientt will match their pain behaviour with the other person, and more empathetic people tend to be more sensitised to pain when witnessing another in pain ∴ should keep waiting patients away from the sight and sounds of other dental patients.<br>Placebos reducing pain has been shown to be a real effect (rather than due to a change in pain reporting) |
| Date of data extraction | 26/04/2023 |
| Person extracting data | SN |
| PMID | 18789200 |
| Date of publication | September 2008 |
| Author(s) | Marco L. Loggia , Petra Schweinhardt, Chantal Villemure,M.Catherine Bushnell |
| Title | Effects of psychological state on pain perception in the dental environment |
| Type of Publication | Journal Article |
| Location of<br>Study |  |
| Study Details |  |
| Study Design | Review/Narrative Article |
| Aims of Study | Examining the psychological factors that influence pain perception and how a deeper understanding of these factors help improve clinical practice |
| Population |  |
| Concept | The article discusses the different psychological ways in which a patient's experience is shaped. |
| Context |  |
| Results of Study | - Distraction is a useful technique that can result in lower pain , but is however influenced by personality type<br>- Negative emotions tend to be more related to pain than positive emotions, which in some circumstances can be seen to reduce pain.<br>- Playing music, nice scents- can help reduce pain in the dental setting<br>- These techniques are subjective to the patient's personality<br>- Distracting post-op can also be seen a beneficial<br>- Seeing someone else in pain could increase pain so keeping patients separated could be beneficial |

Article 17:
| General information |  |
| --- | --- |
| Date of data extraction | 23/4/2023 |
| Person extracting data | RB |
| PMID | 15132272 |
| Date of publication | 2004 |
| Author(s) | Dennis C. Harper, PhD Donna M. D'Alessandro, MD |
| Title | The Child's Voice: Understanding the Contexts of Children and Families Today |
| Type of Publication | Conference paper |
| Location of Study | Children's Hospital of Iowa, Iowa City |
| **Study Details** |  |
| Study Design | NA |
| Aims of Study | The purpose of this report was to explore contemporary themes for improved contextual understanding of children and families today and their relationship to providing optimal paediatric dental care. |
| Population | Children receiving dental care + paediatric dentists |
| Concept | Twelve themes discussed in this paper:<br>- Parenting practices<br>- More permissive in today's society with discipline less rigorously enforced |
|  | - Changing family structure<br>- Work commitments of parents<br>- Multicultural issues<br>- It is impossible for a health care provider to be familiar with all relevant cultural practices<br>- Each family is a unique cultural unit<br>- Communication skills and rapport is key<br>- Challenging children<br>- Paediatric pain management<br>- Acknowledge the child's perception of the experience<br>- Pain medications, noninvasive techniques, family centred therapy<br>- Communication suggestions<br>- Relaxed, acknowledge child's interests, talking at an appropriate development level<br>- Behavioural management<br>- Dynamic process of dialogue, facial expression and voice tone |
| Context | Dental setting (paediatrics) |
| Results of Study | - Child's voice needs to be heard and addressed<br>- Child and parent are in an evolving context with many demands that may impact dental health + care instructions<br>- Cooperation and communication are key |

| General information |  |
| --- | --- |
| Date of data extraction | 24/04/23 |
| Person extracting data | BT |
| PMID | 15132272 |
| Date of publication | 2004 |
| Author(s) | Dennis C Harper<br>Donna M D'Allessandro |
| Title | The Child's Voice: Understanding the Contexts of Children and Families Today |
| Type of Publication | Conference paper |
| Location of Study | Children's Hospital of Iowa, Iowa City, Iowa |
| Study Details |  |
| Study Design | Summary of research ? Narrative review ? |
| Aims of Study | To review contemporary changes in parenting and their potential impact on child behaviour, and offer some brief, family focused suggestions on behavioural management |
| Population | Paediatric dental patients + paediatric dentists |
| Concept | The different changes and effect on behavioural management include:<br>- Parenting practices: Accountability seems to be directed toward the medical or psychological condition rather than the personal control of the child or the families' skills in disciplinary management. Currently, there are less assertive behavioural management techniques being employed, leading to children who are harder to manage<br>- Childhood psychosocial problems: shown to have increased from 1979 to 1996. The use of psychotropic medications, counselling, and referral also increased substantially<br>- Contexts for children's dental interactions: a child's immediate context is a function of age, prior health care experiences, and often families' existing attitudes toward dental health. Should guide initial interaction to be positive, so it sets the tone for future visits.<br>- Parental and family contexts: some expectations include: expectations of the dentist; health care values; ability to pay for the needed services; and lack of knowledge in preparing a young child for the dental interaction.<br>- Context for the paediatric dentist:<br>- Childhood fears and the dentist: hard to assess whether the child is "resisting" because of anxiety or fears vs more general noncompliance behaviours present in their daily activities<br>- Developmental reactions of children: parents need anticipatory guidance on how to respond to their child's questions about discomfort or pain during the dental interaction. A joint parent/child/dentist plan of treatment |
|  | and dialogue is often the best anticipatory strategy for allaying fears<br>- Changing families:<br>- Multicultural issues: need to keep in mind - 1. What is typical of a particular group does not necessarily predict the beliefs of an individual within that group. 2. It is impossible for any health care provider to be familiar with all relevant cultural practices. 3. Each family is a unique cultural unit. 4. Communication skills and partnership building are essential to improving the relationship between the health care provider and family.<br>- Challenging children: need to be managed or assisted according to their developmental age<br>- Paediatric pain management: pain medications; noninvasive techniques; and family-centred therapy<br>- Communication suggestions: always prep children, using a relaxed style in the tone of voice, acknowledging the child's interests, talk to them at an appropriate developmental level.<br>- Behavioural management in paediatric dentistry: Communication between the dentist and the child is built on a dynamic process of dialogue, facial expression, and voice tone. Distraction techniques can be used in a variety of ways to assist the child and family |
| Context | Parenting practices (including discipline) have changed in the past 15 years due to different factors such and therefore, Paediatric dentists often have to deliver care to children who may not always be as compliant as they need to be |
| Results of | When speaking with children, truthfulness and reality in the explanations should |
| Study | always prevail.<br>Child's voice needs to be "heard" and addressed<br>Enhanced communication and partnership building improves comprehension and compliance with dental treatment |

Article 18:
| General information |  |
| --- | --- |
| Date of data extraction | 23/04/23 |
| Person extracting data | BT |
| PMID | 9790006 |
| Date of publication | 1998 |
| Author(s) | Rod Moore, Inger Brodsgaard, Tai-Kum Mao, Marc L Miller, Samuel F Dworkin |
| Title | Perceived need for local anaesthesia in tooth drilling among Anglo-Americans, Chinese, and Scandinavians |
| Type of Publication | Journal Article |
| Location of Study | America, Taiwan, China, Denmark and Sweden |

| Study Details |  |
| --- | --- |
| Study Design | Retrospective cross sectional |
| Aims of Study | Explore the link between pain experience and the social/cultural context in which it is experienced to see if there is any effect.<br>establish and describe ethnic differences or similarities and their importance to the perceived need for local anaesthesia for tooth drilling among adults using semi structured interviews. |
| Population | Dentists from America, Taiwan, China, Sweden and denmark. |
| Concept | A recorded semi structured qualitative interview was taken to determine<br>- the relative frequency of the use or nonuse of anaesthetic for similar tooth drilling in adults<br>- the reasons for nonuse of anaesthetic as reported by these dentists about their adult patients<br>- the distribution of reasons reported by dentists for why some patients were not using anaesthetic. |
| Context | Thematic categories for the non-use of anaesthetics<br>- Biomedical, Ethical Justifications-Dentist Talks Patients into Using Anaesthetic.<br>- Pain Tolerated-Anaesthetic Unnecessary, or pts not used to receiving anaesthetic<br>- Avoiding Disturbing Effects of Numbness: for all but chinese<br>- Fear of anaesthetic injection: which was often stated to be worse than the |
|  | drilling<br>- Barometer of trust in the patient-dentist relationship<br>- Dentist Decides No Anaesthetic Needed-Patient Tolerates Pain.<br>- Economic Incentive-Anaesthetic Costs Extra; unique to the danes |
| Results of Study | "barometer of trust" in the dentist-patient relationship - possible link to a patient's level of relaxation, and its effect on modifying pain threshold.<br>The Chinese concept of suan or "sourish" sensation in tooth drilling is usually thought of as tolerable. Thus, the Chinese concept of suan directly impacts the expected interactions between patients and dentists from within the cultural framework.<br>Studies of dental anxiety in Asiatic patients also suggest that communication between dentist and patients is "one way," with dentists deciding when pain control is appropriate. This study suggests that this one-way communication works for some patients but that undue fear of injections may be reinforced in other patients by Chinese dentists.<br>The possibilities for learning anxiety or pain beliefs through social interactions with dentists or friends and family who express their own ways of thinking about pain and coping are enormous. Self-fulfilling prophecies are perhaps the mechanisms that drive cultural coping strategies, for better or worse. Encouraging a patient to express concerns or discomfort allows the practitioner to communicate without bias and in terms with which the patients feel most comfortable. This can improve the feeling of trust in the interaction, which can in itself have healing effects |

| General information |  |
| --- | --- |
| Date of data extraction | 26/4/23 |
| Person extracting data | JT |
| PMID | 9790006 |
| Date of publication | 1998 |
| Author(s) | Rod Moore, Inger Brodsgaard, Tai-Kum Mao, Marc L Miller, Samual F Dworkin |
| Title | Perceived need for local anaesthesia in tooth drilling among Anglo-Americans, Chinese, and Scandinavians |
| Type of Publication | Journal article |
| Location of Study | Seattle, WA<br>Columbus, OH<br>Taipei, Taiwan<br>China<br>Arhus, Denmark<br>Goteborg and Linkoping, Sweden |
| Study Details |  |
| Study Design | Cross sectional |
| Aims of Study | Explore ethnic differences in perceptions of pain and the need for local anaesthesia for tooth drilling among age- and gender-matched Anglo-American, Mandarin Chinese, and Scandinavian dentists (n = 129) and adult patients |
| Population | Patients and dentists from:<br>Seattle, WA<br>Columbus, OH<br>Taipei, Taiwan<br>China<br>Arhus, Denmark<br>Goteborg and Linkoping, Sweden |
| Concept | Structured interview questions asked to dentists from various geographic locations/ethnic background:<br>- the relative frequency of the use or nonuse of anaesthetic for similar tooth drilling in adults<br>- the reasons for nonuse of anaesthetic as reported by these dentists about their adult patient<br>- the distribution of reasons reported by dentists for why some patients were not using anaesthetic |
| Context | Dental clinic |
| Results of Study | Reasons for not using anaesthetic:<br>- Dentists from Sweden, China, Denmark said that patients are not used to |
|  | anaesthetic so do not feel it is necessary. Chinese patients believe the pain is only “suan”/sourish and not painful<br>- American dentists most frequently said patients wanted to avoid anaesthetic to avoid feeling of numbness after the appointment and embarrassment of drooling<br>- Anglo-American dentists said their patients feared the pain of injections, usually pointing out that this anxiety was worse than the drilling sensations.<br>- Danish dentists report anaesthetic not needed due to “barometer of trust” in dentist-patient relationship<br>Description of anaesthetic by dentists:<br>- American dentists would use the phrase “pinch”, “disfomfort”, “uncomfortable”<br>- Danish dentists described injection as a little “prick” or “pressure” |

Article 19:
| General information |  |
| --- | --- |
| Date of data extraction | 23/4/2023 |
| Person extracting data | RB |
| PMID | ISSN0003-3006/88/ |
| Date of publication | 1987 |
| Author(s) | Peter A. Foreman |
| Title | Practical Patient Management: The Integrated Approach |
| Type of Publication | Journal piece |
| Location of Study | Auckland Hospital, New Zealand |
| **Study Details** |  |
| Study Design | NA |
| Aims of Study | Improve the delivery of dental care to anxious patients |
| Population | Anxious patients requiring dental care |
| Concept | - Pharmacologic approach<br>- GA is risky, IV is a good medium between GA and LA, RA is also a good option<br>- Behaviour approach<br>- Interpersonal and communication skills “iatrosedation” (hypnosis, biofeedback, relaxation training)<br>- Integrated approach<br>- Pharmacologic and behavioural skills |
|  | - Preoperative management = trust building exercise<br>- During sedation recovery, the doctor should stay with the patient so the patient experiences the reduction of stresses associated with the dental procedure in the doctor's presence (this will strengthen the desirable response of being with the doctor) |
| Context | Dental setting |
| Results of Study | - Integrated behavioural and pharmacological approach is most beneficial for the patient<br>- Groundwork for these patient skills should be developed during undergrad years |
| Date of data extraction | 25/04/23 |
| Person extracting data | BT |
| PMID | ISSN0003-3006/88/ |
| Date of publication | 1988 |
| Author(s) | Peter A foreman |
| Title | Practical Pain Management: The Integrated Approach |
| Type of Publication | Opinion piece |
| Location of Study | New Zealand |
| **Study Details** |  |
| Study Design | NA |
| Aims of Study | To bring attention to the need for integrating behavioural approaches to pain management in a dental setting, esp since too much emphasis is made on pharmacological management. |
| Population | Anxious and fearful dental patients. |
| Concept | A dentist recounts his experiences and research regarding the integration of behavioural management techniques to manage dentally fearful and anxious patients.<br>These approaches include: the use of effective interpersonal and communication skills, "iatrosedation" ,30 and patient education; and may include subjects such as the role of suggestion and hypnosis, desensitisation, biofeedback, relaxation training, and others.<br>Also, he states that it is important to understand what the patient fears and their previous dental experiences, so that behavioural and pharmacological management techniques can be catered to the individual. |
| Context | Too much emphasis is placed, during education, to manage pain pharmacologically, which the author thinks is used to circumvent anxiety, rather than attempt to understand, confront, and deal with the problem of fear from a behavioural standpoint.<br>At the same time, steps into behavioural management studies have been started, and the author believes that it is important to integrate these into practice. |
| Results of Study | The clinician's interpersonal and communication skills can exert at least as much sedative or calming effect on the patient as the drug itself<br>Integrated behavioural/pharmacologic approach has been successfully used<br>- Preop management - iatrosedative method: The initial interview is essentially a trust-building exercise, within the context of an empathic emotional environment. Also to elicit what fears they have and previous dental experiences. By interrupting their pain/anxiety cycle, pts become more trusting and receptive<br>- Intraop management: careful pre planning to avoid fear producing situations. Verbal and nonverbal communications are aimed at minimising feelings of helplessness, dependency, and the unknown; along with the use of euphemistic, nontechnical and nonthreatening language, positive suggestion. It is essential that verbal communications are continued throughout.<br>- Post Op management: dr should remain with pt, so that they are in the presence of the dr as they experience the reduction of stresses. Being there while the negative reinforcer is removed helps to reduce the desire to escape the dental office once procedure is over. Should also have a brief, |
|  | friendly farewell to strengthen the desire to return.<br>These cumulative calming effects eventually progress to a stage where many formerly fearful patients are eventually able to accept dental procedures while fully conscious or in the presence of only very light sedation. |

Article 20:
| General information |  |
| --- | --- |
| Date of data extraction | 23/04/23 |
| Person extracting data | BT |
| PMID | 6205032 |
| Date of<br>publication | July 1984 |
| Author(s) | Joel B Epstein |
| Title | Understanding placebos in dentistry |
| Type of Publication | Journal article |
| Location of<br>Study |  |

| Study Details |  |
| --- | --- |
| Study Design | Review Article |
| Aims of Study | To understand placebo actions and limitations so that it may be successfully applied to the management of patients with orofacial pain |
| Population | Dental patients who are suffering from myofascial pain dysfunction syndrome (mainly) |
| Concept | - |
| Context | - |
| Results of Study | Placebos may have pharmacological effects (endorphin, steroid and catecholamine system)<br>Psychological effects such as patient expectation, suggestion and relaxation may be important in placebo reaction<br>Patient practitioner relationship in the delivery of care has been demonstrated<br>Placebo effects may be the most significant in symptoms of chronic conditions including pain, depression and anxiety.<br>Placebos has been shown to work in the context of MPD, and may be applicable to other chronic head and neck conditions. |

| General information |  |
| --- | --- |
| Date of data | 26/4/2023 |
| extraction |  |
| Person extracting data | SN |
| PMID | 6205032 |
| Date of<br>publication | July 1984 |
| Author(s) | Joel B Epstien |
| Title | Understanding Placebos in Dentistry |
| Type of<br>Publication | Review Article |
| Location of<br>Study |  |
| **Study Details** |  |
| Study Design | Review Article |
| Aims of Study | To examine why and how the placebo effect works in the management of patients with orofacial pain. |
| Population | Use of Placebos |
| Concept | There is a current lack of understanding about the relief afforded by placebos , especially in dentistry. This article aims to examine the implication of the use of |
|  | placebos in the broader medical context. |
| Context |  |
| Results of Study | Placebos:<br>- Mechanism: affects blood pressure, lipoproteins ,electrolytes- mechanism of action - physiology and psychological<br>- It is thought that placebos release endorphins<br>- Placebo effect can be seen in isolation or with other treatments<br>- Dizziness, nausea, vomiting include some of the adverse effects of placebos<br>- Patient/doctor relationships can impact the outcome of therapy<br>- Placebo effect most impactful in chronic pain, depression, anxiety<br>- Placebo use has been seen in oro-facial pain |

Article 21:
| General information |  |
| --- | --- |
| Date of data extraction | 25/4/2023 |
| Person extracting data | SN |
| PMID | 29179365 |
| Date of publication | 2015 |
| Author(s) | Council of Clinical Affairs |
| Title | Behaviour Guidance for the Pediatric Dental Patient |
| Type of Publication | Reference manual |
| Location of Study | American Academy of Pediatric Dentistry |
| **Study Details** |  |
| Study Design | N/A |
| Aims of Study |  |
| Population | Dental Practitioners treating childhood dental diseases |
| Concept | This manual discusses the way in which dentists need to employ behaviour guidance to help treat paediatric dental patients. It include traits of empathy, cultural sensitivity, flexibility and awareness of the science behind behaviour |
| Context | Paediatric patients and dental practitioners |
| Results of Study | Discusses:<br>- Child's behaviour- dependent of their attributes, parents, developmental status - importance of being aware and accurately assessing each child<br>- Interaction of the child with the dental environment and dental team<br>- Obtaining informed consent<br>- Accurately diagnosis the clinical condition |
|  | - How to promote through communication appropriate behaviour - behavioural guidance - different techniques it encompasses<br>- Documentation |
| Date of data extraction | 26/4/23 |
| Person extracting data | BT |
| PMID | 29179365 |
| Date of publication | 2015 |
| Author(s) | Council of Clinical Affairs |
| Title | Behaviour Guidance for the Pediatric Dental Patient |
| Type of Publication | Reference manual |
| Location of Study | American Academy of Paediatric Dentistry |
| Study Details |  |
| Study Design | Review of the evidence |
| Aims of Study | Offer an up to date review on the behaviour guidance techniques, both non pharmacological and pharmacological, to manage paediatric dental patients (infants, children, adolescents, and persons with special health care needs) in order to alleviate anxiety, nurture a positive dental attitude, and perform quality oral health care safely and efficiently. |
| Population | Targeting health care providers, parents, and other interested parties about influences on the behaviour of paediatric dental patients and the many behaviour guidance techniques used in contemporary paediatric dentistry |
| Concept | Different behaviour guidance techniques dentists can use in paediatric dental setting<br>Behaviour guidance is the process by which practitioners help patients identify appropriate and inappropriate behaviour, learn problem solving strategies, and develop impulse control, empathy, and self-esteem. Different variables that need to be considered include:<br>- Knowing the predictors of child behaviours<br>- Pain assessment and management during treatment: Prevention or reduction of pain during treatment can nurture the relationship between the dentist and the patient, build trust, allay fear and anxiety, and enhance positive dental attitudes for future visits<br>- Documentation of child behaviour<br>- Deferring tx when needed e.g. pt becomes too hysterical or uncontrollable |
| Context | Dental setting |
| Results of | Predictors of child behaviours: |
| Study | - Patient attributes<br>- Parental Influences<br>- Orientation to dental environment<br>- Patient assessment<br>- Dentist/dental team behaviour: attitude, body language, and communication skills are critical to creating a positive dental visit.<br>Examples of good behaviours include: giving clear and specific instructions, an empathetic communication style, and an appropriate level of physical contact accompanied by verbal reassurance<br>Summary of basic behaviour guidance recommendations<br>- Communication guidance:<br>- Positive pre visit imagery: show positive photographs or images of dentistry and dental treatment in the waiting area<br>- Direct obs: Patients are shown a video or are permitted to directly observe a young cooperative patient undergoing dental treatment<br>- Tell, show, do:<br>- Ask, tell, ask<br>- Voice control: deliberate alteration of voice volume, tone, or pace to influence and direct the patient's behaviour<br>- Nonverbal communication: appropriate contact, posture, facial expression, and body language<br>- Positive reinforcement and descriptive praise: Descriptive praise emphasises specific cooperative behaviours rather than generalised praise<br>- Distraction<br>- Memory restructuring: Memories of negative or difficult events are |
|  | restructured into positive memories by using information suggested after the event has taken place<br>- Parental presence/absence:<br>- Communication techniques for parents<br>- NO inhalation<br>Summary of advanced behavioural guidance:<br>- Protective stabilisation: the restriction of patient's freedom of movement, with or without the patient's permission, to decrease risk of injury while allowing safe completion of treatment<br>- Sedation<br>- GA |

